# A Novel Framework for Evaluating the Clinical Reasoning Process of Large Language Models: A Comparative Study in Nephrology

**DOI:** 10.1101/2025.09.04.25334460

**Authors:** Yuichiro Yano, Hiroaki Kakizaki, Hajime Nagasu, Seiji Kishi, Takeo Koshida, Yoshihito Nihei, Akira Hirano, Masaomi Nangaku, Hirotake Mori, Toshio Naito, Mizuki Ohashi, Shoichi Maruyama, Isao Matsui, Yoshitaka Isaka, Yusuke Suzuki, Naoki Kashihara

**Author notes:** **Corresponding author:** Yuichiro Yano, MD, PhD, FAHA, Professor, Department of General Medicine, Director, AI Incubation Farm, Juntendo University Faculty of Medicine, ZIP: 113-8421 2-1-1, Hongo, Bunkyo-Ku, Tokyo, JAPAN.

## Abstract

Although interest in the application of large language models (LLMs) in medicine is growing, accuracy evaluations have largely relied on static knowledge tests. However, discussions on clinical reasoning, the process most critical to real-world practice, remain limited. In this study, we propose a novel framework to evaluate not the final diagnosis generated by AI, but the reasoning process itself.

This study proposes a novel framework that systematically evaluates the capabilities of LLMs (OpenAI GPT-o3, Gemini 2.5 Pro, DeepSeek-R1, Llxsama4-Marveric) by deconstructing the clinical reasoning process into discrete cognitive steps. We focused on nephrology cases, which often involve multiple organ systems and diverse pathologies, thus requiring a high level of reasoning. The four nephrologists independently evaluated the outputs. Our evaluation of four leading LLMs revealed that while Gemini 2.5 Pro demonstrated the best overall performance, all models exhibited common weaknesses in advanced, synthetic tasks such as “formulating differential diagnoses with rationale” and “treatment planning,” particularly in dynamically changing clinical scenarios. Furthermore, a notable finding of our research is that the highest-performing model was not the most computationally intensive, demonstrating that reasoning quality and computational efficiency are not in a simple trade-off.

In conclusion, our step-by-step evaluation method is an effective approach for identifying the specific strengths and weaknesses in an LLM’s clinical reasoning. The weaknesses identified, particularly in formulating a differential diagnosis with a clear rationale and developing comprehensive treatment plans for dynamic scenarios, should become a primary target for future model development and for the creation of support system.

## Introduction

While the application of large language models (LLMs) in medicine is gaining significant attention, their performance evaluation has predominantly relied on static knowledge tests, such as medical licensing exams. These methods fail to adequately reflect the dynamic reasoning process of actual clinical practice, where physicians iteratively form and revise hypotheses based on ongoing patient interactions and test results.^1,2^

Building on these advancements, our study proposes a novel evaluation framework where specialists scrutinize the “clinical reasoning process itself,”^3^ not just the final output of an AI. We segment each clinical case into three distinct stages: patient history, test results, and subsequent clinical course. At each stage, the validity of the AI’s diagnostic reasoning is assessed in a step-by-step manner by expert physicians. Furthermore, we conduct a comparative analysis of leading LLMs with different architectures and levels of openness (OpenAI GPT-o3, Gemini 2.5 Pro, DeepSeek-R1, Llama4-Marveric) to investigate how these structural differences impact the quality of clinical reasoning. By also recording and analyzing efficiency metrics such as response time and token count, we aim to examine their correlation with reasoning ability and assess each model’s effectiveness from a practical standpoint, considering the balance between accuracy and computational resources.

To ground our evaluation in a domain that is both complex and structured, this study specifically focused on clinical reasoning within the field of nephrology.^4^ This specialty is chosen because kidney diseases are often interconnected with multiple organ systems and present with diverse pathologies, demanding a high degree of clinical reasoning. Despite this complexity, the field has well-systematized diagnostic algorithms and clinical patterns, making it an ideal testbed for validating the reasoning process.

## Methods

A detailed description of the methods can be found in the separate supplementary file. Briefly, four nephrologists used the Delphi method to select ten cases that met the inclusion criteria from over one hundred case reports. As permission could not be obtained from the journal for one of the cases, a total of nine cases were included in the final analysis. For each case, nine questions regarding clinical reasoning were established. The evaluation of the LLM outputs was conducted on July 20, 2025 (JST). Each question was scored on a 3-point scale (0 = Incorrect, 1 = Reasonable but not the best, 2 = Correct). The four nephrologists independently evaluated the outputs in a blinded manner, without knowing the model names. The results were then aggregated and analyzed by a single, blinded researcher.

## Results

### Overall Diagnostic Reasoning Performance

Across all clinical cases and reasoning questions, a statistically significant difference in overall performance was observed among the four LLMs (**Figure 1A**). Gemini 2.5 Pro achieved the highest average score (7.57), followed by OpenAI’s O3 (7.39), DeepSeek-R1 (7.13), and Llama 4 Maverick (6.23).

**Figure 1A.**
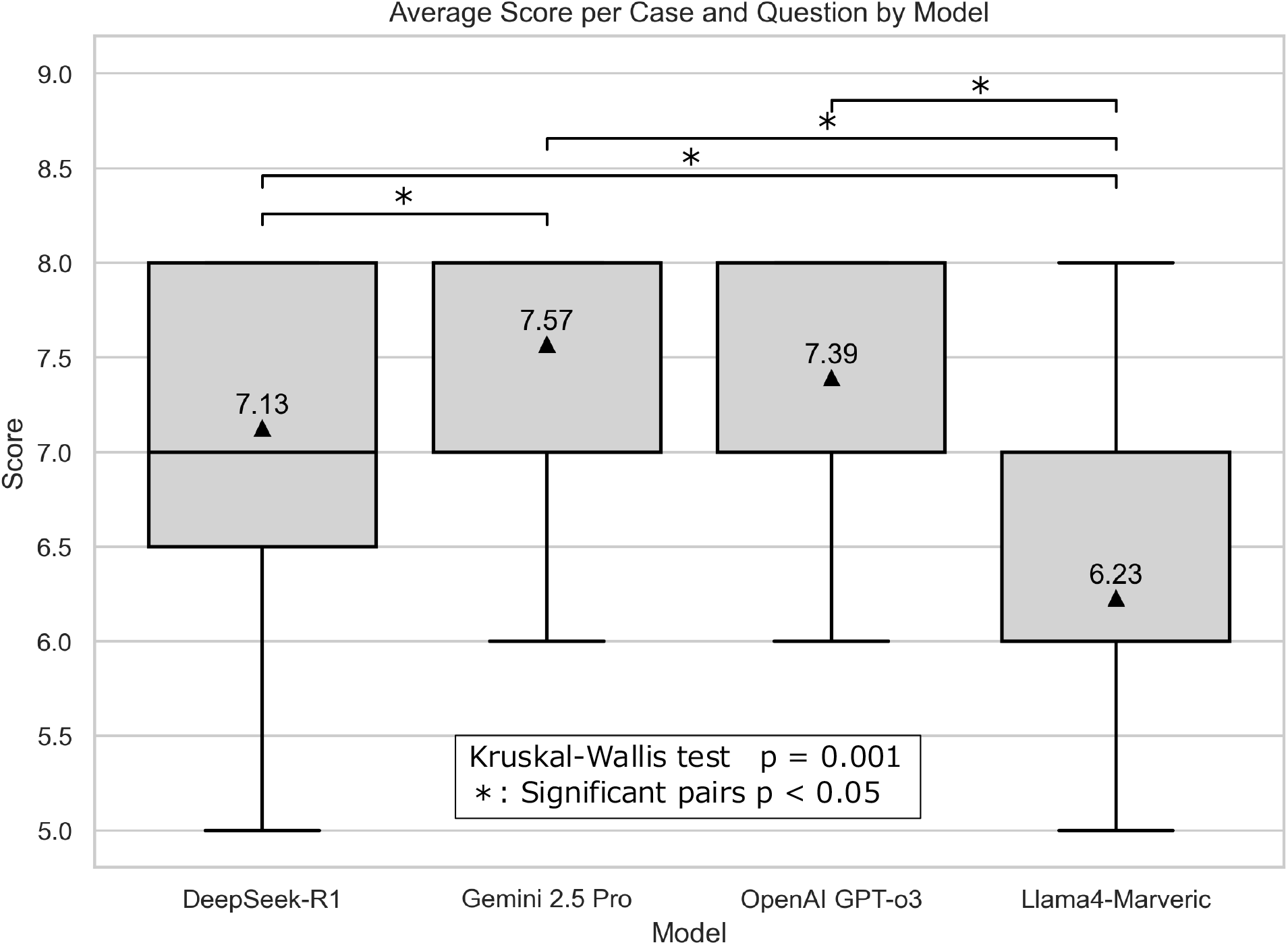
Scores by model for each case and question. Scores for each case and question are displayed by model as box-and-whisker plots. Overall differences were assessed with the Kruskal – Wallis test; when significant, pairwise comparisons between models were performed with Holm’s correction for multiple testing.

### Performance on Specific Reasoning Tasks

Based on the performance across all models ((**Figure 1B**), the tasks that proved most challenging were Question 2 (“Differential diagnoses and rationale”) and Question 7 (“Treatment Planning”), which received the lowest mean scores of 6.56 and 6.58, respectively. In contrast, the models performed best on Question 1 (“Summarizing medical problems”) and Question 6 (“Reassessment of the differential diagnoses”), which both achieved the highest mean score of 7.50.

**Figure 1B.**
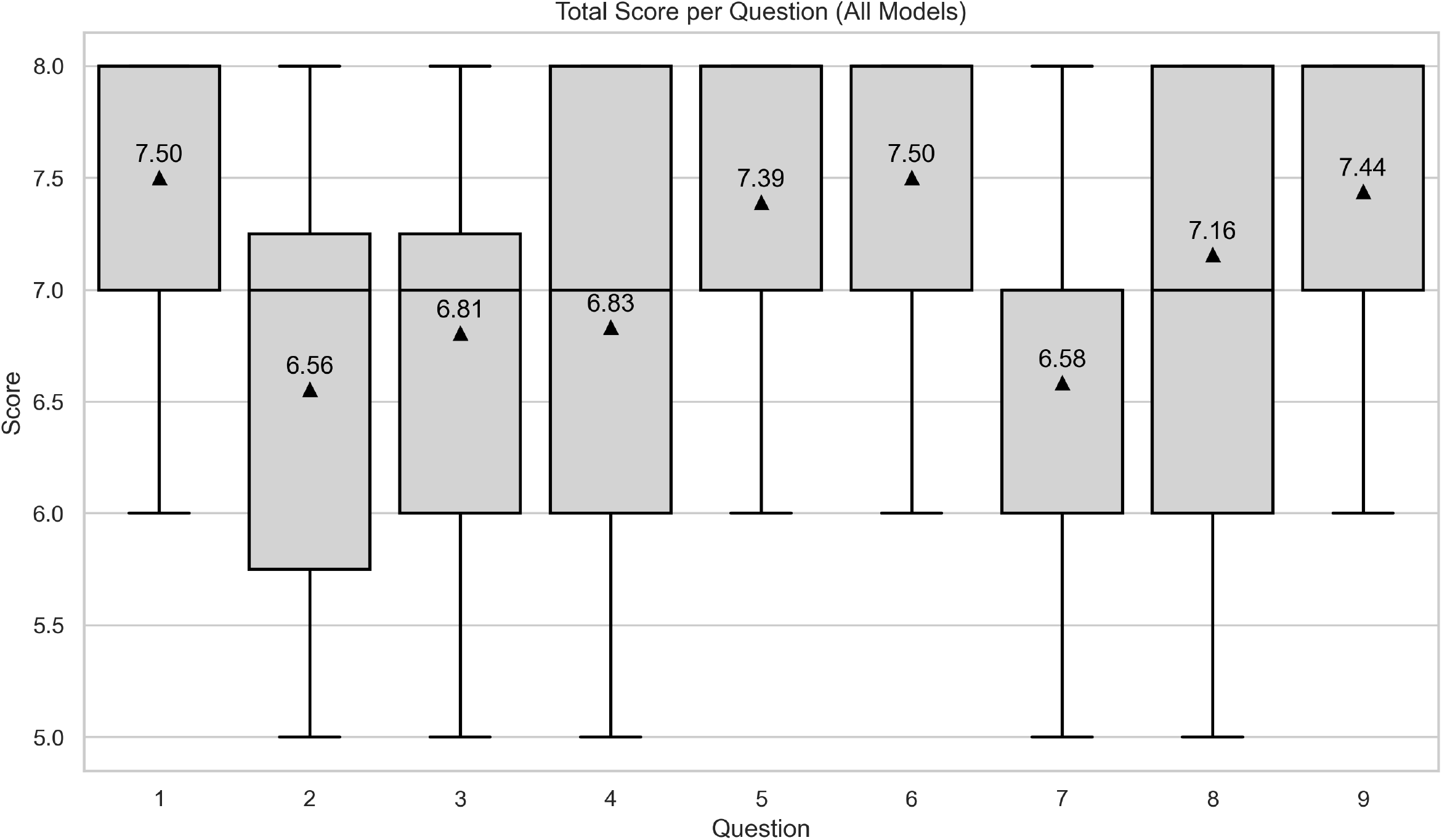
Distribution of Scores by Clinical Reasoning Question (All Models Combined) Box-and-whisker plots show the distribution of scores for each of the nine clinical reasoning questions, aggregating the results from all four LLMs. The box represents the interquartile range (IQR), the line inside the box indicates the median, and the whiskers show the range of the data. Triangles mark the mean score for each question. Questions (Q1–Q9): Q1, summary of the medical problem; Q2, differential diagnoses and rationale; Q3, necessary physical examinations and rationale; Q4, plan for investigations/tests; Q5, interpretation of test results; Q6, reassessment of the differential diagnoses; Q7, treatment planning; Q8, evaluation of treatment; Q9, management in case of clinical worsening.

**Figure 1C** shows heatmap of average scores by model for each clinical reasoning question. Gemini 2.5 Pro demonstrated superior or competitive performance, particularly in complex tasks such as reconsidering the differential diagnosis (Q6, score: 7.89), interpreting test results (Q5, score: 8.00), and formulating plans for worsening conditions (Q9, score: 8.00).

**Figure 1C.**
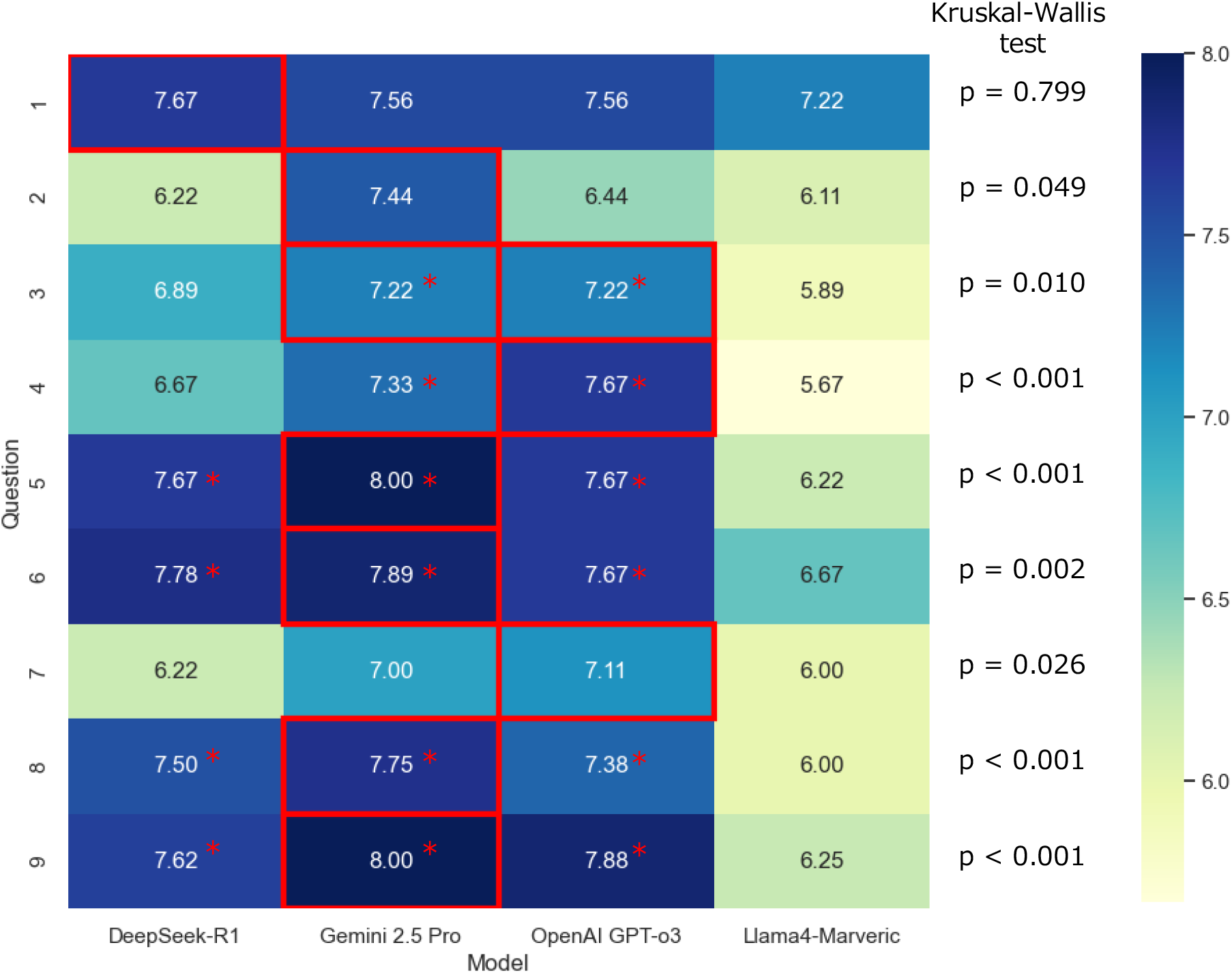
Heatmap of average scores by model for each clinical reasoning question. The values are presented as a heatmap, where red outlines highlight the highest-scoring model for each question. Statistical significance was assessed using the Kruskal–Wallis test, followed by pairwise comparisons with Holm’s correction for multiple testing. An asterisk (^*^) indicates a statistically significant difference in performance compared to Llama 4 Maverick

**Figure 1D.**
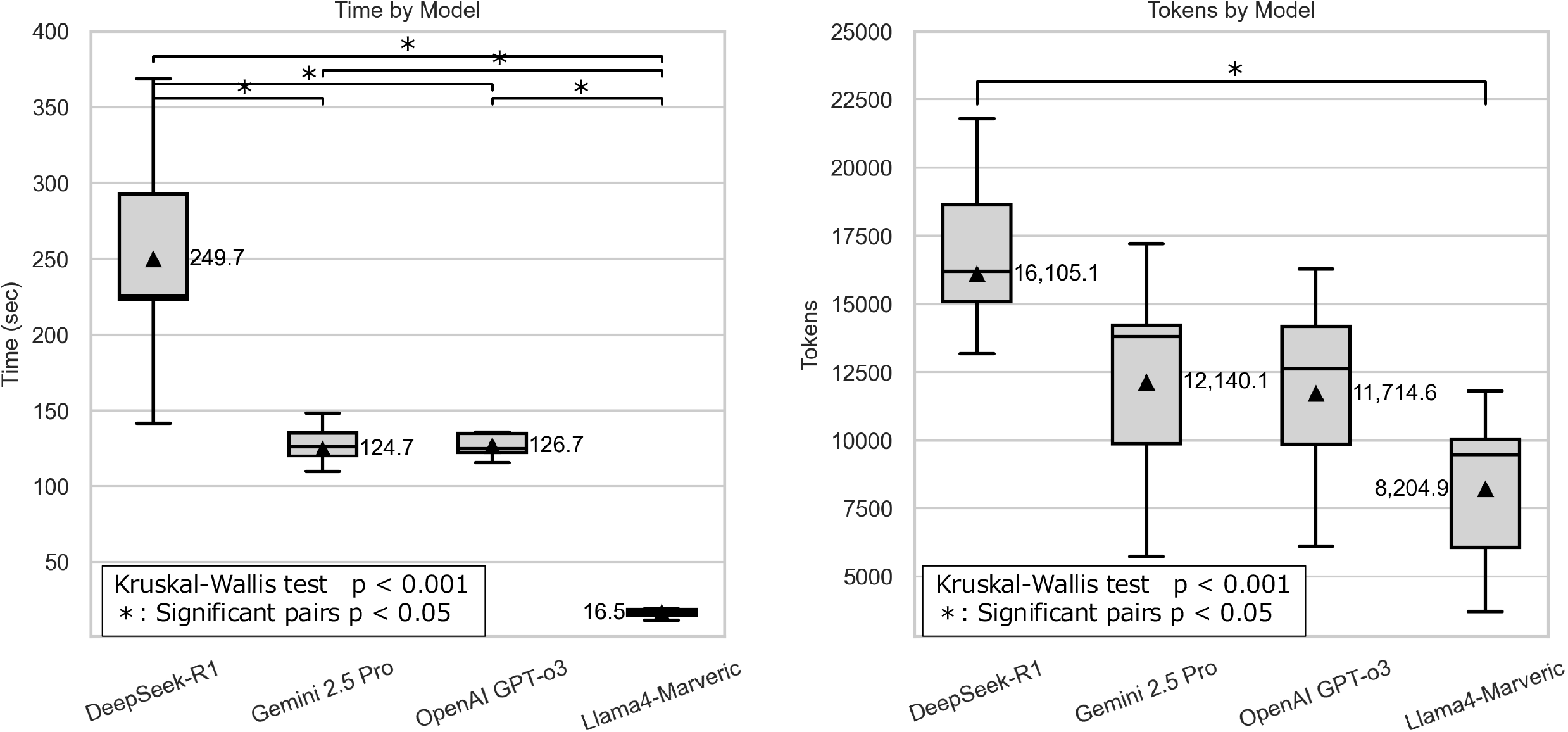
Comparison of Computational Efficiency by Model: Time and Token Usage. Box-and-whisker plots compare the distribution of **(A)** time required and **(B)** tokens consumed per case for each of the four LLMs. The box represents the interquartile range (IQR), the line inside the box is the median, and the whiskers show the range of the data. Triangles mark the mean values for each model. Statistical differences were assessed using the Kruskal–Wallis test, followed by pairwise comparisons with Holm’ s correction. Brackets at the top of the plots indicate statistically significant differences between model pairs

### Efficiency and Resource Usage

While Gemini 2.5 Pro demonstrated high efficiency with a mean response time of 133.2 sec, the greatest computational cost was borne by DeepSeek-R1, which was the slowest (mean time: 260.3 sec) and consumed the most tokens (16,687.2). In contrast, Llama 4 Maverick had the shortest response time and the lowest token usage.

## Discussion

A key novelty of this study lies in its introduction of a framework that deconstructs the clinical reasoning process into discrete cognitive steps and evaluates an AI’s capability at each stage. Previous landmark studies, such as those on Google’s AMIE and Microsoft’s MAI-DxO,^1,2,5^ made significant progress by shifting evaluation away from static tasks toward more dynamic processes like conversational dialogue and sequential information gathering. However, these studies did not pinpoint which specific cognitive tasks within the broader reasoning process represented a model’s weak points.

Our step-by-step evaluation approach revealed specific weaknesses in the models’ clinical reasoning. Notably, the most challenging tasks were Question 2 (“Differential diagnoses and rationale”) and Question 7 (“Treatment Planning”). This suggests that while AIs may excel at discrete tasks like summarizing information and interpreting test results, a capability gap remains in more advanced, synthetic skills like planning optimal interventions.^6^ This inadequacy was particularly evident in cases requiring dynamic adjustments due to a dramatic change in the clinical condition. For example, in a case involving the sudden onset of nocardiosis during treatment for nephrotic syndrome, many of our expert evaluators indicated that the treatment plans proposed by the models were insufficient.

Our research reveals a nuanced relationship between reasoning quality and computational efficiency, challenging the assumption of a simple trade-off. The highest-performing model, Gemini 2.5 Pro, was not the most resource-intensive; in fact, the greatest computational cost was borne by the lower-performing DeepSeek-R1, which was the slowest and consumed the most tokens. This finding is critical for clinical implementation, as it proves that superior reasoning quality does not necessarily demand the highest computational cost, allowing for the selection of AI tools that balance high accuracy with the practical demands of a clinical workflow.

## Acknowledgments

This research was partially funded by the Advanced Medical Personnel Training Program (principal investigator: TN) and was supported by the Ministry of Education, Culture, Sports, Science, and Technology.

## Data Availability

The datasets generated or analyzed during this study are available from the corresponding author on reasonable request.

## Conflicts of Interest

None declared.

## Author contribution

YY conceptualized the study and developed the methodology. HN, SK, TK, and YN curated the data. HK and MO conducted the formal analysis. MN, SM, IM, YI, YS, and NK supervised the study. YY drafted the manuscript, and all authors reviewed, edited, and approved the final version.

## Supplementary Material (PDF)

Supplementary methods

Supplementary Table 1. Prompts and Evaluation Criteria for the Phased Assessment of Clinical Reasoning

Supplementary Table 2. Responses from Four Large Language Models to Clinical Reasoning Questions Across Nine Cases

## Notes

### Competing Interest Statement

The authors have declared no competing interest.

