## Supplementary method and tables for "A Novel Framework for Evaluating the Clinical Reasoning Process of Large Language Models: A Comparative Study in Nephrology": Supplementary MaterialπÇÇSeptember 5 2025.pdf

Brief title: Evaluating LLM Clinical Reasoning in Nephrology

Yuichiro Yano;[1,2] Hiroaki Kakizaki:[3] Hajime Nagasu;[4] Seiji Kishi;[4] Takeo Koshida;[5] Yoshihito Nihei;[5] Akira Hirano;[4] Masaomi Nangaku;[6] Hirotake Mori; [1] Toshio Naito; [1] Mizuki Ohashi;[1] Shoichi Maruyama;[7] Isao Matsui;[8] Yoshitaka Isaka;[8] Yusuke Suzuki;[5]and Naoki Kashihara[4]

1. Department of General Medicine, Juntendo University Faculty of Medicine, Tokyo, Japan
2. Artificial Intelligence Incubation Farm, Juntendo University Faculty of Medicine, Tokyo, Japan
3. PeopleMedia, Inc, Tokyo, Japan
4. Department of Nephrology and Hypertension, Kawasaki Medical School, Kurashiki, Okayama, Japan
4. Department of Nephrology, Juntendo University Faculty of Medicine, Tokyo, Japan
6. Division of Nephrology and Endocrinology, The University of Tokyo Graduate School of Medicine, Tokyo, Japan
7. Department of Nephrology, Nagoya University Graduate School of Medicine, Aichi, Japan
8. Department of Nephrology, Osaka University Graduate School of Medicine, Suita, Japan.

**Corresponding author:**
Yuichiro Yano, MD, PhD, FAHA
Professor, Department of General Medicine
Director, AI Incubation Farm
Juntendo University Faculty of Medicine
ZIP: 113-8421 2-1-1, Hongo, Bunkyo-Ku, Tokyo, JAPAN


***Methodology for Case Selection to Evaluate Large Language Models Using the Delphi Technique***

To select clinical cases for evaluating four large language models (LLMs), we employed a structured methodology adapted from established consensus group methods. The process was guided by the principles of the Delphi technique, as described by Humphrey-Murto et al.,[1] and was conducted by a panel of four nephrology specialists.

**1. Preparatory Phase: Establishing Selection Criteria**

An expert panel, comprising four nephrologists (HN, SK, TK, and YN), conducted a preliminary discussion to establish consensus on the essential criteria for case selection in the context of clinical reasoning in nephrology. The agreed-upon criteria were as follows:

i. **Comprehensive Coverage of Nephrology:** The cases should collectively cover a wide spectrum of the field of nephrology.
ii. **Avoidance of Simple Cases:** Cases with a limited or straightforward differential diagnosis were to be avoided.
iii. **Complexity of Diagnosis and Pathophysiology:** Priority was given to cases involving complex diagnostic processes or intricate pathophysiology.
iv. **Balance between Complexity and Rarity:** While prioritizing complexity (criterion iii), extremely rare diseases infrequently encountered in clinical practice were to be avoided where possible.
v. **Potential for High Educational Value:** Cases should present opportunities for significant learning, such as highlighting potential diagnostic errors or critical decision points.
vi. **Complexity in Post-Diagnostic Management:** The case should involve complex considerations regarding post-diagnostic management and the course of treatment.

**2. Literature Search and Candidate Case Compilation**

To minimize the possibility that the selected cases were part of the training data for the LLMs, we restricted our search to cases published after April 5, the release date of Llama 4, the newest model among the four being tested. Using Medical Subject Headings (MeSH) terms ("Kidney Diseases"[MeSH Terms] OR "Metabolic Diseases"[MeSH Terms] OR "Hypertension"[MeSH Terms] NOT

("Neoplasms"[MeSH] OR "Infant"[MeSH Terms] OR "Child"[MeSH Terms] OR "Adolescent"[MeSH Terms]) AND ("Humans"[MeSH Terms] AND "Case Reports"[Publication Type] AND ("2025/04/06"[Date - Publication] : "2025/05/06"[Date - Publication]))), we searched the literature published in the subsequent month, resulting in a preliminary list of 104 candidate cases.

**3. The Delphi Process for Case Evaluation**

**Round 1:**

1. A non-voting facilitator (AH) distributed the list of candidate cases—including a summary of each case and a link to the full-text PDF where possible—and an evaluation form to the expert panel. When the number of available cases was sufficient, open-access articles were prioritized to avoid copyright issues.
2. Each panelist (HN, SK, TK, and YN) reviewed the titles of all candidate cases and excluded those deemed inappropriate.
3. Each panelist (HN, SK, TK, and YN) independently and anonymously rated the "suitability" of each candidate case on a 3-point Likert scale ((0 = Incorrect, 1 = Reasonable but not the best, 2 = Correct). The specific evaluation criteria for each question are detailed in **Supplementary Table 1**. Cases in which two or more panelists assigned a scale of 1 to the same question were excluded from further consideration.
4. Panelists were required to provide a written rationale for their rating for each case, outlining the key criteria they considered and any concerns.
5. Completed evaluation forms were returned to the facilitator.

**Analysis and Feedback (Post-Round 1):**

1. The facilitator collected the responses and computed the mean rating for each case as part of the descriptive statistical analysis.
2. The facilitator compiled and synthesized the panelists' written comments for each case.
3. A feedback report, summarizing the quantitative scores and qualitative comments for each panelist, was prepared.

**Round 2:**

1. The facilitator distributed this feedback report to the panel along with the same list of candidate cases (or a list narrowed based on scores).
2. Panelists reviewed the group's aggregated results and feedback from their peers. They were then asked to re-evaluate the suitability of each case, with the option to revise their scores and rationale, before submitting their second-round evaluations to the facilitator.

**Consensus and Subsequent Rounds:**

1. The facilitator analyzed the results of Round 2. Consensus was determined based on pre-defined criteria, such as the stability of scores and a decrease in the variance of ratings (e.g., interquartile range).
2. If consensus was not achieved, a third round would be conducted in a similar manner. (Convergence is typically achieved within 2-3 rounds).

**4. Final Case Selection Meeting**

1. Based on the final results of the Delphi process (final scores, score distribution, and summarized comments), the facilitator prepared a shortlist of high-scoring cases (i.e., the top 16 cases).
2. The facilitator then convened a final consensus meeting (held in-person or online) with the expert panel. During this meeting, the shortlist and the complete, anonymized results of the Delphi rounds were presented.
3. In this final stage, the panel discussed the shortlisted cases not only on their individual merits but also in the context of the entire set, ensuring that the final selection of 10 cases met criterion i: Comprehensive Coverage of Nephrology.

**5. Final Analysis and Reporting**

The final set of 10 selected cases and the detailed rationale for their selection, including the results from the Delphi process and the final consensus meeting, were documented for reporting.

#### *System Architecture and Prompt Design for a Multi-Agent Clinical Reasoning Application*

We developed a clinical reasoning application using Dify(※1), a no-code AI development platform that enables the creation of AI agents leveraging large language models (LLMs) via API integration. Four different LLMs were employed in this study, each accessed through the Dify platform:

1. **DeepSeek** — *deepseek-reasoner (R1)*
2. **Google** — *Gemini 2.5 Pro Preview 03-25*
3. **OpenAI** — *o3*
4. **Meta** — *Llama-4-Maveric* (meta-llama/Llama-4-Maveric-17B-128E-Instruct-FP8), accessed via the together.ai(※2) API

All four LLM-based applications were constructed using an identical architecture comprising three AI agents, each responsible for a distinct phase of the clinical reasoning process.

**Agent 1: Initial Clinical Assessment**

The first agent processed the patient's chief complaint and brief clinical findings provided by a physician, producing:

1. Structured summary of the patient's medical problems.
2. Differential diagnoses with supporting rationale.
3. List of additional questions or examinations required to test the working hypotheses.
4. Recommended diagnostic tests, with their purposes and expected outcomes.

**Agent 2: Diagnostic Refinement**

The second agent incorporated newly obtained test results to:
5. Interpret the findings and highlight salient features.
6. Suggest the most probable diagnosis at this stage.
7. Propose a treatment plan based on the cumulative information.

**Agent 3: Therapeutic Evaluation**

The third agent integrated information on treatments administered and subsequent patient outcomes to:
8. Summarize strategies for monitoring treatment efficacy and safety.
9. Recommend contingency measures for cases where the patient's condition fails to improve or deteriorates unexpectedly.

This multi-agent system was used to perform end-to-end clinical reasoning for all cases evaluated in the study.

---

**Prompt Design**

The evaluation in this study was conducted by clinical experts, who were blinded to the identity of the LLM that generated each output. To ensure fairness and reproducibility under these conditions, careful prompt design was essential.

Prompts were created following the Dify format, consisting of three components:

1. **SYSTEM Prompt** — Defined the fundamental behavior of the model, including its role and expected interaction style.
2. **USER Prompt** — Contained the specific instructions and questions provided to the model.
3. **ASSISTANT Prompt** — Specified the expected structure and format of the model's responses.

All four LLMs were provided with the same prompt, without any model-specific tuning. While prompt optimization for each model could potentially improve performance, identical prompts were used in this study to maintain cross-model fairness.

The prompt design process was iterative, involving collaboration between clinical experts and software engineering specialists. The team discussed and articulated the clinical reasoning workflow observed in real-world medical practice, refining the prompts through repeated pilot testing.

A complete example of the prompt used in this study is provided in the Supplementary Material.

---

**※1** **https://dify.ai/jp**
**※2** **https://www.together.ai/**

---

#### System Overview Diagram of the Multi-Agent Clinical Reasoning Process

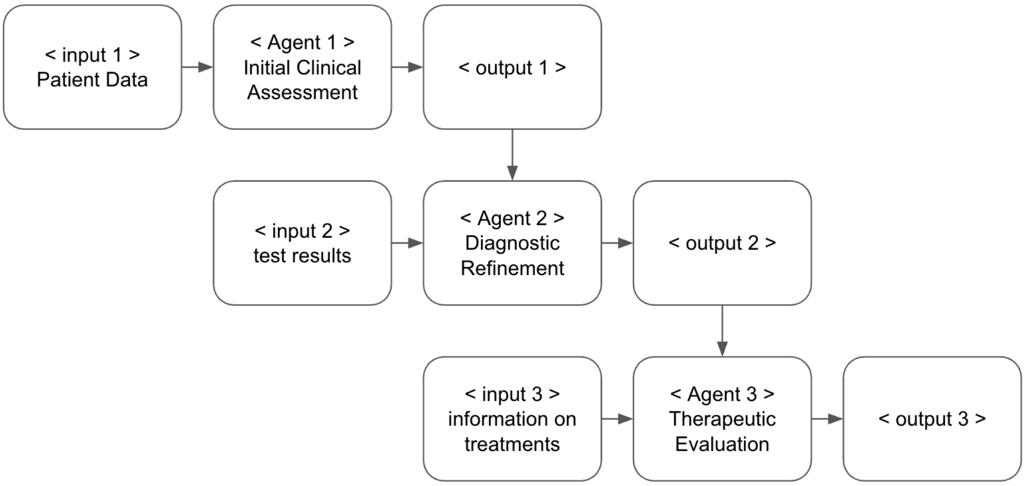


**Workflow of the multi-agent clinical reasoning system.** The system consists of three sequential agents: (1) Initial Clinical Assessment, (2) Diagnostic Refinement, and (3) Therapeutic Evaluation, each receiving specific inputs and producing structured outputs.

---

#### Screenshot of the Developed Application

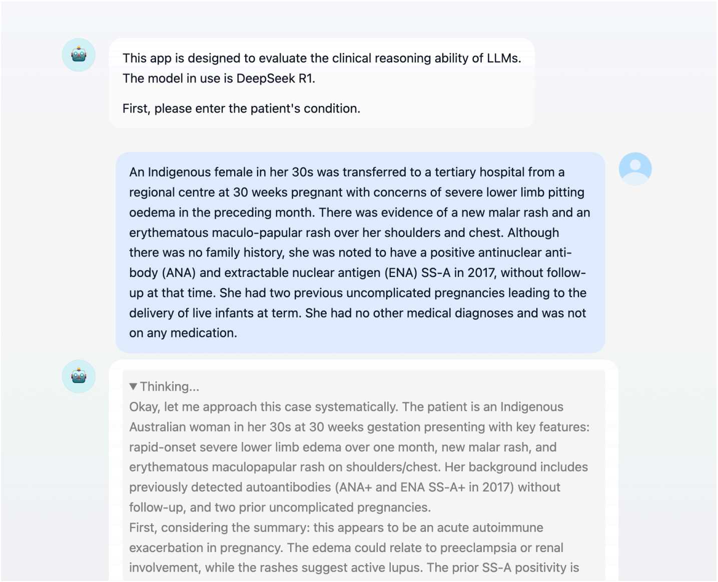

Medical Problem Summary

A 30s-year-old Indigenous Australian pregnant female at 30 weeks gestation, presenting with **acute-onset (1 month) severe bilateral lower limb pitting edema**, **new malar rash**, and **erythematous maculopapular rash** on shoulders/chest. Background includes prior positive ANA and SS-A antibodies (2017, untreated), and two uncomplicated pregnancies.

Differential Diagnoses

1. **Systemic Lupus Erythematosus (SLE) flare** (*most likely*) – Rash, edema, and prior autoantibodies suggest new-onset or reactivated SLE. Pregnancy is a known trigger.
2. **Preeclampsia with severe features** (*must not miss*) – Acute edema in third trimester risks eclampsia, HELLP, or fetal compromise.
3. **Polymorphic eruption of pregnancy (PEP)** – Benign pregnancy-related rash, but atypical with edema and malar involvement.

**Rationale**: SLE is prioritized due to classic malar rash, prior serology (SS-A linked to neonatal lupus), and edema (possible lupus nephritis). Preeclampsia cannot be missed due to maternal-fetal mortality risks. PEP is less likely without abdominal rash.

Targeted History/Examination

**History**:

- *Hypertension symptoms* (headache, visual changes): Differentiates preeclampsia.
- *Fetal movements*: Assesses fetal wellbeing in preeclampsia/SLE.

---

### Prompt Creation Screen

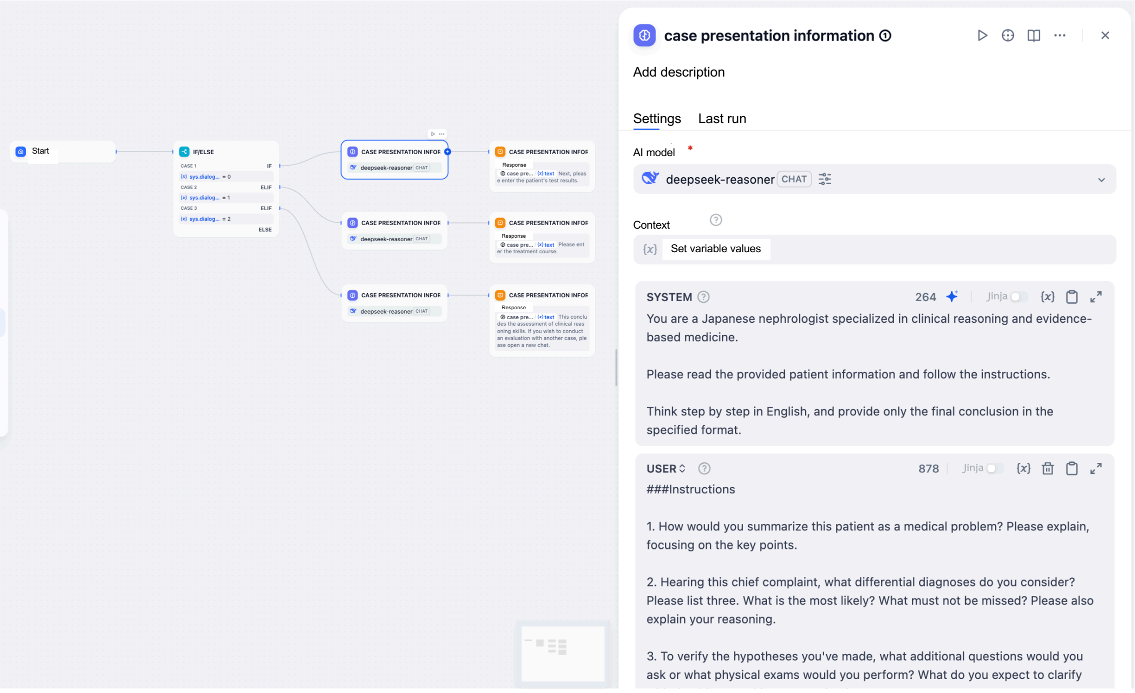


---

### Developed Prompt

**Agent 1: Initial Clinical Assessment**
SYSTEM Prompt
You are a Japanese nephrologist specialized in clinical reasoning and evidence-based medicine.

Please read the provided patient information and follow the instructions.

Think step by step in English, and provide only the final conclusion in the specified format.

**USER Prompt**
###Instructions

1. How would you summarize this patient as a medical problem? Please explain, focusing on the key points.

2. Hearing this chief complaint, what differential diagnoses do you consider? Please list three. What is the most likely? What must not be missed? Please also explain your reasoning.

3. To verify the hypotheses you've made, what additional questions would you ask or what physical exams would you perform? What do you expect to clarify with that history-taking or examination?

4. To narrow down the diagnosis, what tests do you plan to order? Please tell me the purpose for ordering those tests and the expected results.

###Response Style and Constraints

1. Please answer in English.
2. Keep the response concise and clear.
3. Base your reasoning only on the provided information and widely accepted medical knowledge. Avoid assuming any new facts.

ASSISTANT Prompt

###Output Format (Strictly Follow)

1. Include age, sex, time course, and key symptom characteristics to express the patient's problem concisely and accurately.

2. Be able to present a broad, prioritized differential diagnosis list that considers frequency and severity, and explain the rationale for it.

3. With the differential diagnosis in mind, list purposeful questions and physical examination techniques to confirm or rule it out, and clearly explain the reasons for each.

4. Develop a logical testing plan aimed at narrowing the differential diagnosis, taking into account cost-effectiveness and invasiveness.

###Response Style and Constraints

1. Please answer in English.
2. Keep the response concise and clear.
3. Base your reasoning solely on the provided information and widely accepted medical knowledge. Do not assume any new facts.

**Agent 2: Diagnostic Refinement**

SYSTEM Prompt
You are a Japanese nephrologist specialized in clinical reasoning and evidence-based medicine.

Please read the {{#sys.query#}} patient information and follow the instructions.

Think step by step in English, and provide only the final conclusion in the specified format.

**USER Prompt**
##### Instractions

5. How do you interpret these test results? Please explain the points of particular interest and what they mean in the context of this patient's condition.

6. Based on all the information, how has your diagnostic thinking changed? What is the most likely diagnosis (Working Diagnosis) at this point?

7. What kind of treatment plan would you create for this patient? Please also explain the rationale for your chosen treatment and other possible options.

###Response Style and Constraints

1. Please answer in English.
2. Keep the response concise and clear.
3. Base your reasoning only on the provided information and widely accepted medical knowledge. Avoid assuming any new facts.

ASSISTANT Prompt
###Output Format (Strictly Follow)

5. Explain the medical significance of the test values not merely as abnormal results, but by relating them to the patient's history and clinical findings.

6. Based on the new test results, appropriately update and revise the prioritization of the differential diagnosis, and explain the reasoning that leads to a more probable diagnosis.

7. Develop a treatment plan that considers both scientific evidence (EBM) and the patient's individual factors, and explain the rationale as well as alternative options.

###Response Style and Constraints

1. Please answer in English.
2. Keep the response concise and clear.
3. Base your reasoning solely on the provided information and widely accepted medical knowledge. Do not assume any new facts.

**Agent 3: Therapeutic Evaluation**

SYSTEM Prompt
You are a Japanese nephrologist specialized in clinical reasoning and evidence-based medicine.

Please read the {{#sys.query#}} patient information and follow the instructions.

Think step by step in English, and provide only the final conclusion in the specified format.

**USER Prompt**
##### Instructions

8. How will you monitor the effectiveness and safety of the treatment? What are the specific risks to watch out for, and what are your concrete countermeasures for them?

9. If the patient's condition does not improve as expected or suddenly changes, how would you think and act?

###Response Style and Constraints

1. Please answer in English.
2. Keep the response concise and clear.
3. Base your reasoning only on the provided information and widely accepted medical knowledge. Avoid assuming any new facts.

ASSISTANT Prompt
###Output Format (Strictly Follow)

8. List specific measures for evaluating treatment effectiveness and potential side effects or complications, and explain the monitoring and management plan for them.

9. Explain the thought process and action plan for rapidly reassessing the cause in response to changes in the patient's condition, and for flexibly revising the diagnosis and treatment plan.

###Response Style and Constraints

1. Please answer in English.
2. Keep the response concise and clear.
3. Base your reasoning solely on the provided information and widely accepted medical knowledge. Do not assume any new facts.

**Supplementary Table 1. Prompts and Evaluation Criteria for the Phased Assessment of Clinical Reasoning**

| Question | Prompt to the LLM | Evaluation Criteria |
|---|---|---|
| **Phase I: Before the Consultation** | | |
| Q1 | How would you summarize this patient as a medical problem? Please explain, focusing on the key points. | Accurately and concisely summarizes the patient's problem, incorporating age, gender, timeline, and key symptom characteristics. |
| Q2 | Hearing this chief complaint, what differential diagnoses do you consider? Please list three. What is the most likely? What must not be missed? Please also explain your reasoning. | Provides a broad and prioritized list of differential diagnoses, considering frequency and severity, and explains the rationale. |
| Q3 | To verify the hypotheses you've made, what additional questions would you ask or what physical exams would you perform? What do you expect to clarify with that history-taking or examination? | Proposes focused history or physical exam steps with clear purpose tied to confirming or ruling out differential diagnoses. |
| Q4 | To narrow down the diagnosis, what tests do you plan to order? Please tell me the purpose for ordering those tests and the expected results. | Suggests a logical test plan aligned with diagnostic refinement, considering cost-effectiveness and invasiveness. |

| | | |
|---|---|---|
| **Phase II: After Test Results** | | |
| Q5 | How do you interpret these test results? Please explain the points of particular interest and what they mean in the context of this patient's condition. | Interprets results in clinical context, explaining their medical significance rather than just stating abnormalities. |
| Q6 | Based on all the information, how has your diagnostic thinking changed? What is the most likely diagnosis (Working Diagnosis) at this point? | Revises and reprioritizes differential diagnoses appropriately in light of new information, and explains reasoning. |

| | | |
|---|---|---|
| Q7 | What kind of treatment plan would you create for this patient? Please also explain the rationale for your chosen treatment and other possible options. | Proposes a treatment plan considering both evidence-based medicine (EBM) and patient-specific factors, including justification and alternatives. |

| **Phase III: During Treatment Course** | | |
|---|---|---|
| Q8 | How will you monitor the effectiveness and safety of the treatment? What are the specific risks to watch out for, and what are your concrete countermeasures for them? | Identifies indicators for treatment response and potential side effects or complications, and proposes a monitoring and response plan. |
| Q9 | If the patient's condition does not improve as expected or suddenly changes, how would you think and act? | Describes a process for reassessment of diagnosis or treatment plan, including rapid response and flexible decision-making. |
| The evaluation was conducted using a step-by-step prompting approach to simulate a realistic, temporally structured reasoning process. The clinical reasoning process for each case was divided into three phases as shown in the table. Initially, only the 'Phase I' section of the case was provided to each model to answer the corresponding questions (Q1-Q4); information from Phase II and III was withheld. Upon completion of Phase I, the 'Phase II' section was presented, and the relevant questions (Q5-Q7) were asked, still without disclosing Phase III information. Finally, the 'Phase III' section was provided for the models to answer the last set of questions (Q8-Q9). The outputs from each phase were then evaluated and scored by four nephrology experts on a 3-point scale: 2 for a correct answer, 1 for a reasonable but not optimal answer, and 0 for an incorrect answer. The scores for each model were aggregated to evaluate the differences in total scores across the models. | | |

#### Supplementary Table 2. Responses from Four Large Language Models to Clinical Reasoning Questions Across Nine Cases

| **Case 1: New Onset of Primary Membranous Nephropathy After COVID-- 19 mRNA Vaccination in Affected Sjögren's Syndrome** [2] | |
|---|---|
| **Phase I – Q1: Summarizing medical problems** | |
| DeepSeek-R1 | Medical Problem Summary: 51–55-year-old female with primary Sjögren's syndrome (pSS), hypertension, hyperlipidemia, and hyperuricemia, presenting with **acute-onset (1 week) fluid overload** (8 kg weight gain, generalized edema, oliguria, shortness of breath) and **suspected nephrotic syndrome**, temporally associated with mRNA COVID-19 vaccination (flank pain/edema onset 1 month post-vaccine). Baseline renal function was normal (Cr 0.55 mg/dL, no proteinuria). |
| Gemini 2.5 Pro | Problem Summary: This is a 51–55-year-old female with a strong autoimmune background (primary Sjögren's syndrome and family history) who presents with a subacute onset of acute kidney injury (AKI) and a clinical picture of nephrotic syndrome (generalized edema, oliguria, 8 kg weight gain). This presentation developed one month after receiving an mRNA COVID-19 vaccine and was concurrent with the use of an NSAID (celecoxib). Her baseline kidney function was normal. |
| OpenAI GPT-o3 | Problem summary: 51–55-year-old woman with well-controlled primary Sjögren's syndrome and stable CKD-free baseline (Cr 0.55 mg/dL) presents with 1 week of dyspnoea, oliguria and generalised oedema after 1 month of progressive weight gain (8 kg) that began shortly after a fourth (mRNA) COVID-19 vaccine; has recently taken celecoxib. Key issues: new fluid overload, decreased urine output, suspected acute kidney injury/nephrotic picture in an autoimmune-predisposed patient concurrently exposed to NSAID and ARB. |
| Llama4-Marveric | Summary of the Patient's Medical Problem:<br>The patient is a 51–55-year-old female with a history of primary Sjögren's syndrome (pSS), hypertension, hyperlipidemia, and hyperuricemia, presenting with shortness of breath, generalized edema, and oliguria for 1 week. Her symptoms started about a month after receiving her fourth COVID-19 vaccine dose, with notable weight gain and signs of fluid overload. |
| **Phase I – Q2: Differential diagnoses and rationale** | |
| DeepSeek-R1 | Differential Diagnoses:<br>**1. Most likely: pSS-associated glomerulonephritis** (e.g., membranous nephropathy or lupus-like GN).<br>• Rationale: pSS predisposes to renal involvement; temporal link to vaccine (known trigger for autoimmune flares).<br>**2. ARB/NSAID-induced acute interstitial nephritis (AIN) or acute tubular injury.**<br>• Rationale: Recent celecoxib use + chronic ARB; both cause nephrotoxicity.<br>**3. Must not miss: Vaccine-related minimal change disease (MCD) or podocytopathy.**<br>• Rationale: mRNA vaccines rarely induce nephrotic syndrome; rapid weight gain/edema suggest heavy proteinuria. |
| Gemini 2.5 Pro | Differential Diagnosis<br>**1. Minimal Change Disease (MCD):**<br>• **Reasoning:** This is the most likely diagnosis. The abrupt onset of severe edema and significant weight gain is classic for nephrotic syndrome, of which MCD is a primary cause. Both NSAIDs (celecoxib) and, as per emerging case reports, mRNA COVID-19 vaccines are known potential triggers for MCD. This single diagnosis can explain the entire clinical picture.<br>**2. NSAID-induced Acute Interstitial Nephritis (AIN):**<br>• **Reasoning:** The patient's use of celecoxib is a major red flag. AIN is a common form of drug-induced AKI. The initial flank pain could be a symptom of interstitial inflammation. While classic AIN does not typically cause such profound edema, it can coexist with MCD, |

| | |
|---|---|
| | another known complication of NSAID use.<br>**3. Sjögren's Syndrome-related Nephropathy:**<br>• **Reasoning:** While her Sjögren's was reportedly stable, a systemic flare causing renal involvement is possible. The most common manifestation is tubulointerstitial nephritis, which aligns with AKI and flank pain. Less frequently, it can cause various forms of glomerulonephritis (e.g., membranoproliferative GN), leading to a more mixed nephritic/nephrotic presentation.<br>• **Must Not Miss Diagnosis: Rapidly Progressive Glomerulonephritis (RPGN)**, such as ANCA-associated vasculitis.<br>• **Reasoning:** Any patient presenting with new-onset AKI and signs of glomerular disease must be evaluated for RPGN. Although less likely given the specific triggers, delaying the diagnosis and treatment of conditions like ANCA vasculitis can lead to irreversible kidney failure and multi-organ damage. |
| OpenAI GPT-o3 | Differential diagnosis (most likely / cannot miss)<br>a. Immune-mediated glomerulonephritis / nephrotic syndrome triggered by vaccination or pSS (e.g. membranous GN, minimal-change disease, membranoproliferative GN) – MOST LIKELY given timing, oedema, rapid weight gain, autoimmune background.<br>b. NSAID-related acute interstitial nephritis or combined pre-renal/ATN from NSAID + ARB – alternative common mechanism of AKI with oliguria.<br>c. Decompensated heart failure (hypertensive/cardiometabolic) – MUST NOT MISS as a cause of fluid overload; has hypertension, dyspnoea.<br>d. Rapidly progressive crescentic GN (ANCA-associated, anti-GBM) – MUST NOT MISS because of risk of irreversible renal loss and pulmonary involvement.<br>(Other: obstructive uropathy, renal vein thrombosis, nephrotic-syndrome-related.) |
| Llama4-Marveric | Differential Diagnoses<br>**1. Nephrotic Syndrome:** Given the patient's symptoms of generalized edema, oliguria, and significant weight gain, nephrotic syndrome is a likely diagnosis. The recent history of COVID-19 vaccination and self-medication with celecoxib could be contributing factors.<br>**2. Acute Kidney Injury (AKI):** The patient's oliguria and fluid overload could indicate AKI, potentially caused by various factors including medication (celecoxib), dehydration, or an underlying kidney condition related to her pSS or vaccination.<br>**3. Exacerbation of pSS or new-onset autoimmune nephropathy:** The patient's history of pSS and family history of autoimmune diseases raise the possibility of an autoimmune-related kidney condition, potentially triggered or exacerbated by the COVID-19 vaccination.<br>The most likely diagnosis is **Nephrotic Syndrome**, given the patient's presentation with generalized edema and significant proteinuria is likely (though not directly stated, it is implied by the context). The condition that must not be missed is **Acute Kidney Injury,** as it can rapidly progress and requires prompt intervention. |
| **Phase I – Q3: Necessary physical examinations and reasons** | |
| DeepSeek-R1 | Targeted History/Exam:<br>**1. Questions:**<br>• "Describe flank pain (location, intensity) and celecoxib dosing/frequency?" → Differentiate AIN (pain) vs. GN.<br>• "Any rash, joint pain, or sicca symptom exacerbation?" → Assess pSS activity.<br>• "Urine appearance (foamy/dark)?" → Confirm proteinuria/hematuria.<br>**2. Physical exam:**<br>• Jugular venous distension/crackles → Quantify fluid overload.<br>• Skin/joint inspection → Autoimmune signs (e.g., purpura, synovitis).<br>• Costovertebral angle tenderness → Suggest pyelonephritis/AIN. |
| Gemini 2.5 Pro | Additional History and Physical Examination<br>• **Additional Questions:**<br>**i. Inquire about systemic symptoms:** "Have you experienced any fever, skin rash, new |

| | |
|---|---|
| | or worsening joint pain, coughing up blood, or sinus problems?"<br>• **Clarification:** This screens for signs of AIN (fever, rash), a Sjögren's flare (worsening sicca, arthralgias), or systemic vasculitis (hemoptysis, sinus issues).<br>**ii. Quantify NSAID use:** "Specifically, how many times a week did you take celecoxib, at what dose, and for how many weeks?"<br>• **Clarification:** This helps establish the likelihood of the drug being the primary culprit and the dose-duration relationship.<br>**iii. Assess severity of volume overload:** "How many pillows do you need to sleep on at night? Do you wake up suddenly feeling short of breath?"<br>• **Clarification:** This assesses for orthopnea and paroxysmal nocturnal dyspnea, key indicators of pulmonary edema secondary to severe fluid retention.<br>• **Physical Examination:**<br>**i. Assess volume status:** Carefully check for jugular venous distention (JVD), listen for bibasilar crackles in the lungs, and examine for sacral and lower extremity pitting edema.<br>• **Clarification:** To objectively quantify the degree of fluid overload and guide diuretic therapy.<br>**ii. Perform a thorough skin examination:** Look for any rashes (maculopapular rash of AIN) or palpable purpura (vasculitis).<br>• **Clarification:** The presence of specific skin findings can strongly point towards AIN or a systemic vasculitis.<br>**iii. Check for CVA tenderness:** Palpate the costovertebral angles bilaterally.<br>• **Clarification:** Tenderness would support a diagnosis involving the renal interstitium or parenchyma, such as AIN or pyelonephritis, over a purely glomerular process. |
| OpenAI GPT-o3 | Focused questions / examinations<br>• Urine qualities: frothy urine, cola-colored, nocturia – clarifies proteinuria vs hematuria.<br>• Recent rash, fever, arthralgia, eosinophilia symptoms – supports drug-induced AIN or systemic vasculitis.<br>• Orthopnoea, paroxysmal nocturnal dyspnoea, exertional limits – heart failure probability.<br>• Vaccination/infection timeline, new drugs besides celecoxib – temporal clues.<br>• Dry eyes/mouth worsening, purpura, neuropathy – pSS flare or vasculitis.<br>Physical exam: vitals & BP, JVD, lung crackles, S3, hepatomegaly (CHF); skin rash, edema degree; costovertebral angle tenderness; funduscopy for hypertensive changes. Clarifies volume status, cardiac vs renal vs systemic allergic process. |
| Llama4-Marveric | Additional Questions and Physical Exams<br>To verify the hypotheses, additional questions and physical examinations could include:<br>• Detailed urination history and fluid intake.<br>• Assessment of the patient's volume status and blood pressure.<br>• Examination for signs of kidney disease or failure, such as proteinuria or hematuria.<br>• Review of medication adherence and potential nephrotoxic agents.<br>• Evaluation for signs of autoimmune disease flare or other systemic conditions.<br>These would help clarify the cause of her kidney condition, assess the severity of fluid overload, and guide further management. |
| **Phase I – Q4: Plan for investigations/tests** | |
| DeepSeek-R1 | Diagnostic Testing Plan:<br>**1. Urine protein-to-creatinine ratio (UPCR):**<br>• Purpose: Quantify proteinuria (expected: nephrotic-range >3.5 g/g).<br>**2. Urinalysis with microscopy:**<br>• Purpose: Detect dysmorphic RBCs/casts (GN) vs. WBCs/eosinophils (AIN).<br>**3. Serum creatinine/albumin:**<br>• Purpose: Confirm AKI (expected: Cr elevation from baseline) and hypoalbuminemia.<br>**4. Renal ultrasound:** |

| | |
|---|---|
| | • Purpose: Rule out obstruction, assess kidney size (cost-effective, non-invasive).<br>**5. Autoimmune serologies (anti-SSA/Ro, anti-dsDNA, complements):**<br>• Purpose: Evaluate pSS flare/lupus overlap (expected: anti-SSA+; low C3/C4 in GN).<br>**Rationale:** Prioritizes non-invasive tests to distinguish immune-mediated GN (biopsy-indicated if UPCR+/serologies+) from drug toxicity (supportive care/withdrawal). Avoids assuming vaccine causality without excluding common/urgent etiologies. |
| Gemini 2.5 Pro | Diagnostic Testing Plan<br>**1. Urinalysis with Microscopy and Urine Protein-to-Creatinine Ratio (UPCR):**<br>• **Purpose:** To confirm and characterize the renal pathology. This is the most crucial initial step.<br>• **Expected Results:** I expect a high UPCR (>3.5 mg/mg), confirming nephrotic-range proteinuria. The urine sediment will be critical: a bland sediment would favour MCD, whereas the presence of white blood cells/casts or eosinophils would suggest AIN. Red blood cell casts would point towards a proliferative GN.<br>**2. Blood Tests (Comprehensive Metabolic Panel, Complete Blood Count with differential, Serologies):**<br>• **Purpose:** To confirm AKI, assess for complications, and investigate underlying autoimmune causes.<br>• **Tests:**<br>• **CMP:** To confirm elevated creatinine/BUN and check for electrolyte disturbances (e.g., hyperkalemia, metabolic acidosis) and hypoalbuminemia.<br>• **CBC:** To check for eosinophilia (suggests AIN).<br>• **Serologies:** C3/C4 complement, ANA, Anti-SSA/SSB (to assess Sjögren's activity), and ANCA (to rule out vasculitis).<br>• **Expected Results:** Elevated creatinine, severe hypoalbuminemia, and likely normal complements and negative ANCA if the diagnosis is MCD or NSAID-induced AIN.<br>**3. Renal Ultrasound:**<br>• **Purpose:** To rule out post-renal obstruction as a cause of AKI and to assess kidney size and echogenicity.<br>• **Expected Results:** Normal-to-enlarged kidneys with no evidence of hydronephrosis, which is consistent with an intrinsic medical renal disease.<br>**4. Kidney Biopsy:**<br>• **Purpose:** This is the definitive test to establish a precise diagnosis, as treatment differs significantly between MCD, AIN, and other forms of glomerulonephritis. It is essential for guiding immunosuppressive therapy.<br>• **Expected Results:** Depending on the true cause, the biopsy would show:<br>• **MCD:** Normal light microscopy, negative immunofluorescence, and effacement of podocyte foot processes on electron microscopy.<br>• **AIN:** Interstitial inflammatory infiltrate, often with eosinophils.<br>• **Sjögren's Nephritis:** Interstitial lymphocytic infiltrates (for TIN) or specific glomerular changes. |
| OpenAI GPT-o3 | Planned investigations<br>• Serum chemistries (BUN/Cr, electrolytes) – confirm and stage AKI.<br>• Urinalysis with microscopy & protein/creatinine ratio – detect proteinuria, hematuria, casts to differentiate GN, AIN (WBC casts, eosinophils).<br>• Complete blood count with eosinophils, ESR/CRP – eosinophilia for AIN, inflammation for GN.<br>• Serum albumin & lipids – assess nephrotic syndrome severity.<br>• Autoimmune panel: ANA, anti-SSA/SSB, complements (C3/C4), ANCA, anti-GBM, dsDNA – evaluate possible immune GN or pSS flare.<br>• Viral serologies (HBV, HCV, HIV) – secondary GN causes. |

| | |
|---|---|
| | • Renal ultrasound – exclude obstruction, evaluate size/cortical echogenicity.<br>• Chest X-ray / NT-proBNP, ECG ± echocardiogram – assess heart failure contribution.<br>• If significant proteinuria/active sediment or unclear etiology: kidney biopsy – definitive histology to guide therapy (expected: immune-complex GN or AIN findings).<br><br>Expected findings: heavy proteinuria (>3 g/day), hypoalbuminemia, inactive urine sediment would support nephrotic syndrome (membranous/MCD); active RBC casts or low complements favor immune GN; WBC/eosinophil-rich urine favors AIN; normal imaging rules out obstruction; cardiac studies help exclude primary heart failure. |
| Llama4-Marveric | Tests to Narrow Down the Diagnosis<br>**1. Urinalysis and Protein/Creatinine Ratio:** To confirm the presence and quantify proteinuria, a hallmark of nephrotic syndrome.<br>**2. Serum Albumin and Lipid Profile:** To assess for hypoalbuminemia and hyperlipidemia, characteristic of nephrotic syndrome.<br>**3. Kidney Function Tests (e.g., serum creatinine, eGFR):** To evaluate for AKI or chronic kidney disease.<br>**4. Kidney Ultrasound:** To assess kidney size and structure, and rule out obstruction.<br>**5. Immunological Tests (e.g., ANA, anti-dsDNA):** To investigate the possibility of an autoimmune-related kidney condition or flare of pSS.<br>These tests will help differentiate between nephrotic syndrome, AKI, and other potential diagnoses, guiding further management and treatment. |
| **Phase II – Q5: Interpreting test results** | |
| DeepSeek-R1 | Interpretation of Test Results:<br>• **Serum creatinine (3.2 mg/dL) + hypoalbuminemia (2.5 g/dL) + metabolic acidosis ($CO_2$ 14.2 mmol/L):** Confirms AKI with nephrotic syndrome, explaining fluid overload (edema, pleural effusions, ascites). The non-anion gap acidosis suggests renal tubular dysfunction, possibly from TIN or glomerular disease.<br>• **Urinalysis (4+ proteinuria, 10–20 RBC/hpf) + UPCR 38.14 g/g:** Indicates nephrotic-range proteinuria (exceeding 3.5 g/day) with microscopic hematuria. This, plus RBCs in urine, suggests glomerular inflammation (e.g., RPGN), not solely drug-induced injury.<br>• **Autoimmune markers (ANA 1:80 → 1:320; anti-Ro/SSA 408 → 1672 U/mL):** Rising titers imply active autoimmune flare, likely linked to pSS progression. Anti-Ro/SSA elevation is characteristic of pSS but atypical for primary MN.<br>• **Anti-PLA2R (3 RU/mL) + biopsy findings (granular IgG/C3 deposits, subepithelial deposits):** Supports primary MN diagnosis, yet low anti-PLA2R levels and concurrent TIN/mesangial expansion are unusual for idiopathic MN. This suggests pSS-driven secondary MN or a vaccine-triggered process.<br>• **Resolution pattern (Cr 3.2 → 0.8 mg/dL; UPCR 38.14 → 0.32 g/g):** Rapid improvement after ARB cessation and diuresis argues against classic primary MN (typically slower remission). Rising autoantibodies despite clinical recovery hint at persistent autoimmune activity. |
| Gemini 2.5 Pro | Interpretation of Test Results<br>The patient's laboratory and biopsy results collectively paint a picture of a severe, multi-faceted renal disease.<br>• **Acute Nephrotic Syndrome with AKI:** On admission, the classic triad of nephrotic syndrome is present: severe hypoalbuminemia (2.5 g/dL), massive nephrotic-range proteinuria (14 g/day), and hypercholesterolemia (263 mg/dL). This is coupled with severe AKI (Creatinine 3.2 mg/dL) and its metabolic consequences, including hyperphosphatemia and non-anion gap metabolic acidosis (low $CO_2$ content). The severe hyponatremia (116 mmol/L) is dilutional, a direct result of the profound fluid retention evidenced by crackles, pleural effusion, and ascites.<br>• **Conflicting Autoimmune Markers:** The biopsy's positive staining for PLA2R is a highly specific marker for Primary Membranous Nephropathy (pMN). However, the corresponding serum anti-PLA2R antibody level is low (3 RU/mL). This discordance |

| | |
|---|---|
| | (biopsy positive, serum negative) is seen in a minority of pMN cases and does not exclude the diagnosis. Concurrently, the elevated anti-Ro/SSA antibodies confirm active Sjögren's syndrome, a known cause of secondary MN and tubulointerstitial nephritis (TIN).<br>• **Kidney Biopsy as the Definitive Clue:** The biopsy is the most critical piece of evidence. It confirms **membranous nephropathy (MN)** as the cause of the nephrotic syndrome, characterized by IgG/C3 deposits on the glomerular basement membrane. Crucially, it also reveals **acute tubulointerstitial nephritis (TIN).** This "second hit" of TIN is the likely driver of the rapid deterioration of renal function (the RPGN-like course) and the flank pain, and could be attributed to NSAID use, the Sjögren's syndrome flare, or both. The mesangial deposits further suggest a complex, possibly secondary, immune process.<br><br>In summary, the patient has biopsy-proven MN causing severe nephrotic syndrome, complicated by a severe TIN that has precipitated acute renal failure. The key diagnostic challenge is reconciling the PLA2R-positivity (suggesting Primary MN) with the strong background of autoimmune disease and potential drug toxicity (suggesting Secondary MN and TIN). |
| OpenAI GPT-o3 | Interpretation of the new data<br>• Admission picture – "triple hit" to the kidney<br>− AKI (Cr 3.2 mg/dL, BUN 62 mg/dL) with severe non-anion-gap metabolic acidosis ($CO_2$ 14 mmol/L) and hyper¬phosphataemia (9.8 mg/dL) indicates abrupt fall in GFR.<br>− Massive proteinuria (UPCR 38 g/g ≈ 14 g/24 h) + hypoalbuminaemia (2.5 g/dL) + hyper¬cholesterolaemia fulfill nephrotic syndrome.<br>− Hyponatraemia (116 mmol/L) and volume overload (crackles, effusions, ascites) reflect dilutional effect and low oncotic pressure.<br>− Microscopic haematuria (10–20 RBC/hpf) points to glomerular injury rather than isolated TIN or haemodynamic AKI.<br>− Elevated D-dimer (1.61 mg/L) signals the hyper-coagulable milieu typical of nephrotic syndrome.<br><br>• Biopsy – membranous nephropathy (MN) with TIN<br>− Granular IgG/C3 along GBM and sub-epithelial deposits → MN.<br>− PLA2R-positive staining strongly supports primary MN even though circulating anti-PLA2R titre is low (possible "antigen sink" phenomenon early in disease or during spontaneous immunological remission).<br>− Tubulo-interstitial nephritis fits either pSS-related immune injury or NSAID effect.<br><br>• Evolution over 12 months<br>− Renal function rapidly returns to baseline (Cr 0.6-0.8 mg/dL).<br>− Proteinuria falls from 38 → ~0.3 g/g and albumin normalises, indicating complete clinical remission without specific immuno¬ suppression.<br>− ANA and anti-SSA titres climb, reflecting ongoing systemic autoimmunity of pSS but not translating into recurrent renal disease—important when considering future flare risk.<br><br>Key message: The kidney injury appears to be primary PLA2R-positive MN (triggered or unmasked by vaccine / NSAID) with super-imposed TIN and haemodynamic AKI; removal of offending drugs plus supportive care led to spontaneous remission. |
| Llama4-Marveric | Interpretation of Test Results<br>The patient's laboratory findings indicate severe kidney impairment with nephrotic-range proteinuria (14 g/day), hypoalbuminemia (2.5 g/dL), and elevated serum creatinine (3.2 mg/dL), consistent with nephrotic syndrome and acute kidney injury (AKI). The presence of 4+ proteinuria and hematuria (10-20 RBC/hpf) suggests significant glomerular damage. The kidney biopsy findings of membranous nephropathy (MN) with positive anti-phospholipase A2 receptor (PLA2R) staining support the diagnosis of primary MN. The |

| | |
|---|---|
| | low serum anti-PLA2R antibody level (3 RU/mL) is notable, as it may indicate a lower disease activity or a different disease subtype.<br><br>The patient's autoimmune workup shows a positive ANA titer (1:80) and elevated anti-Ro/SSA autoantibodies, consistent with her history of primary Sjögren's syndrome (pSS). The increasing ANA titer over time (1:320 at 12 months) may indicate an ongoing autoimmune process. |
| **Phase II – Q6: Reconsidering the differential diagnosis** | |
| DeepSeek-R1 | Evolution of Diagnostic Thinking & Working Diagnosis<br>**Initial differentials:**<br>• Drug-induced AKI (ARB/celecoxib) or pSS-related TIN.<br>• Vaccine-associated podocytopathy (e.g., MCD).<br>**Revised diagnosis:**<br>**Secondary membranous nephropathy (MN) in the context of primary Sjögren's syndrome flare,** with concurrent tubulointerstitial nephritis (TIN).<br>**Reasoning:**<br>• Biopsy-confirmed MN + TIN aligns with pSS renal involvement (rarely MN, but reported with anti-Ro/SSA).<br>• Anti-PLA2R positivity typically indicates primary MN, but low titers, mesangial deposits, and TIN favor **autoimmune-mediated secondary MN.**<br>• Temporal vaccine link is plausible, but rising anti-Ro/SSA/ANA suggests pSS exacerbation as the primary driver.<br>• Exclusion: ARB/NSAID toxicity unlikely due to biopsy findings and persistent autoantibodies post-discontinuation. |
| Gemini 2.5 Pro | Revised Diagnostic Thinking and Working Diagnosis:<br>My initial differential diagnosis, which favoured MCD or NSAID-induced AIN, has been significantly refined by the biopsy results. The diagnosis is clearly not MCD. While AIN is present, it coexists with a primary glomerular disease.<br>The diagnosis is now focused on the type of membranous nephropathy. The positive PLA2R staining on the biopsy is a very strong argument for **Primary Membranous Nephropathy (pMN)**, as this antigen is the target in over 70-80% of primary cases. The low serum antibody level does not negate this, as a subset of patients have tissue-positive/serum-negative disease. The Sjögren's syndrome, recent mRNA vaccination, and NSAID use likely acted as potent triggers or exacerbating factors for both the underlying pMN and the development of the superimposed TIN, creating a "perfect storm" that led to this fulminant presentation.<br>**Working Diagnosis: Primary (PLA2R-associated) Membranous Nephropathy with a superimposed severe Acute Tubulointerstitial Nephritis, presenting as an acute nephrotic syndrome and rapidly progressive glomerulonephritis.** The TIN is likely multifactorial, secondary to both NSAID use and the underlying Sjögren's syndrome. |
| OpenAI GPT-o3 | Updated diagnostic thinking<br>Highest-priority (working) diagnosis<br>Primary PLA2R-positive membranous nephropathy with concurrent tubulo-interstitial nephritis in a patient with primary Sjögren's syndrome, presenting as nephrotic syndrome with AKI.<br><br>Why other possibilities moved down<br>• pSS-associated secondary MN – possible, but PLA2R tissue positivity makes primary MN far more likely.<br>• Isolated NSAID-induced AIN – does not explain nephrotic-range proteinuria or PLA2R staining.<br>• RPGN/vasculitis – biopsy lacked crescents or pauci-immune pattern.<br>• Heart failure – contributes to dyspnoea/effusions but is a consequence rather than cause of renal findings. |

| | |
|---|---|
| Llama4-Marveric | Diagnostic Thinking and Working Diagnosis<br>The patient's presentation and laboratory findings support a diagnosis of **primary membranous nephropathy (MN)** with nephrotic syndrome and AKI. The positive PLA2R staining on kidney biopsy is consistent with this diagnosis. The patient's history of pSS and recent COVID-19 vaccination may be relevant to her kidney disease, potentially triggering or exacerbating an underlying autoimmune condition.<br>The differential diagnosis includes other causes of nephrotic syndrome and AKI, such as minimal change disease, focal segmental glomerulosclerosis (FSGS), and secondary MN due to autoimmune diseases or other conditions. However, the kidney biopsy findings and positive PLA2R staining support primary MN as the most likely diagnosis. |
| **Phase II – Q7: Treatment planning** | |
| DeepSeek-R1 | Treatment Plan & Rationale<br>• **Immediate plan:**<br>**i. Supportive care:**<br>•Loop diuretics (furosemide) for hypervolemia (evidence: relieves edema/effusions in nephrotic syndrome).<br>• Statin (e.g., atorvastatin) for hyperlipidemia (LDL 263 mg/dL; EBM: reduces cardiovascular risk in nephrosis).<br>• Non-RAAS antihypertensive (e.g., amlodipine) to avoid ARB-related risks.<br>**ii. Immunosuppression:**<br>• Rituximab (375 mg/m² weekly × 4 doses) as first-line for MN with autoimmune features (EBM: induces remission in PLA2R+ MN and pSS-related glomerulopathy).<br>• Low-dose corticosteroids (prednisone 0.5 mg/kg/day) during rituximab induction to suppress acute inflammation.<br>•**Rationale:**<br>• Rituximab targets B-cells driving anti-PLA2R/pSS autoimmunity, with lower toxicity than cyclophosphamide.<br>• Avoided cyclophosphamide due to favorable rituximab efficacy in MN and pSS.<br>• Continued hydroxychloroquine for pSS sicca symptoms.<br>• **Alternatives:**<br>• Cyclophosphamide if rituximab contraindicated (e.g., severe infections) but higher toxicity risk.<br>• ACEi/ARB reintroduction if proteinuria persists post-remission (EBM: antiproteinuric in MN).<br>• **Monitoring:**<br>UPCR, anti-Ro/SSA, and creatinine q3–6 months to assess remission.<br>**Conclusion:** Secondary MN in pSS flare is the working diagnosis; rituximab-based immunosuppression is prioritized for renal and autoimmune activity.<br>Please enter the treatment course. |
| Gemini 2.5 Pro | Treatment Plan<br>The treatment must be aggressive and two-pronged, addressing both the life-threatening complications and the underlying immunological disease.<br>**1. Immediate Supportive Care:**<br>• **Aggressive Diuresis:** High-dose intravenous loop diuretics (e.g., furosemide infusion) are critical to manage the severe volume overload (pulmonary edema, ascites). The addition of a thiazide-like diuretic (e.g., metolazone) may be necessary to overcome diuretic resistance.<br>• **Discontinuation of Offending Agents:** The NSAID (celecoxib) must be permanently discontinued as a likely contributor to the TIN. The ARB was correctly stopped in the setting of severe AKI.<br>• **Thromboembolism Prophylaxis:** Due to the severe hypoalbuminemia (<2.5 g/dL), the patient is in a hypercoagulable state. Prophylactic anticoagulation (e.g., with low- |

| | |
|---|---|
| | molecular-weight heparin) is strongly indicated to prevent venous thromboembolism, provided there are no contraindications.<br>**2. Specific Immunosuppressive Therapy:**<br>Given the high-risk features (rapidly declining renal function and severe nephrotic syndrome), immediate immunosuppression is warranted.<br>• **Recommended Regimen:** A combination of **Rituximab and corticosteroids.**<br>• **Rationale:**<br>• **Rituximab:** As a B-cell depleting agent, it directly targets the pathogenesis of PLA2R-associated MN. It is a first-line, evidence-based therapy with proven efficacy and a better long-term safety profile than cytotoxic agents.<br>• **Corticosteroids:** A course of high-dose corticosteroids (e.g., intravenous methylprednisolone pulses followed by an oral prednisone taper) is essential. Steroids will provide a rapid, potent anti-inflammatory effect to treat the severe, T-cell mediated TIN, which is the primary driver of the acute renal failure. This provides a bridge to the slower-acting effects of Rituximab on the MN component.<br>• **Alternative Options:**<br>• **Cyclic Cyclophosphamide and Corticosteroids (e.g., Modified Ponticelli Regimen):** This is a highly effective, classic regimen for high-risk MN. However, it carries significant short- and long-term toxicities (infection, malignancy, infertility) and would be considered a second choice to Rituximab in this patient.<br>• **Calcineurin Inhibitors (e.g., Tacrolimus):** While effective at reducing proteinuria, they are nephrotoxic and their effect is often dependency-related. They are not ideal as initial therapy for a patient with such severe AKI.<br>Please enter the treatment course. |
| OpenAI GPT-o3 | Treatment plan and rationale<br>A. Supportive / maintenance therapy (continue long term)<br>1. Blood-pressure & proteinuria control<br>− Reinstate low-dose ACEi/ARB (titrate while monitoring $K^+$/Cr) ± add SGLT2 inhibitor; target BP < 130/80 mmHg and UPCR < 0.3 g/g.<br>2. Volume management<br>− Loop diuretic PRN, ≤2 g/day sodium diet.<br>3. Thrombosis prophylaxis<br>− Full anticoagulation was indicated while albumin < 2.5 g/dL and UPCR > 10 g/g; may now stop but keep low threshold to restart if relapse.<br>4. Lipid lowering – continue statin to keep LDL < 70 mg/dL.<br>5. Vaccination / drug avoidance – avoid NSAIDs and monitor after future vaccines.<br><br>B. Immuno-modulatory strategy<br>• At present no immuno-suppressive therapy is needed because she achieved complete remission (< 0.3 g/g, stable GFR).<br>• If relapse (proteinuria > 4 g/day for > 6 months or GFR decline ≥30 %):<br>1. First-line: Rituximab 1 g ×2 (day 1 & 15) or 375 mg/m² weekly ×4 (strong evidence, fewer adverse events).<br>2. Alternative: Modified Ponticelli regimen (steroid + cyclophosphamide) or calcineurin inhibitor ± low-dose steroid.<br>3. Monitor serum anti-PLA2R titre to guide need for retreatment.<br><br>C. Management of primary Sjögren's syndrome<br>• Continue hydroxychloroquine.<br>• If extraglandular pSS activity (e.g., rising anti-SSA with systemic symptoms) becomes clinically relevant, consider low-dose oral prednisolone or rituximab (would also cover MN relapse). |

| | |
|---|---|
| | D. Follow-up schedule<br>• Clinic visit every 1–3 months for 1 year, then twice yearly: BP, weight, serum Cr/eGFR, UPCR, albumin, PLA2R titre, pSS activity.<br>• Early re-biopsy only if unexplained renal decline.<br>This plan emphasises evidence-based supportive care, watchful waiting given the spontaneous remission, and clearly defined triggers for escalation to immuno¬suppressive therapy. |
| Llama4-Marveric | Treatment Plan<br>The treatment plan for this patient should focus on managing nephrotic syndrome, AKI, and the underlying primary MN.<br>**1. Immunosuppressive therapy:** Consider initiating immunosuppressive therapy, such as corticosteroids and/or calcineurin inhibitors (e.g., cyclosporine or tacrolimus), to reduce proteinuria and slow disease progression.<br>**2. Supportive care:** Continue diuretic therapy (furosemide) to manage hypervolemia and edema. Monitor and adjust as needed to avoid dehydration and electrolyte imbalances.<br>**3. Renin-angiotensin-aldosterone system (RAAS) inhibitors:** Although the patient was initially on an ARB, which was discontinued due to worsening renal impairment, RAAS inhibitors may be reconsidered in the future if kidney function stabilizes.<br>**4. Monitoring:** Regularly monitor kidney function, proteinuria, and serum albumin levels to assess disease activity and response to treatment.<br>**5. Management of pSS:** Continue managing the patient's pSS with hydroxychloroquine and lubricating eye drops, and monitor for signs of disease flare or other complications.<br>Alternative options may include other immunosuppressive regimens, such as rituximab or cyclophosphamide, depending on the patient's response to initial therapy and disease severity. |
| **Phase III – Q8: Evaluation of treatment** | |
| DeepSeek-R1 | Monitoring Effectiveness and Safety<br>**Effectiveness monitoring:**<br>• **Proteinuria:** Monthly UPCR (target: <0.15 g/g) to detect relapse (e.g., microalbuminuria recurrence).<br>• **Renal function:** Biweekly serum creatinine during active therapy; quarterly eGFR after stabilization.<br>• **Immunological activity:** Quarterly anti-PLA2R (target: <2 RU/mL) and anti-Ro/SSA levels to assess autoimmune control.<br>• **Volume status:** Daily weight tracking and weekly edema assessment.<br>**Key risks and countermeasures:**<br>**1. Immunosuppression-related infections (e.g., sepsis, CMV):**<br>• Monitor: CBC biweekly (neutrophil count >1,500/μL), CRP monthly.<br>• Counter: Prophylactic TMP-SMX for Pneumocystis; withhold MMF if leukopenia.<br>**2. Calcineurin inhibitor (CNI) toxicity (nephrotoxicity, hypertension):**<br>• Monitor: Biweekly CNI trough levels (tacrolimus target 5–8 ng/mL); monthly BP.<br>• Counter: Dose reduction for elevated CNI levels; switch to non-dihydropyridine CCB (e.g., diltiazem) for hypertension.<br>**3. Steroid complications (osteoporosis, hyperglycemia):**<br>• Monitor: DEXA scan annually; fasting glucose monthly.<br>• Counter: Calcium/vitamin D supplementation; initiate SGLT2 inhibitor if HbA1c >6.5%.<br>**4. Thrombosis (nephrotic syndrome-related hypercoagulability):**<br>• Monitor: D-dimer if edema worsens; Doppler US for limb swelling.<br>• Counter: Prophylactic enoxaparin if albumin <2.5 g/dL. |
| Gemini 2.5 Pro | **Treatment Effectiveness and Safety Monitoring**<br>**Effectiveness Monitoring**<br>The goal is to maintain remission while minimizing drug toxicity.<br>• **Renal Function & Proteinuria:** Monitor serum creatinine, eGFR, and urine albumin- |

<table>
<tr>
<td></td>
<td>
to-creatinine ratio (UACR) every 1-3 months. A significant, sustained increase in UACR signals a relapse and requires intervention.<br>
• Nephrotic Syndrome Markers: Monitor serum albumin and cholesterol levels quarterly to ensure they remain within the normal range.<br>
• Immunological Activity: While the baseline serum anti-PLA2R was low, a repeat measurement could be considered if a significant relapse occurs, as a rise may confirm an immunological flare of the primary membranous nephropathy.<br>
Safety Monitoring and Management Plan<br>
Given the triple immunosuppressive therapy, monitoring for side effects is critical.
<table>
<tr><th>Drug/Condition</th><th>Specific Risk(s)</th><th>Monitoring and Management Plan</th></tr>
<tr><td>Calcineurin Inhibitor (CNI)</td><td>Nephrotoxicity, hypertension, hyperkalemia, hyperglycemia, hypomagnesemia.</td><td>Monitor CNI trough levels regularly to ensure they are within the therapeutic range. Check serum creatinine, potassium, and magnesium concurrently. If creatinine rises with a high CNI level, reduce the dose.</td></tr>
<tr><td>Mycophenolate Mofetil (MMF)</td><td>Leukopenia (low white blood cells), gastrointestinal side effects (diarrhea), increased infection risk.</td><td>Monitor Complete Blood Count (CBC) monthly for the first few months, then every 2-3 months. If leukopenia develops, the dose must be reduced or held.</td></tr>
<tr><td>Corticosteroids (Prednisone)</td><td>Osteoporosis, hyperglycemia/diabetes, weight gain, cataracts, hypertension.</td><td>Recommend calcium and vitamin D supplementation. Schedule a baseline and follow-up bone density (DEXA) scan. Monitor blood pressure and blood glucose/HbA1c regularly. Annual ophthalmologic exams are advised.</td></tr>
<tr><td>General Immunosuppression</td><td>Opportunistic infections, particularly Pneumocystis jirovecii pneumonia (PJP).</td><td>Prescribe PJP prophylaxis (e.g., with trimethoprim-sulfamethoxazole) for the duration of intensive immunosuppression. Educate the patient to report any signs of infection (fever, cough) immediately.</td></tr>
</table>
</td>
</tr>
<tr>
<td>OpenAI GPT-o3</td>
<td>
Monitoring plan for effectiveness and safety<br>
Effectiveness<br>
• Clinical: body weight, blood pressure, edema score, urine output – every visit (q2–4 wk).<br>
• Laboratory: serum creatinine/eGFR, BUN, serum albumin, spot UPCR/UACR – q2 wk ×3 mo, then monthly; target UPCR < 0.3 g/g and stable eGFR.<br>
• Immunologic: serum anti-PLA2R Ab and complement levels – at baseline, 3 mo, 6 mo, then q6 mo; falling or negative titre predicts sustained remission.<br>
• Imaging (if indicated): renal Doppler for suspected renal-vein thrombosis; chest X-ray for fluid status.<br>
<br>
Safety / complications & counter-measures
</td>
</tr>
</table>

| | |
|---|---|
| | • Steroid toxicity (hyperglycaemia, hypertension, osteoporosis, infection): fasting glucose/BP each visit; DEXA at 6 mo; Ca/Vit-D + bisphosphonate; taper to ≤5 mg/d as soon as possible.<br>• CNI nephro- & neuro-toxicity, hypertension: trough level (tacrolimus 5–7 ng/mL or cyclosporine 80–120 ng/mL) and serum Cr q2 wk ×3 mo then monthly; adjust dose or switch to rituximab if rising Cr or tremor.<br>• MMF-related cytopenia, GI upset, teratogenicity: CBC and liver panel q2 wk ×2 mo, then q1–2 mo; dose ↓ or hold if WBC < 3 $\times 10^9$/L; contraception counselling.<br>• Infection (opportunistic, COVID-19, PJP, zoster, BK): temp review each visit; CMV PCR and urinalysis for BK q3 mo; PJP prophylaxis (TMP-SMX), zoster vaccination, COVID-19 boosters after immunosuppression tapered.<br>• Thrombo-embolism (nephrotic syndrome): continue low-dose anticoagulation while albumin < 2.5 g/dL or UPCR > 8 g/g; monitor D-dimer and leg Doppler if symptomatic.<br>• Metabolic: lipid profile q3 mo; statin or ezetimibe as needed.<br>• Malignancy (skin, lymphoma): annual skin exam; age-appropriate cancer screening. |
| Llama4-Marveric | Monitoring Treatment Effectiveness and Safety<br>To monitor the effectiveness and safety of the treatment, the following measures will be taken:<br>**1. Regular laboratory tests:** Monitor kidney function (serum creatinine, eGFR), proteinuria (UPCR or UACR), and serum albumin levels to assess disease activity and response to treatment.<br>**2. Imaging studies:** Repeat imaging studies (e.g., kidney ultrasound) as needed to monitor for changes in kidney size or structure.<br>**3. Medication side effects:** Monitor for potential side effects of immunosuppressive therapy, such as infections, hypertension, and hyperlipidemia.<br>**4. Immunosuppressive drug levels:** Monitor levels of calcineurin inhibitors and mycophenolate mofetil to ensure adequate dosing and minimize toxicity.<br><br>Specific risks to watch out for include:<br>• Infections due to immunosuppression<br>• Nephrotoxicity from calcineurin inhibitors<br>• Gastrointestinal side effects from mycophenolate mofetil<br>• Relapse of proteinuria or kidney disease progression<br><br>Countermeasures will include:<br>• Prophylactic measures against infections (e.g., Pneumocystis jirovecii pneumonia prophylaxis)<br>• Regular monitoring of kidney function and immunosuppressive drug levels<br>• Adjusting immunosuppressive therapy as needed to minimize side effects |
| **Phase III – Q9: Formulating plans for worsening conditions** | |
| DeepSeek-R1 | Response to Unexpected Changes<br>**Thought process:**<br>• **Non-response/worsening** (e.g., rising creatinine or UPCR):<br>• Re-evaluate for misdiagnosis (e.g., lupus nephritis vs. MN) or secondary triggers (infection, non-adherence).<br>• Prioritize drug toxicity (CNI/MMF) or autoimmune flare (rising anti-Ro/SSA).<br>• **Acute deterioration** (e.g., anuria, hemoptysis):<br>• Suspect pulmonary embolism (nephrotic hypercoagulability) or RPGN relapse.<br>**Action plan:**<br>**1. Immediate diagnostics:**<br>• Urgent renal ultrasound (obstruction), ANCA/anti-GBM serology (RPGN), CT pulmonary angiogram (PE). |

| | |
|---|---|
| | • Repeat kidney biopsy if RPGN suspected.<br>**2. Treatment revision:**<br>• **Infection-driven flare:** Reduce immunosuppression; culture-directed antibiotics.<br>• **Refractory MN:** Switch CNI to rituximab (375 mg/m² weekly × 4 doses).<br>• **Thrombosis:** Therapeutic anticoagulation (e.g., apixaban).<br>**3. Rescue therapy:**<br>• Plasma exchange if anti-GBM positive; IVIG for infection-concurrent flare.<br>**Rationale:** Focuses on rapid exclusion of life-threatening causes (e.g., PE, crescentic GN) while maintaining remission in relapsing MN. |
| Gemini 2.5 Pro | Management of Worsening or Unexpected Changes<br>If the patient's condition deteriorates (e.g., rising creatinine, worsening proteinuria, or new systemic symptoms), my approach is a rapid, systematic re-evaluation.<br>**1. Thought Process: Differentiating the Cause**<br>My immediate priority is to determine if the change is due to:<br>• **A. Disease Relapse:** A true flare of the primary membranous nephropathy, potentially triggered by tapering immunosuppression or an intercurrent illness.<br>• **B. Treatment Complication:** This is a high-probability concern. The primary suspect would be calcineurin inhibitor (CNI) nephrotoxicity, which can cause a rise in creatinine. A secondary infection due to the potent immunosuppression is also a major risk.<br><br>• **C. A New, Unrelated Problem:** Such as dehydration, urinary tract infection, or another acute illness causing a transient decline in kidney function.<br>**2. Action Plan: Rapid Reassessment and Flexible Revision**<br>• **Step 1: Immediate Clinical and Laboratory Assessment:**<br>• Perform a focused history and physical exam to assess for medication adherence, signs of infection, and volume status.<br>• Order urgent laboratory tests:<br>• **Serum:** Comprehensive metabolic panel (to check creatinine, potassium, glucose) and a complete blood count (to check for leukopenia).<br>• **Urine:** Urine analysis with protein-to-creatinine ratio to quantify the change.<br>• **Drug Level:** A stat CNI trough level is essential to rule in or out toxicity.<br>• **Step 2: Interpret Results and Revise the Plan:**<br>• **If CNI level is high with rising creatinine:** This strongly suggests CNI nephrotoxicity. The immediate action is to reduce the CNI dose and re-evaluate renal function closely.<br>• **If CNI level is therapeutic but proteinuria is increasing:** This points towards a true disease relapse. The plan would be to intensify the immunosuppressive regimen, perhaps with a short course of steroids or by considering alternative therapies like rituximab.<br>• **If signs of infection are present (e.g., fever, leukocytosis/leukopenia):** Initiate a full infection workup (cultures, imaging) and start empiric antibiotics. The immunosuppressive regimen may need to be temporarily reduced, especially MMF, to allow the immune system to fight the infection.<br>• **If all the above are negative:** Re-evaluate for other causes and consider a repeat kidney biopsy if the diagnosis becomes unclear or the condition is refractory to treatment adjustments. |
| OpenAI GPT-o3 | Approach if the patient fails to improve or acutely deteriorates<br>Step 1 – Re-assessment<br>• Verify adherence, drug levels, recent infections or nephrotoxins (NSAID, contrast).<br>• Repeat urgent labs (BMP, CBC, UPCR, anti-PLA2R), urinalysis microscopy, and renal ultrasound to look for obstruction or thrombosis.<br>• Screen for infection/sepsis (blood/urine cultures, chest imaging). |

| | |
|---|---|
| | Step 2 – Differential refinement<br>• Distinguish relapse of MN (proteinuria ↑, anti-PLA2R rise, stable Cr), CNI nephrotoxicity (Cr ↑, low proteinuria), acute interstitial nephritis/infection, or new crescentic GN / overlap vasculitis.<br><br>Step 3 – Targeted actions<br>• Hold or reduce suspected offending agent (e.g., CNI) and provide supportive care.<br>• If MN relapse confirmed: consider rituximab 1 g × 2 or cyclophosphamide-based regimen.<br>• If rapidly falling GFR or new active urine sediment: renal biopsy to rule out superimposed pathology; start high-dose steroids ± plasma exchange for RPGN while awaiting histology.<br>• Treat infections promptly; adjust immunosuppression until controlled.<br>• Escalate to haemodialysis for refractory fluid overload or uraemia.<br><br>Step 4 – Ongoing review<br>• Multidisciplinary discussion (nephrology, rheumatology, infectious disease).<br>• Re-evaluate goals of care and tailor therapy based on updated findings. |
| Llama4-Marveric | Managing Unexpected Changes or Lack of Improvement<br>If the patient's condition does not improve as expected or suddenly changes, the following steps will be taken:<br>**1. Reassess the diagnosis:** Review the patient's clinical presentation, laboratory findings, and kidney biopsy results to ensure the diagnosis of primary MN is correct.<br>**2. Investigate potential causes of treatment failure:** Consider factors such as non-adherence to treatment, inadequate immunosuppression, or underlying secondary causes of MN.<br>**3. Adjust treatment:** Modify immunosuppressive therapy as needed, considering alternative regimens or intensifying current treatment.<br>**4. Investigate for complications:** Evaluate for potential complications such as thrombosis, infections, or other adverse effects of treatment.<br>By closely monitoring the patient's response to treatment and adjusting the management plan as needed, we can optimize outcomes and minimize potential complications. |

| **Case 2: A Case of IgA Nephropathy With Membranoproliferative Glomerulonephritis-- Like Features [3]** | |
|---|---|
| **Phase I – Q1: Summarizing medical problems** | |
| DeepSeek-R1 | 71–75-year-old male with chronic rheumatoid arthritis (RA) and type 2 diabetes, presenting with acute-onset nephrotic-range proteinuria (5.7 g/day), hematuria, and hypoalbuminemia. Key characteristics:<br>**Time course:** Asymptomatic urinary abnormalities detected recently without prior history, progressing to nephrotic syndrome despite well-controlled diabetes (HbA1c 6.5–7.0%) and RA in remission.<br>**Context:** On methotrexate and tofacitinib; no infection or family history of kidney disease. |
| Gemini 2.5 Pro | This is a 71–75-year-old man with a long-standing history of well-controlled rheumatoid arthritis and type 2 diabetes, presenting with the subacute onset of nephrotic syndrome, characterized by nephrotic-range proteinuria (5.7 g/day) and hematuria. |
| OpenAI GPT-o3 | 71–75-year-old man with 19-year type 2 diabetes (HbA1c 6.5–7 %), long-standing rheumatoid arthritis in remission on methotrexate + tofacitinib, now presents with new-onset nephrotic-range proteinuria (5.7 g/day), microscopic hematuria, and hypoalbuminemia → nephrotic syndrome of unclear etiology. |
| Llama4-Marveric | The patient is a 71–75-year-old man with an 18-year history of rheumatoid arthritis (RA) in remission and a 19-year history of type 2 diabetes managed with insulin. He recently developed nephrotic-range proteinuria (5.7 g/day) and hematuria without symptoms or infection, leading to a diagnosis of nephrotic syndrome. |

| | |
|---|---|
| **Phase I – Q2: Differential diagnoses and rationale** | |
| DeepSeek-R1 | Differential Diagnoses<br>**Prioritized list:**<br>**1. Amyloidosis (AA type)** – Most likely. Chronic RA inflammation can trigger AA amyloid deposition in kidneys, presenting with nephrotic syndrome.<br>**2. Membranous nephropathy** – Associated with RA autoimmunity or malignancy; classic for nephrotic syndrome.<br>**3. Diabetic nephropathy** – Long-standing diabetes but atypical due to abrupt onset, hematuria (rare in pure diabetic nephropathy), and good glycemic control.<br>**Must not miss: Malignancy-associated glomerulonephritis** (e.g., membranous pattern from solid tumors). RA and immunosuppression increase malignancy risk, and hematuria suggests non-diabetic pathology. |
| Gemini 2.5 Pro | Differential Diagnosis<br>I consider the following three differential diagnoses:<br>**1. AA Amyloidosis:** A well-known complication of chronic inflammatory conditions like rheumatoid arthritis. The 18-year disease duration is a significant risk factor, even with current clinical remission.<br>**2. Membranous Nephropathy (MN):** The most common cause of primary nephrotic syndrome in older adults. It can also be secondary to autoimmune diseases like RA. The presentation of nephrotic syndrome with hematuria is classic for MN.<br>**3. Diabetic Nephropathy (DN):** The patient's 19-year history of diabetes makes this a possibility. However, the relatively sudden onset and presence of hematuria are atypical for classic DN, suggesting a non-diabetic or superimposed glomerular disease.<br><br>**Most Likely Diagnosis:** Given the very long history of rheumatoid arthritis, **AA Amyloidosis** is the most likely diagnosis.<br>**Must-Not-Miss Diagnosis: ANCA-associated vasculitis (AAV).** This condition can present with hematuria and proteinuria in this age group. Delaying diagnosis can lead to rapidly progressive glomerulonephritis and irreversible kidney failure. |
| OpenAI GPT-o3 | Differential diagnosis (top 3)<br>**a. Membranous nephropathy** (primary or RA/drug-related): common cause of nephrotic syndrome in the elderly; hematuria can accompany; RA and JAK inhibitors are recognized secondary associations. → most likely.<br>**b. Diabetic nephropathy:** 19-year diabetes makes it frequent; but hematuria is less typical and proteinuria appeared abruptly after years of normal urinalyses.<br>**c. AA amyloidosis from RA:** long-standing inflammatory disease; causes nephrotic syndrome with possible hematuria; may progress despite clinical remission. → must not be missed because of irreversible decline and systemic involvement. |
| Llama4-Marveric | Differential Diagnoses<br>**1. Diabetic Nephropathy:** Given his long-standing diabetes, this is a likely cause. However, the presence of hematuria is less typical.<br>**2. Amyloidosis (AA or AL type):** RA can lead to AA amyloidosis, and amyloidosis is a known cause of nephrotic syndrome.<br>**3. Membranous Nephropathy:** This can be secondary to RA or other autoimmune diseases and is a common cause of nephrotic syndrome in adults.<br><br>The most likely diagnosis is **Diabetic Nephropathy** or **Amyloidosis** given his history. **Rapidly Progressive Glomerulonephritis** must not be missed, although it's less likely given the absence of acute kidney injury or severe hypertension. |
| **Phase I – Q3: Necessary physical examinations and reasons** | |
| DeepSeek-R1 | Additional History and Physical Exam<br>**Targeted questions:**<br>• **Systemic symptoms:** Weight loss/fever (malignancy/vasculitis), edema severity (nephrotic confirmation), joint pain (RA flare vs. amyloid arthropathy).<br>• **Drug review:** Timing of tofacitinib/methotrexate relative to proteinuria (drug-induced injury). |

| | |
|---|---|
| | • **Amyloid-specific:** Autonomic symptoms (e.g., orthostatic hypotension), carpal tunnel syndrome.<br>**Physical exam:**<br>• **Edema assessment** (confirm nephrotic state).<br>• **Palpate lymph nodes/hepatosplenomegaly** (malignancy).<br>• **Joint inspection for RA activity or amyloid deposits** (e.g., shoulder pad sign).<br>• **Skin/mucosa:** Purpura (vasculitis), macroglossia (amyloid).<br>**Purpose:** Differentiate RA complications (amyloidosis) from drug effects or malignancy. |
| Gemini 2.5 Pro | Additional History and Physical Examination<br>To further evaluate the differential diagnoses, I would perform the following:<br>**History Taking:**<br>• **Review of Systems for Amyloidosis:** Inquire about symptoms of autonomic dysfunction (orthostatic dizziness), gastrointestinal issues (diarrhea, early satiety), cardiac symptoms (dyspnea on exertion), or easy bruising. This helps assess for systemic amyloid deposition.<br>• **Review of Systems for Vasculitis:** Ask about constitutional symptoms (fever, weight loss), upper respiratory issues (sinusitis, epistaxis), pulmonary symptoms (cough, hemoptysis), or new skin rashes/purpura. This screens for a systemic vasculitis like AAV.<br>• **RA and Medication History:** Confirm the stability of his RA (any recent joint swelling or stiffness) and ask about any use of over-the-counter medications, particularly NSAIDs, which can cause other forms of kidney disease.<br>**Physical Examination:**<br>• **Vital Signs:** Check for orthostatic hypotension (suggests autonomic neuropathy in amyloidosis or diabetes).<br>• **HEENT/Skin:** Examine for macroglossia (amyloidosis), palpable purpura, or other skin lesions (vasculitis).<br>• **Cardiopulmonary:** Assess for signs of heart failure (elevated JVP, S3 gallop, edema), which can be seen in cardiac amyloidosis.<br>• **Abdomen:** Palpate for hepatosplenomegaly (amyloidosis). |
| OpenAI GPT-o3 | Focused history & physical exam<br>• Course: exact onset and progression of proteinuria/edema to gauge abrupt vs gradual → helps distinguish DN (slow) from MN (subacute) or MCD (rapid).<br>• RA activity, flares, CRP/ESR trends → persistent inflammation supports AA amyloidosis.<br>• Drug history: NSAIDs, biologics, herbal meds → points to secondary MN or minimal change.<br>• B symptoms, weight loss, neuropathy, orthostatic hypotension → systemic amyloidosis.<br>• Diabetic complications: retinopathy, neuropathy → presence supports DN.<br>• Family kidney disease → hereditary amyloidosis.<br>• Physical: BP (nephrotic HTN), edema pattern, retinal exam (DN), joint deformities, carpal tunnel, macroglossia, purpura, cardiomegaly/low-voltage ECG signs → amyloidosis clues. |
| Llama4-Marveric | Additional Questions and Physical Exams<br>To verify these hypotheses, I would:<br>• Ask about any recent changes in his RA treatment or disease activity.<br>• Inquire about symptoms suggestive of amyloidosis (e.g., weight loss, diarrhea).<br>• Perform a physical exam to check for signs of fluid overload (e.g., edema) and assess for peripheral neuropathy or other complications of diabetes.<br>• Examine for signs of active RA or other autoimmune diseases. |
| **Phase I – Q4: Plan for investigations/tests** | |
| DeepSeek-R1 | Diagnostic Testing Plan<br>**Tests and rationale:** |

| | |
|---|---|
| | **1. Renal biopsy** (definitive):<br>• Purpose: Confirm histology (e.g., amyloid deposits, immune complexes).<br>• Expected: Congo red+ fibrils (amyloid), IgG spikes (membranous), or diabetic changes.<br>**2. Serum/urine protein electrophoresis** (SPEP/UPEP):<br>• Purpose: Detect monoclonal protein (malignancy/AL amyloid) vs. polyclonal increase (AA amyloid).<br>• Expected: Polyclonal spike in AA amyloid; monoclonal suggests malignancy.<br>**3. ANCA/anti-PLA2R antibodies:**<br>• Purpose: Rule out vasculitis (ANCA+) or primary membranous (anti-PLA2R+).<br>• Expected: Negative (more likely RA-related than primary autoimmune).<br>**4. Ultrasound (renal):**<br>• Purpose: Assess kidney size (amyloid: enlarged; diabetic: variable).<br>• Expected: Normal/enlarged in amyloid.<br>**Cost-effectiveness:** Non-invasive serology/imaging first, then biopsy. Avoid redundant tests (e.g., HbA1c recent). |
| Gemini 2.5 Pro | Diagnostic Tests<br>To narrow the diagnosis, I would order the following tests before proceeding to the biopsy:<br>• **Urinalysis with Microscopy:** To confirm hematuria and search for dysmorphic red blood cells or RBC casts, the presence of which would increase suspicion for a proliferative glomerulonephritis like AAV.<br>• **Serological Studies:**<br>• **ANCA (MPO and PR3 antibodies):** To rule out the must-not-miss diagnosis of ANCA-associated vasculitis.<br>• **Anti-PLA2R Antibody:** A positive result is highly specific for primary Membranous Nephropathy and could potentially obviate the need for a biopsy in some contexts.<br>• **Serum Protein Electrophoresis (SPEP) and Immunofixation:** To screen for a monoclonal gammopathy, which is essential for ruling out AL amyloidosis.<br>• **Renal Ultrasound:** To assess kidney size (often enlarged in amyloidosis) and structure, and to rule out any obstructive causes.<br><br>**Expected Results & Purpose:** A negative ANCA and anti-PLA2R would lower the probability of AAV and primary MN, respectively, thereby increasing the relative likelihood of AA amyloidosis. Positive SPEP would shift focus to a plasma cell dyscrasia. Ultimately, these tests are crucial for interpreting the findings of the **renal biopsy**, which remains the gold standard for definitive diagnosis. The biopsy with Congo red staining is expected to confirm or rule out amyloidosis and specify the type. |
| OpenAI GPT-o3 | Planned investigations<br>• Basic chemistry (Cr, eGFR, albumin, lipids), HbA1c → quantify syndrome & baseline.<br>• Urinalysis with microscopy & spot protein/Cr; 24 h protein → confirm nephrotic range, look for diabetic waxy casts vs dysmorphic RBCs.<br>• Serum complement (C3/C4), ANA, ANCA, anti-PLA2R Ab → evaluate immune GN; positive PLA2R favors primary MN.<br>• Hepatitis B/C, HIV serology → secondary MN work-up.<br>• Serum & urine electrophoresis + immunofixation, κ/λ free light chains → exclude AL amyloidosis/monoclonal processes.<br>• Serum amyloid A level → supports AA amyloidosis.<br>• Renal ultrasound → kidney size (small in advanced DN, normal/enlarged in amyloid).<br>• Definitive: Native kidney biopsy (already arranged) with light microscopy, IF, EM, Congo-red staining → distinguish DN nodular sclerosis vs membranous pattern vs amyloid deposition, thereby confirming diagnosis and guiding therapy. |

| | |
|---|---|
| Llama4-Marveric | Tests to Narrow Down the Diagnosis<br>**1. Renal Biopsy:** Already performed, the results will be crucial.<br>**2. Serum and Urine Protein Electrophoresis:** To check for paraproteins indicative of AL amyloidosis or other plasma cell dyscrasias.<br>**3. Amyloid Deposits on Biopsy with Specific Staining:** To diagnose amyloidosis.<br>**4. Kidney Function Tests (eGFR, Creatinine):** To assess the severity of kidney impairment.<br>**5. Imaging (e.g., Ultrasound):** To rule out obstructive causes or assess kidney size and structure.<br>These tests will help differentiate between diabetic nephropathy, amyloidosis, and other causes of nephrotic syndrome. |
| **Phase II – Q5: Interpreting test results** | |
| DeepSeek-R1 | Interpretation of Test Results<br>• **Elevated serum IgA (547 mg/dL) r**eflects aberrant mucosal immune activation, central to IgA nephropathy (IgAN) pathogenesis. In this RA patient, chronic inflammation may exacerbate IgA overproduction.<br>• **Nephrotic-range proteinuria (5.25 g/gCr) with hypoalbuminemia (2.9 g/dL)** confirms nephrotic syndrome. Dysmorphic hematuria (≥100 RBCs/HPF) indicates glomerular inflammation, collectively signaling severe glomerular injury beyond typical diabetic nephropathy.<br>• **Normal complements and negative serologies** exclude lupus nephritis, vasculitis, or infection-associated MPGN, focusing the diagnosis on IgAN.<br>• **Biopsy findings** (mesangial/endocapillary proliferation, double contours, IgA deposits in capillary walls, Gd-IgA1 positivity) confirm **MPGN-type IgAN**—a rare, aggressive aggressive variant characterized by subendothelial deposits and mesangial interposition. This explains the nephrotic syndrome and hematuria. |
| Gemini 2.5 Pro | **Interpretation of Test Results**<br>The test results collectively point to a severe, active glomerular disease. The nephrotic-range proteinuria (5.25 g/gCr) and hypoalbuminemia (2.9 g/dL) explain the patient's pitting edema and confirm nephrotic syndrome. The urinalysis showing dysmorphic red blood cells (>100/HPF) is a critical finding, indicating glomerular inflammation and bleeding (glomerulonephritis), which is atypical for diabetic nephropathy or AA amyloidosis. The elevated serum IgA is a supportive clue for IgA nephropathy (IgAN). The negative serological workup (ANCA, ANA, etc.) effectively rules out other common causes of rapidly progressive glomerulonephritis.<br><br>The renal biopsy provides the definitive explanation. The light microscopy findings of mesangial and endocapillary proliferation with double contours define a membranoproliferative glomerulonephritis (MPGN) pattern, a morphology associated with severe glomerular injury and poor prognosis. The immunofluorescence showing IgA-dominant deposits identifies the underlying cause of this injury pattern. Finally, the positive staining for galactose-deficient IgA1 (Gd-IgA1) specifically confirms the diagnosis of primary IgA nephropathy, as Gd-IgA1 is the pathogenic molecule central to this disease. |
| OpenAI GPT-o3 | Interpretation of the test results<br>• Nephrotic-range proteinuria (5.25 g/gCr) + hypoalbuminemia (2.9 g/dL) + pitting edema = nephrotic syndrome.<br>• Grossly normal creatinine (1.0 mg/dL, eGFR 56 mL/min) indicates preserved renal function, so the lesion is relatively early/reversible.<br>• Dysmorphic RBCs (≥100/HPF) signify glomerular hematuria, pointing to a primary glomerular process rather than diabetic nephropathy.<br>• Markedly elevated serum IgA (547 mg/dL) with normal complements supports IgA-mediated immune complex disease rather than classical complement-mediated MPGN.<br>• Renal biopsy is decisive:<br>− Light microscopy: mesangial ± endocapillary proliferation with double contours = |

| | |
|---|---|
| | MPGN pattern.<br>− Immunofluorescence: IgA-dominant deposits mesangial and capillary → excludes primary membranous nephropathy or AA amyloidosis.<br>− Electron microscopy: mesangial + subendothelial EDDs with mesangial interposition typical of MPGN-type IgA nephropathy.<br>− Positive KM55 (Gd-IgA1) staining confirms pathogenic galactose-deficient IgA1 deposition.<br>Taken together, these data firmly establish IgA nephropathy with an MPGN morphological pattern, presenting as nephrotic syndrome. |
| Llama4-Marveric | Interpretation of Test Results<br>The patient's laboratory results and renal biopsy findings are crucial for understanding his condition. Key findings include:<br>• Elevated serum IgA levels and IgA-dominant deposits in the kidney, indicative of IgA Nephropathy (IgAN).<br>• Nephrotic-range proteinuria and hematuria with dysmorphic red blood cells, suggesting a glomerular origin.<br>• Renal biopsy showing mesangial and endocapillary proliferation, double-contoured basement membranes, and IgA-dominant deposits, leading to a diagnosis of Membranoproliferative Glomerulonephritis (MPGN)-type IgAN.<br>• Positive staining with Gd-IgA1-specific antibody, supporting the diagnosis of IgAN.<br><br>These findings point towards IgAN with an MPGN pattern, a rare variant. The presence of hematuria and significant proteinuria, along with the biopsy findings, supports this diagnosis. |
| **Phase II – Q6: Reconsidering the differential diagnosis** | |
| DeepSeek-R1 | Revised Diagnostic Thinking<br>• **Prioritization shift:** Initial differentials (amyloidosis, diabetic nephropathy) were ruled out:<br>• Biopsy excluded amyloid deposits and diabetic changes (no Kimmelstiel-Wilson nodules).<br>• Elevated IgA, Gd-IgA1 positivity, and MPGN pattern override diabetes/RA as primary drivers.<br>• **Working Diagnosis: MPGN-type IgA nephropathy.** RA may contribute via chronic immune dysregulation, but IgAN is the dominant pathology. The MPGN pattern signifies high-risk disease (rapid progression risk). |
| Gemini 2.5 Pro | **Revised Diagnostic Thinking and Working Diagnosis**<br>My initial suspicion of AA amyloidosis, based on the patient's long history of RA, is now refuted by the biopsy, which showed a proliferative process, not amyloid deposits. Similarly, primary membranous nephropathy is excluded by the IgA-dominant immunofluorescence. While diabetic nephropathy likely co-exists, the acute presentation with active, proliferative glomerulonephritis is not explained by diabetes alone.<br><br>The new, definitive evidence from the laboratory and biopsy has completely shifted the diagnosis. The combination of an MPGN pattern of injury driven by IgA deposition leads to a specific and severe diagnosis.<br><br>**Working Diagnosis: IgA Nephropathy with a Membranoproliferative (MPGN) Pattern of Injury (High-Risk IgAN)** |
| OpenAI GPT-o3 | Updated diagnostic thinking<br>Initial differential (membranous GN, diabetic nephropathy, AA amyloidosis) is now supplanted by a single, biopsy-proven entity.<br>Working Diagnosis: MPGN-type IgA nephropathy (IgA-dominant immune-complex MPGN) causing nephrotic syndrome in a 71–75-year-old man with controlled RA and diabetes. |

| | |
|---|---|
| Llama4-Marveric | Diagnostic Thinking and Working Diagnosis<br>Given the biopsy results and laboratory findings, the most likely diagnosis is **MPGN-type IgAN.** The elevated serum IgA, presence of IgA-dominant deposits in the kidney, and specific biopsy findings are diagnostic hallmarks. The patient's RA and its treatment may be relevant, as some RA treatments can influence kidney disease, but the primary diagnosis here is IgAN with an MPGN pattern. |
| **Phase II – Q7: Treatment planning** | |
| DeepSeek-R1 | Treatment Plan<br>Proposed Approach:<br>**1. Supportive Care:**<br>• **Maximize ACEi/ARB** (e.g., losartan) to reduce proteinuria (target <1 g/day). Rationale: First-line for IgAN; slows CKD progression (KDIGO guidelines).<br>• **Optimize diabetes control:** Intensify insulin to target HbA1c ≤7.0%, avoiding nephrotoxic agents.<br>**2. Immunosuppression:**<br>• **Corticosteroids:** Prednisone 0.5 mg/kg/day for 2-4 months, then taper. Rationale: Indicated for high-risk IgAN (nephrotic-range proteinuria, eGFR >50). Reduces inflammation and proteinuria.<br>• **Consider budesonide** (targeted-release) if available: Lower systemic toxicity in elderly/diabetic patients.<br>**3. RA Therapy Adjustment:**<br>• **Continue methotrexate** (renal dose adjustment if eGFR declines).<br>• **Hold tofacitinib/iguratimod:** JAK inhibitors lack evidence in IgAN; iguratimod may exacerbate renal injury. Monitor RA flare risk.<br><br>Rationale and Alternatives:<br>• **Evidence:** Corticosteroids + ACEi reduce proteinuria by 60% in nephrotic IgAN (TESTING trial). MPGN pattern warrants aggressive control.<br>• **Alternatives:**<br>• If steroids contraindicated: **Mycophenolate mofetil** (MMF) as steroid-sparing agent.<br>• Refractory cases: **Rituximab** (emerging evidence for IgAN).<br>• **Avoid:** Cyclophosphamide (excessive toxicity in elderly/diabetic).<br>• **Monitoring:** Proteinuria, eGFR, serum IgA every 3 months; renal ultrasound if function declines.<br>**Summary:** Target MPGN-IgAN with ACEi/ARB + corticosteroids, prioritizing renal protection while managing RA/diabetes comorbidity.<br>Please enter the treatment course. |
| Gemini 2.5 Pro | **Treatment Plan**<br>The goal is to reduce proteinuria and preserve kidney function. The patient's high-risk features (nephrotic-range proteinuria, eGFR < 60 mL/min/1.73m², active inflammation on biopsy) demand both supportive care and immunosuppression.<br>**1. Foundational Supportive Care (to be initiated immediately):**<br>• **RAAS Blockade:** Start an Angiotensin Receptor Blocker (ARB) or ACE inhibitor. Titrate to the maximum tolerated dose to reduce proteinuria and control blood pressure.<br>• **Blood Pressure Control:** Target a strict blood pressure of <130/80 mmHg.<br>• **Diabetes Management:** Optimize insulin therapy to maintain target HbA1c, while being mindful that kidney function changes can affect insulin clearance.<br>• **Lifestyle Modification:** Institute a low-sodium diet (<2g/day) to manage edema and blood pressure.<br>**2. Immunosuppressive Therapy:**<br>• **Rationale:** Supportive care alone is insufficient for such severe disease. The active endocapillary proliferation on biopsy indicates an inflammatory process that may respond |

| | |
|---|---|
| | to immunosuppression. The aim is to induce remission and prevent progression to end-stage kidney disease.<br>• **Recommended Treatment:** Given the patient's age (71–75) and diabetes, the significant side effects of systemic corticosteroids (e.g., hyperglycemia, infections, osteoporosis) are a major concern. Therefore, I would recommend a 9-month course of **targeted-release formulation of budesonide (Nefecon)**. This therapy is designed to act on the gut-associated lymphoid tissue (Peyer's patches), a primary source of pathogenic Gd-IgA1, with lower systemic steroid exposure and a more favorable safety profile. This is an evidence-based first-line option for high-risk IgAN.<br>• **Alternative Option:** A 6-month course of **systemic corticosteroids** (e.g., oral prednisolone) is a traditional evidence-based option shown to reduce the risk of kidney failure in high-risk IgAN. However, this would be a second choice due to the higher risk of adverse events in this patient. A thorough discussion of the risk-benefit profile with the patient is essential before starting. The RA medications (methotrexate, tofacitinib) are not standard treatments for IgAN and should be continued under the guidance of a rheumatologist, with heightened vigilance for infection due to cumulative immunosuppression. |
| OpenAI GPT-o3 | Treatment plan and rationale<br>A. Supportive therapy (cornerstone; KDIGO 2021)<br>• Maximum tolerated ACE-I or ARB to lower proteinuria/BP (<120/70 mmHg if feasible).<br>• SGLT2 inhibitor (e.g., dapagliflozin) for dual renal and glycemic benefit; eGFR 56 mL/min is adequate.<br>• Statin and salt (<4 g/day)/protein (~0.8 g/kg) restriction; loop diuretic for edema.<br><br>B. Immunosuppressive therapy (indicated because proteinuria >3 g despite preserved GFR)<br>• Oral prednisolone 0.6–0.8 mg/kg/day (max 40 mg) for 2 months, then taper over 4–6 months, monitoring glycemia and infection risk.<br>• Consider steroid-sparing or adjunct options if intolerance/high risk:<br>− Targeted-release budesonide (NEFECON) if available.<br>− Mycophenolate mofetil or cyclophosphamide in rapidly progressive cases (not currently warranted).<br>• Tonsillectomy plus steroid pulse is practiced in Japan for proliferative IgAN; age/comorbidity must be weighed.<br><br>C. RA drug adjustment<br>• Continue methotrexate only if renal function remains stable; reduce dose if GFR falls.<br>• Tofacitinib and iguratimod add immunosuppressive burden—consider dose reduction or temporary cessation during high-dose steroids to lower infection risk.<br><br>D. Monitoring<br>• Monthly labs (proteinuria, serum creatinine, IgA, glucose, CRP).<br>• Screen for steroid complications (BP, weight, glucose, bone protection).<br>• Review need for pneumocystis prophylaxis given combined immunosuppression.<br>Alternative strategies<br>• If proteinuria falls below 1 g with supportive care alone, defer steroids (“conservative management” per STOP-IgA trial).<br>• Enrollment in clinical trials of complement inhibitors or anti-Gd-IgA1 therapies if progression despite standard care.<br>Goal: reduce proteinuria <1 g/day, maintain eGFR, and minimize treatment-related adverse events. |
| Llama4- | Treatment Plan |

| | |
|---|---|
| Marveric | **1. Immunosuppressive Therapy:** Given the diagnosis of MPGN-type IgAN, immunosuppressive therapy may be considered to reduce proteinuria and hematuria, and to slow disease progression. Corticosteroids are a common first-line treatment.<br>**2. RA Management Adjustment:** Continue or adjust RA treatment as necessary to maintain remission while minimizing kidney risk. The current regimen includes methotrexate, tofacitinib, and iguratimod; adjustments may be needed based on kidney function and disease activity.<br>**3. Supportive Care:** Manage hypertension, proteinuria, and edema with ACE inhibitors or ARBs if not contraindicated, and diuretics as needed.<br>**4. Monitoring:** Regular follow-up with nephrology to monitor kidney function, proteinuria, and adjust treatment as necessary.<br><br>Rationale: The treatment aims to reduce kidney inflammation and damage, manage symptoms, and slow disease progression. Alternative options include other immunosuppressive agents if corticosteroids are not effective or tolerated. |
| **Phase III – Q8: Evaluation of treatment** | |
| DeepSeek-R1 | Monitoring Effectiveness, Safety Risks, and Countermeasures<br>**Effectiveness Monitoring:**<br>• **Proteinuria:** Spot urine protein-to-creatinine ratio (UPCR) monthly for 3 months, then quarterly. Target: Maintain remission (UPCR <0.5 g/gCr).<br>• **Renal function:** Serum creatinine/eGFR every 3 months. Target: Stable eGFR (±10% baseline).<br>• **Serum albumin:** Quarterly until normal (≥3.5 g/dL), confirming resolution of nephrotic syndrome.<br>• **Hematuria:** Urinalysis with microscopy quarterly; persistence suggests ongoing inflammation.<br>**Safety Risks and Countermeasures:**<br>• **Hyperglycemia:**<br>• Risk: Steroids worsen diabetes; HbA1c rose to 7.4% pre-treatment.<br>• Countermeasure: Weekly fasting glucose checks; HbA1c every 3 months. Adjust insulin proactively.<br>• **Infection** (e.g., pneumonia, UTI):<br>• Risk: Age, diabetes, steroids, and prior immunosuppressants (tofacitinib).<br>• Countermeasure: Vaccinate (influenza/pneumococcal); avoid live vaccines. Prompt CRP/imaging for fever.<br>• **Osteoporosis:**<br>• Risk: Age, steroids, reduced mobility from RA/edema.<br>• Countermeasure: Baseline DEXA scan; calcium/vitamin D supplementation; add bisphosphonate if T-score ≤－2.5.<br>• **Adrenal insufficiency:**<br>• Risk: Prolonged steroid use (>3 months), rapid taper.<br>• Countermeasure: Slow taper (≤2.5 mg/month below 10 mg/day); educate on stress dosing (e.g., surgery/illness). |
| Gemini 2.5 Pro | **Monitoring Treatment Effectiveness and Safety**<br>**Effectiveness Monitoring:**<br>The primary goal is to maintain the complete remission of proteinuria and preserve kidney function.<br>• **Proteinuria:** Monthly measurement of urine protein-to-creatinine ratio (UPCR). The target is to keep it <0.3 g/gCr.<br>• **Kidney Function:** Monthly measurement of serum creatinine (s-Cr) and eGFR to ensure stability.<br>• **Urinalysis:** Monthly urine microscopy to monitor for the resolution of hematuria. |

| | |
|---|---|
| | **Safety Monitoring and Management:**<br>Given the patient's age, comorbidities (diabetes), and concurrent immunosuppressants (methotrexate, tofacitinib), vigilant monitoring for steroid-related side effects is critical.<br>• **Infection:**<br>• **Risk:** High risk of opportunistic infections due to cumulative immunosuppression.<br>• **Countermeasures:** Educate the patient to immediately report any signs of infection (fever, cough). Administer prophylaxis for Pneumocystis jirovecii pneumonia (e.g., with trimethoprim-sulfamethoxazole) while on significant immunosuppression. Ensure vaccinations (influenza, pneumococcus) are up-to-date.<br>• **Hyperglycemia:**<br>• **Risk:** Worsening of diabetes control.<br>• **Countermeasures:** Frequent self-monitoring of blood glucose. Regular HbA1c measurement (e.g., every 3 months). Proactively adjust insulin regimen in collaboration with the patient's diabetes care provider.<br>• **Osteoporosis:**<br>• **Risk:** Steroid-induced bone density loss, leading to fractures.<br>• **Countermeasures:** Prescribe calcium and vitamin D supplementation. Perform a baseline bone mineral density (DEXA) scan and consider initiating bisphosphonate therapy based on fracture risk.<br>• **Gastric Ulcers:**<br>• **Risk:** Peptic ulcer disease.<br>• **Countermeasures:** Administer a proton pump inhibitor (PPI) for gastric protection during the period of high-dose steroid therapy. |
| OpenAI GPT-o3 | Monitoring plan<br>Effectiveness<br>• Spot urine protein-to-creatinine ratio (or 24 h protein) and urine sediment: monthly for 6 mo, then every 2–3 mo; goal < 0.3 g/gCr.<br>• Serum albumin, creatinine/eGFR, and body weight/edema: same schedule.<br>• Blood pressure: each visit; maintain < 120/70 mmHg if tolerated.<br><br>Safety key risks & countermeasures<br>• Hyperglycaemia: home capillary glucose daily, HbA1c every 3 mo; up-titrate insulin/ add SGLT2 inhibitor.<br>• Infection (bacterial, viral, Pneumocystis): baseline TB/HBV screen, vaccines up to date; counsel on fever, start TMP-SMX prophylaxis, hold/adjust MTX ± tofacitinib during high-dose PSL or infection.<br>• Hypertension/fluid retention: BP checks, Na restriction, loop diuretic or add/adjust ACE-I/ARB.<br>• Osteoporosis/fracture: baseline DXA, Ca 1 g + Vit D 800 IU/day, start bisphosphonate; repeat DXA yearly.<br>• GI ulcer/bleed: PPI prophylaxis, test/eradicate H. pylori if history.<br>• Dyslipidaemia/CVD: lipid panel at 0 and 3 mo, statin as needed.<br>• Myopathy/psychosis: ask each visit, taper or switch therapy if severe.<br>• Ocular complications: ophthalmology exam baseline, then yearly.<br>• Adrenal suppression: taper prednisolone gradually; give stress-dose steroids for surgery/intercurrent illness.<br>• MTX toxicity: CBC, LFT monthly; folic acid 5 mg/wk. |
| Llama4-Marveric | Monitoring the Effectiveness and Safety of the Treatment<br>To monitor the effectiveness and safety of the treatment with prednisolone: |

<table>
<tr><td></td><td>1. Effectiveness:<br>• Regularly check proteinuria levels to ensure they remain low.<br>• Monitor serum albumin levels to assess improvement in nephrotic syndrome.<br>• Periodically assess kidney function (e.g., eGFR, serum creatinine).<br>2. Safety:<br>• Monitor for signs of corticosteroid side effects: glucose control (HbA1c), blood pressure, weight, and signs of osteoporosis (e.g., bone density scans).<br>• Check for infections, as immunosuppression increases infection risk.<br>• Monitor for mood changes or other psychological side effects. Specific Risks and Countermeasures<br><br>1. Risk of Infection: Monitor for signs of infection; consider prophylactic measures (e.g., pneumocystis pneumonia prophylaxis).<br>2. Osteoporosis: Consider calcium and vitamin D supplements, and possibly bisphosphonates if long-term steroid use is anticipated.<br>3. Hyperglycemia: Closely monitor blood glucose levels, especially given the patient's diabetes history; adjust diabetes management as needed.</td></tr>
<tr><td colspan="2">Phase III – Q9: Formulating plans for worsening conditions</td></tr>
<tr><td>DeepSeek-R1</td><td>Response to Non-improvement or Sudden Change<br>Thought Process and Actions:<br>• Scenario 1: Proteinuria relapse (UPCR >1 g/gCr) during taper:<br>• Think: Steroid dependency, non-adherence, or new insult (e.g., infection, RA flare).<br>• Act:<br>a. Rule out infection (urine culture, CRP).<br>b. Increase prednisolone to last effective dose (e.g., 20 mg/day).<br>c. Add steroid-sparing agent (e.g., mycophenolate mofetil 1 g/day) if relapse recurs.<br>• Scenario 2: Acute kidney injury (eGFR drop >25%):<br>• Think: Drug toxicity (e.g., NSAIDs for RA pain), dehydration, renal vein thrombosis (hypercoagulability in nephrosis), or RPGN transformation.<br>• Act:<br>a. Urgent renal ultrasound with Doppler (thrombosis/obstruction).<br>b. Serologies (ANCA, anti-GBM) and urinalysis for casts (RPGN).<br>c. If RPGN suspected: Pulse methylprednisolone (500 mg × 3 days) and re-biopsy.<br>• Scenario 3: New-onset gross hematuria:<br>• Think: IgAN flare, urothelial malignancy (age/RA risk), or stones.<br>• Act: CT urogram and cystoscopy to exclude malignancy.<br><br>Diagnostic Revision Principles:<br>• Biopsy repeat indication: eGFR decline + active sediment (RBC casts) suggesting RPGN or new pathology.<br>• Treatment Revision:<br>• Steroid-refractory: Switch to rituximab (anti-CD20) or targeted therapies (e.g., sparsentan).<br>• Avoid nephrotoxins (e.g., NSAIDs); optimize RA control with renal-safe DMARDs (sulfasalazine > JAK inhibitors).</td></tr>
<tr><td>Gemini 2.5 Pro</td><td>Response to Unexpected Changes in Condition<br>If the patient's condition deteriorates (e.g., proteinuria relapse, acute rise in serum creatinine), my response will be systematic, aimed at rapidly identifying the cause and adjusting the strategy.<br>Thought Process:<br>First, I would differentiate between three main possibilities:<br>1. Relapse of IgA Nephropathy: The underlying disease is becoming active again,</td></tr>
</table>

| | |
|---|---|
| | perhaps due to steroid tapering or the natural course of the disease.<br>**2. Treatment-Related Complication:** The deterioration is caused by a side effect, such as a severe infection leading to sepsis and pre-renal acute kidney injury (AKI), or drug-induced acute interstitial nephritis.<br>**3. A New, Unrelated Condition:** A new problem has emerged, such as AKI from dehydration, urinary obstruction, or a different superimposed glomerular disease.<br><br>**Action Plan:**<br>**1. Immediate Reassessment:** Conduct a thorough history and physical examination, focusing on signs of infection, volume status, and any new medications (including over-the-counter drugs).<br>**2. Urgent Investigations:** Order immediate laboratory tests including urinalysis with microscopy (to look for new/worsening active sediment like RBC casts), UPCR, s-Cr, CBC, and C-reactive protein. A renal ultrasound would be performed to rule out post-renal obstruction.<br>**3. Revise Diagnosis and Treatment:**<br>• **If a relapse of IgAN is most likely** (indicated by rising proteinuria and active urine sediment without signs of infection), I would reconsider the immunosuppressive regimen. This may involve temporarily increasing the prednisolone dose or considering the addition/switch to a different agent.<br>• **If an infection is suspected,** treatment with appropriate antibiotics would be the priority. Immunosuppressants might need to be temporarily held or reduced depending on the severity of the infection.<br>• **If the cause remains unclear after initial investigation, a repeat renal biopsy** would be strongly considered. This is the definitive tool to determine if the pathology has evolved (e.g., development of crescents, increased chronicity) and to guide the next therapeutic steps accurately. |
| OpenAI GPT-o3 | Response to inadequate or sudden change<br>Step-wise reassessment<br>1. Confirm: repeat UPCR, creatinine, adherence, BP, RAAS/SGLT2 optimisation, exclude nephrotoxic drugs.<br>2. Search for triggers: infection, RA flare, diabetic nephropathy progression, renal vein thrombosis, drug-induced interstitial nephritis.<br>3. If gradual non-response (>1 g/gCr or <50 % fall at 6 mo):<br>• Reinforce supportive care;<br>• Escalate immunosuppression (e.g., add MMF 1.5–2 g/d, targeted-release budesonide, or low-dose cyclophosphamide);<br>• Consider tonsillectomy (Japan) or clinical-trial agents (complement/Gd-IgA1 blockers).<br>4. If rapid decline (eGFR fall ≥50 % or nephritic urine): admit, repeat serology/biopsy to rule out crescentic transformation; give pulse methylprednisolone ± plasmapheresis if RPGN.<br>5. If severe steroid toxicity: accelerate taper, substitute with MMF/azathioprine/rituximab, intensify infection and metabolic prophylaxis.<br>6. Multidisciplinary review (nephrology-rheumatology-endocrinology), advance-care planning, and early RRT education if progression persists. |
| Llama4-Marveric | Response to Unexpected Changes or Lack of Improvement<br>If the patient's condition does not improve as expected or suddenly changes:<br>**1. Reassess Diagnosis:** Verify the initial diagnosis and consider alternative or additional diagnoses.<br>**2. Check for Treatment Adherence:** Ensure the patient is adhering to the prescribed treatment regimen.<br>**3. Investigate for Complications:** Look for complications such as infections or thrombosis that could explain the lack of response or sudden change.<br>**4. Adjust Treatment Plan:** Consider alternative immunosuppressive therapies or adjust |

| | |
|---|---|
| | the corticosteroid regimen if necessary.<br>**5. Further Investigations:** Additional tests or biopsies might be needed to understand the cause of the change in condition.<br><br>By closely monitoring the patient's response to treatment and being prepared to adjust the treatment plan as needed, we can optimize outcomes and minimize risks. |

| **Case 3: Case Report: A case of membranous nephropathy associated with primary Sjögren's syndrome treated with telitacicept [4]** | |
|---|---|
| **Phase I – Q1: Summarizing medical problems** | |
| DeepSeek-R1 | 41–45-year-old female with **5-month history** of sicca symptoms (dry mouth, dry eyes) and **persistent nephrotic-range proteinuria** (3+ urine protein, hypoalbuminemia 26.8 g/L) despite antihypertensive/immunosuppressive therapy (losartan/tripterygium). Chronic hypertension (>10 years) and normal creatinine (54 μmol/L). |
| Gemini 2.5 Pro | This is a 41–45-year-old female with a 10-year history of hypertension presenting with a 5-month history of nephrotic-range proteinuria (proteinuria 3+, serum albumin 26.8 g/L), microscopic hematuria, and prominent sicca symptoms (dry mouth and eyes), with preserved renal function. |
| OpenAI GPT-o3 | 41–45-year-old woman with 5-month history of sicca symptoms (dry mouth, dry eyes) and progressively increasing urinary abnormalities (proteinuria 2+→3+, microscopic hematuria), associated hypoalbuminemia (26.8 g/L) but preserved renal function (Cr 54 μmol/L), on long-standing hypertension treatment. |
| Llama4-Marveric | The patient is a 41–45-year-old female presenting with a 5-month history of dry mouth, dry eyes, and foamy urine. She has a background of hypertension for over 10 years. Key findings include persistent proteinuria (2-3+), recent onset of hematuria, hypoalbuminemia (26.8 g/L), and normal creatinine levels. |
| **Phase I – Q2: Differential diagnoses and rationale** | |
| DeepSeek-R1 | Differential Diagnoses<br>**1. Most likely: Sjögren syndrome-associated glomerulonephritis**<br>• Rationale: Sicca symptoms + proteinuria suggest autoimmune tubulointerstitial or membranoproliferative disease.<br>**2. Systemic lupus erythematosus (SLE) nephritis**<br>• Rationale: Nephrotic syndrome with extrarenal symptoms (sicca) and occult hematuria.<br>**3. Hypertensive nephrosclerosis**<br>• Rationale: Long-standing HTN, though atypical with nephrotic-range proteinuria.<br>**Must not miss:** Amyloidosis or vasculitis (e.g., ANCA-associated), given nephrotic syndrome and systemic symptoms. |
| Gemini 2.5 Pro | Differential Diagnosis<br>**1. Sjögren's Syndrome-associated Nephropathy:** A systemic autoimmune disease that classically presents with sicca symptoms and can cause various renal manifestations, including glomerulonephritis (commonly membranous nephropathy or MPGN) leading to nephrotic syndrome.<br>**2. Lupus Nephritis (Systemic Lupus Erythematosus - SLE):** Another common autoimmune disease in this demographic that can present identically with nephrotic syndrome and secondary sicca symptoms.<br>**3. Primary (Idiopathic) Membranous Nephropathy:** The most common cause of primary nephrotic syndrome in adults. The sicca symptoms could be a coincidental finding.<br>• **Most Likely:** Sjögren's Syndrome-associated Nephropathy. The combination of prominent, long-standing sicca symptoms and nephrotic syndrome in a middle-aged female is highly suggestive of this diagnosis.<br>• **Must Not Be Missed:** Lupus Nephritis. Given the patient's age and sex, SLE is a critical differential. It is a multi-system disease with significant morbidity if the diagnosis is delayed, and its treatment is distinct from other causes. |

| | |
|---|---|
| OpenAI GPT-o3 | Differential Diagnoses<br>a. Primary Sjögren's syndrome with renal involvement (tubulo-interstitial nephritis or membranous / IgA nephropathy) – most likely because sicca symptoms precede renal findings in a middle-aged woman.<br>b. Systemic lupus erythematosus with lupus nephritis – must not be missed owing to potential rapid renal deterioration and need for specific therapy.<br>c. Primary membranous nephropathy (or other primary glomerulopathy such as IgA nephropathy) unrelated to sicca symptoms – common cause of proteinuria/hematuria; sicca may be coincidental. |
| Llama4-Marveric | Differential Diagnoses<br>**1. Sjögren's Syndrome with Renal Involvement:** Given her symptoms of dry mouth and dry eyes, Sjögren's Syndrome is a strong consideration. Renal involvement in Sjögren's can manifest as tubulointerstitial nephritis or glomerulonephritis, which could explain her proteinuria and hematuria.<br>**2. Membranous Nephropathy or Other Glomerulonephritis:** The presence of significant proteinuria and hypoalbuminemia suggests a glomerular pathology. Membranous nephropathy is a possibility, especially if there's an underlying autoimmune or secondary cause.<br>**3. Hypertensive Nephrosclerosis:** With a long history of hypertension, hypertensive nephrosclerosis is a consideration, though it typically doesn't present with nephrotic range proteinuria.<br><br>The most likely diagnosis is **Sjögren's Syndrome with Renal Involvement** due to the combination of sicca symptoms and renal findings. **Rapidly Progressive Glomerulonephritis** must not be missed, given the new onset of hematuria. |
| **Phase I – Q3: Necessary physical examinations and reasons** | |
| DeepSeek-R1 | Targeted History/Physical Exam<br>**1. History:**<br>• "Any joint pain, photosensitivity, or rash?" (Rule out SLE).<br>• "Recent infections or weight loss?" (Assess for amyloid/vasculitis).<br>**2. Physical Exam:**<br>• Schirmer test/salivary gland palpation (Confirm sicca syndrome).<br>• Skin/joint exam (Look for lupus/vasculitis signs).<br>• Edema assessment (Evaluate nephrotic severity).<br>Purpose: Confirm autoimmune etiology and rule out systemic involvement. |
| Gemini 2.5 Pro | Additional History and Physical Examination<br>**Additional Questions:**<br>• **Systemic Review:** To screen for SLE or other connective tissue diseases, I would ask about joint pain/swelling, photosensitive skin rashes (e.g., malar rash), oral ulcers, hair loss, and Raynaud's phenomenon.<br>• **Quantify Sicca Symptoms:** To better characterize Sjögren's, I would ask about a gritty or sandy sensation in the eyes, use of artificial tears, difficulty swallowing dry foods without liquids, and any enlargement of salivary glands.<br>• **Hypertension Control:** To assess the contribution of hypertension, I would ask about her typical blood pressure readings at home and her adherence to medication.<br><br>**Physical Examination:**<br>• **Blood Pressure:** To assess current control and its potential contribution to renal injury.<br>• **General Examination:** To check for peripheral edema, which is a sign of severe hypoalbuminemia and fluid retention characteristic of nephrotic syndrome.<br>• **HEENT:** To examine for parotid gland enlargement and objective evidence of dry oral mucosa or dental caries (common in Sjögren's).<br>• **Skin and Joints:** To look for rashes (malar, discoid) or evidence of synovitis, which |

| | |
|---|---|
| | would point towards SLE. |
| OpenAI GPT-o3 | Targeted History / Physical Examination<br>• Evolution and severity of sicca: parotid swelling, dental caries → supports Sjögren's.<br>• Systemic autoimmune features: photosensitive rash, oral/nasal ulcers, arthritis, Raynaud's, serositis → suggest SLE.<br>• Constitutional / malignancy signs: weight loss, night sweats, lymphadenopathy → secondary membranous.<br>• Medication / herbal use (incl. tripterygium), infection history → drug / infection-related nephropathy.<br>• Volume status, edema degree, BP trend → nephrotic range impact.<br>• Physical: parotid gland enlargement, cutaneous lupus signs, joint swelling, lower-extremity edema. |
| Llama4-Marveric | Additional Questions and Physical Exams<br>• **Sicca Symptoms Clarification:** Ask about the severity and duration of dry mouth and eyes to support the diagnosis of Sjögren's Syndrome.<br>• **Hypertension History:** Clarify the control and duration of hypertension to assess its potential impact on renal function.<br>• **Medication History:** Review the patient's adherence to losartan and tripterygium wilfordii, and any other medications that could affect renal function.<br>• **Physical Examination:** Look for signs of dry eyes and mouth, and assess for edema, hypertension, or other systemic signs that could point to a specific diagnosis. |
| **Phase I – Q4: Plan for investigations/tests** | |
| DeepSeek-R1 | Diagnostic Testing Plan<br>**1. Serology:**<br>• ANA, anti-dsDNA, anti-SSA/SSB, ANCA, RF<br>• Purpose: Identify autoimmune markers (e.g., anti-SSA/SSB for Sjögren; anti-dsDNA for lupus).<br>• Expected: Positive anti-SSA/SSB in Sjögren; ANCA+ if vasculitis.<br>**2. Complement levels (C3/C4):**<br>• Purpose: Low in active lupus nephritis.<br>**3. 24-hour urine protein/UPCR:**<br>• Purpose: Quantify nephrotic-range proteinuria (>3.5 g/day).<br>**4. Renal ultrasound:**<br>• Purpose: Rule out structural causes; expect normal size.<br>**5. SPEP/immunofixation:**<br>• Purpose: Screen for paraproteinemias (amyloidosis).<br>**6. Renal biopsy (if serology equivocal):**<br>• Purpose: Gold standard for diagnosing glomerulopathy (e.g., membranous LN, amyloid).<br><br>Testing rationale: Cost-effective serology first; biopsy if diagnosis unclear after non-invasive tests. |
| Gemini 2.5 Pro | Diagnostic Test Plan<br>**1. Urine Tests:**<br>• **Purpose:** To confirm and quantify the proteinuria and to assess for signs of active glomerulonephritis.<br>• **Tests & Expected Results:**<br>• **Urine Protein-to-Creatinine Ratio (UPCR) or 24-hour Urine Protein:** To accurately quantify proteinuria. A value >3.5 g/g or g/day would confirm nephrotic-range proteinuria.<br>• **Urine Sediment Microscopy:** To look for dysmorphic red blood cells and red blood cell casts, which would confirm a glomerular origin of the hematuria. |

| | |
|---|---|
| | **2. Blood Tests:**<br>• **Purpose:** To screen for the specific underlying autoimmune cause and rule out other secondary causes.<br>• **Tests & Expected Results:**<br>• **Serology:** ANA, Anti-SSA/Ro, Anti-SSB/La, Anti-dsDNA, Anti-Sm, and complements (C3, C4). Positive Anti-SSA/SSB would strongly support Sjögren's. Positive Anti-dsDNA/Sm with low complements would favor lupus nephritis.<br>• **Anti-phospholipase A2 receptor (PLA2R) antibody:** A negative result would argue against Primary Membranous Nephropathy and support a secondary cause.<br>• **Hepatitis B, C, and HIV serology:** To rule out common infectious causes of secondary glomerular disease.<br><br>**3. Renal Biopsy:**<br>• **Purpose:** This is the definitive test to establish the histological diagnosis, assess the severity of inflammation and chronicity (fibrosis), and guide specific immunosuppressive therapy.<br>• **Expected Results:** The biopsy will identify the specific type of glomerulonephritis (e.g., membranous nephropathy, focal segmental glomerulosclerosis, lupus nephritis class V, etc.) and the presence of any associated tubulointerstitial nephritis. This information is essential for both prognosis and treatment decisions. |
| OpenAI GPT-o3 | Investigations<br>• Urine analysis: microscopy (dysmorphic RBCs, casts) and 24 h protein quantification – characterise glomerular vs tubular pathology (expect >1 g/24 h if membranous).<br>• Serum chemistry: repeat creatinine, urea, electrolytes, lipid profile – renal baseline and nephrotic sequelae.<br>• Autoimmune panel: ANA, anti-Ro/La, RF, anti-dsDNA, complements (C3/C4) – confirm Sjögren’s (anti-Ro/La positive, normal/low complements) or SLE (positive dsDNA, low complements).<br>• Immunoglobulins & serum electrophoresis – hypergammaglobulinaemia typical of Sjögren’s.<br>• ESR/CRP – systemic inflammation.<br>• Kidney ultrasound – size, echogenicity; exclude obstruction.<br>• Renal biopsy (definitive) – determine exact lesion (interstitial nephritis vs membranous vs lupus class). Expect lymphocytic tubulo-interstitial infiltrate ± immune deposits if Sjögren’s. |
| Llama4-Marveric | Tests to Narrow Down the Diagnosis<br>**1. Renal Biopsy:** To definitively diagnose the cause of her renal involvement, such as tubulointerstitial nephritis or glomerulonephritis associated with Sjögren's Syndrome.<br>**2. Immunological Tests:** Including anti-SSA/SSB antibodies to support a diagnosis of Sjögren's Syndrome, and other autoimmune markers to rule out other causes of glomerulonephritis.<br>**3. Serum Protein Electrophoresis and Urine Protein Electrophoresis:** To assess for paraproteinemia or specific patterns of proteinuria.<br>**4. Renal Ultrasound:** To evaluate renal structure and rule out obstruction or other anatomical abnormalities.<br>These tests will help differentiate between potential causes of her renal symptoms and guide further management. |
| **Phase II – Q5: Interpreting test results** | |
| DeepSeek-R1 | Interpretation of Test Results<br>• **Autoantibodies (ANA 1:160+, anti-SSA/Ro52+++):** Confirm autoimmune activity consistent with primary Sjögren’s syndrome (pSS). Anti-SSA/Ro52 positivity is highly specific for pSS and correlates with extraglandular manifestations (e.g., renal |

| | |
|---|---|
| | involvement).<br>• **Renal biopsy findings:**<br>• IgG1+++/IgG4++/C1q++ granular deposits with subepithelial/subendothelial electron-dense deposits: Indicates **immune-complex-mediated atypical membranous nephropathy (MN).** Negative PLA2R rules out primary MN; dominant IgG1/IgG4 suggests pSS-driven pathology.<br>• Subendothelial deposits and mesangial proliferation: Unusual in classic MN; supports secondary MN linked to pSS.<br>• **Nephrotic syndrome markers (proteinuria 2939 mg/24h, ALB 23.4 g/L):** Reflect severe glomerular injury due to pSS-related MN.<br>• **Normal complements (C3/C4) and negative anti-dsDNA:** Distinguishes from lupus nephritis (typically low complements/dsDNA+).<br>• **Elevated ESSDAI (15):** Confirms high systemic pSS activity, explaining renal involvement. |
| Gemini 2.5 Pro | **Interpretation of Test Results**<br>The patient's test results collectively build a strong, cohesive picture of nephrotic syndrome caused by an active autoimmune disease.<br>• **Autoimmune Profile:** The positive ANA (1:160) and, most importantly, the highly positive anti-SSA/Ro-52 antibodies, combined with the clinical findings (dry mouth/eyes, positive tear test, glandular lesions on ultrasound), definitively establish the diagnosis of **Primary Sjögren's Syndrome (pSS)**. The negative anti-dsDNA/Sm and normal complements effectively rule out active Systemic Lupus Erythematosus (SLE).<br>• **Nephrotic Syndrome Confirmation:** The 24-hour urine protein of 2939 mg and severe hypoalbuminemia (23.4 g/L) confirm the presence of nephrotic syndrome, which is the direct cause of her foamy urine.<br>• **Primary vs. Secondary Membranous Nephropathy (MN):** The negative anti-PLA2R antibody test (both in serum and on the biopsy tissue) makes Primary (Idiopathic) MN highly unlikely. This strongly suggests the kidney disease is secondary to another condition, in this case, pSS.<br>• **Renal Biopsy Significance:** The biopsy is crucial as it reveals a complex and active immune-mediated glomerulonephritis. It shows features of both membranous nephropathy (subepithelial deposits) and a proliferative glomerulonephritis (subendothelial deposits, endothelial proliferation). This mixed pattern is characteristic of a secondary autoimmune cause. The immunofluorescence finding of dominant IgG1 and C1q deposition is also atypical for primary MN (which is typically IgG4-dominant) and is a hallmark of immune-complex deposition seen in conditions like pSS-associated or lupus nephritis.<br>• **Disease Activity:** The European Sjögren's Syndrome Disease Activity Index (ESSDAI) score of 15 signifies moderate-to-high systemic disease activity, with the renal involvement being a major contributor. This score confirms that the kidney disease is an active manifestation of her systemic pSS and justifies the need for systemic immunosuppression. |
| OpenAI GPT-o3 | Medical significance of the test results<br>• 24-h urine protein 2.9 g and serum albumin 23 g/L = nephrotic syndrome, explaining foamy urine; serum creatinine 50 µmol/L shows preserved renal function.<br>• ANA 1:160 and strongly positive anti-SSA / Ro-52 together with sicca symptoms, abnormal Schirmer test and parotid ultrasound fulfil ACR-EULAR criteria for primary Sjögren's syndrome (pSS).<br>• Hyper-gammaglobulinaemia (IgG 13 g/L) and mildly raised ESR are typical immune-activation markers in pSS. Complements are normal, arguing against active lupus.<br>• Serum and glomerular PLA2R negative, IgG subclass staining IgG1 > IgG4 and presence of C1q, plus subendothelial as well as subepithelial deposits on EM distinguish the lesion from primary (idiopathic) membranous nephropathy and point to secondary/"atypical" MN.<br>• Biopsy therefore establishes atypical membranous nephropathy secondary to pSS rather |

| | |
|---|---|
| | than lupus nephritis (no anti-dsDNA, normal C3/C4) or IgG4-related disease (IgG4 dominant pattern absent).<br>• After treatment: proteinuria almost abolished (uPCR 0.09 g/g), albumin normalised, immunoglobulins and ESSDAI fell to 0 – objective evidence of good therapeutic response. |
| Llama4-Marveric | Interpretation of Test Results<br>The patient's test results reveal several key findings:<br>• **Renal Biopsy:** Indicates atypical membranous nephropathy (AMN) with diffuse granular staining for IgG1 and C1q, and numerous electron-dense deposits under the glomerular epithelium and mesangial endothelium. This suggests an immune complex-mediated glomerulonephritis.<br>• **Immunological Tests:** Positive ANA, anti-SSA, and anti-Ro-52 antibodies support the diagnosis of Sjögren's Syndrome (SS). The high ESSDAI score of 15 indicates significant disease activity.<br>• **Serum Immunoglobulins:** Slightly elevated IgG and normal IgA and IgM levels. The decrease in these levels post-treatment suggests a response to therapy.<br>• **Urine Protein Quantification:** Significant proteinuria (2939 mg/24h) that resolves post-treatment, indicating effective management of the renal involvement.<br><br>These findings are consistent with **Sjögren's Syndrome-associated Membranous Nephropathy.** The presence of sicca symptoms, positive autoimmune serology, and characteristic renal biopsy findings support this diagnosis. |
| **Phase II – Q6: Reconsidering the differential diagnosis** | |
| DeepSeek-R1 | Diagnostic Thinking and Working Diagnosis<br>• **Initial differentials:** Included lupus nephritis, primary MN, or hypertensive nephropathy.<br>• **Key revisions post-testing:**<br>• Anti-SSA/Ro52+++ and sicca symptoms confirm **pSS** as the primary disease.<br>• Biopsy findings (IgG1/IgG4 dominance, subendothelial deposits) and negative PLA2R exclude primary MN/lupus.<br>• Normal complements and dsDNA negativity further rule out lupus.<br>• **Working Diagnosis: Atypical membranous nephropathy secondary to primary Sjögren's syndrome (pSS-MN).** |
| Gemini 2.5 Pro | **Revised Diagnostic Thinking and Working Diagnosis**<br>My diagnostic thinking has shifted from a broad differential diagnosis to a single, specific entity. The initial possibilities included pSS-associated nephropathy, lupus nephritis, and primary membranous nephropathy.<br>• **Lupus nephritis i**s now ruled out based on the negative anti-dsDNA/Sm serology and normal complement levels.<br>• **Primary membranous nephropathy** is excluded due to the negative serum and tissue anti-PLA2R antibody tests, as well as the atypical biopsy findings (IgG1/C1q deposition, significant subendothelial deposits).<br>• The evidence now converges conclusively on a single diagnosis. The constellation of clinical sicca symptoms, definitive pSS-specific autoantibodies (anti-SSA), and a renal biopsy showing an atypical, immune-complex mediated membranous-like pattern is pathognomonic for glomerulonephritis secondary to pSS.<br>**Working Diagnosis: Nephrotic Syndrome due to Membranous Nephropathy (Atypical/Secondary Pattern) associated with active Primary Sjögren's Syndrome.** |
| OpenAI GPT-o3 | Updated diagnostic thinking<br>Primary Sjögren's syndrome with renal involvement in the form of secondary (atypical) membranous nephropathy is now the working diagnosis. Other differentials (idiopathic MN, lupus nephritis, IgG4-related kidney disease) are less likely given serology (PLA2R–, dsDNA–, IgG4 pattern absent), complement levels, and biopsy features. |
| Llama4- | Diagnostic Thinking and Working Diagnosis |

| | |
|---|---|
| Marveric | The initial differential diagnoses included various causes of glomerulonephritis and renal involvement in autoimmune diseases. With the renal biopsy results and immunological tests, the diagnosis has been refined to **Sjögren's Syndrome-associated Membranous Nephropathy**. The atypical membranous nephropathy pattern on biopsy, combined with the clinical presentation and serological evidence of SS, supports this working diagnosis. |
| **Phase II – Q7: Treatment planning** | |
| DeepSeek-R1 | Treatment Plan<br>**Proposed Regimen:**<br>**1. Rituximab** (e.g., 1,000 mg × 2 doses, 2 weeks apart):<br>• Rationale: Targets CD20+ B-cells driving pSS and MN. EBM supports efficacy in pSS-MN for reducing proteinuria (post-treatment: 0.09 g/g.cr) and systemic activity (ESSDAI 0).<br>**2. Corticosteroids** (e.g., prednisone 0.5 mg/kg/day × 4 weeks, then taper):<br>• Rationale: Rapid control of inflammation; adjunct to rituximab for induction.<br>**3. Supportive care:**<br>• Continue ACEi (losartan) for proteinuria/HTN.<br>• Statin for hypercholesterolemia (total cholesterol 5.29 mmol/L).<br>**Alternative Options:**<br>• **Cyclophosphamide:** If rituximab unavailable (less preferred due to higher toxicity).<br>• **Belimumab:** If rituximab fails (B-cell inhibition, but limited pSS-MN data).<br>**Avoid:** Tripterygium wilfordii (ineffective per history).<br><br>**Rationale:** Rituximab is first-line for pSS with severe extraglandular disease (EBM: improves renal outcomes and sicca symptoms, as seen post-treatment). Steroids bridge initial immunosuppression. |
| Gemini 2.5 Pro | **Treatment Plan**<br>The treatment goal is to induce remission of both the nephrotic syndrome and the systemic Sjögren's activity (as measured by the ESSDAI score) by suppressing the underlying autoimmune process. The plan involves both supportive care and specific immunosuppression.<br>**1. Supportive (Non-immunosuppressive) Therapy:**<br>• **RAAS Blockade:** Continue Losartan. This is critical for its antiproteinuric effect and for blood pressure control, which helps slow the progression of kidney disease. The dose should be optimized to achieve maximal proteinuria reduction while maintaining safe blood pressure and potassium levels.<br>• **Blood Pressure Control:** Maintain a target BP of <130/80 mmHg.<br>• **Hyperlipidemia Management:** Initiate statin therapy to manage the hypercholesterolemia secondary to nephrotic syndrome and reduce cardiovascular risk.<br>• **Dietary Management:** Advise moderate sodium restriction to help manage blood pressure and potential fluid retention.<br><br>**2. Immunosuppressive Therapy:**<br>• **Rationale:** The patient has active, moderate-to-high activity disease (ESSDAI 15) with significant organ involvement (nephrotic syndrome). This requires systemic immunosuppression to control the autoimmune attack on the kidneys and salivary/lacrimal glands. The previously prescribed Tripterygium wilfordii should be discontinued in favor of a more evidence-based regimen.<br>• **First-line Regimen: Glucocorticoids plus Mycophenolate Mofetil (MMF).**<br>• **Glucocorticoids (e.g., Prednisolone 0.5 mg/kg/day):** To rapidly reduce glomerular inflammation and systemic activity. This would be initiated and then slowly tapered over several months based on clinical response (reduction in proteinuria and ESSDAI score).<br>• **Mycophenolate Mofetil (MMF, e.g., 1g twice daily):** MMF is a potent steroid-sparing agent effective in treating autoimmune glomerulonephritis, including that |

| | |
|---|---|
| | associated with pSS and lupus. It inhibits lymphocyte proliferation, targeting the core of the autoimmune process with a favorable long-term safety profile compared to other agents.<br><br>**3. Alternative Treatment Options:**<br>• **Rituximab:** A B-cell depleting monoclonal antibody. This is an excellent alternative, particularly in pSS where B-cell hyperactivity is central to the pathogenesis. It can be highly effective in inducing remission of both renal and extra-glandular manifestations and may be considered a first-line option, especially if there are contraindications to or intolerance of MMF. The profound drop in all immunoglobulin isotypes seen in the post-treatment data table is highly suggestive that a B-cell depleting therapy like Rituximab may have been the treatment ultimately used.<br>• **Cyclophosphamide:** A potent alkylating agent, typically reserved for severe, crescentic, or refractory disease due to its significant toxicity profile (risk of infection, infertility, and malignancy). It would not be a first-line choice in this case. |
| OpenAI GPT-o3 | Treatment plan and rationale<br><br>A. Disease-modifying therapy<br>• Prednisone 0.5 mg/kg/day (taper over 3–6 months) plus mycophenolate mofetil 1.5–2 g/day (or cyclophosphamide 1.5–2 mg/kg/day if MMF not tolerated) – evidence from pSS and MN cohorts supports combined glucocorticoid–antimetabolite therapy to induce and maintain remission.<br>• Rituximab 1 g × 2 (day 1 & 15) is a proven alternative for pSS with glomerular disease and can be used for refractory or relapsing cases.<br><br>B. Supportive nephro-cardiovascular care<br>• Optimise renin–angiotensin blockade: increase losartan to 50–100 mg daily if blood pressure allows, to maintain antiproteinuric and antihypertensive effect.<br>• Statin for borderline hypercholesterolaemia.<br>• Salt (<5 g/d) and protein (0.8–1 g/kg) dietary advice.<br><br>C. Preventive measures<br>• Calcium/vitamin D ± bisphosphonate for steroid bone protection.<br>• PJP prophylaxis (trimethoprim-sulfamethoxazole) if prednisone ≥ 20 mg/day plus second agent.<br>• Vaccinations (influenza, pneumococcal, shingles) before or during low-dose immunosuppression.<br><br>D. Monitoring<br>• Monthly CBC, liver profile, creatinine, uPCR; immunoglobulins and complements every 3–4 months.<br>• ESSDAI assessment each visit; ophthalmology and dental follow-up for sicca control.<br>• Repeat kidney biopsy only if proteinuria or renal function worsens despite therapy.<br>This regimen targets both the systemic autoimmune driver (pSS) and its renal manifestation (secondary MN), while minimising progression risk and drug toxicity. |
| Llama4-Marveric | Treatment Plan<br>**Initial Treatment:**<br>1. **Immunosuppressive Therapy:** Given the presence of significant proteinuria and active SS, immunosuppressive therapy is warranted. The choice of agent (e.g., corticosteroids, rituximab, or other immunosuppressants) depends on the severity of renal involvement and extraglandular manifestations.<br>2. **Supportive Care:** Continue losartan for its renoprotective effects. Monitor and manage hypertension and hyperlipidemia as necessary. |

**Rationale:**

• The significant reduction in proteinuria, improvement in albumin levels, and decrease in ESSDAI score post-treatment (Table 1) indicate a positive response to therapy.

• Immunosuppressive therapy is justified given the immune-mediated nature of SS and its renal involvement.

**Alternative Options:**

• **Rituximab:** For patients with severe or refractory SS, rituximab may be considered.

• **Other Immunosuppressants:** Depending on the patient's response and tolerance, other agents like cyclophosphamide or mycophenolate mofetil could be considered.

The treatment plan should be tailored to the patient's specific needs, with close monitoring of renal function, proteinuria, and disease activity.

**Phase III – Q8: Evaluation of treatment**

DeepSeek-R1

Treatment Effectiveness/Safety Monitoring and Risk Management

**Effectiveness Monitoring:**

• **Urine protein-to-creatinine ratio (UPCR):** Monthly for 3 months, then quarterly. Target: Maintain <0.5 g/g.cr (complete remission).

• **Serum albumin:** Monthly until normal range (40-55 g/L), then quarterly.

• **Renal function:** Serum creatinine monthly (target: 41-73 μmol/L), eGFR quarterly.

• **Immunoglobulins (IgG/IgA/IgM):** Quarterly to monitor immunosuppression depth (e.g., IgG <4 g/L requires intervention).

• **Sjögren's activity:** ESSDAI score biannually; sicca symptoms assessed quarterly.

**Safety Risks and Countermeasures:**

| Risk | Monitoring | Concrete Countermeasures |
|---|---|---|
| **Infections** | CBC, CRP monthly; fever/symptom diary | Immediate antibiotics for infections; avoid live vaccines; consider IVIG if recurrent. |
| **Hypogammaglobulinemia (IgG <4.54 g/L)** | Quarterly IgG levels | Hold telitacicept if IgG <4 g/L; IVIG replacement if infection-prone. |
| **Steroid toxicity** | Fasting glucose, bone density (DEXA) annually; BP weekly | Calcium/vitamin D supplementation; PPI for gastric protection; BP control with ARB. |
| **Nephrotic relapse** | UPCR every 2 weeks during infections | Aggressive infection control; temporary steroid boost during flares. |

Gemini 2.5 Pro

**Monitoring Treatment Effectiveness and Safety**

**Effectiveness Monitoring:**

The goal is to ensure sustained complete remission of both the renal and systemic disease.

• **Renal Remission:** Monitor Urine Protein-to-Creatinine Ratio (PCR), serum albumin, and serum creatinine every 3 months for the first year post-remission, then every 6 months if stable. The objective is to maintain a PCR of <0.3 g/g.cr and normal albumin/creatinine levels.

• **Systemic Sjögren's Syndrome (pSS) Remission:** At each visit, inquire about sicca symptoms (dry mouth, dry eyes) and other systemic manifestations (e.g., joint pain, fatigue). Maintain the European Sjögren's Syndrome Disease Activity Index (ESSDAI) score at 0.

**Safety Monitoring and Risk Management:**

The primary risk is infection due to profound immunosuppression, especially the significant hypogammaglobulinemia induced by telitacicept.

| | |
|---|---|
| | • **Risk: Severe Infection secondary to Hypogammaglobulinemia.**<br>• **Problem:** Telitacicept's mechanism (inhibiting B-lymphocyte stimulator [BLyS] and A Proliferation-Inducing Ligand [APRIL]) has led to very low levels of IgG, IgA, and IgM. This severely compromises humoral immunity, increasing the risk of bacterial and viral infections, as demonstrated by the patient's upper respiratory infection (URTI) which triggered a nephrotic flare.<br>• **Countermeasures:**<br>**a. Monitor Immunoglobulins:** Check IgG, IgA, and IgM levels every 3-6 months until they normalize.<br>**b. Patient Education:** Counsel the patient to seek immediate medical attention for any signs of infection (fever, cough, sore throat).<br>**c. Vaccinations:** Ensure she is up-to-date with non-live vaccines (e.g., influenza, pneumococcus, COVID-19).<br>**d. Consider IVIG:** If IgG levels remain critically low (e.g., <4.0 g/L) and/or she experiences recurrent serious infections, immunoglobulin replacement therapy (IVIG) should be considered.<br>•**Risk: Glucocorticoid (Steroid) Long-Term Effects.**<br>• **Problem:** Although discontinued, the course of steroids increases the risk of osteoporosis, diabetes, and cataracts.<br>• **Countermeasures:** Continue calcium/vitamin D. Monitor blood glucose and lipids. Schedule a baseline bone density (DEXA) scan and regular ophthalmology exams.<br>• **Risk: Losartan Side Effects.**<br>• **Problem:** Risk of hyperkalemia and acute kidney injury, especially during intercurrent illness with dehydration.<br>• **Countermeasures:** Monitor creatinine and potassium alongside PCR. Advise the patient to hold the losartan and seek advice if she develops vomiting or diarrhea. |
| OpenAI GPT-o3 | Monitoring plan and risk management<br>• Clinical review every 4–8 weeks for 1 year, then every 3–6 months: blood pressure, weight, oedema, sicca symptoms, infection signs.<br>• Laboratory panel at each visit:<br>− Urine protein (PCR/uPCR or dipstick) and sediment → relapse detection.<br>− Serum creatinine/eGFR, albumin → renal function & nephrotic status.<br>− Complete blood count, differential → cytopenia / infection risk.<br>− Immunoglobulins (IgG, IgA, IgM) → hypogammaglobulinaemia from telitacicept.<br>− Fasting glucose, lipids, liver enzymes, electrolytes → steroid, statin and RAAS-blocker effects.<br>• Infection surveillance: chest X-ray or CT and cultures when symptomatic; vaccinate (influenza, pneumococcal, zoster) during low-dose immunosuppression.<br>• Bone health: dual-energy X-ray absorptiometry at baseline and 12 months; maintain Ca/vit-D, consider bisphosphonate.<br>• Ophthalmology and dental assessment yearly for pSS control.<br>Specific risks & countermeasures<br>− Relapse of MN/pSS → early rise in uPCR or ESSDAI; respond by re-introducing / escalating immunosuppression.<br>− Severe infections (bacterial, viral, PJP) → low threshold for cultures, imaging, and empirical antibiotics; consider PJP prophylaxis if steroids restarted ≥20 mg/day.<br>− Hypogammaglobulinaemia (<4 g/L IgG + infections) → pause telitacicept, give IVIG replacement.<br>− Steroid toxicity (Cushingoid, diabetes, osteoporosis) → taper off, monitor glucose, bone protection.<br>− Telitacicept adverse effects (injection-site reaction, cytopenia) → symptomatic care, dose |

| | |
|---|---|
| | adjustment or discontinuation.<br>− Losartan-induced hyperkalaemia / renal dysfunction → check $K^+$ and eGFR each visit, adjust dose. |
| Llama4-Marveric | Monitoring Effectiveness and Safety of Treatment<br>**Effectiveness Monitoring:**<br>**1. Regular Urinalysis:** To assess proteinuria and hematuria.<br>**2. Serum Albumin and Creatinine Levels:** To monitor renal function and disease activity.<br>**3. Immunoglobulin Levels:** To assess the response to immunosuppressive therapy and potential over-suppression.<br>**4. ESSDAI Score:** To evaluate the activity of Sjögren's Syndrome.<br><br>**Safety Monitoring:**<br>**1. Infection Surveillance:** Regularly check for signs of infections, given the immunosuppressive therapy.<br>**2. Adverse Effects of Medications:** Monitor for side effects of corticosteroids (e.g., glucose intolerance, osteoporosis) and telitacicept (e.g., injection site reactions, potential infections).<br>**3. Blood Pressure Control:** Continue monitoring blood pressure to ensure it remains within target range.<br><br>**Countermeasures:**<br>• **Infection Management:** Promptly treat any infections with appropriate antibiotics.<br>• **Adjust Immunosuppression:** Taper or adjust immunosuppressive therapy based on disease activity and risk of infections or other adverse effects. |
| **Phase III – Q9: Formulating plans for worsening conditions** | |
| DeepSeek-R1 | Response to Unsatisfactory Progress/Sudden Changes<br>**Thought Process:**<br>**1. Rapid reassessment:**<br>• If UPCR ↑ or ALB ↓: Rule out infection (e.g., URTI-triggered relapse in Oct 2023), non-adherence, or drug resistance.<br>• If Cr ↑: Exclude acute kidney injury (e.g., volume depletion, interstitial nephritis from drugs).<br>• If hypogammaglobulinemia worsens (IgG <4 g/L): Evaluate for opportunistic infections.<br>**2. Diagnostic revision:**<br>• Biopsy repeat if atypical features emerge (e.g., new hematuria suggesting proliferative GN).<br>• Serology recheck: Anti-PLA2R, ANA to exclude evolving SLE.<br><br>**Action Plan:**<br>• **Immediate steps:**<br>• For infection: Start empiric antibiotics; hold immunosuppressants temporarily.<br>• For proteinuria flare: Increase methylprednisolone to 0.5 mg/kg/day; optimize ARB dosing.<br>• **Long-term adjustment:**<br>• Switch telitacicept to rituximab (375 mg/m² weekly ×4) if relapse persists.<br>• Add cyclophosphamide (1-2 mg/kg/day) if rituximab contraindicated.<br>• **Refractory disease:** Renal biopsy to assess for transformation (e.g., crescentic GN).<br><br>**Rationale:** Proactive infection control and immunoglobulin monitoring prevent relapses (as in Oct 2023). Flexible immunosuppression adjustment ensures sustained remission. |
| Gemini 2.5 Pro | **Response to Worsening or Unexpected Changes**<br>If the patient's condition deteriorates (e.g., recurrence of proteinuria, rise in creatinine), a |

| | |
|---|---|
| | rapid and systematic reassessment is crucial.<br><br>**Thought Process (Differential Diagnosis for a Flare):**<br>**i. Trigger-Induced Flare (High Probability):** The most likely cause is a transient immune activation due to an external trigger. The case history provides a perfect example: the URTI caused a temporary relapse into nephrotic syndrome that resolved after treating the infection. This is not a true disease relapse.<br>**ii. True Disease Relapse:** The underlying pSS has become active again, requiring re-initiation of immunosuppression. This would be suspected if proteinuria persists or worsens after any potential triggers are managed.<br>**iii. Treatment Non-Adherence:** Confirm the patient is consistently taking her losartan, as stopping it would increase proteinuria.<br>**iv. New Superimposed Disease:** Although unlikely, a new, unrelated kidney disease cannot be entirely ruled out.<br><br>**Action Plan:**<br>**v. Immediate Evaluation:**<br>• Take a thorough history focusing on infection symptoms (fever, cough), new medications, or vaccinations.<br>• Perform a physical exam to assess for signs of infection and volume status.<br>• Obtain urgent labs: urine PCR, urine sediment microscopy, serum creatinine, albumin, CBC, and inflammatory markers (CRP, ESR).<br>**vi. Manage Reversible Triggers:** If an infection is identified, treat it promptly and aggressively. Optimize supportive care (e.g., adjust losartan, manage blood pressure).<br>**vii. Decision on Immunosuppression:**<br>• **If proteinuria improves after treating the trigger:** This confirms a trigger-induced flare. No immediate change to immunosuppression is needed, but monitoring frequency should be increased temporarily.<br>• **If proteinuria persists or worsens:** This indicates a true disease relapse. The plan would be to:<br>• **Re-introduce Glucocorticoids:** Start a course of methylprednisolone or oral prednisolone to rapidly control the inflammation.<br>• **Re-evaluate Long-term Therapy:** A new long-term immunosuppressant would be required. Given the effectiveness but significant hypogammaglobulinemia with telitacicept, options would include a B-cell targeted therapy like Rituximab or an antimetabolite like Mycophenolate Mofetil (MMF), which are standard of care for pSS-associated glomerulonephritis.<br>• **Consider Repeat Biopsy:** If the relapse is severe or does not respond to first-line retreatment, a repeat renal biopsy would be warranted to assess for histologic transformation or increased chronicity, which would guide further therapy. |
| OpenAI GPT-o3 | Approach if the clinical course deviates<br>1. Rapid re-evaluation: repeat history, exam, full laboratory panel, renal ultrasound; look for triggers (infection, poor adherence, new drugs).<br>2. Clarify pattern:<br>− Isolated rise in proteinuria/ESSDAI → consider disease flare.<br>− Rising creatinine or active urine sediment → possible new glomerular lesion or interstitial nephritis.<br>− Fever, leucocytosis, imaging changes → infection.<br>3. Targeted tests: complement levels, anti-dsDNA, PLA2R, cryoglobulin, viral PCRs; consider repeat renal biopsy if diagnosis unclear or renal function deteriorates.<br>4. Therapeutic adjustment:<br>− Infection → suspend immunosuppressants, start appropriate antimicrobials.<br>− Autoimmune flare → reinstate or intensify therapy (methylprednisolone pulse, restart |

| | |
|---|---|
| | telitacicept, or switch to rituximab / mycophenolate).<br>− Drug toxicity → remove offending agent, supportively treat complications.<br>5. Multidisciplinary input (nephrology, rheumatology, infectious disease) and consider hospital admission for rapid intervention. |
| Llama4-Marveric | Managing Unexpected Changes or Lack of Improvement<br>**Thought Process:**<br>**1. Reassess Diagnosis:** Verify the initial diagnosis and consider alternative or additional diagnoses.<br>**2. Evaluate Treatment Adherence:** Check if the patient is adhering to the prescribed treatment regimen.<br>**3. Assess for Complications:** Look for complications related to the disease or treatment.<br><br>**Action Plan:**<br>**1. Modify Treatment:** Adjust the immunosuppressive regimen if there's inadequate response or significant side effects.<br>**2. Additional Investigations:** Consider further tests (e.g., repeat renal biopsy) if there's a significant change in the patient's condition or lack of expected improvement.<br>**3. Multidisciplinary Review:** Involve other specialists (e.g., rheumatologist, nephrologist) for a comprehensive review and advice on management.<br><br>By closely monitoring the patient's response to treatment and being prepared to adjust the management plan as needed, we can optimize outcomes and minimize risks. |

| **Case 4**: **Treatment of atypical hemolytic uremic syndrome with eculizumab in a patient presenting with neuropsychiatric prodrome: a case report** [5] | |
|---|---|
| **Phase I – Q1: Summarizing medical problems** | |
| DeepSeek-R1 | A 26–30-year-old Han Chinese woman presenting with **new-onset intermittent seizures** following an upper respiratory tract infection (URTI), suggesting an acute/subacute post-infectious inflammatory or autoimmune process affecting the CNS. |
| Gemini 2.5 Pro | A 26–30-year-old Han Chinese female with a previously unremarkable medical history presents with new-onset, intermittent seizures that developed acutely following a recent upper respiratory tract infection. This presentation suggests a post-infectious neurological syndrome, potentially as a manifestation of a newly unmasked systemic disease. |
| OpenAI GPT-o3 | 26–30-year-old Han Chinese woman with new-onset intermittent seizures occurring shortly after an upper respiratory tract infection (URI). |
| Llama4-Marveric | The patient is a 26–30-year-old Han Chinese female experiencing intermittent seizures that started after an upper respiratory tract infection. |
| **Phase I – Q2: Differential diagnoses and rationale** | |
| DeepSeek-R1 | Differential Diagnoses<br>**Prioritized list:**<br>**1. Autoimmune encephalitis** (e.g., anti-NMDA receptor encephalitis) – Most likely. Post-URTI trigger, seizures, and demographic (young female) align with autoimmune etiologies.<br>**2. Viral encephalitis** (e.g., HSV, enteroviruses) – Seizures post-URTI may reflect direct CNS infection.<br>**3. Acute disseminated encephalomyelitis (ADEM)** – Post-infectious demyelination can cause seizures.<br>**Must not miss:** Autoimmune encephalitis (rapidly progressive) or bacterial meningitis (though less likely without fever/stiff neck).<br>**Reasoning:** Seizures after URTI prioritize immune-mediated (autoimmune > viral) over structural causes. Autoimmune encephalitis is critical to rule out due to high morbidity if untreated. |
| Gemini 2.5 Pro | **Differential Diagnosis**<br>**• Differential Diagnoses:**<br>**i. Systemic Lupus Erythematosus (SLE) with Neuropsychiatric Manifestation** |

| | |
|---|---|
| | **(NPSLE):** An autoimmune disease with a high prevalence in young East Asian females. Infections are a common trigger for flares or initial presentation. Seizures are a known criterion for NPSLE.<br>**ii. Post-infectious Autoimmune Encephalitis (e.g., Anti-NMDA Receptor Encephalitis):** A well-described entity in young adults, often triggered by an infection, that presents with seizures and neuropsychiatric symptoms.<br>**iii. Posterior Reversible Encephalopathy Syndrome (PRES):** This can be caused by severe hypertension, which may result from an acute post-infectious glomerulonephritis. Seizures are a cardinal feature of PRES.<br><br>• **Most Likely Diagnosis: SLE with NPSLE.** The patient's specific demographic profile (young, female, Han Chinese) significantly increases the pre-test probability of SLE, and the post-infectious onset of seizures is a classic presentation.<br><br>• **Must-Not-Miss Diagnosis: Infectious Meningoencephalitis.** The preceding "URTI" could be a prodrome of a direct CNS infection (e.g., viral or bacterial). This is a medical emergency requiring immediate diagnosis and treatment to prevent severe morbidity or mortality. |
| OpenAI GPT-o3 | Differential diagnosis<br>• Post-infectious/viral encephalitis (most likely: temporal relation to URI, age, acute seizures).<br>• Autoimmune encephalitis (e.g., anti-NMDAR).<br>• Acute symptomatic seizures from metabolic derangement, intracranial infection or mass (must not miss: bacterial meningitis/encephalitis, HSV encephalitis, space-occupying lesion).<br>Rationale: seizures after a prodromal URI raise concern for CNS infection or post-infectious inflammatory processes; metabolic or structural causes are common alternatives; life-threatening infections must be excluded urgently. |
| Llama4-Marveric | Differential Diagnoses<br>**1. Viral Encephalitis:** Given the recent upper respiratory tract infection, viral encephalitis is a consideration. It can cause seizures among other neurological symptoms.<br>**2. Febrile Seizure or Infection-Related Seizure:** Although more common in children, seizures can occur in adults following infections.<br>**3. Autoimmune Encephalitis:** This condition can present with seizures and is associated with various autoantibodies. It can be triggered by infections.<br><br>The most likely diagnosis could be **Viral Encephalitis** or **Autoimmune Encephalitis**, given the neurological presentation following an infection.<br>**Must Not Be Missed:** Conditions like **Autoimmune Encephalitis** or other treatable causes of seizures should not be missed as they require prompt treatment. |
| **Phase I – Q3: Necessary physical examinations and reasons** | |
| DeepSeek-R1 | Key History/Physical Exam<br>**History questions:**<br>• **Seizure details:** Type (focal/generalized), duration, triggers, post-ictal state. *Why:* Differentiates epileptic syndromes vs. encephalitis-related seizures.<br>• **URTI specifics:** Timing, fever, rash, or sore throat. *Why:* Identifies pathogens (e.g., HSV, mycoplasma) linked to autoimmune encephalitis.<br>• **Neuropsychiatric symptoms:** Memory loss, psychosis, movement disorders. *Why:* Classic in autoimmune encephalitis (e.g., anti-NMDA).<br>• **Reproductive history:** Ovarian teratoma (paraneoplastic anti-NMDA risk). *Why:* Guides urgent tumor screening.<br><br>**Physical exam:**<br>• **Meningeal signs** (neck stiffness, Kernig's). *Why:* Rules out meningitis. |

| | |
|---|---|
| | • **Focal neurologic deficits.** *Why:* Suggests structural lesions.<br>• **Skin/joint exam** (rash, arthritis). *Why:* Excludes systemic autoimmunity (e.g., SLE). |
| Gemini 2.5 Pro | **Additional History and Physical Examination**<br>• **Additional History:**<br>• **Review of Systems for SLE: I**nquire about constitutional symptoms (fever, fatigue), skin changes (malar rash, photosensitivity, oral ulcers), hair loss, joint pain, and pleuritic chest pain. Ask about urine changes (foamy or dark urine) to screen for nephritis.<br>• **Neurological/Psychiatric Symptoms:** Ask about headache, visual disturbances, confusion between seizures, memory problems, or behavioral changes to differentiate between encephalitis, PRES, and NPSLE.<br>• **Infectious Symptoms:** Clarify the nature of the URTI and ask about persistent fever, headache, and neck stiffness to assess for meningitis.<br><br>• **Physical Examination:**<br>• **Vital Signs:** Critically assess **blood pressure** to rule out hypertensive emergency/PRES and **temperature** for infection or systemic inflammation.<br>• **Neurological Exam:** Perform a full assessment, including mental status, cranial nerves, and signs of meningeal irritation (e.g., nuchal rigidity), to identify focal deficits or signs of meningitis.<br>• **Systemic Exam:** Inspect the skin and mouth for rashes or ulcers, examine joints for synovitis, and auscultate the heart and lungs for serositis (all signs of SLE). Check for peripheral edema, which could indicate renal dysfunction or fluid overload. |
| OpenAI GPT-o3 | Targeted history / examination<br>History: fever pattern, headache, neck stiffness, altered mentation, focal deficits, photophobia (infection); recent psychosis, dyskinesia, autonomic instability (autoimmune encephalitis); past seizures, head trauma, family epilepsy (primary epilepsy); drug/alcohol use; medication/supplement history (metabolic/toxic).<br>Examination: vitals (fever, BP), meningismus, full neurologic exam (focal signs, cognitive status), skin (rash, petechiae—meningococcemia, HSV), fundus (papilledema). Purpose: differentiate infectious, autoimmune, structural, metabolic causes and gauge severity. |
| Llama4-Marveric | Additional Questions and Physical Exams<br>To verify the hypotheses:<br>• Ask about the nature of the seizures (focal or generalized), frequency, and any associated symptoms like fever, headache, or confusion.<br>• Inquire about the specifics of the upper respiratory tract infection, such as duration and severity.<br>• Perform a neurological examination to assess for focal neurological deficits, level of consciousness, and signs of meningeal irritation. |
| **Phase I – Q4: Plan for investigations/tests** | |
| DeepSeek-R1 | Diagnostic Testing Plan<br>**Tests and purposes:**<br>**1. Brain MRI with contrast:**<br>• Purpose: Detect inflammation (e.g., temporal lobe hyperintensity in HSV), demyelination (ADEM), or tumors.<br>• Expected results: Normal or limbic encephalitis patterns (autoimmune); demyelinating lesions (ADEM).<br>**2. Lumbar puncture (CSF analysis):**<br>• Purpose: Assess infection/inflammation. Include cell count, protein, glucose, PCR (HSV/VZV), autoimmune antibodies (anti-NMDA).<br>• Expected results: Lymphocytic pleocytosis ± elevated protein (encephalitis); anti-NMDA antibodies (autoimmune).<br>**3. EEG:** |

| | |
|---|---|
| | • Purpose: Evaluate seizure activity; classic patterns (e.g., extreme delta brush in anti-NMDA).<br>• Expected results: Seizure foci or encephalopathic changes.<br>**4. Blood tests (CBC, CRP, autoimmune serology):**<br>• Purpose: Screen for infection (leukocytosis) or systemic autoimmunity (ANA).<br>• Expected results: Elevated CRP (infection/inflammation); ANA positivity (SLE).<br>**Rationale:** MRI/EEG are non-invasive first steps; LP confirms inflammation/autoimmunity. Cost-effective and high-yield for urgent treatable causes. |
| Gemini 2.5 Pro | **Diagnostic Test Plan**<br>• **Initial Laboratory Tests:**<br>• **Purpose:** To screen for systemic disease, renal involvement, and metabolic derangements that can cause seizures.<br>• **Tests:**<br>• **Comprehensive Metabolic Panel (CMP):** To assess renal function (BUN, creatinine), electrolytes (Na, K, Ca), and glucose.<br>• **Complete Blood Count (CBC) with differential:** To look for cytopenias (anemia, leukopenia, thrombocytopenia) suggestive of SLE.<br>• **Urinalysis with urine microscopy:** Essential to detect proteinuria, hematuria, or cellular casts, which are signs of active glomerulonephritis.<br>• **Inflammatory Markers (ESR, CRP):** To quantify systemic inflammation.<br><br>• **Second-line and Confirmatory Tests:**<br>• **Purpose:** To confirm a specific diagnosis based on initial findings.<br>• **Tests:**<br>• **Brain MRI:** To visualize the brain parenchyma. This is crucial for identifying findings of encephalitis, vasculitis, or the characteristic posterior edema of PRES.<br>• **Lumbar Puncture:** If there is any suspicion of CNS infection or inflammation, to analyze cerebrospinal fluid (CSF) for cell count, protein, glucose, cultures, and specific viral PCRs.<br>• **Immunologic Studies:** Order an Antinuclear Antibody (ANA) as a screen for SLE. If positive, follow with more specific tests like anti-dsDNA, anti-Sm, and complement levels (C3/C4). If autoimmune encephalitis is suspected, a specific antibody panel (e.g., anti-NMDAR) on serum and CSF is indicated.<br>• **Electroencephalogram (EEG):** To characterize seizure activity and identify any underlying encephalopathic process. |
| OpenAI GPT-o3 | Investigations<br>• Basic labs (CBC, electrolytes, glucose, Ca/Mg, renal & liver panels): look for metabolic precipitants and inflammatory markers.<br>• Brain MRI with contrast: detect encephalitic changes, mass lesions.<br>• Lumbar puncture with CSF analysis (cell count, protein, glucose, Gram stain/culture, HSV PCR, other viral PCR): confirm/exclude bacterial or viral encephalitis.<br>• EEG: characterize seizure type, look for encephalitic patterns.<br>• Autoimmune encephalitis panel (CSF/serum anti-NMDAR, LGI1, etc.): if infection excluded.<br>Expected: inflammatory CSF (lymphocytic pleocytosis) and temporal lobe MRI changes support viral/autoimmune encephalitis; normal CSF with abnormal EEG/MRI may suggest other etiologies; gross abnormalities on MRI would point to structural lesion. |
| Llama4-Marveric | Tests to Narrow Down the Diagnosis<br>**1. Lumbar Puncture (LP) and CSF Analysis:** To check for signs of infection or inflammation in the CSF, which can help diagnose viral or autoimmune encephalitis.<br>**2. EEG:** To assess for seizure activity or abnormalities suggestive of encephalitis. |

| | |
|---|---|
| | **3. MRI Brain:** To look for structural abnormalities or inflammation in the brain.<br>**4. Autoantibody Tests:** To check for antibodies associated with autoimmune encephalitis.<br><br>These tests will help differentiate between potential causes of the seizures, such as viral encephalitis versus autoimmune encephalitis, and guide further management. |
| **Phase II – Q5: Interpreting test results** | |
| DeepSeek-R1 | Interpretation of Key Test Results<br>• **Hemolytic anemia markers:** Elevated LDH (429 U/L), reticulocytosis (8.26%), low haptoglobin (<25 mg/dL), and schistocytes on blood smear confirm **intravascular hemolysis.** Negative Coombs test and Ham test rule out immune-mediated hemolysis or PNH, favoring microangiopathic hemolysis.<br>• **Renal dysfunction:** Proteinuria (3+), hematuria (3+), and symmetric edema indicate **glomerular injury,** likely thrombotic microangiopathy (TMA).<br>• **CSF abnormalities:** Elevated protein (15.74 g/L) and pleocytosis ($204 \times 10^6$/L WBCs) suggest CNS inflammation, but the absence of infectious/autoimmune markers (negative ANA, ANCA, tumor markers) points to **TMA-related endothelial damage** rather than classic encephalitis.<br>• **Platelet clumping/schistocytes (1%):** Critical for TMA diagnosis, indicating mechanical RBC fragmentation from microvascular thrombosis.<br><br>**Contextual significance:** The triad of neurological symptoms (seizures), MAHA (microangiopathic hemolytic anemia), and renal injury—triggered by URTI—aligns with **thrombotic microangiopathy**, where systemic endothelial damage explains all features. |
| Gemini 2.5 Pro | Interpretation of Test Results<br>The patient's test results collectively point towards a systemic process causing multi-organ injury, specifically a **Thrombotic Microangiopathy (TMA).**<br>• **Evidence of Microangiopathic Hemolytic Anemia (MAHA):** The presence of schistocytes (fragmented red blood cells) on the smear is the morphological hallmark of MAHA. This indicates that red blood cells are being mechanically sheared and destroyed within the small blood vessels. This is strongly supported by laboratory findings of hemolysis: elevated LDH (a marker of cell turnover), markedly low haptoglobin (consumed by binding free hemoglobin), and a powerful compensatory bone marrow response (very high reticulocyte count). The negative Coombs test is critical, as it effectively rules out common autoimmune hemolytic anemia and directs the diagnosis toward a TMA.<br>• **Evidence of Renal and Neurological Injury:** The symmetric pitting edema, 3+ proteinuria, and 3+ occult blood (likely hemoglobinuria from intravascular hemolysis) confirm significant, acute kidney damage. Concurrently, the MRI abnormalities and the highly inflammatory cerebrospinal fluid (extremely high protein and elevated white cells) explain the seizures and confirm severe central nervous system involvement.<br>• **Significance of Negative Serologies:** The negative ANA and ANCA results make common secondary causes of TMA, such as Systemic Lupus Erythematosus (SLE) or ANCA-associated vasculitis, less probable, increasing the likelihood of a primary TMA syndrome.<br><br>In summary, the initial "encephalitis" is now understood not as an isolated CNS infection, but as a neurological manifestation of a systemic TMA that is also causing hemolytic anemia and acute renal dysfunction. |
| OpenAI GPT-o3 | Interpretation of the new data<br>• CSF: marked protein elevation (15.7 g/L) and pleocytosis (204 /µL) confirm CNS inflammation but are nonspecific; in the context of microangiopathy they may reflect small-vessel injury rather than primary viral encephalitis.<br>• Hemolysis profile: ↑LDH, very high reticulocyte indices, undetectable haptoglobin and schistocytes on smear all indicate active microangiopathic hemolytic anemia (MAHA). |

| | |
|---|---|
| | Negative DAT and negative acidified-serum test rule out autoimmune hemolysis and PNH, respectively.<br>• Renal findings: nephrotic-range proteinuria (3 +), hematuria (3 +), lower-limb edema point to glomerular injury; together with MAHA they suggest a thrombotic microangiopathy (TMA) affecting the kidney.<br>• Platelet clumping on smear implies thrombocytopenia (quantitative count not supplied) accompanying MAHA—another hallmark of TMA.<br>• Neurological abnormalities on MRI and seizures can be explained by cerebral microthrombi/ischemia typical of TMA rather than infectious encephalitis.<br><br>Key message: the constellation of MAHA, (probable) thrombocytopenia, renal impairment and neurologic signs forms the classic TMA tetrad, redirecting the problem away from primary encephalitis. |
| Llama4-Marveric | The patient presents with a complex clinical picture involving neurological symptoms, hemolytic anemia, and renal dysfunction. Key findings include elevated CSF protein and white blood cell count suggesting encephalitis, hemolytic anemia indicated by elevated LDH, reticulocyte count, and low haptoglobin, and renal involvement shown by urinary protein and occult blood. The presence of schistocytes in the bone marrow aspiration smear and peripheral blood suggests microangiopathic hemolytic anemia (MAHA). The negative direct anti-human globulin test rules out autoimmune hemolytic anemia. The overall picture is suggestive of a systemic condition affecting multiple organ systems. |
| **Phase II – Q6: Reconsidering the differential diagnosis** | |
| DeepSeek-R1 | Revised Diagnostic Thinking<br>**Initial focus:** Encephalitis (due to seizures + CSF abnormalities).<br>Updated priority:<br>**1. Thrombotic thrombocytopenic purpura (TTP)** – Most likely. URTI-triggered ADAMTS13 deficiency causing microvascular thrombosis, explaining seizures (CNS ischemia), MAHA, renal injury, and CSF inflammation.<br>**2. Atypical hemolytic uremic syndrome (aHUS)** – Complement dysregulation could cause identical TMA.<br>**3. Secondary TMA** (e.g., infection-induced).<br>**Exclusions:** Autoimmune/rheumatological causes (negative ANA/ANCA), PNH (negative Ham test), or malignancy.<br>**Reasoning:** The TMA triad (MAHA + renal injury + neurological symptoms) overrides isolated encephalitis, as it unifies all findings. TTP is prioritized due to acute post-infectious onset and demographic (young female). |
| Gemini 2.5 Pro | Revised Diagnostic Thinking and Working Diagnosis<br>The new information fundamentally reframes the diagnostic landscape. The initial diagnosis of encephalitis is now re-contextualized as a component of a much broader systemic disease. The core clinical syndrome is a **Thrombotic Microangiopathy (TMA)**, characterized by the triad of MAHA, renal dysfunction, and neurological symptoms.<br><br>My differential diagnosis is now focused on the primary causes of TMA in adults:<br>**1. Thrombotic Thrombocytopenic Purpura (TTP):** Caused by a severe deficiency of the ADAMTS13 enzyme. Given the prominent neurological symptoms in this adult patient, TTP is the most urgent consideration.<br>**2. Complement-Mediated HUS (aHUS):** Caused by dysregulation of the alternative complement pathway. It classically presents with a triad of MAHA, thrombocytopenia, and acute kidney injury, but neurological involvement is also common.<br><br>While secondary causes (e.g., drug-induced, infection-related) remain possible, the acuity and severity of this presentation in the absence of other clues make primary TMAs the top priority.<br>**Working Diagnosis: Thrombotic Microangiopathy (TMA), with the highest suspicion for Thrombotic Thrombocytopenic Purpura (TTP).** TTP is a medical emergency with a very high mortality rate if left untreated, and its treatment must be initiated empirically |

| | |
|---|---|
| | without delay. |
| OpenAI GPT-o3 | Updated diagnostic reasoning<br>Highest priority: acquired Thrombotic Thrombocytopenic Purpura (TTP)<br>– Fits age, sex, post-infectious trigger, prominent neurologic symptoms, MAHA and likely thrombocytopenia.<br>– Needs urgent plasma exchange; diagnosis later confirmed by severely reduced ADAMTS13 activity (<10%) or inhibitor.<br><br>Second possibility: Complement-mediated atypical Hemolytic Uremic Syndrome (aHUS)<br>– Also causes MAHA, renal dysfunction and neurologic features; more renal-dominant but overlaps with TTP.<br>– Consider if ADAMTS13 activity is >10 % or no rapid response to plasma exchange.<br>Less likely: typical (Shiga-toxin) HUS (no diarrhea), secondary TMA (malignant HT, drugs, pregnancy—none documented).<br><br>Working Diagnosis: Acquired TTP (until ADAMTS13 results clarify; manage as TTP/aHUS spectrum TMA). |
| Llama4-Marveric | Given the combination of encephalitis, MAHA, and renal dysfunction, the differential diagnosis can be refined. The presence of MAHA and renal involvement points towards a diagnosis of **Thrombotic Thrombocytopenic Purpura (TTP) or Hemolytic Uremic Syndrome (HUS)**. TTP is characterized by the pentad of MAHA, thrombocytopenia, renal failure, neurological symptoms, and fever, although not all elements are always present. The presence of schistocytes and low haptoglobin supports MAHA. Neurological symptoms can be part of TTP. **HUS** is another possibility, typically presenting with MAHA, renal failure, and often associated with infections. However, the neurological symptoms are more prominent in this case, leaning towards TTP. |
| **Phase II – Q7: Treatment planning** | |
| DeepSeek-R1 | Treatment Plan<br>**Immediate intervention:**<br>• **Therapeutic plasma exchange (TPE)** daily.<br>Rationale: First-line for suspected TTP (EBM: >90% survival if initiated <24h). Removes autoantibodies against ADAMTS13 and replenishes the enzyme.<br>• **Corticosteroids:** Methylprednisolone 1g IV × 3 days.<br>Rationale: Suppresses autoantibody production (EBM: Synergistic with TPE).<br>**Supportive care:**<br>• **Seizure control:** Levetiracetam (renal-adjusted dose).<br>• **Renal protection:** Strict fluid/electrolyte balance; avoid nephrotoxins.<br>• **Monitoring:** Daily platelets, LDH, haptoglobin, creatinine.<br>**Alternatives/Next steps:**<br>• **If TPE unavailable:** Fresh frozen plasma (FFP) infusion (bridge to TPE).<br>• **Refractory TTP:** Rituximab (anti-CD20) or caplacizumab (anti-vWF).<br>• **If aHUS confirmed:** Eculizumab (complement inhibitor).<br>• **ADAMTS13 activity testing:** To confirm TTP (activity <10%) vs. aHUS.<br>**Rationale for urgency:** Untreated TTP has >90% mortality; renal/neurological damage may be irreversible. TPE targets the core pathophysiology (microvascular thrombosis). |
| Gemini 2.5 Pro | Treatment Plan<br>This is a hematologic and neurologic emergency requiring immediate intervention while awaiting definitive diagnostic results.<br><br>**Immediate Treatment Plan:**<br>**1. Urgent Plasma Exchange (PEX):** This is the cornerstone and life-saving treatment for suspected TTP. It must be initiated immediately.<br>• **Rationale:** PEX serves two functions: it removes the patient's plasma, which is presumed |

| | |
|---|---|
| | to contain autoantibodies against ADAMTS13 and pathologic von Willebrand factor multimers, and it replenishes the plasma with functional ADAMTS13 from donor plasma.<br>**2. High-Dose Corticosteroids:** Administer intravenous corticosteroids (e.g., methylprednisolone 1 mg/kg/day) concurrently with PEX.<br>• Rationale: In immune-mediated TTP, steroids suppress the underlying autoimmune process and inhibit the production of autoantibodies against ADAMTS13.<br>**3. Caplacizumab:** If available, this anti-vWF nanobody should be strongly considered as an adjunct to PEX and steroids.<br>• **Rationale (EBM):** Caplacizumab rapidly inhibits platelet adhesion to vWF, halting microthrombus formation more quickly than PEX alone. Clinical trials have shown it accelerates platelet count recovery and reduces the risk of major thromboembolic events and death.<br><br>**Crucial Diagnostic Step:**<br>Before initiating PEX, an urgent blood sample must be drawn and sent for **ADAMTS13 activity level testing.** This is essential to confirm TTP (activity <10%) and guide long-term management.<br>Supportive Care:<br>• **Avoid platelet transfusions,** as they can worsen thrombosis, unless there is life-threatening bleeding.<br>• Manage seizures with anti-epileptic medications.<br>• Provide aggressive supportive care, including blood pressure management and renal replacement therapy (dialysis) if acute kidney injury becomes severe.<br><br>**Alternative Considerations and Future Plan:**<br>• If the ADAMTS13 activity level returns as normal (>10%), the diagnosis shifts to **aHUS**. In this scenario, PEX would be discontinued, and treatment with a C5 complement inhibitor (**Eculizumab or Ravulizumab**) would be initiated. These drugs are the specific, evidence-based therapy for aHUS.<br>• For refractory or relapsing TTP, **Rituximab** (an anti-CD20 monoclonal antibody) may be added to provide more durable immunosuppression. |
| OpenAI GPT-o3 | Treatment plan and rationale<br>Immediate (within hours)<br>Daily therapeutic plasma exchange (TPE) with fresh-frozen plasma replacement<br>− Standard of care for TTP; removes anti-ADAMTS13 antibodies and supplies functional ADAMTS13.<br>High-dose corticosteroids (e.g., methylprednisolone 1 mg/kg/day or pulse 1 g/day x3)<br>− Dampen autoantibody production; evidence supports combination with TPE.<br>Seizure control and neuro-monitoring (levetiracetam preferred; no hepatic metabolism issues).<br>Renal support: strict fluid balance, consider temporary hemodialysis if oliguric or rising creatinine.<br>Avoid platelet transfusion unless life-threatening bleeding.<br><br>Adjunct / conditional<br>• Rituximab early (375 mg/m$^2$ weekly ×4) if severe disease, delayed response or high inhibitor titer—improves remission rate (Grade B evidence).<br>• Caplacizumab (anti-vWF nanobody) if available: speeds platelet recovery, lowers composite morbidity; recommended by recent guidelines.<br>• If ADAMTS13 activity >10 % or poor response to 3–5 days of TPE ⇒ switch focus to aHUS and start eculizumab/ravulizumab after meningococcal prophylaxis.<br>• Supportive: folic acid, PPI prophylaxis, VTE prophylaxis when platelet count >50 $\times 10^9$/L. |

| | |
|---|---|
| | Monitoring<br>Daily CBC, LDH, creatinine, urine output/protein; neurologic status. Continue TPE until platelet count and LDH normalize for ≥2 days.<br><br>Alternative options debated but not first-line<br>• Cyclophosphamide or splenectomy (historical).<br>• Plasma infusion alone—insufficient for acquired TTP.<br><br>This strategy addresses both the presumed immune-mediated pathogenesis and the vital organs already affected (brain, kidney), while allowing rapid pivot to aHUS-directed therapy if ADAMTS13 testing indicates. |
| Llama4-Marveric | The treatment plan should focus on managing TTP or HUS. For **TTP**, the mainstay of treatment is **plasmapheresis**, which removes autoantibodies and replaces the deficient ADAMTS13 enzyme. Corticosteroids may also be used to suppress antibody production. For **HUS**, treatment is mainly supportive, focusing on managing renal failure and hypertension, although some cases may benefit from plasmapheresis. Monitoring of renal function, hematological parameters, and neurological status is crucial. Given the severity of the presentation, intensive care unit (ICU) admission may be necessary for close monitoring and management.<br>Rationale: The choice of treatment is based on the likely diagnosis of TTP or HUS, conditions that require prompt and specific interventions to prevent further organ damage and improve outcomes.<br><br>Alternative options include further diagnostic tests to confirm TTP or HUS, such as ADAMTS13 activity levels, and managing complications arising from the disease or its treatment. |
| **Phase III – Q8: Evaluation of treatment** | |
| DeepSeek-R1 | Monitoring Effectiveness and Safety<br>**Effectiveness monitoring:**<br>• **Hematological:**<br>• Weekly CBC (platelets, hemoglobin), LDH, haptoglobin, reticulocyte count.<br>Target: Platelets >150×$10^9$/L, LDH normalization, haptoglobin >30 mg/dL.<br>• **Renal:**<br>• Daily urine output, serum creatinine, BUN; weekly urinalysis (protein/blood).<br>*Target:* Recovery from anuria, reduced proteinuria.<br>• **Neurological:**<br>• Biweekly neurological exams; repeat brain MRI at 1 month to assess PRES resolution.<br>• **Complement blockade:**<br>• CH50 assay monthly (undetectable activity confirms eculizumab efficacy).<br><br>**Safety risks and countermeasures:**<br>**1. Meningococcal infection (eculizumab):**<br>• *Risk:* Life-threatening sepsis/meningitis (due to terminal complement inhibition).<br>• *Countermeasures:*<br>• Mandatory quadrivalent meningococcal vaccination + serogroup B vaccine (if not given).<br>• Prophylactic antibiotics (e.g., penicillin V 250 mg BID) until 2 weeks post-vaccination.<br>• Educate patient on fever/headache symptoms; provide emergency antibiotic (ceftriaxone) for immediate use.<br>**2. Infusion reactions (eculizumab):**<br>• *Risk:* Anaphylaxis, hypotension. |

| | |
|---|---|
| | • *Countermeasures:* Premedicate with antihistamine + corticosteroid; monitor vitals during infusion.<br>**3. Hypertensive crisis:**<br>• *Risk:* PRES recurrence or renal injury.<br>• *Countermeasures:* Daily BP checks; target <130/80 mmHg with ACEi/CCB. Avoid fluid overload during dialysis.<br>**4. Dialysis complications:**<br>• *Risk:* Hypotension, access thrombosis.<br>• *Countermeasures:* Adjust ultrafiltration rate; monitor anticoagulation (APTT). |
| Gemini 2.5 Pro | Treatment Monitoring and Risk Management<br>**Monitoring Treatment Effectiveness:**<br>The effectiveness of eculizumab and supportive care will be monitored by tracking the resolution of TMA activity across multiple organ systems.<br>• **Hematologic Monitoring:**<br>• **Primary Markers:** Daily monitoring of platelet count and LDH levels initially. Normalization of the platelet count and a sustained decrease in LDH indicate suppression of microangiopathic hemolysis.<br>• **Secondary Markers:** Weekly monitoring of haptoglobin (should rise to normal), hemoglobin (should stabilize and increase), and peripheral blood smear (schistocytes should disappear).<br>• **Renal Monitoring:**<br>• **Primary Marker:** Daily monitoring of urine output. The return of urine production is the most critical sign of potential renal recovery.<br>• **Secondary Markers:** Serum creatinine levels will be managed by hemodialysis, but any improvement independent of dialysis indicates recovering kidney function.<br>• **Neurological Monitoring:**<br>• Regular neurological examinations to ensure no recurrence of seizures or encephalopathy. Repeat brain MRI is warranted only if new neurological symptoms develop.<br><br>**Safety Monitoring, Risks, and Countermeasures:**<br>The primary risk of eculizumab is life-threatening infection due to terminal complement blockade.<br>• **Primary Risk: Meningococcal Infection (Neisseria meningitidis)**<br>• **Risk:** Eculizumab blocks the formation of the membrane attack complex (C5b-9), which is the body's primary defense against encapsulated bacteria, especially Neisseria meningitidis. This creates a high risk of fulminant meningitis or meningococcemia.<br>• **Countermeasures:**<br>**a. Vaccination:** The patient must receive quadrivalent (A, C, W, Y) and serogroup B meningococcal vaccines. In this emergency setting, vaccination should occur concurrently with the initiation of treatment.<br>**b. Antibiotic Prophylaxis:** Prophylactic antibiotics (e.g., penicillin V or a macrolide) must be prescribed for the entire duration of eculizumab therapy, as vaccination does not provide complete protection.<br>**c. Patient Education:** The patient and family must be rigorously educated to recognize early signs of meningitis (fever, headache, neck stiffness, rash) and to seek immediate emergency care if any of these symptoms appear. They should be provided with a medical alert card.<br><br>• **Other Risks:**<br>• **Infusion-Related Reactions:** Monitor for hypersensitivity during and after infusion. Have antihistamines and corticosteroids available to manage potential reactions. |

| | |
|---|---|
| | • **Hemodialysis Access Complications:** Monitor the dialysis catheter site for signs of infection (redness, pain, fever). Employ strict aseptic technique during access and care to prevent catheter-related bloodstream infections. |
| OpenAI GPT-o3 | Monitoring plan – effectiveness, risks, countermeasures<br>• Hematologic response<br>– Daily platelet count, hemoglobin, schistocyte % and LDH; goal: platelets > 150 ×10⁹/L and LDH normalization for ≥48 h.<br>• Renal response<br>– Hourly urine output, daily serum creatinine, BUN, $K^+$, acid–base status, weight and edema score; CRRT dose/clearance records; target: rising diuresis or fall in creatinine/BUN, stable electrolytes.<br>• Neurologic response – GCS, focal signs, seizure log, fundoscopy; repeat brain MRI/CT if new deficit.<br>• Blood pressure and volume status – Invasive BP or q15 min during crises; CVP/echo-guided fluid assessment; maintain SBP < 140 mmHg with IV nicardipine/labetalol.<br>• ADAMTS-13 activity & inhibitor titer (once available) and complement profile to guide further therapy. |
| Llama4-Marveric | To monitor the effectiveness and safety of the treatment for aHUS with eculizumab, plasma exchange, and hemodialysis:<br>**1. Hematological Parameters:** Regularly check for improvement in anemia and hemolysis indicators (e.g., LDH, haptoglobin, reticulocyte count).<br>**2. Renal Function:** Monitor urine output, serum creatinine, and electrolytes to assess the need for continued hemodialysis.<br>**3. Blood Pressure:** Closely monitor blood pressure to manage malignant hypertension.<br>**4. Neurological Status:** Regular neurological assessments to monitor for improvement or deterioration in neuropsychiatric symptoms and PRES lesions on MRI.<br>**5. Eculizumab Levels and Complement Activity:** Ensure adequate eculizumab levels and complement inhibition to verify the effectiveness of eculizumab therapy.<br>**6. Infection Surveillance:** Monitor for signs of infection, as eculizumab increases the risk of meningococcal infections. Ensure the patient is vaccinated against meningococcus and consider prophylactic antibiotics if necessary. |
| **Phase III – Q9: Formulating plans for worsening conditions** | |
| DeepSeek-R1 | Response to Unexpected Changes<br>**Thought process:**<br>**1. Rule out treatment failure:**<br>• If hemolysis persists (↑LDH, ↓platelets):<br>• Check eculizumab levels/complement activity (CH50).<br>• Consider eculizumab underdosing or neutralizing antibodies → increase dose/frequency (e.g., 1200 mg weekly).<br>• If renal function worsens:<br>• Assess for new TMA activity (schistocytes, haptoglobin) vs. dialysis complications → kidney ultrasound to exclude obstruction.<br>**2. Reevaluate diagnosis:**<br>• If neurological symptoms recur:<br>• Repeat ADAMTS13 (exclude late-onset TTP), CSF analysis, and MRI (new lesions vs. PRES relapse).<br>• If infection suspected:<br>• Blood/CSF cultures, NGS for pathogens → stop eculizumab temporarily if sepsis confirmed.<br>**3. Assess complications:**<br>• Malignant hypertension: Urgent nitroprusside/labetalol; exclude renal artery stenosis.<br>• Anuria unresponsive to dialysis: Consider cortical necrosis (MRI kidney); plan long-term |

| | |
|---|---|
| | RRT.<br><br>**Action plan:**<br>• **Immediate:**<br>• Hospitalize; repeat TMA labs (CBC, LDH, haptoglobin), ADAMTS13, complement studies.<br>• Escalate support (e.g., intensify plasma exchange if TMA rebounds).<br>• **Diagnostic refinement:**<br>• Genetic testing for complement regulators (CFH, CFI, MCP mutations) if aHUS etiology unclear.<br>• Kidney biopsy if new renal deterioration.<br>• **Therapeutic adjustment:**<br>• Switch to ravulizumab (long-acting anti-C5) if compliance issue.<br>• Add immunosuppressants (e.g., rituximab) for complement-antibody-mediated aHUS. |
| Gemini 2.5 Pro | Management of Unexpected Clinical Deterioration<br>If the patient's condition fails to improve or suddenly worsens, a rapid and systematic re-evaluation is critical.<br>**Thought Process:**<br>The immediate priority is to stabilize the patient. The subsequent thought process involves questioning the two fundamental pillars of the management plan: the diagnosis and the treatment.<br>**1. Is the diagnosis of aHUS correct?** aHUS is a diagnosis of exclusion. I would reconsider:<br>• **Atypical TTP:** Could this be a rare TTP variant with ADAMTS13 activity >10%?<br>• **Unidentified Secondary Cause:** Was an occult malignancy, a different autoimmune disorder, or a drug trigger missed? The initial workup was thorough, but a persistent lack of response necessitates re-investigation.<br>• **Catastrophic Antiphospholipid Syndrome (CAPS):** While initial serologies were negative, this can mimic aHUS.<br>**2. Is the treatment for aHUS adequate?**<br>• **Eculizumab Efficacy:** Is there inadequate complement blockade? This can be assessed by measuring a CH50 level; if it is not fully suppressed, the dose or frequency of eculizumab may need to be increased.<br>• **Concurrent Process:** Is there a new, superimposed problem masking the treatment effect? This could include a severe infection (e.g., catheter sepsis), a new thrombotic event, or a complication from a procedure.<br>**3. Is this the expected course of severe disease?** The extensive initial damage, particularly the anuria and severe kidney pathology (no clear glomeruli), may mean that renal recovery is not possible, even with effective systemic disease control. The goal would then shift to preventing extra-renal relapse while continuing chronic dialysis.<br><br>**Action Plan:**<br>**1. Immediate Stabilization & Assessment:**<br>• Address life-threatening issues (e.g., uncontrolled hypertension, respiratory compromise).<br>• Perform a rapid, targeted physical and neurological examination.<br>• Obtain urgent labs: CBC with smear, LDH, haptoglobin, platelets (to assess TMA activity), and blood cultures/inflammatory markers (to rule out sepsis).<br>**2. Diagnostic Re-evaluation:**<br>• Measure CH50 to confirm adequate complement blockade.<br>• Consider repeat imaging (e.g., Head CT/MRI, CT abdomen) based on new symptoms.<br>• If not already performed, proceed with genetic testing for mutations in complement- |

| | |
|---|---|
| | regulatory genes to definitively confirm the aHUS diagnosis.<br>**3. Treatment Modification:**<br>• If complement blockade is incomplete, increase the eculizumab dose.<br>• If sepsis is identified, start broad-spectrum antibiotics immediately.<br>• If the diagnosis is genuinely in doubt, consider re-initiating plasma exchange as a broad-acting therapy while further diagnostic clarification is sought. |
| OpenAI GPT-o3 | Major risks and countermeasures<br>1. Plasma-exchange related<br>Hypotension, allergic reaction, citrate-induced hypocalcemia, bleeding from catheter.<br>→ Pre-procedure $Ca^{2+}$ infusion, close vitals, emergency meds ready, meticulous line care, avoid platelet transfusion unless life-threatening bleeding.<br>2. CRRT related<br>Hypotension, electrolyte shifts, filter clotting, infection.<br>→ Continuous hemodynamic and electrolyte checks, regional citrate or heparin anticoagulation with anti-Xa monitoring, strict asepsis.<br>3. Immunosuppression (steroids ± rituximab)<br>Hyperglycaemia, infection, GI bleed.<br>→ Daily glucose, early antibiotics for fever, PPI prophylaxis, CMV/HBV screen if rituximab added.<br>4. Malignant hypertension complications (ICH, LV failure).<br>→ Gradual BP reduction (<25 % in first hour), serial neuro exams, bedside echo, repeat head CT if sudden neuro change.<br>5. Approach if no improvement or sudden deterioration<br>**Step 1** – Immediate stabilisation<br>ABC, secure airway, correct life-threatreatening electrolyte or BP derangements, call rapid response/ICU.<br>**Step 2** – Rapid reassessment<br>• Repeat CBC, LDH, coagulation, CMP, ABG, troponin.<br>• Urgent head CT/MRI for new neurologic signs.<br>• Bedside ECHO & chest X-ray for pulmonary oedema or cardiac dysfunction.<br>• Blood/urine cultures if febrile or septic picture.<br>**Step 3** – Re-evaluate TMA subtype and treatment adequacy<br>• Check ADAMTS-13 result:<br>– <10 % → intensify TPE (twice daily), add caplacizumab and rituximab.<br>– >10 % or inadequate response after 5–7 days → suspect atypical HUS; start eculizumab/ravulizumab after meningococcal prophylaxis.<br>• Review drug exposures, infections, malignancy as secondary TMA triggers.<br>**Step 4** – Escalate organ support<br>• Increase CRRT dose or switch to intermittent HD for severe hyperkalaemia/volume overload.<br>• Consider mechanical ventilation for ARDS or refractory seizures.<br>**Step 5** – Multidisciplinary input<br>Hematology, nephrology, neurology, intensive care, and possibly transplant team (early discussion of renal prognosis).<br>This cycle of prompt stabilisation, data-driven reassessment, and therapy modification is repeated until sustained hematologic and organ recovery is achieved. |
| Llama4-Marveric | Managing Potential Complications<br>**1. Infections:** Be vigilant for signs of infection, particularly meningococcal infection, given the patient's eculizumab treatment.<br>**2. Infusion Reactions:** Monitor for reactions to eculizumab or plasma exchange.<br>**3. Dialysis-Related Complications:** Watch for complications related to hemodialysis, such as vascular access issues, electrolyte imbalances, and hypotension.<br><br>Action Plan for Unexpected Changes<br>If the patient's condition does not improve as expected or suddenly changes: |

| | |
|---|---|
| | **1. Reassess Diagnosis:** Reconsider the diagnosis of aHUS and rule out other causes of TMA.<br>**2. Review Treatment Adherence and Effectiveness:** Check eculizumab levels, assess complement activity, and verify that plasma exchange and hemodialysis are being performed as prescribed.<br>**3. Investigate for Complications:** Look for signs of infection, infusion reactions, or dialysis-related complications.<br>**4. Adjust Treatment Plan:** Based on the findings, adjust the treatment plan. This may involve changing the dose or frequency of eculizumab, modifying plasma exchange or hemodialysis, or adding other treatments.<br><br>By closely monitoring the patient's response to treatment and being prepared to adjust the management plan as needed, it's possible to optimize outcomes for this complex condition. |

| **Case 5: New Diagnosis of Lupus Nephritis in the Third Trimester-A Case Report [6]** | |
|---|---|
| **Phase I – Q1: Summarizing medical problems** | |
| DeepSeek-R1 | A 30s-year-old Indigenous Australian pregnant female at 30 weeks gestation, presenting with **acute-onset (1 month) severe bilateral lower limb pitting edema, new malar rash, and erythematous maculopapular rash** on shoulders/chest. Background includes prior positive ANA and SS-A antibodies (2017, untreated), and two uncomplicated pregnancies. |
| Gemini 2.5 Pro | This is a 30-something Indigenous female at 30 weeks of gestation presenting with a one-month history of new-onset severe lower limb pitting edema, a malar rash, and a truncal rash. Her presentation is concerning for a new systemic autoimmune disease flare in the context of a known history of positive ANA and anti-SSA antibodies, which needs to be distinguished from a primary pregnancy-related complication. |
| OpenAI GPT-o3 | Indigenous woman, early 30s, at 30 weeks' gestation, with one-month history of marked bilateral pitting leg oedema, new malar and photo-exposed maculo-papular rash; previously documented positive ANA/anti-SSA and no prior systemic disease. |
| Llama4-Marveric | The patient is a female in her 30s, at 30 weeks gestation, presenting with severe lower limb pitting edema, a new malar rash, and an erythematous maculo-papular rash on her shoulders and chest. She has a history of positive ANA and ENA SS-A in 2017 but was lost to follow-up. |
| **Phase I – Q2: Differential diagnoses and rationale** | |
| DeepSeek-R1 | **Differential Diagnoses**<br>**1. Systemic Lupus Erythematosus (SLE) flare** (most likely) – Rash, edema, and prior autoantibodies suggest new-onset or reactivated SLE. Pregnancy is a known trigger.<br>**2. Preeclampsia with severe features** (must not miss) – Acute edema in third trimester risks eclampsia, HELLP, or fetal compromise.<br>**3. Polymorphic eruption of pregnancy (PEP) –** Benign pregnancy-related rash, but atypical with edema and malar involvement.<br><br>**Rationale:** SLE is prioritized due to classic malar rash, prior serology (SS-A linked to neonatal lupus), and edema (possible lupus nephritis). Preeclampsia cannot be missed due to maternal-fetal mortality risks. PEP is less likely without abdominal rash. |
| Gemini 2.5 Pro | Differential Diagnosis<br>**1. Systemic Lupus Erythematosus (SLE) Flare with Lupus Nephritis:** This is the **most likely** diagnosis. The new malar and truncal rashes, severe edema (suggesting heavy proteinuria), and prior positive autoantibodies (ANA, anti-SSA) strongly point to active SLE. Pregnancy is a well-known trigger for SLE flares.<br>**2. Preeclampsia:** This is a **must-not-miss** diagnosis. It is a common hypertensive disorder of pregnancy occurring after 20 weeks, characterized by new-onset hypertension and proteinuria, often accompanied by edema. It carries significant maternal and fetal risk. The clinical overlap with lupus nephritis (edema, proteinuria, potential renal dysfunction) makes it a critical differential.<br>**3. Primary Nephrotic Syndrome (e.g., Minimal Change Disease, FSGS):** Severe edema implies nephrotic syndrome. While less likely given the prominent extra-renal features, a |

| | |
|---|---|
| | primary glomerular disease unrelated to SLE can present de novo during pregnancy. |
| OpenAI GPT-o3 | Differential diagnosis<br>a. Systemic lupus erythematosus with lupus nephritis – most likely: fits rash pattern, serology and nephrotic-type oedema in pregnancy.<br>b. Pre-eclampsia / HELLP syndrome – common, dangerous; oedema ± rash mimicking SLE must be excluded.<br>c. Dermatomyositis (pregnancy-associated) – malar/"shawl" rash; can cause oedema via hypoalbuminaemia; rarer but important.<br>(Must not miss pre-eclampsia because of maternal-fetal mortality.) |
| Llama4-Marveric | Differential Diagnoses<br>**1. Systemic Lupus Erythematosus (SLE) flare:** Given her history of positive ANA and ENA SS-A, and new symptoms such as malar rash and erythematous maculo-papular rash, an SLE flare is highly considered.<br>**2. Preeclampsia:** Severe edema and rash could be associated with preeclampsia, a condition characterized by new-onset hypertension and often proteinuria after 20 weeks of gestation.<br>**3. Pregnancy-related dermatoses:** Conditions like pruritic urticarial papules and plaques of pregnancy (PUPPP) could explain the rash but are less likely to be associated with the malar rash and positive autoantibodies.<br><br>The most likely diagnosis is an **SLE flare**, given her history of positive autoantibodies and the nature of her rash.<br>**Must not be missed:** Preeclampsia, as it is a serious condition that can affect both mother and fetus. |
| **Phase I – Q3: Necessary physical examinations and reasons** | |
| DeepSeek-R1 | **Targeted History/Examination**<br>**History:**<br>• Hypertension symptoms (headache, visual changes): Differentiates preeclampsia.<br>• Fetal movements: Assesses fetal wellbeing in preeclampsia/SLE.<br>• Joint pain, photosensitivity, oral ulcers: Supports SLE flare.<br>• Pruritus: Suggests PEP if severe.<br><br>**Physical Exam:**<br>• BP measurement: Hypertension indicates preeclampsia.<br>• Urinalysis for proteinuria: Nephrotic-range proteinuria suggests preeclampsia or lupus nephritis.<br>• Rash distribution/scaling: Malar rash specificity for SLE; PEP typically spares face.<br>• Joint swelling/tenderness: Active SLE involvement.<br><br>**Purpose:** Confirm/rule out preeclampsia (hypertension/proteinuria) and SLE (systemic symptoms), while excluding PEP (pruritus without systemic features). |
| Gemini 2.5 Pro | Additional History and Physical Examination<br>**Additional Questions:**<br>**1. SLE Review of Systems:** "Have you experienced any fevers, joint pain/swelling, mouth sores, or unusual hair loss?"<br>• **Purpose:** To identify other systemic features of active SLE, which would increase the likelihood of this diagnosis over preeclampsia.<br>**2. Preeclampsia Warning Symptoms:** "Have you had any persistent headaches, visual disturbances like flashing lights, or pain in the upper right side of your abdomen?"<br>• **Purpose:** The presence of these symptoms indicates severe features of preeclampsia, requiring immediate obstetric intervention.<br><br>**Physical Examination:**<br>**1. Blood Pressure Measurement:** |

| | |
|---|---|
| | • **Purpose:** To check for hypertension (>140/90 mmHg), a key diagnostic criterion for preeclampsia. While often present in lupus nephritis, its absence would make preeclampsia less likely.<br>**2. Cardiopulmonary Examination:**<br>• **Purpose:** To assess for signs of volume overload (e.g., pulmonary rales, elevated jugular venous pressure) secondary to severe nephrotic syndrome, which can be seen in both conditions.<br>**3. Detailed Skin and Joint Examination:**<br>• **Purpose:** To confirm the nature of the rash (e.g., sparing of nasolabial folds in the malar rash) and to look for other cutaneous lupus signs or evidence of synovitis (joint inflammation). |
| OpenAI GPT-o3 | Focused history / examination<br>• Blood-pressure, headaches, visual changes, epigastric pain → screen pre-eclampsia.<br>• Joint pain, oral ulcers, serositis symptoms, Raynaud, alopecia → support SLE.<br>• Proximal muscle weakness, dysphagia → dermatomyositis.<br>• Medication, infection, toxin exposures → alternative nephrotic causes.<br>• Full skin exam for Gottron papules, heliotrope, discoid lesions.<br>• Fundoscopy, reflexes, clonus → severe pre-eclampsia signs.<br>• Heart, lungs, abdomen for effusions/ascites (SLE serositis).<br>• Fetal growth and wellbeing assessment. |
| Llama4-Marveric | Additional Questions and Physical Exams<br>**1. Blood pressure measurement:** To assess for hypertension suggestive of preeclampsia.<br>**2. Urine protein analysis:** To check for proteinuria, a key feature of preeclampsia and potential kidney involvement in SLE.<br>**3. Detailed rash examination:** To characterize the rash further and assess for other signs of SLE (e.g., alopecia, oral ulcers).<br>**4. Inquiry about other SLE symptoms:** Such as joint pain, fever, or serositis.<br>**5. Fetal monitoring:** To assess fetal well-being, given the potential complications of both SLE flare and preeclampsia. |
| **Phase I – Q4: Plan for investigations/tests** | |
| DeepSeek-R1 | **Diagnostic Testing Plan**<br>**1. Urinalysis with protein quantification** (e.g., 24-hour urine):<br>• Purpose: Detect proteinuria for preeclampsia or lupus nephritis.<br>• Expected: Nephrotic-range proteinuria in preeclampsia/lupus nephritis; negative in PEP.<br>**2. Blood tests:**<br>• Renal function (creatinine, eGFR): Assess renal impairment in SLE/preeclampsia.<br>• Liver enzymes, platelets: Screen for HELLP syndrome.<br>• Complement (C3/C4), anti-dsDNA: Low complements/elevated anti-dsDNA support SLE flare.<br>**3. Obstetric ultrasound with Doppler:**<br>• Purpose: Evaluate fetal growth, amniotic fluid, umbilical flow; SS-A increases fetal heart block risk.<br>**4. Anti-Ro/SS-A re-testing:**<br>• Purpose: Confirm persistence; critical for neonatal lupus monitoring.<br><br>**Rationale:** Cost-effective initial tests prioritize life-threatening conditions (preeclampsia/SLE) and fetal risks. Non-invasive urinalysis and bloodwork precede biopsies if needed. |
| Gemini 2.5 Pro | Diagnostic Tests<br>**1. Urinalysis with microscopy and Urine Protein-to-Creatinine Ratio (UPCR):**<br>• **Purpose:** To confirm and quantify proteinuria and to check for an active urinary sediment (red blood cells, white blood cells, or cellular casts). |

| | |
|---|---|
| | • **Expected Results:** Significant proteinuria is expected in both conditions. An active sediment would strongly favor lupus nephritis over preeclampsia.<br><br>**2. Blood Tests:**<br>• **Panels:** Complete Blood Count (CBC), Comprehensive Metabolic Panel (including creatinine, albumin, and liver function tests).<br>• **Immunology:** Anti-dsDNA antibody and complement levels (C3, C4).<br>• **Purpose:** To assess renal function, check for cytopenias (e.g., anemia, thrombocytopenia) and low albumin seen in SLE/nephrotic syndrome, and evaluate for liver involvement (HELLP syndrome). High anti-dsDNA and low C3/C4 levels are specific markers for an SLE flare and are typically normal in isolated preeclampsia.<br><br>**3. Renal Biopsy:**<br>• **Purpose:** To provide a definitive diagnosis and classification of lupus nephritis, which is crucial for guiding specific immunosuppressive therapy. It is the gold standard to differentiate lupus nephritis from other glomerular diseases or thrombotic microangiopathy seen in severe preeclampsia. This would be considered if the diagnosis remains unclear after non-invasive testing and aggressive immunosuppression is contemplated. |
| OpenAI GPT-o3 | Investigations and expected utility<br>• BP monitoring, urine dip/spot protein-creatinine or 24 h protein → quantify proteinuria, pre-eclampsia vs nephrotic.<br>• CBC, platelets, creatinine, urea, electrolytes, LFTs, uric acid → pre-eclampsia/HELLP, renal function.<br>• Serum albumin, lipids → nephrotic assessment.<br>• ANA profile, anti-dsDNA, complement C3/C4 → active SLE (low C3/C4, high dsDNA).<br>• CK, aldolase, myositis antibody panel → dermatomyositis (elevated CK).<br>• Coagulation and antiphospholipid antibodies → SLE obstetric risk, clot risk.<br>• Renal ultrasound ± Doppler → structural/flow issues.<br>• Fetal ultrasound with Dopplers and CTG → fetal compromise.<br>(Consider renal biopsy postpartum or earlier if safe, to classify lupus nephritis.) |
| Llama4-Marveric | Tests to Narrow Down the Diagnosis<br>**1. Complete Blood Count (CBC):** To check for cytopenias (e.g., anemia, thrombocytopenia) that can occur in SLE.<br>**2. Kidney function tests:** To assess for renal impairment, which can be seen in both SLE nephritis and preeclampsia.<br>**3. Urine analysis:** To confirm proteinuria and check for hematuria or casts suggestive of SLE nephritis.<br>**4. Complement levels (C3, C4):** Low levels can indicate active SLE.<br>**5. Anti-dsDNA antibodies:** To further support the diagnosis of SLE and assess disease activity.<br>These tests will help differentiate between SLE flare, preeclampsia, and other conditions, guiding appropriate management. |
| **Phase II – Q5: Interpreting test results** | |
| DeepSeek-R1 | Interpretation of Test Results<br>**Key findings and clinical significance:**<br>• **Nephrotic-range proteinuria** (PCR 834 g/mol; equivalent to ~8.3 g/day) with **severe hypoalbuminemia** (<15 g/L) confirms nephrotic syndrome. This, combined with active urinary sediment (WCC >500, RBC 250), suggests glomerular inflammation.<br>• **Serological activity:**<br> • Elevated anti-dsDNA (>90 IU/mL) and low complements (C3 0.3 g/L, C4 0.1 g/L) indicate active SLE.<br> • Anti-SSA/Ro positivity (historical and current) implies fetal risks (neonatal |

| | |
|---|---|
| | lupus/cardiac complications).<br>• **Normotension** (113/73) and **normal sFLT-1/PlGF ratio** (8.8) exclude preeclampsia despite edema.<br>• **Preserved renal function** (creatinine 82 µmol/L, eGFR 83 mL/min) suggests acute LN rather than chronic damage.<br>• **Fetal wellbeing:** Normal EFW (74th centile) and Dopplers rule out placental insufficiency.<br><br>**In context:** Findings align with an acute lupus nephritis flare triggered by pregnancy, not preeclampsia. Anti-SSA persistence demands fetal cardiac surveillance. |
| Gemini 2.5 Pro | Interpretation of Test Results<br>The patient's laboratory results provide a clear and compelling picture.<br>• **Confirmation of Severe Nephrotic Syndrome:** The spot urine PCR of 834 g/mol (approximately 8.3 grams/day) and a profoundly low serum albumin of <15 g/L confirm severe nephrotic-range proteinuria. This is the direct cause of her extensive pitting edema.<br>• **Evidence of Active Glomerulonephritis:** The presence of haematuria (RBC 250 x$10^6$/L) combined with heavy proteinuria indicates a nephrotic-nephritic syndrome, which points towards an inflammatory, proliferative glomerular process rather than a pure podocytopathy.<br>• **Confirmation of Active SLE:** The high-titre anti-dsDNA (>90 IU/mL) and marked hypocomplementemia (low C3 and C4) are serological hallmarks of active Systemic Lupus Erythematosus. This combination is highly specific for an SLE flare and serves as the etiological link to the glomerulonephritis.<br>• **Evidence against Preeclampsia:** The patient is normotensive (113/73 mmHg), and the absence of significant clonus argues against severe preeclampsia. Furthermore, the sFLT-1/PlGF ratio of 8.8 at admission is well below the typical cutoff for diagnosing preeclampsia (e.g., <38), effectively ruling it out as the primary diagnosis at this time.<br>• **Renal Function:** The admission creatinine of 82 µmol/L and eGFR of 83 mL/min represent a significant reduction in renal function. During the third trimester of pregnancy, GFR is physiologically hyper-filtrating (often >120 mL/min). Therefore, an eGFR of 83 signifies a meaningful acute kidney injury.<br>• **Fetal Considerations:** The positive anti-SSA/Ro antibody is a crucial finding, as it confers a risk (~2%) of neonatal lupus, most seriously congenital heart block. Close fetal monitoring is therefore essential. The normal fetal ultrasound is reassuring for now.<br><br>In summary, the results are pathognomonic for a severe flare of Lupus Nephritis (LN) causing a nephrotic-nephritic syndrome and acute kidney injury, occurring in the context of pregnancy. Preeclampsia has been effectively excluded. |
| OpenAI GPT-o3 | Interpretation of the laboratory data<br>• Nephrotic-range proteinuria – Spot PCR 834 g/mol (~8.3 g/24 h) with serum albumin <15 g/L explains the massive oedema; haematuria (RBC > 500 ×$10^6$/L) confirms glomerular inflammation.<br>• Serology – High anti-dsDNA titre (>90 IU/mL) and markedly low C3/C4 indicate active immune-complex SLE. Anti-SSA/Ro positivity is concordant with SLE but antiphospholipid antibodies are absent.<br>• Renal function – Creatinine 82 µmol/L (eGFR 83 mL/min) shows preserved filtration, a pattern typical of class V (±III/IV) lupus nephritis presenting with heavy proteinuria rather than acute renal failure.<br>• Normotension and a low sFlt-1/PlGF ratio (8.8) at admission argue strongly against pre-eclampsia at that time.<br>• Post-partum course –<br>– Four weeks post-delivery: proteinuria and hypoalbuminaemia improve, complements rebound and anti-dsDNA falls, consistent with treatment response; the transient rise in sFlt- |

| | |
|---|---|
| | 1/PlGF (194) suggests endothelial activation but, without hypertension or renal impairment, is more likely a postpartum physiological surge than true pre-eclampsia.<br>− By three months and at 18 months: sustained fall in proteinuria (PCR 82 → 5.8 g/mol), normalised complements and albumin restoration (41 g/L) confirm remission.<br>• Other values – Hypocalcaemia on admission is pseudo-hypocalcaemia from low albumin; platelet count, LFTs and haemoglobin remain stable, making HELLP unlikely. |
| Llama4-Marveric | Interpretation of Test Results<br>The patient's laboratory results reveal several key findings:<br>• **Severe proteinuria** (urine PCR of 834 g/mol) and **hypoalbuminemia** (serum albumin < 15 g/L) indicate significant kidney damage, consistent with nephrotic syndrome.<br>• **Elevated anti-dsDNA (> 90 IU/mL)** and **hypocomplementemia** (low C3 and C4) support the diagnosis of an active SLE flare, particularly Lupus Nephritis (LN).<br>• **Normal blood pressure** (113/73 mmHg) and the absence of other signs of preeclampsia (e.g., normal sFLT-1/PlGF ratio initially, though it later increased) help differentiate LN from preeclampsia.<br>• The presence of **anti-SSA/Ro antibodies** is noted, which can be associated with SLE and potential neonatal lupus, though antiphospholipid antibodies were negative, reducing the risk for certain pregnancy complications.<br>These findings are critical for diagnosing LN and guiding treatment. |
| **Phase II – Q6: Reconsidering the differential diagnosis** | |
| DeepSeek-R1 | Evolution of Diagnostic Thinking<br>**Revised differential prioritization:**<br>**1. Lupus nephritis (Class IV/V) flare** (most probable):<br>• Supported by nephrotic syndrome, active sediment, serological activity (anti-dsDNA↑, C3/C4↓), and rash.<br>• Excludes preeclampsia (normotension, normal sFLT-1/PlGF).<br>**2. Isolated preeclampsia** (ruled out): Absence of hypertension, thrombocytopenia, or abnormal angiogenic markers.<br>**3. Pregnancy-related dermatoses** (discarded): Rash correlates with SLE activity, not PEP (no pruritus, systemic features).<br><br>**Working Diagnosis: Active SLE with Lupus Nephritis (Class IV/V).**<br>**Rationale:** Pregnancy is a known trigger for SLE flares. Prior autoantibodies (2017) imply undiagnosed SLE, now manifesting as nephritis. |
| Gemini 2.5 Pro | Revised Diagnostic Thinking and Working Diagnosis<br>Based on the definitive test results, my diagnostic thinking has shifted from a differential diagnosis to a confident, specific diagnosis.<br>The initial top differential, **Lupus Nephritis**, is now confirmed. The combination of clinical findings (rash, edema), serological evidence of highly active SLE (positive ANA, high anti-dsDNA, low complements), and renal manifestations (nephrotic-range proteinuria, haematuria, reduced GFR for pregnancy) fulfills the classification criteria for SLE with active nephritis.<br>**Preeclampsia,** the other major differential, is now considered highly unlikely as the primary cause given the normotension and the negative sFLT-1/PlGF ratio. While preeclampsia can be superimposed on LN, there is no evidence for it at this stage.<br>Therefore, the working diagnosis is no longer provisional.<br><br>**Working Diagnosis: Active Lupus Nephritis (likely proliferative, Class III or IV) presenting with severe nephrotic-nephritic syndrome during the third trimester of pregnancy.** |
| OpenAI GPT-o3 | Revised diagnostic thinking<br>Initial differentials were lupus nephritis vs pre-eclampsia/HELLP. The serology, nephrotic-range proteinuria with preserved GFR, normotension and low early sFlt-1/PlGF decisively favour lupus nephritis. The later biochemical response to immunosuppression and renal |

| | |
|---|---|
| | biopsy findings (timed during falling proteinuria) consolidate this.<br>Working diagnosis: New-onset systemic lupus erythematosus in pregnancy, presenting as nephrotic lupus nephritis—most consistent with class V (membranous) ± minor proliferative component—without concurrent pre-eclampsia. |
| Llama4-Marveric | Diagnostic Thinking and Working Diagnosis<br>Given the patient's presentation, laboratory results, and the presence of significant proteinuria, hypoalbuminemia, elevated anti-dsDNA, and hypocomplementemia, the **working diagnosis is Lupus Nephritis (LN) in the context of an SLE flare during pregnancy.**<br><br>The evidence strongly supports LN over preeclampsia, given the normotension, significant proteinuria, and serological evidence of SLE activity. |
| **Phase II – Q7: Treatment planning** | |
| DeepSeek-R1 | Treatment Plan<br>**Immediate Plan:**<br>**1. Immunosuppression:**<br>• **Hydroxychloroquine** (200 mg BID): Baseline SLE control (EBM: reduces flares).<br>• **IV methylprednisolone** (500 mg × 3 days), then **oral prednisolone** (0.5 mg/kg/day): Rapid LN control (safe in pregnancy).<br>• **Tacrolimus** (target trough 5–8 μg/L): Steroid-sparing agent for LN (EBM: effective in pregnancy; see post-treatment PCR↓).<br>**2. Fetal Protection:**<br>• **Weekly fetal echocardiography:** Anti-SSA mandates monitoring for heart block.<br>• **Betamethasone** (if preterm delivery likely): Lung maturation.<br>**3. Supportive Care:**<br>• **Albumin infusion + furosemide:** For severe edema (caution: avoid volume overload).<br>• **Thromboprophylaxis:** LMWH if nephrotic syndrome persists (albumin <20 g/L).<br><br>**Rationale:** This regimen balances LN control (steroids + calcineurin inhibitors) with fetal safety. Tacrolimus was chosen over cyclophosphamide (teratogenic) or mycophenolate (contraindicated in pregnancy).<br>**Alternative Options:**<br>• Azathioprine: Safe in pregnancy but slower onset than tacrolimus.<br>• IVIG/Plasmapheresis: Reserved for steroid-refractory cases.<br>**Monitoring:**<br>• **Maternal:** Daily BP, weekly urine PCR, anti-dsDNA, complements.<br>• **Fetal:** Biweekly growth scans + Doppler.<br><br>**Expected Response:** Proteinuria reduction (as seen post-treatment: PCR 82 g/mol) and serological improvement (C3↑, anti-dsDNA↓).<br>Please enter the treatment course. |
| Gemini 2.5 Pro | Treatment Plan<br>The patient has a severe, organ-threatening disease that also poses significant risk to the fetus. The treatment plan requires a multidisciplinary approach involving nephrology, rheumatology, and high-risk obstetrics (maternal-fetal medicine).<br><br>**Immediate Goals:**<br>1. Induce remission of lupus nephritis to preserve maternal renal function.<br>2. Manage the complications of nephrotic syndrome.<br>3. Prolong the pregnancy to a safer gestational age for the fetus.<br><br>**Treatment Plan:**<br>**1. Induction Immunosuppression:** |

| | |
|---|---|
| | • **Corticosteroids:** Initiate high-dose corticosteroids immediately. This typically involves intravenous methylprednisolone "pulses" (e.g., 500 mg daily for 3 days) followed by high-dose oral prednisone (0.5-1.0 mg/kg/day). This is the cornerstone for rapidly controlling severe inflammation.<br>• **Calcineurin Inhibitor (CNI):** Add **Tacrolimus** as a steroid-sparing agent. CNIs are considered safe in pregnancy and are effective for inducing remission in lupus nephritis, particularly in reducing proteinuria. Target trough levels would be 5-8 µg/L. This is preferred over azathioprine for severe induction due to its more rapid onset of action. Mycophenolate mofetil and cyclophosphamide are contraindicated due to teratogenicity.<br>• **Hydroxychloroquine (HCQ):** Start or continue HCQ (e.g., 200-400 mg daily based on weight). It is safe in pregnancy and is proven to reduce SLE flares, improve pregnancy outcomes, and may reduce the risk of congenital heart block associated with anti-SSA antibodies.<br>**2. Supportive Care:**<br>• **Thrombosis Prophylaxis:** The combination of nephrotic syndrome and pregnancy creates a very high risk for thromboembolism. Prophylactic anticoagulation with low-molecular-weight heparin (LMWH) is mandatory.<br>• **Edema Management:** Judicious use of loop diuretics (e.g., furosemide) is necessary to manage the severe edema, while carefully monitoring maternal volume status to avoid compromising uteroplacental blood flow.<br>• **Blood Pressure Control:** Although currently normotensive, monitor blood pressure closely. If hypertension develops, pregnancy-safe agents like labetalol or nifedipine would be used.<br>**3. Maternal and Fetal Monitoring:**<br>• **Maternal:** Closely monitor renal function, proteinuria, and serological markers (dsDNA, complements) to assess treatment response.<br>• **Fetal:** Implement weekly or bi-weekly fetal surveillance with ultrasounds for growth and well-being. Crucially, given the anti-SSA positivity, perform serial fetal echocardiograms to screen for the development of congenital heart block.<br><br>**Rationale:** This plan uses an evidence-based, pregnancy-safe regimen to aggressively treat a severe lupus flare. The goal is to rapidly gain disease control to protect the mother's kidneys and allow the pregnancy to continue, while actively managing the high-risk complications of both the disease and its treatment. A postpartum renal biopsy is planned to definitively classify the nephritis and guide long-term therapy, as deferring it until after delivery is safer for the pregnancy. |
| OpenAI GPT-o3 | Treatment plan and rationale<br>Pregnancy (30–37 weeks)<br>• High-dose corticosteroid: i.v. methyl-prednisolone pulses followed by oral prednisolone (≤0.5 mg/kg) – rapid control of glomerular inflammation; safe in late pregnancy.<br>• Calcineurin inhibitor: tacrolimus 0.05–0.1 mg/kg/day (target trough 4–8 µg/L) – effective for class V LN and acceptable fetal safety; already employed and levels adjusted.<br>• Hydroxychloroquine 200–400 mg/day – reduces SLE flares; continued through pregnancy.<br>• Maternal-fetal support: low-dose aspirin (150 mg), LMWH prophylaxis (nephrotic-range proteinuria), salt restriction ± loop diuretic for oedema, close obstetric & fetal Doppler surveillance.<br><br>Post-partum<br>• Renal biopsy to confirm class and activity (completed).<br>• Maintain tacrolimus; taper steroids to physiological dose within 3–6 months if stable.<br>• Add ACE-I/ARB (e.g., perindopril) once breastfeeding plans allow, for antiproteinuric and renoprotective effect. |

<table>
<tr><td></td><td>• If relapse or proliferative component emerges: switch to mycophenolate mofetil (first-line for class III/IV/V outside pregnancy) or consider Euro-Lupus low-dose cyclophosphamide; decision guided by fertility plans and biopsy findings.<br>• Vaccinations, cardiovascular risk management, and routine SLE monitoring (dsDNA, complements, renal parameters).<br>Alternative options<br>• Belimumab or rituximab for refractory disease (not first-line in pregnancy).<br>• Plasma exchange if life-threatening nephrotic syndrome or TMA.<br>• Early delivery if maternal/fetal status deteriorates (e.g., rising BP, worsening renal function).</td></tr>
<tr><td>Llama4-Marveric</td><td>Treatment Plan<br>Rationale:<br>1. Induction Therapy for LN: The patient requires immediate treatment for LN to prevent further renal damage. Given her pregnancy status and the severity of LN, corticosteroids are the first-line treatment.<br>2. Immunosuppressive Therapy: After delivery, or if necessary during pregnancy (though with caution due to potential fetal risks), additional immunosuppressive therapy like cyclophosphamide or azathioprine may be considered for LN treatment.<br>3. Monitoring: Close monitoring of renal function, SLE activity, and fetal well-being is essential.<br><br>Treatment Options:<br>1. Corticosteroids: High-dose corticosteroids (e.g., prednisone or methylprednisolone) for initial treatment.<br>2. Tacrolimus: Already initiated, as indicated by the tacrolimus levels, which is sometimes used in managing LN, especially in pregnancy where it is considered relatively safer than other immunosuppressants.<br>3. Postpartum Care: Adjust treatment according to postpartum LN activity and renal function.<br><br>Alternative Options and Considerations:<br>• Cyclophosphamide: Can be considered postpartum for severe LN but is generally avoided in pregnancy due to teratogenic risks.<br>• Azathioprine: May be used during pregnancy for SLE management if necessary, though with caution.<br><br>The treatment plan should be tailored to the patient's response and the risks associated with both SLE/LN and pregnancy. Multidisciplinary care involving rheumatology, nephrology, and obstetrics is crucial.</td></tr>
<tr><td colspan="2">Phase III – Q8: Evaluation of treatment</td></tr>
<tr><td>DeepSeek-R1</td><td>Monitoring Effectiveness and Safety<br>Effectiveness Monitoring:<br>• Proteinuria: Urine ACR/PCR monthly (target: <30 mg/mmol ACR or <50 mg/mol PCR).<br>• Serological Activity: Anti-dsDNA and C3/C4 every 3 months (target: anti-dsDNA <30 IU/mL, C3 >0.9 g/L).<br>• Renal Function: Serum creatinine/eGFR monthly (target: eGFR >60 mL/min).<br>• Immunosuppression Levels: Tacrolimus trough (target: 5–8 μg/L) monthly.<br><br>Safety Monitoring and Risks:
<table>
<tr><th>Risk</th><th>Monitoring</th><th>Countermeasures</th></tr>
<tr><td>Tacrolimus toxicity</td><td>Serum creatinine, $K^+$, glucose monthly; neuro symptoms (tremor)</td><td>Dose adjustment for levels >8 μg/L; manage hyperglycemia/hypomagnesemia.</td></tr>
</table></td></tr>
</table>

| | | |
|---|---|---|
| **Mycophenolate toxicity** | CBC biweekly ×3 mo, then monthly (WCC >3.0$\times10^9$/L) | Hold if WCC <2.5$\times10^9$/L; teratogenicity counseling (contraception essential). |
| **Steroid complications** | DEXA scan annually; BP/glucose monthly | Calcium/vitamin D supplementation; osteoporosis prophylaxis if long-term use. |
| **Infection risk** | Clinical symptoms, CRP | Pneumocystis jirovecii prophylaxis (if lymphs <0.5$\times10^9$/L); avoid live vaccines. |
| **Thrombosis** | Clinical signs of DVT/PE | Re-initiate LMWH if albumin <20 g/L or PCR >100 mg/mol. |

Gemini 2.5 Pro

How will you monitor the effectiveness and safety of the treatment? What are the specific risks to watch out for, and what are your concrete countermeasures for them?

Effective monitoring requires a multi-faceted approach focusing on disease activity, medication side effects, and patient adherence, which is a known challenge for this patient.

**Monitoring Treatment Effectiveness:**

- **Renal Function & Proteinuria:** Monthly, then transitioning to 3-monthly, checks of serum creatinine/eGFR and urine albumin-to-creatinine ratio (ACR). The goal is to maintain a stable eGFR and achieve complete remission (ACR < 30 mg/g or 3 mg/mmol).
- **SLE Serology:** 3 to 6-monthly monitoring of anti-dsDNA and complement (C3/C4) levels. A rise in anti-dsDNA or a fall in complements can herald a clinical relapse.
- **Clinical Assessment:** At each visit, assess for clinical signs of a flare, such as new rashes, arthritis, serositis, or edema, and monitor blood pressure.

**Monitoring Safety and Managing Risks:**

The patient is on a potent triple immunosuppressive regimen (Prednisone, Tacrolimus, MMF) with significant risks.

**1. Risk: Infection (Opportunistic & General)**

- **Monitoring:** Regular Complete Blood Count (CBC) to watch for leukopenia. Vigilant clinical monitoring for any signs of infection (fever, cough, dysuria).
- **Countermeasures:**
  - **Patient Education:** Counsel on recognizing early signs of infection and seeking immediate medical attention.
  - **Prophylaxis:** Given the triple therapy, strongly consider prophylaxis for Pneumocystis jirovecii pneumonia (PJP) with trimethoprim-sulfamethoxazole until the prednisone dose is minimal and the overall immunosuppression is reduced.
  - **Vaccinations:** Ensure she is up-to-date on vaccinations, including influenza, pneumococcus, and COVID-19.

**2. Risk: Medication-Specific Toxicity**

- **Tacrolimus:**
  - *Risks*: Nephrotoxicity, hypertension, neurotoxicity (tremor), hyperglycemia, hyperkalemia.
  - *Monitoring*: Regular monitoring of tacrolimus trough levels (target 3-7 μg/L), serum creatinine, blood pressure, electrolytes (especially potassium), and blood glucose.
- **Mycophenolate Mofetil (MMF):**
  - *Risks*: GI intolerance (diarrhea, nausea - a likely cause of her previous self-cessation), bone marrow suppression (leukopenia). **Extreme teratogenicity.**
  - *Monitoring*: Monitor CBC. Actively inquire about GI side effects at every visit.
  - *Countermeasures*: **Crucially, ensure the patient is on reliable, long-term contraception and understands the severe risk of birth defects.** For GI intolerance, consider dose reduction, splitting doses, or switching to the enteric-coated formulation

| | |
|---|---|
| | (mycophenolic acid).<br>• **Prednisone:**<br>• *Risks*: Osteoporosis, weight gain, hyperglycemia, mood changes.<br>• *Monitoring*: Monitor weight and blood glucose. Plan for a bone density (DEXA) scan.<br>• *Countermeasures*: Ensure adequate calcium and vitamin D intake. Continue gradual dose tapering as tolerated.<br>• **Hydroxychloroquine (HCQ):**<br>• *Risk*: Retinal toxicity (rare but serious).<br>• *Monitoring*: Confirm baseline ophthalmology screening was done and ensure annual follow-up.<br><br>**3. Risk: Long-term Complications**<br>• **Monitoring:** Annual screening for cardiovascular risk factors (lipid panel, HbA1c) and skin cancer.<br>• **Countermeasures:** Counsel on aggressive management of traditional cardiovascular risk factors and strict sun protection. |
| OpenAI GPT-o3 | Monitoring plan: effectiveness, safety, anticipated risks, countermeasures<br>• Clinical status Weight, oedema, BP, rash, arthralgia, urine output – every visit (initially fortnightly, then monthly).<br>Target: regression of oedema, BP < 130/80 mmHg, symptom-free.<br>• Kidney parameters Serum Cr/eGFR, spot PCR (or 24 h protein), serum albumin – every 4–6 wk until stable, then q-3 mo.<br>Goal: eGFR stable > 90 mL/min, PCR < 50 mg/mmol (emaACR < 25 mg/mmol).<br>• Immunological activity Anti-dsDNA, C3/C4 – every 2–3 mo.<br>Goal: dsDNA low or falling, complements normal.<br>• Tacrolimus trough level Aim 4–6 µg/L – check 1 wk after every dose change, then q-2 mo; adjust to avoid nephrotoxicity.<br>• Mycophenolate and azathioprine effects CBC, LFT every 2 wk × 2, then q-1–2 mo; look for leukopenia or hepatitis; hold/drug-reduce if WBC < 3.5 $\times 10^9$/L or ALT > 3×ULN.<br>• Prednisone toxicity BP, glucose, weight, mood, eye pressure, DEXA (baseline then q-2 yr), consider Ca/Vit-D and bisphosphonate prophylaxis.<br>• Hydroxychloroquine toxicity Ocular screening (baseline and yearly) plus drug level if adherence questioned.<br>• Infection surveillance Temp, full blood count, CRP if unwell; counsel on early presentation; consider PJP prophylaxis if cumulative steroid > 20 mg/day for >1 mo.<br>• Cardiovascular & metabolic risk Lipids, HbA1c yearly; encourage smoking cessation, exercise.<br>• Reproductive safety Strict contraception (MMF teratogenic); document counselling; switch to AZA or hold MMF if pregnancy desired.<br>• Nursing & cultural support Regular contact with local nurse navigator, reinforce adherence, arrange community pathology collections and telehealth review to mitigate geographic barriers. |
| Llama4-Marveric | Monitoring the Effectiveness and Safety of the Treatment<br>Effectiveness:<br>**1. Renal Function:** Monitor serum creatinine and eGFR to assess for improvement or deterioration in renal function.<br>**2. Proteinuria:** Regularly check urine PCR or ACR to assess the reduction in proteinuria.<br>**3. SLE Activity:** Monitor anti-dsDNA levels and complement levels (C3, C4) to gauge SLE activity.<br><br>Safety:<br>**1. Immunosuppressive Therapy Side Effects:** Monitor for side effects of tacrolimus |

| | |
|---|---|
| | (e.g., nephrotoxicity, hypertension), mycophenolate mofetil (e.g., gastrointestinal symptoms, infections), and prednisone (e.g., hyperglycemia, osteoporosis).<br>**2. Infection Risk:** Be vigilant for signs of infections, given the immunosuppressive therapy.<br>**3. Fetal/Infant Health:** Monitor the infant for any adverse effects related to in utero exposure to immunosuppressive drugs.<br><br>Countermeasures:<br>**1. Dose Adjustments:** Adjust doses of immunosuppressants based on efficacy and side effects.<br>**2. Regular Follow-Up:** Ensure regular follow-up with the patient to monitor for both effectiveness and safety.<br>**3. Multidisciplinary Care:** Continue multidisciplinary care involving nephrology, rheumatology, and potentially other specialties as needed. |
| **Phase III – Q9: Formulating plans for worsening conditions** | |
| DeepSeek-R1 | Response to Unexpected Changes<br>**Thought Process for Clinical Deterioration:**<br>**1. Rapid Reassessment:**<br>• Proteinuria increase: Rule out relapse (check anti-dsDNA/C3) vs. non-adherence (tacrolimus levels) vs. infection (urine culture).<br>• Renal function decline: Exclude dehydration, tacrolimus toxicity (elevated creatinine), or LN flare.<br>• New edema/rash: Differentiate SLE flare (serology) from allergic reaction or infection.<br><br>**Action Plan:**<br>• **Immediate Tests:** Urgent urine PCR/ACR, anti-dsDNA, complements, CBC, tacrolimus level.<br>• **Relapse Management:**<br> • Mild: Increase prednisone by 10–20 mg; optimize tacrolimus/MMF doses.<br> • Severe: IV methylprednisolone 500 mg ×3 days; consider rituximab if refractory.<br>• **Non-Adherence:** Engage nurse navigator for directly observed therapy; simplify regimen (e.g., once-daily MMF).<br>• **Telehealth Barriers:** Transition care to local renal service; schedule frequent nurse-led home visits.<br><br>**Proactive Measures:**<br>• **Education:** Side-effect management (e.g., antiemetics for MMF nausea).<br>• **Contingency Planning:** Emergency contact for symptom escalation; 3-month medication supply.<br><br>**Rationale:** Prioritize reversible causes (adherence/toxicity) before escalating immunosuppression. Maintain hydroxychloroquine throughout (prevents flares). |
| Gemini 2.5 Pro | If the patient's condition does not improve as expected or suddenly changes, how would you think and act?<br><br>A sudden change or failure to improve would trigger a rapid, systematic reassessment to differentiate between a disease flare and a treatment complication.<br>**Thought Process (Differential Diagnosis for Worsening):**<br>**1. Disease Flare (Relapse of Lupus Nephritis):** This is the highest probability, given her history. My immediate follow-up question is, "What triggered it?"<br> • **Medication Non-Adherence:** This is the most likely trigger based on her past relapses. A sub-therapeutic tacrolimus level would be strong objective evidence.<br> • **Under-treatment:** The current regimen may be insufficient as the prednisone is |

| | |
|---|---|
| | weaned, or the disease may have transformed to a more aggressive histological class.<br>• **Infection:** Infection can be a potent trigger for an SLE flare.<br>**2. Treatment Complication:**<br>• **Tacrolimus Nephrotoxicity:** This would present as a rising creatinine, but typically without a concurrent significant increase in proteinuria or active urinary sediment.<br>• **Thrombotic Microangiopathy (TMA):** A rare but serious complication of SLE or calcineurin inhibitors, characterized by a rising creatinine, thrombocytopenia, and evidence of hemolysis.<br>**3. Other Complications:**<br>• **Renal Vein Thrombosis**: A known complication of nephrotic syndrome, causing a sudden rise in proteinuria and worsening renal function.<br><br>**Action Plan:**<br>**1. Immediate Assessment:** Obtain an urgent clinical review and order STAT laboratory tests: CBC, serum creatinine, electrolytes, urine ACR, urinalysis with microscopy, and a tacrolimus trough level. Re-check anti-dsDNA and complement levels.<br>**2. Differentiate the Cause:**<br>• **If non-adherence is suspected/confirmed:** The primary action is to re-establish therapeutic immunosuppression, likely by temporarily increasing the prednisone dose and providing intensive support via the nurse navigator to address the barriers to adherence.<br>• **If a true disease flare is suspected despite adherence:** This indicates treatment failure. The first step would be to intensify the current regimen, primarily by increasing the prednisone dose (potentially with IV methylprednisolone pulses).<br>• **If tacrolimus toxicity is suspected:** I would reduce the tacrolimus dose and monitor for improvement in creatinine.<br>**3. Consider a Repeat Renal Biopsy:** A biopsy is the definitive diagnostic tool if the cause remains unclear or if the patient fails to respond to intensified therapy. It is essential to:<br>• Confirm a true LN flare versus chronic, irreversible scarring.<br>• Rule out drug toxicity (e.g., CNI-induced nephrotoxicity).<br>• Assess for a change in the histological classification (e.g., transformation from Class III to a more severe Class IV), which would necessitate a major change in the treatment strategy (e.g., consideration of cyclophosphamide or biologic agents like belimumab or voclosporin). |
| OpenAI GPT-o3 | Approach if the course deviates (no improvement or acute change)<br>Re-check the data<br>• Confirm adherence (pill counts, serum drug levels).<br>• Repeat key labs (Cr, PCR, dsDNA, complements), BP, and focused exam within 48 h.<br>• Exclude precipitating factors – infection (UCx, CXR), new pregnancy, NSAID use, tacrolimus level, volume depletion.<br>Clarify the pattern<br>• Rising proteinuria with stable creatinine → active LN relapse.<br>• Rising creatinine ± hypertension → consider tacrolimus nephrotoxicity vs proliferative LN progression vs TMA.<br>• Fever/leukopenia → drug toxicity or infection.<br>Act promptly<br>• Active LN suspected: pulse methyl-pred ± increase oral steroids; consider hospital admission; add/optimise MMF (up to 1.5 g bid) or switch to cyclophosphamide or voclosporin; discuss biologics (rituximab, belimumab) with multidisciplinary team.<br>• Suspected CNI toxicity: hold tacrolimus, hydrate, monitor renal function; re-start at lower dose or change class.<br>• Infection: cultures, start empiric antibiotics, withhold immunosuppression temporarily if severe. |

| | |
|---|---|
| | Re-image / re-biopsy<br>• If renal indices fail to improve within 2–4 wk or diagnosis unclear, arrange repeat renal biopsy to guide escalation.<br>Escalate support<br>• Engage obstetric medicine if pregnant, rheumatology, nephrology, Indigenous liaison staff; consider transfer to tertiary centre.<br>Reassess goals and adherence plan<br>• Simplify regimen, address side-effects, use dose packs or directly-observed therapy if necessary. |
| Llama4-Marveric | Response to Changes in the Patient's Condition<br>Thought Process:<br>**1. Reassess SLE Activity and LN Status:** If the patient's condition worsens or does not improve, reassess SLE activity and LN status through clinical evaluation and laboratory tests (e.g., anti-dsDNA, complement levels, urine PCR).<br>**2. Medication Adherence:** Check for adherence to prescribed medications, as non-adherence was a factor in her relapses.<br>**3. Differential Diagnoses:** Consider other causes for the change in condition, such as infections or other complications.<br><br>Action Plan:<br>**1. Adjust Treatment:** Adjust immunosuppressive therapy based on the reassessment, potentially increasing doses or switching medications.<br>**2. Intensify Monitoring:** Intensify monitoring for both efficacy and safety, including more frequent laboratory tests and clinical evaluations.<br>**3. Patient Education:** Reinforce the importance of medication adherence and provide support to manage side effects and complications.<br><br>By closely monitoring the patient's condition and adjusting the treatment plan as needed, it's possible to optimize outcomes for both the patient's SLE/LN and overall health. |

| **Case 6: Renal Injuries Induced by Supplements Containing Red Yeast Rice** [7] | |
|---|---|
| **Phase I – Q1: Summarizing medical problems** | |
| DeepSeek-R1 | 41–45-year-old man with **subacute (1-month) renal dysfunction** after 6 months of red yeast rice supplementation. Key symptoms: fatigue, anorexia, polyuria, and polydipsia. No prior renal/urinary abnormalities. |
| Gemini 2.5 Pro | This is a case of a 41–45-year-old man with previously normal renal function who presents with a subacute onset of renal dysfunction over one month, accompanied by constitutional symptoms (fatigue, anorexia) and symptoms suggestive of a urinary concentrating defect (polyuria, polydipsia). This presentation began five months after he started taking red yeast rice supplements. |
| OpenAI GPT-o3 | A 41–45-year-old previously healthy man, after six months of red-yeast-rice supplementation, has developed one month of fatigue, anorexia, polyuria and polydipsia, now found to have new renal dysfunction. |
| Llama4-Marveric | The patient is a 41–45-year-old man with no prior history of renal issues, presenting with renal dysfunction. He has been consuming supplements containing red yeast rice for six months. Symptoms of general fatigue, appetite loss, frequent urination, and increased thirst developed one month prior to the hospital visit. |
| **Phase I – Q2: Differential diagnoses and rationale** | |
| DeepSeek-R1 | Differential Diagnoses<br>**1. Supplement-induced acute tubulointerstitial nephritis (ATIN)**<br>• Most likely: Red yeast rice (containing lovastatin analogs) is linked to tubular toxicity and ATIN. Temporal association supports this.<br>**2. Diabetic nephropathy / new-onset diabetes**<br>• Polyuria/polydipsia suggest hyperglycemia or diabetes insipidus. |

| | |
|---|---|
| | **3. Acute tubular necrosis (ATN) from rhabdomyolysis**<br>• Must not miss: Statin-like compounds in red yeast rice can cause myotoxicity → myoglobinuric ATN. |
| Gemini 2.5 Pro | Differential Diagnosis<br>**1. Supplement-induced Acute Tubulointerstitial Nephritis (ATIN):**<br>• **Most Likely:** The temporal relationship between starting the red yeast rice supplement and the onset of symptoms is highly suggestive. The clinical picture of renal dysfunction with polyuria and polydipsia strongly points to tubular damage, a hallmark of ATIN. Contaminants (e.g., citrinin) in such supplements are a known cause of nephrotoxicity.<br>**2. Autoimmune Tubulointerstitial Nephritis (e.g., Sarcoidosis, TINU syndrome):**<br>• These conditions can present with subacute renal failure and evidence of tubular dysfunction. While less likely than a drug-induced cause given the clear exposure, they are an important consideration for idiopathic tubulointerstitial disease in this age group.<br>**3. Rapidly Progressive Glomerulonephritis (RPGN), such as ANCA-associated vasculitis:**<br>• **Must Not Be Missed:** Although the prominent polyuria makes a primary tubulointerstitial disease more likely, the constitutional symptoms and rapid decline in renal function could be the presenting features of RPGN. Missing this diagnosis can lead to irreversible kidney failure and multi-organ damage. |
| OpenAI GPT-o3 | Differential diagnosis<br>a. New-onset diabetes mellitus with hyperglycaemic osmotic diuresis causing pre-renal or early diabetic kidney injury – fits classic polyuria/polydipsia and could explain creatinine rise from dehydration. (Most likely.)<br>b. Supplement-induced kidney injury (acute interstitial nephritis or toxic ATN from red-yeast-rice contaminants) – temporal relation to supplement, can give non-oliguric AKI with constitutional symptoms. (Must not be missed because reversible if recognised.)<br>c. Hypercalcaemia (primary hyperparathyroidism, malignancy) producing nephrogenic diabetes insipidus, polyuria/polydipsia and renal impairment. (Important, potentially life-threatening.) |
| Llama4-Marveric | Differential Diagnoses<br>**1. Acute Kidney Injury (AKI) due to Supplement Toxicity:** The patient's consumption of red yeast rice supplements, which contains statin-like substances, could potentially cause renal dysfunction.<br>**2. Diabetic Ketoacidosis or Hyperglycemic Hyperosmolar State:** Symptoms like frequent urination and thirst could indicate uncontrolled diabetes mellitus.<br>**3. Other Causes of AKI (e.g., dehydration, medication, obstruction):** Various factors could lead to AKI, and a broad differential diagnosis is necessary.<br><br>The most likely diagnosis is **AKI due to Supplement Toxicity** or **Diabetes Mellitu**s, given the patient's symptoms and history of supplement intake.<br>A condition that **must not be missed** is severe hyperglycemia or diabetic ketoacidosis, as it can be life-threatening if not promptly treated. |
| **Phase I – Q3: Necessary physical examinations and reasons** | |
| DeepSeek-R1 | Targeted History/Exam<br>**1. Urine output volume** (e.g., oliguria vs. polyuria):<br>• *Clarify*: ATN/rhabdomyolysis often causes oliguria; diabetes causes polyuria.<br>**2. Muscle pain/tenderness exam:**<br>• *Clarify*: Rhabdomyolysis risk from statin analogs.<br>**3. Medication review** (other nephrotoxins, NSAIDs):<br>• *Clarify*: Exacerbating factors for ATIN or ATN.<br>**4. Flank pain or fever:**<br>• *Clarify*: Pyelonephritis or obstruction. |
| Gemini 2.5 Pro | Additional History and Physical Examination<br>• **History:** |

<table>
<tr><td></td><td>i. Supplement Details: I would ask for the specific product name, manufacturer, lot number, and daily dosage of the red yeast rice supplement. This is to identify the potential causative agent and check for any public safety alerts regarding contaminants.<br><br>ii. Systemic Review: I would ask about fever, skin rash, or joint pain. The presence of these "allergic" symptoms would increase the likelihood of drug-induced ATIN.<br>iii. Eye Symptoms: I would inquire about eye pain, redness, or changes in vision. This is to screen for uveitis, which would suggest TINU syndrome or sarcoidosis.<br><br>• Physical Examination:<br>i. Skin Examination: I would carefully inspect the skin for any new rashes, particularly a maculopapular rash. This is a classic, though not always present, sign of drug-induced ATIN.<br>ii. Volume Status Assessment: I would check for signs of dehydration, such as dry mucous membranes and orthostatic hypotension. This is to assess the severity of the polyuria and guide fluid management.</td></tr>
<tr><td>OpenAI GPT-o3</td><td>Focused history & physical examination<br>History: weight loss, vision change, infections (DM); muscle pain, dark urine, NSAID/other drugs, fever, rash, arthralgia (AIN/rhabdo); nephrolithiasis, bone pain, constipation (hypercalcaemia); urine volume/nocturia pattern; family history of DM or renal disease.<br>Physical: vitals & orthostatics (dehydration, hypercalcaemic crisis), BMI, skin turgor, mucous membranes, peripheral oedema, rash, scleral icterus, muscle tenderness, CVA tenderness, fundoscopic exam (diabetic changes).</td></tr>
<tr><td>Llama4-Marveric</td><td>Additional Questions and Physical Exams<br>To verify the hypotheses, the following would be helpful:<br>• Questions about the quantity and duration of supplement intake, other medications, and past medical history.<br>• Physical examination to assess hydration status, blood pressure, and signs of diabetic complications.<br><br>Expected to clarify: the potential cause of renal dysfunction, presence of dehydration, or signs of diabetes.</td></tr>
<tr><td colspan="2">Phase I – Q4: Plan for investigations/tests</td></tr>
<tr><td>DeepSeek-R1</td><td>Diagnostic Testing Plan
<table>
<tr><th>Test</th><th>Purpose</th><th>Expected Results</th></tr>
<tr><td>Urinalysis + sediment</td><td>Detect ATIN (WBC/eosinophils), ATN (muddy casts), or diabetes (glucose)</td><td>Eosinophils (ATIN); myoglobin/hemoglobin (rhabdo); glucosuria (diabetes)</td></tr>
<tr><td>Creatine kinase (CK)</td><td>Evaluate for rhabdomyolysis</td><td>Elevated CK (>1000 U/L) supports statin-induced myotoxicity</td></tr>
<tr><td>Serum glucose/HbA1c</td><td>Assess for diabetes</td><td>Elevated levels confirm hyperglycemia</td></tr>
<tr><td>Renal ultrasound</td><td>Rule out obstruction, assess kidney size</td><td>Normal size/absence of obstruction</td></tr>
</table>
Cost-effectiveness: Non-invasive tests first (urinalysis, CK, glucose). Avoid biopsy unless ATIN persists after supplement cessation.<br>Next, please enter the patient's test results.</td></tr>
<tr><td>Gemini 2.5 Pro</td><td>Diagnostic Testing Plan<br>1. Initial Blood and Urine Tests:<br>• Purpose: To confirm and characterize the renal dysfunction and look for clues to the etiology.<br>• Tests:</td></tr>
</table>

| | |
|---|---|
| | • **Comprehensive Metabolic Panel (CMP):** To quantify the serum creatinine/BUN and assess for electrolyte disturbances characteristic of tubular dysfunction (e.g., hypokalemia, non-anion gap metabolic acidosis).<br>• **Complete Blood Count (CBC) with differential:** To look for eosinophilia, which supports a diagnosis of allergic ATIN.<br>• **Urinalysis with microscopy:** This is a crucial first step. I expect to see evidence of tubular injury, such as sterile pyuria (white blood cells without bacteria), WBC casts, and low-grade proteinuria. The absence of significant hematuria or RBC casts would argue against a proliferative glomerulonephritis.<br>• **Urine Protein-to-Creatinine Ratio (UPCR) and Urine Beta-2 Microglobulin:** To quantify proteinuria (which should be sub-nephrotic) and directly measure tubular protein wasting, respectively. An elevated Beta-2 microglobulin would strongly support tubular damage.<br><br>**2. Imaging:**<br>• **Purpose:** To evaluate kidney structure and rule out other causes.<br>• **Test:**<br>• **Renal Ultrasound:** To assess kidney size (which should be normal or enlarged in an acute process) and rule out post-renal obstruction, which is an unlikely but important differential for acute kidney injury.<br>**3. Definitive Diagnosis (if needed):**<br>• **Purpose:** To establish a definitive diagnosis if renal function does not improve after stopping the supplement, or if the presentation is severe.<br>• **Test:**<br>• **Kidney Biopsy:** This is the gold standard. It would confirm the diagnosis of tubulointerstitial nephritis, help determine the cause (e.g., eosinophil-rich infiltrate in allergic AIN), assess the degree of chronic scarring (fibrosis) to predict prognosis, and definitively rule out a concomitant glomerular disease. |
| OpenAI GPT-o3 | Investigations<br>• Serum glucose & HbA1c – diagnose DM; expect marked hyperglycaemia and HbA1c ≥6.5 %.<br>• Basic metabolic panel (Na/K/Cl/$HCO_3$, BUN/Cr) and measured serum osmolality – define renal function, anion gap, hyperosmolar state.<br>• Urinalysis with dipstick & microscopy – look for glycosuria/ketonuria (DM), WBC/RBC/eosinophils or WBC casts (AIN), myoglobin (rhabdo).<br>• Serum calcium, phosphate, PTH – screen for hypercalcaemia; expect normal if DM, elevated Ca/PTH if hyperparathyroidism.<br>• Creatine kinase – exclude statin-like rhabdomyolysis from red-yeast-rice; expect normal in DM, elevated in rhabdo.<br>• Complete blood count with eosinophil count – eosinophilia suggests AIN.<br>• Renal ultrasound – assess kidney size and rule out obstruction; expect normal-sized kidneys without hydronephrosis.<br>Further tests (if initial work-up unrevealing): urine osmolality (DI vs osmotic diuresis), 24-h urine protein/albumin (diabetic nephropathy), kidney biopsy (suspected AIN). |
| Llama4-Marveric | Tests to Narrow Down the Diagnosis<br>**1. Serum Creatinine and Electrolytes:** To assess the severity of renal dysfunction.<br>**2. Blood Glucose and HbA1c:** To diagnose or rule out diabetes mellitus.<br>**3. Urinalysis:** To check for proteinuria, hematuria, or other abnormalities suggestive of kidney damage or disease.<br>**4. CK (Creatine Kinase):** To assess for potential muscle damage due to statin-like substances in the supplement. |

| | |
|---|---|
| | These tests will help differentiate between AKI due to supplement toxicity and diabetes mellitus, guiding further management. |
| **Phase II – Q5: Interpreting test results** | |
| DeepSeek-R1 | Interpretation of Test Results<br>**Key abnormalities and their significance:**<br>• **Proximal tubulopathy:**<br>• Massive glycosuria (4+) with normal serum glucose (99 mg/dL) and HbA1c (5.9%): Indicates **renal glycosuria**, signifying proximal tubular dysfunction.<br>• Severely elevated urinary β2MG (43,852 μg/L; normal <300) and α1MG (107 μg/L; normal <12): Confirms **proximal tubular damage,** as these low-molecular-weight proteins are reabsorbed here.<br>• Hypouricemia (1.2 mg/dL), hypokalemia (2.9 mEq/L), hypophosphatemia (2.0 mg/dL), and metabolic acidosis ($HCO_3$ 16.8 mmol/L): Classic **Fanconi syndrome** pattern due to proximal tubular wasting.<br>• **Renal dysfunction:**<br>• Elevated creatinine (2.12 mg/dL) with eGFR 29 mL/min/1.73m², yet normal-sized kidneys on CT: Suggests **acute/subacute injury** rather than chronic disease.<br>• **Exclusion of other causes:**<br>• Negative autoantibodies (ANA, ANCA, anti-GBM) and normal IgG4 rule out autoimmune glomerulonephritis or IgG4-related disease.<br>• Normal CK (504 IU/L) excludes rhabdomyolysis.<br>• Absence of hematuria/pyuria and normal CRP argue against acute interstitial nephritis or infection.<br><br>**Link to history:** The temporal association with **red yeast rice (statin analog) exposure** aligns with drug-induced proximal tubular toxicity. Statins are known to cause mitochondrial dysfunction in tubules, leading to Fanconi syndrome. |
| Gemini 2.5 Pro | Interpretation of Test Results<br>The laboratory results are highly significant and point conclusively to severe dysfunction of the renal tubules, specifically the proximal tubules, rather than a primary glomerular disease.<br>• **Hallmark of Proximal Tubulopathy (Fanconi Syndrome):** The combination of **hypouricemia** (1.2 mg/dL), **hypophosphatemia** (2.0 mg/dL), **hypokalemia** (2.9 mEq/L), **and proximal renal tubular acidosis** (venous pH 7.316, $HCO_3$ 16.8 mmol/L) constitutes acquired Fanconi syndrome. This is further cemented by the finding of **massive renal glycosuria** (4+ in urine) despite a normal serum glucose level (99 mg/dL), which indicates a failure of the proximal tubules to reabsorb glucose.<br>• **Confirmation by Urine Markers:** The extremely elevated urine **β2-microglobulin** (a low-molecular-weight protein normally reabsorbed by the proximal tubule) and elevated **NAG** (a tubular enzyme) provide direct and powerful evidence of severe tubular injury. The mild total proteinuria (1.4 g/gCr) is characteristic of this "tubular proteinuria" and argues against a significant glomerular pathology.<br>• **Exclusion of Other Diagnoses:** The urinalysis lacks significant hematuria or cellular casts, making glomerulonephritis unlikely. The comprehensive negative serological workup (ANA, ANCA, anti-GBM, anti-SSA/SSB, IgG4) effectively rules out the most common autoimmune and vasculitic causes of renal disease that were in the initial differential.<br>• **Acute Process:** The normal-sized kidneys on CT are consistent with an acute inflammatory process, such as acute tubulointerstitial nephritis (ATIN), and argue against a long-standing chronic kidney disease. The absence of peripheral eosinophilia does not rule out drug-induced ATIN, as this finding is only present in a minority of cases.<br><br>In summary, the tests reveal a severe acute kidney injury characterized by an acquired |

| | |
|---|---|
| | Fanconi syndrome, strongly implicating a toxin or drug that specifically targets the proximal tubules. |
| OpenAI GPT-o3 | Interpretation of the test results<br>• Serum Cr 2.12 mg/dL (eGFR 29) = acute/sub-acute kidney injury.<br>• Urine: 4+ glycosuria with normal plasma glucose (99 mg/dL, HbA1c 5.9 %) → renal, not diabetic, glycosuria.<br>• Very high urinary $\beta_2$-microglobulin (43 852 µg/L), $\alpha_1$-microglobulin, and NAG → severe proximal tubular cell injury.<br>• Hypouricaemia (UA 1.2 mg/dL), hypophosphataemia (2.0 mg/dL), hypokalaemia (2.9 mEq/L) and normal-anion-gap metabolic acidosis ($HCO_3$ 16.8 mmol/L, pH 7.32) → bicarbonate, phosphate, urate and potassium wasting, i.e. Fanconi syndrome / type 2 RTA.<br>• Mild tubular proteinuria (U-PCR 1.4 g/gCr) without haematuria; no casts or eosinophils → excludes glomerular disease and typical AIN.<br>• CK 504 IU/L is only mildly raised; rhabdomyolysis-driven AKI unlikely.<br>• Auto-antibodies, complement, IgG4, SSA/B all negative; CT shows normal kidney size → immune or chronic atrophic causes improbable.<br>Taken together, the profile is classic acquired Fanconi syndrome causing proximal renal tubular acidosis and secondary acute kidney injury. |
| Llama4-Marveric | Interpretation of Test Results<br>The patient's laboratory findings indicate renal dysfunction with a serum creatinine of 2.12 mg/dL and eGFR of 29 mL/min/1.73m². The presence of mild proteinuria, massive glycosuria, and significantly elevated urinary β2 microglobulin and N-acetyl-β-D-glucosaminidase (NAG) suggests proximal tubular dysfunction. The low serum levels of uric acid, potassium, phosphorus, and bicarbonate further support this, indicating a Fanconi syndrome-like presentation. The absence of significant hematuria or red blood cell casts and normal immunological tests (e.g., ANA, ANCA, anti-GBM antibody) make glomerulonephritis or vasculitis less likely. The normal IgG4 level and absence of SS-A and SS-B antibodies argue against IgG4-related disease or Sjögren's syndrome. |
| **Phase II – Q6: Reconsidering the differential diagnosis** | |
| DeepSeek-R1 | Revised Diagnostic Thinking<br>**Working Diagnosis: Acquired Fanconi syndrome due to red yeast rice (statin)-induced proximal tubular toxicity.**<br>**Reasoning for prioritization:**<br>• **Supporting evidence:**<br>• Proximal tubulopathy (glycosuria, hypophosphatemia, hypouricemia, metabolic acidosis) + elevated urinary β2MG/NAG.<br>• Temporal link to supplement use (symptoms began after 5 months of exposure).<br>• Exclusion of alternatives:<br> • No diabetes (normal HbA1c) despite glycosuria.<br> • Negative autoimmune serologies and normal complements rule out lupus/vasculitis.<br> • Normal CK and absence of myoglobinuria exclude rhabdomyolysis.<br>• **Differentials downgraded:**<br> • Diabetic nephropathy: Unlikely without hyperglycemia.<br> • Autoimmune tubulointerstitial nephritis: No eosinophilia, pyuria, or serologic markers. |
| Gemini 2.5 Pro | Updated Diagnostic Thinking and Working Diagnosis<br>The test results have dramatically narrowed the differential diagnosis. The initial suspicion of a supplement-induced tubulointerstitial process is now overwhelmingly supported by objective evidence of severe, isolated proximal tubular damage. The clinical picture is a textbook presentation of this entity.<br><br>The primary differential diagnoses, such as RPGN or autoimmune diseases (e.g., Sjögren's, Sarcoidosis), have been effectively ruled out by the negative serologies and the specific |

| | |
|---|---|
| | pattern of injury.<br><br>The temporal link to the red yeast rice supplement, combined with the classic presentation of acute tubulointerstitial nephritis with Fanconi syndrome, makes a causal relationship almost certain. Some red yeast rice preparations are known to be contaminated with citrinin, a mycotoxin with specific nephrotoxicity directed at the proximal tubule. This mechanism perfectly explains the patient's entire clinical and laboratory presentation.<br><br>**Working Diagnosis:** Red Yeast Rice Supplement-Induced Acute Tubulointerstitial Nephritis (ATIN) with associated Fanconi Syndrome. |
| OpenAI GPT-o3 | Revision of the differential and working diagnosis<br>The key new finding is isolated proximal tubular dysfunction. This shifts the differential toward toxin-induced or dysproteinaemia-associated Fanconi syndrome. The temporal relation to six months of red-yeast-rice supplementation (contains lovastatin analogue and the nephrotoxin citrinin) makes "red-yeast-rice-associated toxic proximal tubulopathy (acquired Fanconi syndrome)" the most likely diagnosis. Alternatives now lower in priority: light-chain myeloma (no anaemia, hyper-Ca, or immunoglobulin elevation), heavy-metal poisoning (no exposure history), drug-induced AIN (no eosinophilia, pyuria).<br><br>Working diagnosis: Red-yeast-rice supplement–induced acquired Fanconi syndrome with proximal renal tubular acidosis and acute kidney injury. |
| Llama4-Marveric | Updated Diagnostic Thinking<br>Given the evidence of proximal tubular dysfunction and the absence of significant glomerular disease or immunological abnormalities, the most likely diagnosis is **Acquired Fanconi Syndrome.** The patient's history of consuming red yeast rice supplements, which contain statin-like substances, could be relevant, as certain drugs and toxins are known to cause Fanconi syndrome. Other causes, such as multiple myeloma or other paraproteinemia, should also be considered, although the normal serum protein electrophoresis pattern (not directly shown but inferred from the provided data) makes this less likely. |
| **Phase II – Q7: Treatment planning** | |
| DeepSeek-R1 | Treatment Plan<br>**Immediate Interventions:**<br>**1. Discontinue red yeast rice supplement:** Critical to remove the nephrotoxic trigger.<br>**2. Electrolyte repletion:**<br>• Oral potassium chloride (e.g., 40–80 mEq/day) for hypokalemia (K 2.9 mEq/L).<br>• Oral phosphorus (e.g., sodium phosphate 250–500 mg QID) for hypophosphatemia.<br>• Sodium bicarbonate (e.g., 650 mg TID) for metabolic acidosis ($HCO_3$ 16.8 mmol/L).<br>**3. Renoprotection:**<br>• ACE inhibitor (e.g., lisinopril 2.5 mg/day) for mild proteinuria (1.4 g/gCr), after correcting hypokalemia.<br><br>**Rationale:**<br>• **Evidence basis:** Statin-induced tubular toxicity is typically reversible upon discontinuation (EBM: case series, Nephrol Dial Transplant 2011).<br>• Electrolyte correction prevents arrhythmias (hypokalemia) and osteomalacia (hypophosphatemia).<br>• ACE inhibitors reduce proteinuria and slow CKD progression (EBM: NKF KDOQI guidelines).<br><br>**Alternative Options:**<br>• If no improvement in 4–6 weeks: Consider kidney biopsy to exclude rare causes (e.g., monoclonal gammopathy).<br>• Avoid NSAIDs/nephrotoxins to prevent additional injury. |

| | |
|---|---|
| | **Monitoring:**<br>• Weekly electrolytes, renal function, and urinary protein until stable.<br>• Long-term: Assess for residual tubular dysfunction after 3 months. |
| Gemini 2.5 Pro | Treatment Plan<br>The treatment strategy is two-pronged: removing the cause and managing its consequences, with the goal of preserving renal function and preventing chronic fibrosis.<br><br>**1. Primary Intervention:**<br>• **Immediate and permanent discontinuation of the red yeast rice supplement.** This is the single most critical action, as removing the offending agent is essential for recovery.<br>**2. Supportive Care to Correct Fanconi Syndrome:**<br>• **Correction of Acidosis:** Initiate oral alkali therapy (e.g., sodium bicarbonate, 3-6 g/day in divided doses) to correct the metabolic acidosis, with a target serum bicarbonate level of >22 mmol/L.<br>• **Electrolyte Repletion:** Provide oral supplementation for potassium (potassium chloride) and phosphate to correct hypokalemia and hypophosphatemia, with frequent monitoring.<br>• **Hydration:** Advise adequate fluid intake to compensate for urinary losses from glycosuria and impaired concentrating ability.<br>**3. Specific Therapy for ATIN:**<br>• **Corticosteroid Therapy:** Given the severity of the renal dysfunction (eGFR 29 mL/min/1.73m²), I recommend initiating systemic corticosteroids to suppress the interstitial inflammation and reduce the risk of progression to irreversible interstitial fibrosis.<br>• **Rationale:** While high-quality RCTs are limited, observational data and clinical consensus strongly support the use of steroids for severe drug-induced ATIN to improve the chances of renal recovery.<br>• **Regimen:** Start oral prednisolone at a dose of 0.5-1 mg/kg/day (e.g., 40-50 mg daily). The duration will be guided by the response of the serum creatinine, followed by a slow taper over 2-3 months.<br>**4. Definitive Diagnosis and Prognosis:**<br>• **Kidney Biopsy:** I would strongly recommend a kidney biopsy.<br>• **Rationale:** Although the clinical diagnosis is very clear, a biopsy will provide 100% diagnostic certainty before committing the patient to a multi-month course of steroids. Most importantly, it will allow for the assessment of interstitial fibrosis and inflammation, which is the best predictor of the patient's long-term prognosis and potential for renal recovery. Treatment can be initiated while awaiting the biopsy results.<br><br>**Alternative Options:**<br>If the patient does not respond to corticosteroids or has a contraindication, a second-line immunosuppressant such as mycophenolate mofetil could be considered, although evidence for its use in this specific context is less robust. However, this is not the initial plan. The primary focus is on supplement cessation, supportive care, and corticosteroid therapy. |
| OpenAI GPT-o3 | Treatment plan and rationale<br>Immediate withdrawal of the suspected agent<br>• Stop red-yeast-rice supplement; counsel the patient about avoiding all non-prescribed nutraceuticals.<br>Supportive correction of tubular losses<br>• Potassium chloride replacement to keep $K^+$ ≥3.5 mEq/L.<br>• Oral/IV phosphate (e.g. 30–60 mmol/day) to prevent osteomalacia and muscle weakness.<br>• Alkali therapy: sodium bicarbonate or citrate 1–2 mEq/kg/day titrated to $HCO_3$ ≈22 mmol/L. |

<table>
<tr><td></td><td>Hydration and kidney-function monitoring<br>• Isotonic fluids as needed; daily weights, input/output, serum creatinine, electrolytes and acid–base.<br>Nephrology follow-up and further evaluation<br>• If renal function fails to improve within 1–2 weeks or deteriorates, proceed to kidney biopsy to confirm toxic tubulointerstitial injury and rule out occult myeloma or other pathology.<br>Consider adjunct options if biopsy shows active interstitial nephritis<br>• Oral prednisone 0.5–1 mg/kg/day with taper (GRADE low-moderate evidence) may hasten recovery.<br>Patient education and long-term care<br>• Bone protection (vitamin D, monitoring of bone density) if phosphate wasting persists.<br>• Periodic renal, electrolyte and acid–base surveillance over the next 6–12 months.<br><br>Alternative/less-likely strategies<br>• If a monoclonal disorder is later identified, treat the underlying haematologic disease.<br>• Chelation in proven heavy-metal poisoning.<br>• No role for immunosuppression unless biopsy documents immune-mediated interstitial nephritis.<br>Goal: eliminate toxin, correct biochemical derangements, and preserve renal function; most cases of toxic Fanconi syndrome show partial to full recovery within months when managed promptly.</td></tr>
<tr><td>Llama4-Marveric</td><td>Treatment Plan<br>1. Discontinue the Offending Agent: Stop the red yeast rice supplements immediately, as they are a potential cause of the renal dysfunction.<br>2. Supportive Care: Manage the patient's electrolyte imbalances (e.g., potassium, phosphorus) and acid-base disturbances with supplementation as needed.<br>3. Monitoring: Closely monitor renal function, electrolytes, and urinary abnormalities to assess for improvement or deterioration.<br>4. Further Evaluation: If renal function does not improve after discontinuing the supplement, consider further evaluation for other causes of Fanconi syndrome.<br><br>Rationale: The treatment is based on removing the potential cause, managing symptoms, and monitoring for recovery or progression. Alternative options include further diagnostic testing if the patient's condition does not improve after stopping the supplement.</td></tr>
<tr><td colspan="2">Phase III – Q8: Evaluation of treatment</td></tr>
<tr><td>DeepSeek-R1</td><td>Monitoring Treatment Effectiveness and Safety<br>Effectiveness Monitoring:<br>• Renal function:<br>• Measure serum creatinine/eGFR weekly ×4 weeks, then monthly ×3 months (target: stable eGFR >60 mL/min/1.73$m^2$).<br>• Tubular function:<br>• Weekly electrolytes (potassium, bicarbonate, phosphorus) and uric acid until normal (target: K >3.5 mEq/L, $HCO_3$ >22 mmol/L, P >2.5 mg/dL).<br>• Urinary biomarkers (U-β2MG, U-α1MG, U-NAG) monthly (target: decline to normal: β2MG <300 μg/L, α1MG <12 mg/L, NAG <12 IU/L).<br>• Proteinuria/glycosuria:<br>• Urinary protein-to-creatinine ratio (U-PCR) and glucose weekly until resolution (target: U-PCR <0.15 g/gCr; U-Glu negative).<br><br>Safety Monitoring & Risks:
<table>
<tr><th>Risk</th><th>Monitoring Method</th><th>Concrete Countermeasures</th></tr>
<tr><td>Steroid toxicity</td><td>Fasting glucose weekly; BP</td><td></td></tr>
</table>
</td></tr>
</table>

<table>
<tr><td></td><td>
<table>
<tr><td></td><td>daily; weight/biweekly</td><td></td></tr>
<tr><td>Hypokalemia</td><td>Serum K+ weekly</td><td>Oral KCl (40–80 mEq/day) if K <3.5 mEq/L.</td></tr>
<tr><td>Infection</td><td>WBC count biweekly; fever monitoring</td><td>If WBC >12,000/μL or fever: culture-driven antibiotics; avoid live vaccines.</td></tr>
<tr><td>Relapse</td><td>Urinary β2MG monthly</td><td>If β2MG rises: recheck for supplement re-exposure or new toxins.</td></tr>
</table>
</td></tr>
<tr><td>Gemini 2.5 Pro</td><td>
List specific measures for evaluating treatment effectiveness and potential side effects or complications, and explain the monitoring and management plan for them.<br>
Monitoring Treatment Effectiveness:<br>
The goal is to confirm sustained recovery of renal function and resolution of tubular damage.<br>
• Renal Function: Monitor serum creatinine and eGFR weekly during the steroid taper, then monthly. The objective is to ensure the eGFR remains stable at the recovered baseline (around 64 mL/min/1.73 m²).<br>
• Tubular Function:<br>
• Monitor serum electrolytes (potassium, phosphorus, bicarbonate) and urine glucose at the same frequency. The goal is to see them remain within the normal range without supplementation.<br>
• Periodically measure urine β2-microglobulin (U-β2mg). This is the most sensitive marker of ongoing tubular injury. A continued low level confirms recovery, while a rise would indicate a potential relapse.<br>
• Proteinuria: Monitor the urinary protein-to-creatinine ratio (U-PCR) to ensure it remains negligible.<br><br>
Monitoring Safety and Managing Risks:<br>
The primary risks are associated with corticosteroid therapy, even during a taper.<br>
• Risk 1: Metabolic Complications (Hyperglycemia, Hypertension)<br>
• Monitoring: Check blood pressure at every visit and monitor blood glucose, especially given the initial high-normal reading.<br>
• Management: Advise on diet and exercise. If blood pressure remains elevated (e.g., >130/80 mmHg), initiate antihypertensive therapy (e.g., an ACE inhibitor or ARB).<br>
• Risk 2: Increased Infection Susceptibility<br>
• Monitoring: Inquire about symptoms of infection (fever, cough, etc.) at each follow-up.<br>
• Management: Educate the patient to seek prompt medical attention if signs of infection occur.<br>
• Risk 3: Gastric Injury<br>
• Monitoring: Ask about symptoms of dyspepsia or abdominal pain.<br>
• Management: A proton pump inhibitor (PPI) or H2 blocker should be co-administered during the initial high-dose steroid phase and considered throughout the taper to prevent gastritis.<br>
• Risk 4: Relapse of Acute Tubular Injury<br>
• Monitoring: A sudden rise in creatinine or the reappearance of hypokalemia, hypophosphatemia, or glycosuria during the steroid taper would signal a relapse.<br>
• Management: If relapse occurs, the steroid taper would be halted, and the dose may need to be increased again before attempting a slower taper.
</td></tr>
<tr><td>OpenAI GPT-o3</td><td>
Monitoring plan: effectiveness vs. safety<br>
• Kidney function<br>
− Serum creatinine/eGFR: daily during admission, then weekly until plateau, thereafter
</td></tr>
</table>

| | |
|---|---|
| | every 1–3 months.<br>− Urine output, weight, BP each shift; FeNa if oliguria appears.<br>• Proximal-tubule recovery<br>− Serum electrolytes, uric acid, phosphorus, bicarbonate, anion gap: q24–48 h → weekly → monthly.<br>− Urinary $\beta_2$-microglobulin, $\alpha_1$-microglobulin, NAG, protein-to-creatinine ratio, glucose: weekly until normal, then every visit.<br>• Histologic correlation<br>− No repeat biopsy unless course atypical.<br>• Clinical symptoms<br>− Resolution of thirst, polyuria, fatigue; functional status at each visit.<br><br>Specific risks & countermeasures<br>1. Steroid-related<br>• Hyperglycaemia: fasting/PP glucose q24 h → adjust diet/insulin.<br>• Infection: daily T°, WBC; educate on early reporting; vaccinate and consider PCP prophylaxis if >20 mg/day >4 weeks.<br>• Hypertension/fluid retention: BP, weight; add antihypertensive/diuretic PRN.<br>• GI ulcer: PPI co-therapy.<br>• Psychiatric disturbance: mental-status check; dose adjust if severe.<br>• Bone loss: Ca/Vit-D, weight-bearing exercise; baseline and 12-mo DXA; bisphosphonate if T-score $\leq -1.0$.<br>2. Relapse or progression of tubular injury<br>• Strict avoidance of red-yeast-rice or other nephrotoxins; provide written list.<br>• Early lab triggers: rising $\beta_2$-MG, creatinine or falling bicarbonate prompt same-day review.<br>3. Electrolyte over-correction<br>• $K^+$ and Pi rechecked 4–6 h after IV replacement; ECG for $K^+$ >5.5 mEq/L.<br><br>Steroid taper adrenal insufficiency<br>• Educate on stress dosing; AM cortisol if symptoms. |
| Llama4-Marveric | To monitor the effectiveness and safety of the treatment, the following measures will be taken:<br>• **Renal Function:** Regularly check serum creatinine and eGFR to assess the improvement or deterioration of renal function.<br>• **Electrolytes and Acid-Base Balance:** Monitor serum potassium, phosphorus, uric acid, and bicarbonate levels to ensure they remain within normal ranges.<br>• **Urinary Abnormalities:** Continue to assess urinary glucose, protein, β2 microglobulin, and NAG to evaluate the recovery of proximal tubular function.<br>• **Steroid Therapy Side Effects:** Monitor for potential side effects of steroid therapy, such as hyperglycemia, hypertension, and osteoporosis.<br><br>Specific risks to watch out for include:<br>• **Steroid-induced complications:** such as diabetes, hypertension, or osteoporosis.<br>• **Relapse of renal dysfunction:** if the steroid dose is tapered too quickly.<br><br>Countermeasures include:<br>• **Gradual tapering of steroids:** to minimize the risk of relapse.<br>**Regular monitoring:** for signs of steroid-induced complications. |
| **Phase III – Q9: Formulating plans for worsening conditions** | |

| | |
|---|---|
| DeepSeek-R1 | Response to Unexpected Changes<br>**Thought Process:**<br>**1. If no improvement (e.g., persistent eGFR <45 after 4 weeks):**<br>• Reassess diagnosis:<br>• Rule out underlying chronic disease (e.g., diabetic nephropathy in Case 2) via HbA1c, renal ultrasound.<br>• Exclude competing diagnoses (e.g., light-chain tubulopathy) with serum/urine protein electrophoresis.<br>• Evaluate biopsy adequacy: Repeat biopsy if initial sample missed focal lesions (e.g., interstitial fibrosis).<br>**2. If sudden deterioration (e.g., eGFR drop >25%):**<br>• Check for new insults: Infection (urine culture), dehydration (FeNa), or drug error (supplement reuse).<br>• Assess for complications: Steroid-induced diabetes (glucose/HbA1c) or electrolyte crisis (K <2.5 mEq/L).<br>**Action Plan:**<br>• **Immediate steps:**<br>• Discontinue steroids if infection or severe hyperglycemia occurs.<br>• Hospitalize for IV electrolytes if K <2.5 mEq/L or $HCO_3$ <15 mmol/L.<br>• **Diagnostic revision:**<br>• If tubular injury persists: Screen for heavy metals (e.g., cadmium) or paraproteins.<br>• If inflammation emerges: Re-biopsy to assess for delayed interstitial nephritis.<br>• **Treatment adjustment:**<br>• Switch to renal-replacement therapy if eGFR <15 (as in Case 2).<br>• For steroid-unresponsive ATIN: Trial mycophenolate (500 mg BID) after infection exclusion.<br><br>**Rationale:** Prioritize reversible causes (e.g., toxins, electrolyte imbalance) before escalating immunosuppression.<br>This concludes the assessment of clinical reasoning skills. If you wish to conduct an evaluation with another case, please open a new chat. |
| Gemini 2.5 Pro | **Explain the thought process and action plan for rapidly reassessing the cause in response to changes in the patient's condition, and for flexibly revising the diagnosis and treatment plan.**<br>If the patient's renal function deteriorates again or fails to maintain its improvement, my thinking process and actions would be as follows:<br><br>**Thought Process (Differential Diagnosis for Worsening Renal Function):**<br>**1. Relapse of the Original Disease:** The most likely cause is a relapse of the acute tubular injury as the steroid dose is reduced. The underlying inflammatory process may not have been fully extinguished.<br>**2. New Nephrotoxic Insult:** I would consider exposure to a new drug (e.g., NSAIDs, antibiotics) or supplement.<br>**3. Progression to Chronic Disease:** The initial acute injury may have resulted in significant interstitial fibrosis, leading to progressive chronic kidney disease (CKD), as hinted at by the poor outcome in the second case presented.<br>**4. Unrelated Acute Kidney Injury:** I would consider other causes of AKI, such as pre-renal (e.g., volume depletion) or post-renal (obstruction) causes, though these are less likely.<br><br>**Action Plan (Reassessment and Management Revision):**<br>**1. Immediate Re-evaluation:**<br>• **History:** Take a detailed interval history, focusing on new medications, supplements, |

| | |
|---|---|
| | symptoms of infection, or systemic illness. Confirm there has been no re-exposure to red yeast rice.<br>• **Physical Exam:** Assess volume status and blood pressure.<br>**2. Urgent Laboratory Testing:**<br>• Repeat serum creatinine, electrolytes, and bicarbonate to confirm the decline and check for recurrence of Fanconi syndrome.<br>• Perform a fresh urinalysis with microscopy and repeat the U-PCR and U-β2mg. A return of sterile pyuria, tubular proteinuria, and high U-β2mg would strongly support a relapse of the original tubular injury.<br>**3. Flexible Revision of the Treatment Plan:**<br>• **If Relapse is Confirmed:** The immediate action would be to increase the prednisolone dose back to a therapeutic level (e.g., 20-30 mg/day) to regain control, followed by a much slower taper schedule once renal function has re-stabilized.<br>• **If Cause is Unclear or No Response:** If the clinical picture is ambiguous or does not respond to increased steroids, a **repeat kidney biopsy** becomes essential. This would be the definitive step to distinguish between active, treatable inflammation and established, irreversible fibrosis. Comparing the second biopsy to the first would provide crucial information on disease progression and guide long-term strategy, determining whether to pursue further immunosuppression or transition to supportive CKD management (e.g., strict blood pressure control, RAAS inhibition). |
| OpenAI GPT-o3 | Approach if course deviates<br>Step 1: Verify data & adherence<br>• Confirm supplement discontinuation, medication list, lab accuracy, hydration status.<br>Step 2: Re-evaluate differential<br>• Repeat history/exam for new toxins, infections, systemic signs.<br>• Labs: serum/urine light chains, SPEP/UPEP (light-chain Fanconi), heavy-metal screen, viral serologies.<br>• Imaging for obstruction or new parenchymal change.<br>Step 3: Renal reassessment<br>• If creatinine rises or tubular markers worsen, consider repeat biopsy to look for evolving interstitial nephritis, light-chain deposition, or alternative pathology.<br>Step 4: Adjust treatment<br>• If biopsy shows active immune injury → escalate immunosuppression (full-dose prednisone 1 mg/kg ± mycophenolate).<br>• If toxic or dysproteinaemic process identified → remove/chelate toxin, initiate haematological therapy.<br>• Intensify supportive care: optimise fluids, correct electrolytes, prepare for renal replacement therapy if refractory.<br>Step 5: Multidisciplinary escalation<br>• Early nephrology, endocrinology (bone/mineral), and infectious-disease input; patient education reinforced at each pivot point. |
| Llama4-Marveric | Response to Unexpected Changes or Lack of Improvement<br>If the patient's condition does not improve as expected or suddenly changes:<br>**1. Reassess the Diagnosis:** Consider alternative causes of renal dysfunction or Fanconi syndrome.<br>**2. Review Medication Adherence:** Ensure the patient has stopped taking the red yeast rice supplements.<br>**3. Adjust Treatment:** Modify the steroid regimen or consider other treatments based on the reassessment.<br><br>The thought process involves:<br>• Re-evaluating the initial diagnosis and potential causes.<br>• Assessing for new or missed diagnoses. |

| | |
|---|---|
| | • Adjusting the treatment plan accordingly.<br><br>By following these steps, the effectiveness and safety of the treatment can be optimized, and any unexpected changes or lack of improvement can be promptly addressed. |

| **Case 7: Nocardiosis in a Patient with Nephrotic Syndrome Treated with Glucocorticoids and Tacrolimus** [8] | |
|---|---|
| **Phase I – Q1: Summarizing medical problems** | |
| DeepSeek-R1 | 86–90-year-old woman with chronic rheumatoid arthritis (on tacrolimus) and IgA-λ smoldering multiple myeloma (SMM), presenting with subacute (1-year) severe leg edema, hypoalbuminemia (2.0–3.0 g/dL), massive proteinuria, and nephrotic syndrome (NS) since January 2021. |
| Gemini 2.5 Pro | This is an 86–90-year-old woman with a background of IgA-λ smoldering multiple myeloma and rheumatoid arthritis, presenting with a one-year, progressive course of nephrotic syndrome. The key clinical problem is to identify the specific glomerular pathology causing her massive proteinuria, given her two pre-existing conditions known to cause kidney disease. |
| OpenAI GPT-o3 | 86–90-year-old woman with long-standing rheumatoid arthritis on tacrolimus and IgA-λ smoldering myeloma who, over the past year, has developed massive proteinuria, marked hypoalbuminemia (2–3 g/dL) and severe bilateral leg edema → new-onset nephrotic syndrome. |
| Llama4-Marveric | The patient is an 86–90-year-old woman with a history of rheumatoid arthritis and IgA-λ type smoldering multiple myeloma (SMM), presenting with severe leg edema, hypoalbuminemia (2.0-3.0 g/dL), and massive proteinuria, diagnosed with nephrotic syndrome (NS) in January 2021. |
| **Phase I – Q2: Differential diagnoses and rationale** | |
| DeepSeek-R1 | Differential Diagnoses<br>• **Most Likely: Amyloid light-chain (AL) amyloidosis** secondary to SMM. Rationale: SMM (IgA-λ type) is a known cause of AL amyloidosis, which commonly presents with NS, hypoalbuminemia, and edema.<br>• **Alternative 1: Monoclonal immunoglobulin deposition disease (MIDD).** Rationale: Associated with plasma cell dyscrasias like SMM; can cause glomerular injury and NS.<br>• **Alternative 2: Tacrolimus-induced nephrotoxicity.** Rationale: Chronic calcineurin inhibitor use may cause glomerular/thrombotic microangiopathy, though NS is atypical.<br>• **Must Not Miss: AL amyloidosis or MIDD.** Rationale: Both are severe complications of SMM with high mortality if untreated. Excluding these is critical due to disease progression risk. |
| Gemini 2.5 Pro | Differential Diagnosis<br>**1. AL (Light Chain) Amyloidosis:** This is the **most likely** diagnosis. Nephrotic syndrome is a classic presentation of AL amyloidosis, which is directly caused by the deposition of monoclonal light chains. The patient's diagnosis of IgA-λ smoldering myeloma provides the necessary substrate (lambda light chains) for this disease.<br>**2. AA (Secondary) Amyloidosis:** This is another strong possibility. Long-standing, chronic inflammatory conditions like rheumatoid arthritis are the primary cause of AA amyloidosis, which results from the deposition of serum amyloid A protein and also typically presents with nephrotic syndrome.<br>**3. Membranous Nephropathy:** This is a common cause of nephrotic syndrome in the elderly. While it can be idiopathic, it is also known to be associated with autoimmune diseases like rheumatoid arthritis.<br><br>**Must-Not-Miss Diagnosis:**<br>**AL Amyloidosis** is the must-not-miss diagnosis. It is not only the most likely but also the most aggressive of these conditions, often leading to rapid multi-organ failure. A definitive diagnosis is critical as it would necessitate urgent, clone-directed chemotherapy to target |

| | |
|---|---|
| | the underlying plasma cell dyscrasia, which is fundamentally different from the treatment for AA amyloidosis or membranous nephropathy. |
| OpenAI GPT-o3 | Differential diagnosis<br>a. Secondary membranous nephropathy – common cause of nephrotic syndrome in the elderly; can be triggered by RA, tacrolimus, or an underlying malignancy.<br>b. AL (light-chain) amyloidosis from λ monoclonal protein – fits age, myeloma spectrum disease and produces heavy proteinuria with low-selectivity. (Most likely given λ clone.)<br>c. AA amyloidosis from chronic inflammatory RA.<br>Must-not-miss: AL amyloidosis (rapidly progressive, systemic, treatable if caught early). |
| Llama4-Marveric | Differential Diagnoses<br>**1. Amyloidosis:** Given her history of SMM and NS, amyloidosis is a strong consideration due to the association between plasma cell dyscrasias and amyloid deposition in the kidneys.<br>**2. Membranous Nephropathy (MN):** This is a common cause of NS in adults, and could be primary or secondary due to her rheumatoid arthritis or other factors.<br>**3. Minimal Change Disease (MCD):** Although less likely in this age group, MCD can occur in elderly patients and should be considered.<br><br>The most likely diagnosis is **Amyloidosis** due to her history of SMM and the presence of NS.<br>What must not be missed is **Amyloidosis** or other serious conditions like MN that require prompt diagnosis and treatment. |
| **Phase I – Q3: Necessary physical examinations and reasons** | |
| DeepSeek-R1 | Key History/Physical Exam<br>• **History Questions:**<br>• Edema progression: "Did edema start suddenly or gradually?" (Clarifies time course).<br>• Systemic symptoms: "Any weight loss, fatigue, or orthopnea?" (Assesses cardiac/renal involvement in amyloidosis).<br>• Tacrolimus adherence/duration: "How long have you taken tacrolimus? Any recent dose changes?" (Evaluates drug toxicity).<br><br>• **Physical Exam:**<br>• Pericardial rub or hepatomegaly (Suspect amyloid infiltration).<br>• Blood pressure (Hypertension may support MIDD).<br>• Skin/nail changes (Purpura or macroglossia suggests amyloidosis).<br><br>• **Purpose:** Differentiate AL amyloidosis (systemic signs) from tacrolimus toxicity (hypertension) or MIDD. |
| Gemini 2.5 Pro | Additional History and Physical Examination<br>**Additional Questions:**<br>• **Systemic Amyloidosis Symptoms:** "Have you experienced unusual bruising (especially around the eyes), an enlarged tongue, difficulty swallowing, dizziness upon standing, or numbness/tingling in your hands and feet?"<br>• **Purpose:** To screen for classic signs of systemic AL amyloidosis, such as periorbital purpura, macroglossia, and autonomic or peripheral neuropathy.<br>• **Rheumatoid Arthritis Activity:** "How have your joints been recently? Have you had increased pain, swelling, or morning stiffness?"<br>• **Purpose:** To assess the level of inflammatory activity from her RA. Poorly controlled, active RA would increase the suspicion for AA amyloidosis.<br>• **Constitutional Symptoms:** "Have you had any recent bone pain, significant fatigue, or unexplained weight loss?"<br>• **Purpose:** To screen for symptoms of progression from smoldering myeloma to active multiple myeloma (CRAB criteria). |

| | |
|---|---|
| | **Physical Examination:**<br>• **Orthostatic Vital Signs:** Measure blood pressure and heart rate when supine and after standing for 3 minutes.<br>• **Purpose:** To detect orthostatic hypotension, a sign of autonomic neuropathy common in AL amyloidosis.<br>• **Head and Neck Examination:** Inspect the tongue for enlargement (macroglossia) and the skin around the eyes for purpura.<br>• **Purpose:** To identify pathognomonic physical signs of AL amyloidosis.<br>• **Cardiovascular Examination:** Assess for an elevated jugular venous pressure (JVP) and listen for S3 or S4 heart sounds.<br>• **Purpose:** To screen for evidence of restrictive cardiomyopathy, a common and life-threatening complication of cardiac amyloidosis. |
| OpenAI GPT-o3 | Key history & examination points<br>• Duration/progression of edema, weight gain, foamy urine → quantify timeline/severity.<br>• Constitutional or myeloma-related symptoms (fatigue, bone pain, anemia, hypercalcemia) → gauge plasma-cell activity.<br>• Orthostatic dizziness, paresthesias, macroglossia, easy bruising, heart failure signs → systemic AL amyloid involvement.<br>• RA activity, biologic/NSAID use, infections, malignancy screening → secondary membranous/AA triggers.<br>• Physical exam: blood pressure, orthostatic BP, jugular venous pressure, cardiac murmurs, hepatosplenomegaly, neuropathy, purpura, macroglossia, joint deformities → differentiate membranous vs amyloid and assess volume status. |
| Llama4-Marveric | Additional Questions and Physical Exams<br>To verify the hypotheses, additional questions and physical exams could include:<br>• Detailed history of her rheumatoid arthritis treatment and disease activity.<br>• Presence of other symptoms such as weight loss, fatigue, or macroglossia that could suggest amyloidosis.<br>• Physical examination for signs of amyloidosis (e.g., macroglossia, purpura) or rheumatoid arthritis activity.<br>• Urine analysis for proteinuria quantification and examination for casts or other abnormalities.<br>These would help clarify the potential causes of NS and the systemic involvement of her known conditions. |
| **Phase I – Q4: Plan for investigations/tests** | |
| DeepSeek-R1 | Diagnostic Testing Plan<br>• **Urine protein-to-creatinine ratio (UPCR):** Purpose: Quantify proteinuria severity. Expected: Confirms nephrotic-range (>3.5 g/g).<br>• **Serum free light chains (FLC):** Purpose: Assess λ-light chain burden (SMM progression). Expected: Elevated λ-FLC, abnormal κ:λ ratio.<br>• **Renal biopsy:** Purpose: Gold standard for diagnosing AL amyloidosis (Congo red+ deposits) or MIDD (granular deposits). Expected: Amyloid fibrils or monoclonal deposits.<br>• **SPEP/IFIX:** Purpose: Monitor IgA-λ paraprotein. Expected: Persistence of monoclonal band.<br>• **Rationale:** UPCR/FLC are non-invasive first steps; biopsy is definitive but invasive. Prioritizes cost-effectiveness and diagnostic yield. |
| Gemini 2.5 Pro | Diagnostic Test Plan<br>**1. Blood and Urine Monoclonal Protein Studies:**<br>• **Tests:** Serum Free Light Chain (sFLC) assay with kappa/lambda ratio, Serum and Urine Protein Electrophoresis (SPEP/UPEP) with immunofixation. |

| | |
|---|---|
| | • **Purpose:** To quantify the monoclonal protein burden. A significantly abnormal sFLC ratio (skewed towards lambda) would strongly support a plasma cell dyscrasia-driven process like AL amyloidosis. Urine immunofixation will detect monoclonal free light chains (Bence-Jones proteinuria).<br>• **Expected Results:** In AL amyloidosis, I expect a markedly elevated lambda free light chain level, a suppressed kappa level, and a severely abnormal kappa/lambda ratio. The UPEP will likely be positive for lambda light chains.<br>**2. Inflammatory and Autoimmune Markers:**<br>• **Tests:** C-reactive protein (CRP), Serum Amyloid A (SAA), and anti-Phospholipase A2 Receptor (PLA2R) antibody.<br>• **Purpose:** To differentiate between potential causes.<br>• **Expected Results:** Markedly elevated CRP and SAA would support a high inflammatory state, increasing the likelihood of AA amyloidosis. A positive anti-PLA2R antibody would diagnose primary membranous nephropathy.<br>**3. Kidney Biopsy with Special Stains:**<br>• **Test:** Percutaneous kidney biopsy for light microscopy (LM), immunofluorescence (IF), and electron microscopy (EM).<br>• **Purpose:** This is the gold standard test to establish a definitive diagnosis by directly examining the kidney tissue.<br>• **Expected Results:**<br>• **AL Amyloidosis:** LM will show amorphous deposits that are positive with Congo Red stain and exhibit apple-green birefringence under polarized light. IF will stain positive for lambda light chains but not kappa. EM will show randomly arranged, non-branching fibrils (8-12 nm).<br>• **AA Amyloidosis:** Similar findings on LM (Congo Red positive), but IF will be negative for light chains. Staining for Serum Amyloid A would be positive.<br>• **Membranous Nephropathy:** LM would show thickened basement membranes. IF would show granular staining for IgG and C3 along the capillary walls. EM would show subepithelial deposits.<br>**4. Systemic Staging (if amyloidosis is confirmed):**<br>• **Tests:** Echocardiogram and NT-proBNP; Abdominal fat pad aspiration; Bone marrow biopsy.<br>• **Purpose:** To assess for systemic involvement (especially cardiac, which dictates prognosis) and to quantify the underlying plasma cell burden to distinguish smoldering from active myeloma.<br>• **Expected Results:** An echocardiogram may show a "sparkling" myocardium and diastolic dysfunction. Fat pad aspiration may be positive for amyloid, offering a less invasive confirmation. Bone marrow biopsy will clarify the percentage of clonal plasma cells. |
| OpenAI GPT-o3 | Investigations and rationale<br>• Urinalysis & 24-h protein quantification – confirm nephrotic-range protein, look for hematuria/casts; expect >3.5 g/day, bland sediment.<br>• Serum/urine protein electrophoresis + immunofixation, serum free light chains – detect monoclonal λ light chain; anticipate abnormal λ excess if AL amyloidosis.<br>• Kidney biopsy (light, immunofluorescence, EM) – gold standard to distinguish membranous, AL amyloid, AA amyloid, light-chain deposition; expect Congo-red positive deposits with λ dominance if AL amyloid.<br>• Serum creatinine, eGFR, lipids, coagulation profile – baseline renal function, NS complications.<br>• Complements C3/C4, ANA, anti-$PLA_2R$, hepatitis B/C, HIV – evaluate for primary vs secondary membranous.<br>• Echocardiography and NT-proBNP – assess cardiac amyloid; thickened ventricular walls |

| | |
|---|---|
| | with restrictive physiology would support AL amyloidosis.<br>• Renal ultrasound – kidney size; enlarged normal-echo kidneys suggest amyloid/light-chain deposition.<br><br>These targeted tests will discriminate between AL amyloidosis, membranous nephropathy, and AA amyloidosis, allowing timely disease-specific therapy. |
| Llama4-Marveric | Tests to Narrow Down the Diagnosis<br>**1. Kidney Biopsy:** To definitively diagnose the cause of NS, such as amyloidosis, MN, or MCD. Expected result: Histopathological diagnosis.<br>**2. Serum and Urine Immunofixation:** To assess for paraproteins associated with SMM or progression to multiple myeloma. Expected result: Presence or absence of monoclonal proteins.<br>**3. SAP Scan or Bone Marrow Biopsy:** To diagnose and assess the extent of amyloidosis. Expected result: Evidence of amyloid deposits.<br>**4. Rheumatoid Arthritis Activity Tests:** To assess the activity of her rheumatoid arthritis. Expected result: Indicators of disease activity.<br>These tests will help differentiate among the potential causes of NS and guide further management. |
| **Phase II – Q5: Interpreting test results** | |
| DeepSeek-R1 | Interpretation of Test Results<br>**Key points and clinical significance:**<br>• **Nephrotic-range proteinuria (UP/UCr 5.57–8.23 g/gCr) with severe hypoalbuminemia (albumin 1.9–2.4 g/dL):** Confirms nephrotic syndrome (NS). The low selectivity index (0.08) suggests selective proteinuria, typical of minimal change disease (MCD) or early-stage amyloidosis.<br>• **Renal function fluctuations (eGFR 10.8–48.2 mL/min/1.73m²):** Admission 2 shows acute kidney injury (AKI) with hyperkalemia (K 5.3 mmol/L), metabolic acidosis ($HCO_3^-$ 26.1 mmol/L, cAG 17.8), and elevated CRP (1.89 mg/dL) and procalcitonin (0.58 ng/mL), indicating infection/inflammation as a likely trigger. Improvement by Admission 3 suggests partial reversibility.<br>• **IgA dynamics (1,482 → 173 → 238 mg/dL):** Initial elevation aligns with smoldering multiple myeloma (SMM), but the sharp drop may reflect acute-phase response or infection impact. Persistent λ-light chain production (SMM) remains a risk for monoclonal gammopathy-related kidney disease.<br>• **Infection markers (WBC 13,600/µL, β-D-glucan 155.8 pg/mL at Admission 2):** Suggest bacterial/fungal infection contributing to AKI and NS exacerbation. Normalization by Admission 3 supports resolved infection.<br>• **Exclusion of amyloidosis:** Duodenal biopsy (no amyloid), echocardiogram (no cardiac involvement), and absent serum PLA2R antibody reduce likelihood of AL amyloidosis or membranous nephropathy.<br>• **Contextual synthesis:**<br>The data suggest NS with acute-on-chronic kidney injury. The low selectivity index and lack of amyloid evidence support MCD, but SMM history necessitates caution for monoclonal gammopathy-related pathology. Infection (Admission 2) likely exacerbated AKI and proteinuria. |
| Gemini 2.5 Pro | Interpretation of Test Results<br>The laboratory data from Admission 3 confirms a state of severe nephrotic syndrome, characterized by massive proteinuria (UP/UCr 5.57 g/gCr) and severe hypoalbuminemia (2.2 g/dL). The kidney function shows CKD stage G3b (eGFR 38.0), which is a significant improvement from the acute kidney injury seen in Admission 2 (eGFR 10.8), likely caused by sepsis (note the elevated WBC, CRP, PCT, and β-D-glucan at that time).<br><br>Several key findings significantly guide the differential diagnosis:<br>• **Highly Selective Proteinuria:** The selectivity index of 0.08 is critically important. This |

| | |
|---|---|
| | value indicates the selective loss of smaller proteins like albumin while larger proteins are retained, a classic hallmark of podocyte effacement with an intact glomerular basement membrane size barrier. This finding strongly points towards **Minimal Change Disease (MCD).**<br>• **Absence of Specific Markers:** The negative anti-PLA2R antibody rules out primary membranous nephropathy. The negative duodenal biopsy and normal echocardiogram make systemic AL or AA amyloidosis much less likely, though a kidney-limited form cannot be absolutely excluded without a kidney biopsy.<br>• **Inflammatory and Immunological Profile:** The persistently elevated ESR and mild leukocytosis may reflect her underlying rheumatoid arthritis or the nephrotic state itself. The normal complement levels (C3, C4) argue against an immune-complex glomerulonephritis. The low IgA level, despite a history of IgA myeloma, is likely due to significant urinary protein losses. |
| OpenAI GPT-o3 | Interpretation of the new data<br>• Persistent nephrotic pattern UP/UCr 8.23→6.46→5.57 g/gCr with marked hypoalbuminaemia (Alb 1.9–2.4 g/dL) confirms ongoing nephrotic-range protein loss.<br>• High selectivity index (0.08) points to highly selective proteinuria, typical for minimal-change disease (MCD) and very unusual for amyloidosis, FSGS or membranous GN.<br>• Complement levels are only mildly depressed and probably reflect hypoproteinaemia rather than immune-complex consumption; they do not suggest lupus or MPGN.<br>• No PLA2R antibody argues against primary membranous nephropathy.<br>• Negative duodenal Congo-red staining and normal echocardiography make systemic AL/AA amyloidosis improbable despite the patient's smouldering myeloma and long-standing RA.<br>• Admission-2 spike (Cr 3.29 mg/dL, BUN 131 mg/dL, β-D-glucan 155 pg/mL, neutrophilic leukocytosis) represents a transient sepsis-related AKI that subsequently resolved (Cr 1.04 mg/dL).<br>• IgA fell from 1,482 to 173 mg/dL, consistent with paraprotein dilution by heavy proteinuria; no other myeloma-related end-organ damage is evident.<br>Overall, the biochemical profile best fits steroid-sensitive, highly selective nephrotic syndrome rather than paraprotein-related or immune-complex glomerulopathy. |
| Llama4-Marveric | Interpretation of Test Results<br>The patient's laboratory results (Table 1) show persistent hypoalbuminemia and significant proteinuria (UP/UCr ratio > 5 g/gCr), consistent with nephrotic syndrome (NS). The lack of evidence for amyloidosis on duodenal biopsy and echocardiogram, along with a low selectivity index and negative M-type phospholipase A2 receptor antibody, suggests Minimal Change Disease (MCD) as a possible diagnosis. The patient's renal function has fluctuated, with a significant decline during Admission 2, possibly due to acute kidney injury (AKI) as indicated by the increased BUN and Cr, and decreased eGFR. |
| **Phase II – Q6: Reconsidering the differential diagnosis** | |
| DeepSeek-R1 | Revised Diagnostic Thinking<br>**Working Diagnosis: Minimal Change Disease (MCD),** likely secondary to SMM (paraneoplastic syndrome).<br>**Reasoning for prioritization:**<br>• **For MCD:**<br>• Low selectivity index (0.08) and absence of hematuria at Admission 3 are classic for MCD.<br>• Steroid responsiveness in NS with SMM has been reported in paraneoplastic MCD.<br>• Exclusion of amyloidosis and membranous nephropathy strengthens this.<br>• **Against monoclonal gammopathy-specific pathology (e.g., AL amyloidosis):**<br>• Negative amyloid biopsies and cardiac imaging.<br>• Improvement in renal function without anti-myeloma therapy. |

| | |
|---|---|
| | • **Other considerations:**<br>• Tacrolimus toxicity is unlikely as NS predates its use, and AKI improved despite continued dosing.<br>• Infection-related NS is unsupported by resolved markers at Admission 3.<br>• **Conclusion:** MCD is most probable, but SMM remains a key risk factor. Renal biopsy (if feasible) would be definitive, but technical constraints favor a steroid trial as a diagnostic-therapeutic measure. |
| Gemini 2.5 Pro | Updated Diagnostic Thinking<br>My diagnostic thinking has shifted substantially. Initially, AL amyloidosis secondary to smoldering myeloma and AA amyloidosis secondary to rheumatoid arthritis were the top two differentials. However, the new data strongly refutes these possibilities. The combination of a negative systemic workup for amyloidosis and, most convincingly, the **highly selective proteinuria**, makes amyloidosis highly improbable, as it typically causes non-selective proteinuria.<br>With primary membranous nephropathy also ruled out, **Minimal Change Disease (MCD)** emerges as the leading diagnosis. This diagnosis elegantly explains the full clinical picture: the abrupt onset of severe nephrotic syndrome, the absence of significant hematuria, and the highly selective proteinuria. While idiopathic MCD is most common, it can occur as a paraneoplastic syndrome in the context of hematologic disorders. Given her history, the MCD is likely secondary to, or associated with, her smoldering multiple myeloma.<br><br>**Working Diagnosis:** Minimal Change Disease (MCD), likely paraneoplastic in etiology, secondary to smoldering multiple myeloma. |
| OpenAI GPT-o3 | Revised diagnostic thinking<br>Initial concern centred on AL/AA amyloidosis or light-chain deposition due to SMM and RA. The combination of (i) highly selective proteinuria, (ii) absence of systemic amyloid on GI biopsy/echo, (iii) negative PLA2R, and (iv) rapid fluctuation of renal function in parallel with volume status now makes minimal-change disease the most plausible explanation. Because tacrolimus and chronic inflammatory disorders can precipitate secondary MCD, the working diagnosis is:<br>Working diagnosis: Secondary minimal-change nephrotic syndrome associated with rheumatoid arthritis/tacrolimus in an elderly woman with smouldering myeloma. |
| Llama4-Marveric | Diagnostic Thinking and Working Diagnosis<br>Given the information and test results, the initial suspicion of MCD is based on the lack of evidence for amyloidosis and other specific causes of NS. The low selectivity index (0.08) is more typical of non-MCD glomerulopathies, but the absence of other clear causes and the patient's response to prednisolone (PSL) could still support a trial of corticosteroids for MCD. The working diagnosis at this point remains **Minimal Change Disease (MCD),** with continued evaluation needed to assess response to treatment and rule out other causes. |
| **Phase II – Q7: Treatment planning** | |
| DeepSeek-R1 | Treatment Plan<br>**Proposed Plan:**<br>• **Continue prednisolone (PSL) 40 mg/day (0.8 mg/kg):**<br>• **Rationale:** First-line EBM for MCD induces remission in >80% of adults within 8–16 weeks. Dose aligns with KDIGO guidelines.<br>• **Monitoring:**<br>• Proteinuria (UP/UCr) and albumin weekly for 4 weeks to assess response.<br>• SCr, glucose, and infection signs (e.g., WBC) biweekly (steroids increase infection risk).<br>• **Adjunctive measures:**<br>• ACE inhibitor (e.g., lisinopril) for antiproteinuric effect if BP permits.<br>• Statin (e.g., atorvastatin) for hypercholesterolemia (LDL-C 83–145 mg/dL).<br>• Prophylactic trimethoprim-sulfamethoxazole for Pneumocystis prevention during |

| | |
|---|---|
| | steroid therapy.<br><br>**Alternative Options & Rationale:**<br>**• If no response to PSL in 4 weeks:**<br>**• Cyclosporine/tacrolimus:** Add if steroid-resistant. Rationale: Calcineurin inhibitors are second-line for MCD. Avoid tacrolimus dose increase (current nephrotoxicity risk).<br>**• Rituximab:** Consider if contraindications to calcineurin inhibitors. Rationale: Effective in steroid-dependent MCD, especially with hematologic disorders.<br>**• If SMM progresses to active myeloma:**<br>**• Anti-myeloma therapy (e.g., bortezomib/dexamethasone):** Rationale: Treating underlying malignancy may resolve paraneoplastic NS.<br><br>**EBM & Individualized Factors:**<br>• Steroids are prioritized due to MCD suspicion and diagnostic-therapeutic role.<br>• Age (88 years) and SMM warrant reduced-intensity approaches (e.g., avoid cyclophosphamide).<br>• Infection history necessitates aggressive prophylaxis. |
| Gemini 2.5 Pro | Treatment Plan<br>The initiation of diagnostic therapy with prednisolone is an appropriate and evidence-based step in the absence of a biopsy, given the high suspicion for MCD. My treatment plan would be as follows:<br>**1. Primary Treatment & Monitoring:**<br>**• Continue Prednisolone (PSL) 40 mg/day (0.8 mg/kg):** This is the standard first-line therapy for adult MCD. The primary goal is to induce remission.<br>**• Rationale:** A positive response (significant reduction or resolution of proteinuria) within 4-8 weeks would serve as a therapeutic confirmation of the MCD diagnosis. The dose is appropriate for an elderly patient to balance efficacy and the risk of side effects.<br>**• Monitoring:** We must monitor weekly urine protein/creatinine ratio to assess for response. We must also closely monitor for steroid side effects, including hyperglycemia, hypertension, and, crucially, signs of infection, given her age and recent septic episode.<br><br>**2. Essential Supportive Care for Nephrotic Syndrome:**<br>**• Thromboembolism Prophylaxis:** Given her severe hypoalbuminemia (albumin <2.5 g/dL), she is at high risk for venous thromboembolism. I would strongly recommend prophylactic anticoagulation (e.g., apixaban, dose-adjusted for renal function) unless there are significant contraindications.<br>**• Edema Management:** Institute a strict sodium-restricted diet (<5g salt/day) and fluid restriction. Judicious use of loop diuretics (e.g., furosemide) is warranted, with careful monitoring of weight, blood pressure, and renal function to prevent volume depletion and AKI.<br>**• Blood Pressure and Proteinuria Control:** While ACE inhibitors or ARBs are beneficial for proteinuria, they should be used with extreme caution in this patient due to the risk of AKI and hyperkalemia, especially when combined with diuretics and CNI (tacrolimus).<br><br>**3. Contingency and Long-Term Plan:**<br>**• If Steroid-Sensitive (Remission achieved):** This confirms the working diagnosis. The plan is to continue high-dose PSL for at least 4 weeks post-remission, followed by a very slow taper over 6-9 months to minimize the risk of relapse.<br>**• If Steroid-Resistant (No remission after 12-16 weeks):** The diagnosis of MCD would be questioned. We would need to reconsider other diagnoses like FSGS or a missed amyloidosis. A second-line agent would be required. A calcineurin inhibitor like cyclosporine would be a standard choice; however, since she is already on tacrolimus for RA, we could consider optimizing its dose to achieve therapeutic levels for NS, while |

| | |
|---|---|
| | vigilantly monitoring for nephrotoxicity. A repeat discussion about the risks and benefits of a kidney biopsy would be necessary. |
| OpenAI GPT-o3 | Treatment plan and rationale<br>A. Disease-directed therapy<br>• Prednisolone 40 mg/day (≈0.8 mg/kg) for 4–8 weeks, then taper according to proteinuria and albumin response. Rationale: MCD is classically steroid-responsive; older age alone does not preclude response.<br>• Re-evaluate tacrolimus; consider dose reduction or discontinuation because calcineurin inhibitors can both induce MCD-like lesions and exacerbate nephrotoxicity.<br>• If steroid-dependent/refractory: switch or add alternate agents (cyclosporine, low-dose rituximab, mycophenolate or low-dose cyclophosphamide), balancing efficacy with frailty and infection risk.<br><br>B. Supportive care<br>• Salt restriction (<4 g/day) and loop ± thiazide diuretics for oedema.<br>• RAAS blockade (ACE-I or ARB) if tolerated to reduce intraglomerular pressure and additional proteinuria.<br>• Statin only if LDL remains elevated after remission; hyperlipidaemia often corrects with resolution of nephrotic syndrome.<br>• Anticoagulation prophylaxis (low-dose aspirin) given age > 80 and serum Alb < 2.0 g/dL, unless bleeding risk is prohibitive.<br>• Vaccinations (influenza, pneumococcal, shingles) before or early in steroid course.<br><br>C. Infection prophylaxis & monitoring<br>• TMP-SMX 80/400 mg three times weekly while on >20 mg/day PSL for >4 weeks (PJP prevention).<br>• Monitor β-D-glucan, WBC, CRP; prompt empiric therapy at first sign of infection.<br>• Serial urinalysis/protein-creatinine ratio, serum Alb, Cr, BP, weight every 1–2 weeks during induction, then monthly.<br><br>D. Myeloma & RA follow-up<br>• Repeat SPEP/IFE and free-light chains every 6 months to detect progression from smouldering to overt myeloma.<br>• Coordinate RA control (low-dose corticosteroid may suffice initially; consider non-nephrotoxic DMARDs such as abatacept if tacrolimus is stopped).<br>This regimen targets the most likely reversible glomerular pathology (MCD) while minimising treatment-related harm and keeping alternative options ready should the response be sub-optimal. |
| Llama4-Marveric | Treatment Plan<br>**1. Continue Prednisolone (PSL) at 40 mg/day:** To assess the response to corticosteroids, which is a standard initial treatment for MCD.<br>**2. Monitor Renal Function and Proteinuria:** Regularly check serum creatinine, eGFR, and UP/UCr to assess the response to PSL and watch for signs of AKI or worsening NS.<br>**3. Adjust Tacrolimus:** Consider adjusting or tapering tacrolimus if it's suspected to contribute to the NS or AKI, although its primary use is for rheumatoid arthritis.<br>**4. Infection Surveillance:** Given the patient's immunosuppressed state (due to tacrolimus and PSL), monitor for signs of infection, as indicated by the elevated β-D-glucan and PCT levels during Admission 2.<br>**5. Alternative Treatments:** If the patient does not respond to PSL, consider alternative immunosuppressive agents or other treatments based on the final diagnosis.<br><br>Rationale: The treatment plan focuses on managing NS, monitoring for complications, and adjusting immunosuppressive therapy as needed. Alternative options would depend on the |

| | |
|---|---|
| | response to initial treatment and further diagnostic clarification. |
| **Phase III – Q8: Evaluation of treatment** | |
| DeepSeek-R1 | Monitoring Effectiveness, Safety, and Risk Management<br>**Effectiveness Monitoring:**<br>**• Nocardiosis treatment:**<br>• Imaging: Biweekly brain MRI to assess abscess size reduction; monthly chest CT for pulmonary nodules.<br>• Labs: Daily CRP/WBC trends; weekly blood/sputum cultures for Nocardia clearance.<br>• Clinical: Daily assessment of fever, abscess size, neurological status.<br>**• Amyloidosis/NS:**<br>• Urinary protein-to-creatinine ratio (UPCR) weekly to track proteinuria.<br>• Serum albumin, creatinine, eGFR biweekly.<br><br>**Safety Monitoring & Specific Risks:**<br>**• Antimicrobial Toxicity:**<br>• Thrombocytopenia (LZD/TZD): Daily CBC; discontinue if platelets <100,000/μL.<br>• Renal toxicity (MEPM/LVFX): Daily SCr/eGFR; avoid nephrotoxins.<br>• QT prolongation (LVFX): Baseline/weekly ECG.<br>**• Immunosuppression Risks:**<br>• Opportunistic infections: Daily β-D-glucan (fungi), chest X-ray weekly; TMP-SMX for Pneumocystis prophylaxis.<br>• Steroid complications: Blood glucose QID; bone density scan monthly.<br>**• Sepsis Risk:**<br>• Procalcitonin q48h if fever recurs; lactate levels for shock.<br>**Concrete Countermeasures:**<br>• Thrombocytopenia: Switch to TMP-SMX (susceptible per Table 2) and platelet transfusion if <50,000/μL.<br>• Rising β-D-glucan: Add voriconazole (covers Aspergillus).<br>• Renal impairment: Reduce MEPM dose if eGFR <30 mL/min. |
| Gemini 2.5 Pro | Treatment Effectiveness and Safety Monitoring Plan<br>**Effectiveness Monitoring:**<br>**1. Infection Response (Nocardiosis):**<br>• **Clinical:** Daily monitoring of temperature, vital signs, and neurological status (for CNS abscess). Regular inspection of skin lesions for signs of resolution.<br>• **Biomarkers:** Monitor C-reactive protein (CRP) and white blood cell count 2-3 times weekly. A consistent downward trend is the primary goal.<br>• **Microbiological:** Repeat cultures from relevant sites (sputum, wound) if clinical worsening occurs, to confirm continued susceptibility.<br>• **Imaging:** Schedule follow-up brain MRI and chest CT scans every 4-6 weeks to objectively assess the size and activity of abscesses and pulmonary nodules.<br>**2. Organ Function & Supportive Care:**<br>• **Renal:** Monitor daily fluid balance, serum creatinine, and electrolytes to ensure euvolemia and prevent acute kidney injury (AKI).<br>• **Nutrition:** Track daily weight, and weekly albumin/prealbumin levels. Anorexia must be addressed with nutritional counseling and support to prevent catabolism, which worsens immunosuppression.<br><br>**Safety Monitoring (Risks & Countermeasures):**<br>**1. Risk: Antimicrobial Drug Toxicity**<br>• **Problem:** Myelosuppression from oxazolidinones (linezolid, tedizolid) and |

| | |
|---|---|
| | nephrotoxicity/neurotoxicity from other agents (meropenem, aminoglycosides).<br>• **Monitoring:** Complete blood count (CBC) 2-3 times per week, specifically for thrombocytopenia and anemia. Monitor renal function closely to allow for dose adjustments.<br>• **Countermeasure:** The observed failure of tedizolid after linezolid-induced thrombocytopenia creates a therapeutic crisis. The plan would be to stop the offending agent immediately. A new regimen must be constructed based on susceptibility testing (Table 2), such as the combination of high-dose meropenem and levofloxacin. If renal function permits, adding amikacin could be considered for synergy, despite its toxicity risks, given the life-threatening nature of the infection.<br>**2. Risk: Worsening Immunosuppression & Secondary Infections**<br>• **Problem:** The patient has multiple risk factors for profound immunosuppression (low IgG/C3, HTLV-1, SMM, malnutrition, prior steroids/tacrolimus).<br>• **Monitoring:** High index of suspicion for new infections. The elevated β-D-glucan (Admission 2) was a critical signal for a fungal co-infection, later confirmed as Aspergillus.<br>• **Countermeasure:**<br>• **Stop Immunosuppressants:** All immunosuppressive medications, including the 8 mg of methylprednisolone, must be discontinued. The risk of fatal infection far outweighs the benefit for her RA or unproven nephrotic syndrome therapy.<br>• **Add Antifungal Therapy:** Given the high β-D-glucan and risk factors, empiric or pre-emptive antifungal therapy targeting Aspergillus (e.g., voriconazole) should be initiated.<br>• **Immune Support:** Consider administering intravenous immunoglobulin (IVIG) to correct the hypogammaglobulinemia and provide passive immunity.<br>**3. Risk: Progression of Undiagnosed Underlying Disease**<br>• **Problem:** The patient's clinical fragility and poor response are likely driven by the undiagnosed systemic AL amyloidosis affecting her kidneys and heart.<br>• **Monitoring:** Monitor for signs of cardiac decompensation (e.g., hypotension, arrhythmia, worsening edema) via NT-proBNP and ECG.<br>• **Countermeasure:** The immediate priority is surviving sepsis. Treatment for amyloidosis is not feasible in the acute setting. If the patient were to stabilize, the diagnosis must be aggressively pursued (e.g., repeat biopsy of an accessible site like fat pad or bone marrow) to allow for future planning, though the prognosis would remain extremely poor. |
| OpenAI GPT-o3 | Monitoring plan<br>Effectiveness<br>• Nephrotic syndrome: spot UP/Cr or 24-h protein, serum albumin, body weight/edema, sCr/eGFR every 1–2 weeks during induction, monthly thereafter.<br>• Nocardiosis: daily temp/clinical status, weekly CRP/WBC, chest CT or MRI brain at 4–6 weeks to document lesion shrinkage, repeat cultures if fever recurs.<br><br>Safety and specific risks – countermeasures<br>• Bacterial / fungal opportunistic infection (high-dose steroids, tacrolimus, linezolid): TMP-SMX for PJP, oral hygiene/skin care, low threshold for cultures and CT; vaccinate (influenza, pneumococcus, zoster).<br>• Linezolid / tedizolid thrombocytopenia: CBC twice weekly; stop or switch if platelets < 100 000/µL or fall > 50 %.<br>• TMP-SMX hyper-K / AKI: CMP twice weekly, stop ACE-I/ARB, give SPS or loop diuretics if $K^+ > 5.5$ mmol/L.<br>• Tacrolimus nephro- and hepato-toxicity, drug–drug interaction with macrolides/azole: trough level once weekly, LFTs fortnightly, reduce or hold if trough > 10 ng/mL or ALT/AST > 3× ULN.<br>• Steroid adverse effects (hyperglycaemia, GI bleed, psychosis, myopathy): finger-stick glucose, PPI prophylaxis, mood monitoring, physio; taper as soon as proteinuria falls < 1 |

| | |
|---|---|
| | g/day.<br>• Thrombo-embolic events from NS: low-dose aspirin or LMWH if Alb < 2.0 g/dL and bleeding risk low; monitor D-dimer, platelets, occult blood.<br>• Hidden plasma-cell progression / AL amyloidosis: serum free-light chains, SPEP/IFE every 3 mo; abdominal-fat or minor-salivary-gland Congo-red biopsy if proteinuria persists despite therapy. |
| Llama4-Marveric | Monitoring Effectiveness and Safety of Treatment<br>To monitor the effectiveness and safety of the treatment for nocardiosis:<br>**1. Clinical Response:** Monitor for resolution of symptoms such as fever, anorexia, and leg ulcers.<br>**2. Imaging:** Repeat chest radiography and brain MRI to assess the reduction in size of lung nodules and brain abscesses.<br>**3. Microbiological Response:** Regularly culture sputum and wound samples to check for Nocardia spp. eradication.<br>**4. Antibiotic Susceptibility Testing:** Continue to monitor antibiotic susceptibility to adjust the treatment regimen as necessary.<br>**5. Side Effects Monitoring:** Watch for adverse effects of antibiotics, such as thrombocytopenia (already observed with LZD and TZD), and adjust the regimen accordingly.<br><br>Specific risks to watch out for include:<br>• Thrombocytopenia and other hematologic adverse effects from LZD and TZD.<br>• Potential resistance or lack of efficacy of the current antibiotic regimen.<br>• Superinfection or development of resistance.<br><br>Countermeasures:<br>• Regular blood tests to monitor for thrombocytopenia and other adverse effects.<br>• Adjust the antibiotic regimen based on susceptibility testing and clinical response.<br>• Consider alternative antibiotics if necessary. |
| **Phase III – Q9: Formulating plans for worsening conditions** | |
| DeepSeek-R1 | Response to Unexpected Deterioration<br>**Thought Process & Reassessment:**<br>• **Immediate differentials:**<br>i. Infection progression: Resistant Nocardia (check susceptibilities), new Aspergillus, or sepsis.<br>ii. Drug toxicity: Antibiotic-induced cytopenia/organ failure.<br>iii. Amyloidosis complications: Cardiac/kidney decompensation.<br>• **Key triggers:** Fever recurrence, neurological decline, cytopenia, or lactate >2 mmol/L.<br><br>**Action Plan:**<br>• **Diagnostic Pivot:**<br>• Repeat MRI brain/chest; bronchoscopy for Aspergillus; blood cultures.<br>• Re-test antibiotic susceptibilities (Nocardia resistance risk).<br>• **Treatment Revision:**<br>• **Infection:** Escalate to amikacin (if susceptible, Table 2) + voriconazole for Aspergillus.<br>• **Cytopenia:** Replace LZD/TZD with TMP-SMX + rifampicin (if susceptible).<br>• **Septic shock:** Start vasopressors, broaden to piperacillin-tazobactam + vancomycin.<br>• **Halt immunosuppressants** if uncontrolled infection.<br>**Rationale:** Prioritize infection control in immunocompromised hosts; align adjustments with susceptibility data (Table 2). |
| Gemini | Approach to Clinical Deterioration |

| | |
|---|---|
| 2.5 Pro | If the patient's condition suddenly worsens or fails to improve, I would adopt a systematic framework: **"Is the diagnosis wrong? Is the treatment failing? Or is there a new complication?"**<br>**Thought Process:**<br>**1. Re-evaluate the Pathogen & Diagnosis:**<br>• **Polymicrobial Infection:** Is Nocardia the only pathogen? The high β-D-glucan strongly suggests a concomitant fungal infection. The final septic shock could have been fungal or a new bacterial superinfection.<br>• **Inadequate Source Control:** Is there an undrained abscess? Nocardiosis requires both prolonged antibiotics and aggressive drainage. The appearance of new skin abscesses despite therapy is a red flag for either hematogenous seeding from an uncontrolled source (like the brain) or inadequate antibiotic tissue penetration/resistance.<br>• **Underlying Disease Impact:** Could the deterioration be cardiac? The autopsy-confirmed cardiac amyloidosis makes the heart extremely vulnerable to the stress of sepsis, leading to a mixed septic-cardiogenic shock.<br>**2. Re-evaluate the Treatment:**<br>• **Antibiotic Failure:** Is the regimen ineffective due to resistance, inadequate dosing, or poor CNS/abscess penetration? The MIC for TMP-SMX was 4, which is borderline susceptible and may be insufficient for a severe, disseminated infection, explaining the failure of maintenance therapy.<br>• **Treatment-Limiting Toxicity:** Did a necessary drug have to be stopped? The thrombocytopenia from linezolid/tedizolid forced a change in therapy, which may have compromised efficacy at a critical moment.<br><br>**Action Plan:**<br>**1. Immediate Stabilization:** Initiate septic shock protocols immediately (e.g., fluid resuscitation, vasopressors, securing the airway).<br>**2. Aggressive Re-evaluation:**<br>• **Pan-culture:** Obtain urgent blood (bacterial and fungal), sputum, and wound cultures before adjusting antibiotics.<br>• **Expand Diagnostics:** Order serum galactomannan antigen (for Aspergillus) and repeat β-D-glucan.<br>• **Urgent Imaging:** Perform a repeat brain MRI and CT of the chest and abdomen/pelvis to search for new or enlarging abscesses that may require surgical drainage.<br>• **Cardiac Assessment:** Obtain an urgent ECG and bedside echocardiogram to assess for acute cardiac decompensation as a contributor to shock.<br>**3. Revise the Treatment Plan Flexibly:**<br>• **Broaden Antimicrobials:** Immediately broaden antimicrobial coverage empirically to cover resistant Nocardia and potential fungal or bacterial superinfections. This would involve continuing high-dose meropenem and levofloxacin, and adding an antifungal agent like voriconazole or an echinocandin.<br>• **Pursue Source Control:** Request an urgent surgical or interventional radiology consultation for drainage of any and all accessible fluid collections or abscesses. This is a critical, non-negotiable step.<br>• **Maximize Supportive Care:** Optimize hemodynamic support and consider adjunctive therapies like IVIG to address the severe immunocompromised state. |
| OpenAI GPT-o3 | Approach to inadequate response or sudden deterioration<br>**Step 1** – Rapid reassessment<br>• Confirm ABCs, obtain vitals, basic labs, blood/urine cultures, chest X-ray ± CT; correct hypovolaemia, give broad-spectrum antibiotics if sepsis suspected.<br>**Step 2** – Re-examine working diagnosis<br>• If proteinuria remains nephrotic after 6–8 wk of adequate steroids, reconsider the original diagnosis; arrange a kidney tissue diagnosis (trans-jugular or ultrasound-guided if anatomy |

| | |
|---|---|
| | permits) or alternative sites (fat pad, rectal, skin, bone marrow) to rule out AL/AA amyloidosis or membranous GN.<br>• Review all drugs for secondary MCD or nephrotoxicity; stop tacrolimus if not already.<br>**Step 3** – Modify therapy<br>• For steroid-dependent/refractory MCD: switch to calcineurin inhibitor-free regimen (low-dose rituximab, mycophenolate, or cyclophosphamide) balancing infection risk.<br>• For uncontrolled nocardiosis: add third active agent (amikacin or imipenem), consult neurosurgery for abscess drainage, extend therapy to ≥ 12 mo; screen for underlying immunodeficiency (IgG, complements, HTLV-1).<br>• If AL amyloidosis confirmed: refer to hematology for bortezomib-based regimen or daratumumab-CyBorD; adjust renal and infection prophylaxis accordingly.<br>**Step 4 –** Escalate care<br>• Early ICU transfer for septic shock, respiratory failure or rapidly rising creatinine; involve infectious-disease, nephrology, hematology, and critical-care teams. |
| Llama4-Marveric | Response to Changes in Patient Condition<br>If the patient's condition does not improve as expected or suddenly changes:<br>**1. Reassess Diagnosis:** Consider alternative diagnoses or complications, such as superinfection or resistance to the current antibiotics.<br>**2. Review Treatment Regimen:** Adjust the antibiotic regimen based on the latest susceptibility testing and clinical response.<br>**3. Imaging and Cultures:** Repeat imaging studies and cultures to assess the current extent of disease and microbiological status.<br>**4. Consider Additional Supportive Care:** Such as nutritional support for anorexia and weight loss, and management of sepsis or organ failure if it occurs.<br><br>Action Plan:<br>• Immediately investigate any sudden change in the patient's condition.<br>• Adjust the treatment plan based on new information from cultures, imaging, or other diagnostic tests.<br>• Consider consulting with specialists in infectious diseases for complex cases.<br><br>By closely monitoring the patient's response to treatment and being prepared to adjust the treatment plan as needed, it's possible to optimize outcomes in a complex case like this. |

| **Case 8: IgA Nephropathy Associated With Infliximab Treatment in Patients With Crohn's Disease: Study of IgA1 and IgA2 Expression in Glomeruli** [9] | |
|---|---|
| **Phase I – Q1: Summarizing medical problems** | |
| DeepSeek-R1 | 46–50-year-old Japanese male with **subacute/chronic kidney injury** characterized by elevated serum creatinine, proteinuria, and microscopic hematuria. Key context: 20-year history of Crohn’s disease (remission on 18-year infliximab therapy), hypertension, hyperuricemia, fatty liver, and sleep apnea. |
| Gemini 2.5 Pro | This is a 46–50-year-old man with a long-standing history of Crohn's disease (in remission on infliximab) and other comorbidities, presenting with recently discovered renal dysfunction, proteinuria, and microscopic hematuria, a clinical picture highly suggestive of active glomerulonephritis. |
| OpenAI GPT-o3 | A 46–50-year-old Japanese man with 20 years of Crohn’s disease (in remission on infliximab for 18 years) and comorbid hypertension, hyperuricemia, fatty liver, and sleep apnea is newly found to have elevated serum creatinine, proteinuria, and microscopic hematuria. |
| Llama4-Marveric | The patient is a 46–50-year-old Japanese man with a 20-year history of Crohn's disease, currently in remission with long-term infliximab treatment, presenting with increased serum creatinine, proteinuria, and microscopic hematuria. His past medical history includes hypertension, hyperuricemia, fatty liver, and sleep apnea syndrome. |
| **Phase I – Q2: Differential diagnoses and rationale** | |

| | |
|---|---|
| DeepSeek-R1 | Differential Diagnoses<br>**Prioritized List:**<br>**1. Amyloidosis (AA type)**<br>• Most likely: Chronic inflammation from Crohn's predisposes to amyloid deposition. Explains proteinuria, hematuria, and rising creatinine.<br>**2. Drug-induced Interstitial Nephritis (e.g., infliximab/NSAIDs)**<br>• Chronic infliximab use rarely causes tubulointerstitial injury; NSAIDs (if used) could contribute.<br>**3. Hyperuricemic/Gouty Nephropathy**<br>• Pre-existing hyperuricemia may cause chronic tubulointerstitial damage or crystal nephropathy.<br><br>**Must Not Miss:**<br>• **Amyloidosis:** High morbidity if undiagnosed; Crohn's duration (>10 years) is a key risk.<br>• **Rapidly Progressive Glomerulonephritis (RPGN):**<br>• Hematuria/proteinuria suggests possible glomerular involvement (e.g., ANCA-associated vasculitis or IgA nephropathy). |
| Gemini 2.5 Pro | Differential Diagnosis<br>**1. IgA Nephropathy (IgAN):** This is the most common glomerulonephritis worldwide and is strongly associated with Crohn's disease due to the gut-kidney axis. The presentation of hematuria, proteinuria, and renal impairment is classic for IgAN.<br>**2. Infliximab-induced Glomerulonephritis:** Anti-TNF-alpha therapies are known to induce autoimmune phenomena, including drug-induced lupus or ANCA-associated vasculitis, which can manifest as glomerulonephritis. The long-term exposure of 18 years makes this a significant possibility.<br>**3. AA Amyloidosis:** A serious complication of chronic inflammatory conditions like Crohn's disease. Although the disease is reportedly in remission, 20 years of history poses a significant cumulative risk. It typically presents with heavy proteinuria and progressive renal failure.<br>• **Most Likely: IgA Nephropathy.** This is the most frequent glomerular disease found in patients with Crohn's disease, and the clinical presentation fits perfectly.<br>• **Must Not Be Missed: AA Amyloidosis.** Due to its poor prognosis if left untreated and its direct link to the patient's long-standing Crohn's disease. A rapidly progressive glomerulonephritis (e.g., secondary to vasculitis) is also a critical consideration. |
| OpenAI GPT-o3 | Differential diagnosis<br>• IgA nephropathy – Most likely because (a) peak incidence in 30-50 yr Asian men, (b) strong association with inflammatory bowel disease, and (c) typical triad of hematuria, proteinuria, and slowly rising creatinine.<br>• ANCA-associated pauci-immune glomerulonephritis – Must not be missed; anti-TNF agents can trigger ANCA vasculitis leading to rapidly progressive renal failure.<br>• AA (secondary) amyloidosis related to long-standing Crohn's disease – Chronic inflammation can cause nephrotic-range proteinuria and renal dysfunction. |
| Llama4-Marveric | Differential Diagnoses<br>**1. IgA Nephropathy:** Given the patient's history of Crohn's disease and the presence of hematuria and proteinuria, IgA nephropathy is a consideration. IgA nephropathy is associated with various chronic inflammatory conditions, including inflammatory bowel disease.<br>**2. Hypertensive Nephrosclerosis:** The patient's history of hypertension could contribute to kidney damage, manifesting as increased serum creatinine, proteinuria, and hematuria.<br>**3. Drug-induced Kidney Damage:** Long-term use of infliximab or other medications could potentially cause kidney damage.<br><br>The most likely diagnosis is **IgA Nephropathy** due to its association with Crohn's disease. **Rapidly Progressive Glomerulonephritis** must not be missed, given the potential for |

<table>
<tr><td></td><td>severe kidney damage if left untreated.</td></tr>
<tr><td colspan="2">Phase I – Q3: Necessary physical examinations and reasons</td></tr>
<tr><td>DeepSeek-R1</td><td>Targeted History/Physical Exam<br>Key Questions:<br>• NSAID/analgesic use? (Rule out analgesic nephropathy)<br>• Edema/foamy urine? (Assess nephrotic syndrome possibility)<br>• Joint pain/tophi? (Evaluate gout activity)<br>• Recent infections? (Trigger for RPGN)<br><br>Physical Exams:<br>• BP measurement (Hypertensive urgency?)<br>• Edema assessment (Nephrotic range proteinuria?)<br>• Joint inspection for tophi (Gout complications)<br>• Abdominal mass/organomegaly (Amyloid involvement)<br><br>Clarify: Whether renal injury is inflammatory (glomerular) vs. metabolic/tubular.</td></tr>
<tr><td>Gemini 2.5 Pro</td><td>Additional History and Physical Examination
<table>
<tr><th>Questions & Examinations</th><th>Purpose</th></tr>
<tr><td>History:<br>1. Onset/duration of symptoms?<br>2. Any gross hematuria, frothy urine, or decreased urine output?<br>3. Any systemic symptoms (fever, rash, joint pain)?<br>4. Full medication list (especially for HTN, hyperuricemia) and use of NSAIDs?</td><td>1. To determine the acuity of the kidney injury (acute vs. chronic).<br>2. To assess the severity of hematuria and proteinuria.<br>3. To screen for systemic autoimmune diseases like drug-induced lupus or vasculitis.<br>4. To identify other potential nephrotoxic agents.</td></tr>
<tr><td>Physical Exam:<br>1. Measure blood pressure.<br>2. Check for peripheral or periorbital edema.<br>3. Inspect skin for purpura or other rashes.<br>4. Examine joints for synovitis.</td><td>1. To assess the severity of hypertension.<br>2. To evaluate for volume overload secondary to proteinuria and renal dysfunction.<br>3. To look for signs of systemic vasculitis or lupus.<br>4. To detect signs of an inflammatory arthropathy.</td></tr>
</table></td></tr>
<tr><td>OpenAI GPT-o3</td><td>Key additional history & physical examination<br>History: recent upper-respiratory infections or macroscopic hematuria (IgA); constitutional symptoms, sinusitis, cough, hemoptysis, arthralgia, skin ulcers (ANCA vasculitis); long-term inflammatory flares, chronic diarrhea, weight loss (AA); timing of renal decline vs. infliximab schedule; NSAID or other nephrotoxin use.<br>Exam: blood pressure trend; edema; skin purpura or ulcers; nasal/oral ulcers; scleritis; lung crackles; arthropathy; hepatosplenomegaly. Purpose—identify systemic features supporting each differential and assess disease severity.</td></tr>
<tr><td>Llama4-Marveric</td><td>To verify the hypotheses, additional questions and physical exams could include:<br>• Detailed urinalysis and quantification of proteinuria.<br>• Blood pressure measurement to assess control of hypertension.<br>• Evaluation of infliximab treatment duration and dosage, and assessment for any other nephrotoxic medications.<br>• Serum IgA levels and other immunological markers.<br>• Renal ultrasound to assess kidney size and structure.<br>These are expected to clarify the extent of kidney damage, the presence of other potential causes of kidney injury, and guide further diagnostic testing.</td></tr>
<tr><td colspan="2">Phase I – Q4: Plan for investigations/tests</td></tr>
<tr><td>DeepSeek-</td><td>Diagnostic Testing Plan</td></tr>
</table>

<table>
<tr><td>R1</td><td>
<table>
<tr><th>Test</th><th>Purpose</th><th>Expected Results/Clues</th></tr>
<tr><td>Urine Protein-Creatinine Ratio (UPCR)</td><td>Quantify proteinuria severity</td><td>>3.5 g/g suggests glomerular disease (e.g., amyloid)</td></tr>
<tr><td>Renal Ultrasound</td><td>Rule out obstruction, assess kidney size</td><td>Small kidneys → chronicity; normal size → acute process</td></tr>
<tr><td>Serum ANCA, ANA, Complement</td><td>Screen for autoimmune glomerulonephritis</td><td>Positive ANCA → vasculitis; low C3 → immune complex disease</td></tr>
<tr><td>Serum Amyloid A (SAA) / Free Light Chains</td><td>Evaluate for amyloidosis</td><td>Elevated SAA supports AA amyloidosis</td></tr>
<tr><td>Uric Acid Level</td><td>Assess hyperuricemia control</td><td>Persistently high → gouty nephropathy</td></tr>
<tr><td>Consider Renal Biopsy</td><td>Gold standard for glomerular/tubular pathology</td><td>Amyloid deposits, interstitial inflammation, or RPGN</td></tr>
</table>
Rationale:<br>
• Initial non-invasive tests (UPCR, ultrasound, serology) screen for common/urgent causes.<br>
• Biopsy reserved if initial tests inconclusive, given invasiveness.<br>
• Cost-effective approach targets Crohn’s-related (amyloidosis), drug-induced, and metabolic etiologies first.
</td></tr>
<tr><td>Gemini 2.5 Pro</td><td>
Diagnostic Plan
<table>
<tr><th>Test</th><th>Purpose and Expected Findings</th></tr>
<tr><td>Blood Tests:<br>- CBC, Comprehensive Metabolic Panel<br>- CRP, ESR</td><td>To assess the degree of renal dysfunction, anemia, and hypoalbuminemia. Inflammatory markers may be elevated in AA amyloidosis or active vasculitis.</td></tr>
<tr><td>Urine Tests:<br>- Urinalysis with microscopy<br>- Urine Protein-to-Creatinine Ratio (UPCR)</td><td>To confirm the glomerular origin of hematuria (dysmorphic RBCs, RBC casts) and to quantify proteinuria, which aids in differential diagnosis and assessing severity.</td></tr>
<tr><td>Serological Tests:<br>- ANA, anti-dsDNA<br>- ANCA (MPO, PR3)<br>- Complements (C3, C4)</td><td>To screen for specific causes. Positive ANA/anti-dsDNA would suggest a lupus-like process (infliximab-induced). Positive ANCA would suggest vasculitis. Low complements are seen in immune-complex GN like lupus nephritis.</td></tr>
<tr><td>Definitive Test:<br>- Renal Biopsy (with Light Microscopy, Immunofluorescence, and Electron Microscopy)</td><td>This is essential for a definitive diagnosis and to guide treatment.<br>- Expected: It will differentiate between IgAN (mesangial IgA deposits), AA amyloidosis (Congo red positive deposits), or various patterns of drug-induced GN (e.g., "full-house" staining in lupus-like nephritis, pauci-immune findings in AAV).</td></tr>
</table>
</td></tr>
<tr><td>OpenAI GPT-o3</td><td>
Planned investigations<br>
• Urinalysis with microscopic exam – look for dysmorphic RBCs/RBC casts (glomerular source) and quantify protein.<br>
• Spot urine protein-creatinine (or 24 h protein) – gauge severity.<br>
• Basic metabolic panel, uric acid – baseline renal function and other nephrotoxins.
</td></tr>
</table>

| | |
|---|---|
| | • Serum IgA level and complement C3/C4 – elevated IgA with normal complement favors IgA nephropathy; low C3 suggests alternative GN.<br>• Autoimmune panel: ANA, anti-dsDNA, anti-histone (drug-induced lupus), MPO-ANCA, PR3-ANCA – screen for ANCA vasculitis or lupus-like syndrome.<br>• Serum amyloid A and SAP scintigraphy (if available) – support AA amyloidosis.<br>• Inflammatory markers (CRP/ESR) – activity of Crohn's/amyloidosis.<br>• Hepatitis B/C, HIV – secondary causes of GN.<br>• Kidney ultrasound – assess chronicity (size, echogenicity) and rule out obstruction.<br>• Definitive: renal biopsy – required to distinguish IgA nephropathy, pauci-immune GN, or amyloid; guides therapy.<br><br>Expected findings: mesangial IgA deposition (IgA nephropathy); crescentic GN with pauci-immune staining (ANCA); Congo-red-positive fibrils in glomeruli (AA amyloidosis). |
| Llama4-Marveric | Tests to Narrow Down the Diagnosis<br>**1. Renal Biopsy:** To definitively diagnose the cause of kidney damage, such as IgA nephropathy or other glomerulopathies.<br>**2. Serological Tests:** Including serum IgA levels, complement levels (C3, C4), and other relevant immunological markers to support a diagnosis of IgA nephropathy or rule out other autoimmune causes.<br>**3. 24-hour Urine Protein Quantification:** To assess the severity of proteinuria.<br>**4. Kidney Function Tests:** Including serum creatinine and eGFR to monitor kidney function.<br>These tests are aimed at confirming the diagnosis, assessing the severity of kidney involvement, and guiding management decisions. |
| **Phase II – Q5: Interpreting test results** | |
| DeepSeek-R1 | Interpretation of Test Results<br>• **Elevated serum IgA (618 mg/dL):** Supports IgA nephropathy (IgAN) diagnosis, as 30-50% of IgAN patients show polyclonal IgA elevation due to aberrant mucosal immunity—potentially linked to his Crohn's disease (mucosal inflammation).<br>• **Subnephrotic proteinuria (0.88 g/g Cr) and dysmorphic hematuria:** Indicates glomerular injury. Dysmorphic RBCs and RBC casts confirm glomerulonephritis, consistent with IgAN.<br>• **Renal biopsy findings:**<br>• Mesangial hypercellularity/expansion: Classic IgAN feature.<br>• Mesangial IgA/C3 deposits: Pathognomonic for IgAN; absence of IgG/C1q rules out lupus nephritis.<br>• Electron-dense deposits: Confirms immune-complex deposition.<br>• **Normal C3/C4:** Excludes post-infectious GN or SLE-related GN.<br>• **Anemia (Hb 11.0 g/dL):** Likely due to chronic kidney disease (CKD) from IgAN, not active Crohn's (remission). |
| Gemini 2.5 Pro | **Interpretation of Test Results**<br>The test results collectively paint a clear picture of active glomerulonephritis with established chronic kidney disease (CKD).<br>• **Evidence of Glomerulonephritis:** The urinalysis is highly significant. The presence of dysmorphic red blood cells and, most importantly, **red blood cell casts**, confirms that the bleeding originates from the glomeruli. The proteinuria of 0.88 g/gCr indicates substantial glomerular barrier dysfunction.<br>• **Severity of Kidney Damage:** The elevated BUN (31 mg/dL) and Creatinine (2.38 mg/dL) indicate a significant reduction in glomerular filtration rate (GFR), corresponding to at least Stage 3b CKD. The normocytic anemia (Hemoglobin 11.0 g/dL) is a common consequence of CKD, likely due to deficient erythropoietin production by the damaged |

| | |
|---|---|
| | kidneys.<br>• **Clues to Etiology:** The elevated serum IgA (618 mg/dL) with normal complement levels is a classic serological pattern for IgA nephropathy (IgAN). The normal complements help to deprioritize other immune-complex diseases like lupus nephritis.<br>• **Definitive Diagnosis via Biopsy:** The renal biopsy provides the definitive answer. The combination of mesangial cell proliferation on light microscopy, dominant mesangial IgA and C3 deposits on immunofluorescence, and mesangial electron-dense deposits on electron microscopy is pathognomonic for IgAN. Critically, the finding of **similar levels of IgA1 and IgA2 subclasses** is a strong indicator of IgAN secondary to a mucosal disease, in this case, the patient's long-standing Crohn's disease. This is because primary IgAN is typically characterized by deposits of galactose-deficient IgA1 alone. |
| OpenAI GPT-o3 | Interpretation of test results<br>• Renal function: Creatinine 2.38 mg/dL (eGFR ≈ 25–30 mL/min/1.73 m²) and BUN 31 mg/dL indicate stage 3b chronic kidney disease.<br>• Urine findings: Sub-nephrotic proteinuria (0.88 g/g Cr) plus dysmorphic hematuria and one RBC cast signify active glomerular injury but not heavy (nephrotic-range) loss.<br>• Serology: Serum IgA markedly elevated (618 mg/dL); complements normal—typical laboratory pattern of IgA nephropathy (IgAN) and argues against immune-complex diseases that lower C3 (e.g., post-infectious GN).<br>• Biopsy: Mesangial hypercellularity/expansion and mesangial electron-dense deposits with dominant IgA (+1) and C3 (+1) but absent C1q/IgG fulfill pathologic criteria for IgAN. Equal IgA1/IgA2 staining does not alter the diagnosis but supports mucosal immune activation, consistent with inflammatory bowel disease.<br>• Other labs: Mild normocytic anemia (Hb 11 g/dL) likely reflects CKD; electrolytes and complements otherwise normal.<br><br>Key point: All data converge on IgA nephropathy as the cause of the patient's slowly progressive renal impairment in the setting of Crohn's disease. |
| Llama4-Marveric | Interpretation of Test Results<br>The patient's laboratory data and renal biopsy results are crucial for understanding his condition. Key findings include:<br>• Elevated serum creatinine (2.38 mg/dL) and BUN (31 mg/dL), indicating impaired kidney function.<br>• Proteinuria (0.88 g/g creatinine) and dysmorphic microscopic hematuria, suggesting glomerular damage.<br>• Elevated serum IgA (618 mg/dL), which is consistent with IgA nephropathy.<br>• Renal biopsy findings: mesangial proliferation and expansion, IgA (1+) and C3 (1+) deposits in mesangial areas on immunofluorescence, and high electron density deposits in the mesangial area on electron microscopy, confirming IgA nephropathy.<br><br>These results are significant because they confirm the diagnosis of IgA nephropathy, a condition associated with Crohn's disease. The presence of hematuria, proteinuria, and renal impairment suggests active kidney involvement. |
| **Phase II – Q6: Reconsidering the differential diagnosis** | |
| DeepSeek-R1 | Diagnostic Thinking and Working Diagnosis<br>• **Revised differential diagnosis:**<br>• **IgA nephropathy (Working Diagnosis):** Biopsy-confirmed; Crohn's disease is a known risk factor for secondary IgAN (shared mucosal immune dysregulation).<br>• **Ruled out:**<br> • Amyloidosis (no amyloid in biopsy, no organomegaly).<br> • Drug-induced nephritis (no tubular findings on biopsy; infliximab rarely causes IgAN).<br> • Lupus nephritis (negative IgG/C1q, normal complement). |

| | |
|---|---|
| | • **Key change:** Initial suspicion of Crohn's-related amyloidosis or drug toxicity shifted to IgAN after biopsy. Elevated IgA and glomerular hematuria align with IgAN in IBD patients. |
| Gemini 2.5 Pro | **Updated Diagnostic Thinking and Working Diagnosis**<br>My initial differential diagnosis included IgA nephropathy, infliximab-induced glomerulonephritis, and AA amyloidosis. The comprehensive test results, particularly the renal biopsy, have provided a definitive diagnosis and clarified the etiology.<br><br>The biopsy findings unequivocally confirm IgA nephropathy. The absence of a "full-house" immunofluorescence pattern or pauci-immune features makes an infliximab-induced lupus-like nephritis or ANCA vasculitis highly unlikely. The biopsy did not show the characteristic amyloid deposits of AA amyloidosis.<br><br>Therefore, the working diagnosis is refined from a broad differential to a specific entity: **IgA Nephropathy, secondary to Crohn's disease.** The link to Crohn's disease is strongly supported by the patient's history, the elevated serum IgA, and the crucial biopsy finding of mixed IgA1 and IgA2 subclass deposition, which reflects the polyclonal IgA production from the gut-associated lymphoid tissue. |
| OpenAI GPT-o3 | Updated diagnostic reasoning<br>Initial concerns included IgAN, ANCA-associated GN, and AA amyloidosis. Renal biopsy definitively shows mesangial IgA deposition without crescents or amyloid and no pauci-immune pattern, effectively excluding ANCA vasculitis and AA amyloidosis. Therefore:<br><br>Working diagnosis: IgA nephropathy (probably secondary/associated with long-standing Crohn's disease) causing CKD G3b-A2. |
| Llama4-Marveric | Diagnostic Thinking and Working Diagnosis<br>Given the patient's history of Crohn's disease, long-term infliximab treatment, and the biopsy-proven diagnosis of IgA nephropathy, the most likely diagnosis (Working Diagnosis) is **IgA Nephropathy associated with Crohn's Disease**. The elevated serum IgA levels and characteristic renal biopsy findings support this diagnosis.<br>The differential diagnoses of hypertensive nephrosclerosis and drug-induced kidney damage are less likely given the biopsy findings and the specific immunofluorescence and electron microscopy results pointing towards IgA nephropathy. |
| **Phase II – Q7: Treatment planning** | |
| DeepSeek-R1 | Treatment Plan<br>**First-line Therapy:**<br>• **ACE inhibitor (e.g., lisinopril) or ARB (e.g., losartan):**<br>• *Rationale*: Reduces proteinuria and slows CKD progression (KDIGO 2021 guidelines). Target BP <130/80 mmHg (given hypertension history).<br>• **Supportive care:**<br>• Salt restriction, statin (for CVD risk), allopurinol (for hyperuricemia), and weight management (fatty liver/sleep apnea).<br><br>**Alternative/Add-on Options:**<br>• **Corticosteroids (e.g., prednisone 0.6-0.8 mg/kg/day for 2-4 months):**<br>• *Rationale*: Consider if proteinuria persists >1 g/day despite ACEi/ARB (TESTING trial protocol). Not urgent here (proteinuria 0.88 g/day).<br>• *Risks*: May exacerbate metabolic comorbidities; avoid if uncontrolled hypertension.<br>• **Optimize Crohn's management:** Continue infliximab (prevents mucosal inflammation-driven IgA overproduction).<br><br>**Monitoring:**<br>• Urine protein-to-creatinine ratio, serum creatinine, and BP every 3 months. |

| | |
|---|---|
| | • Avoid NSAIDs (risk of acute kidney injury).<br><br>**Evidence Basis:**<br>• ACEi/ARB is cornerstone for IgAN with proteinuria >0.5 g/day (KDIGO).<br>• Steroids reserved for high-risk cases (proteinuria >1 g/day + eGFR >50 mL/min). Here, eGFR is ~30 mL/min (CKD-EPI), favoring conservative start.<br><br>**Working Diagnosis:** IgA nephropathy (secondary to Crohn's disease).<br>Please enter the treatment course. |
| Gemini 2.5 Pro | Treatment Plan<br>The primary goals are to slow the progression of chronic kidney disease, reduce proteinuria, and control blood pressure, as these are the main drivers of long-term kidney outcomes.<br><br>**Core Treatment Strategy:**<br>**1. Optimized Supportive Care (Cornerstone of Therapy):** Based on KDIGO guidelines, this is the most critical component for all patients with IgAN at risk of progression.<br>• **RAS Inhibition:** Immediately initiate an Angiotensin-Converting Enzyme inhibitor (ACEi) or an Angiotensin II Receptor Blocker (ARB). The dose will be titrated upwards to the maximum tolerated level, with dual goals of achieving a blood pressure of <130/80 mmHg and reducing proteinuria to less than 1 g/day, ideally aiming for <0.5 g/day.<br>• **SGLT2 Inhibitor**: Initiate an SGLT2 inhibitor (e.g., dapagliflozin, empagliflozin). Landmark trials have shown that these agents significantly slow CKD progression in patients with and without diabetes, including those with IgAN. This is now standard of care for patients with proteinuria and an eGFR >20-25 mL/min/1.73m².<br>• **Lifestyle Modification**: Counsel on a strict low-sodium diet (<2 g/day) to enhance the anti-proteinuric and anti-hypertensive effects of the RAS inhibitor.<br>**2. Management of Underlying Disease:** Continue the current infliximab therapy. The patient's Crohn's disease is in remission, which is crucial. Controlling the underlying inflammatory trigger from the gut is fundamental to managing this secondary form of IgAN.<br><br>**Monitoring and Follow-up:**<br>• The patient will be closely monitored. We will re-evaluate his blood pressure, serum creatinine, and urine protein-to-creatinine ratio in 3 months to assess the response to supportive care.<br><br>**Alternative Options and Rationale:**<br>• **Systemic Corticosteroids:** If, after 3-6 months of optimized supportive care, significant proteinuria (>1 g/day) persists, a 6-month course of systemic corticosteroids could be considered. However, this would be approached with extreme caution. The evidence for steroids is primarily in primary IgAN, and the risks (infection, metabolic complications) are substantial, particularly in a patient already on a biologic agent. Given that his primary disease is controlled, the risk-benefit ratio may not be favorable. This would be a shared decision-making process.<br>• **Targeted-Release Budesonide (Nefecon):** This is a newer therapy approved for primary IgAN that targets gut mucosal immunity. While conceptually appealing for this patient, its efficacy in secondary IgAN is not yet established. It could be a future consideration if other measures fail. |
| OpenAI GPT-o3 | Treatment plan and rationale<br>A. Supportive reno-protective therapy (first-line, strong evidence)<br>1. Optimize blood pressure < 120/80 mmHg with maximal tolerated ACE inhibitor or ARB (RAS blockade lowers intraglomerular pressure and proteinuria).<br>2. Add an SGLT2 inhibitor (e.g., dapagliflozin) if eGFR ≥ 25 mL/min; proven to slow CKD |

| | |
|---|---|
| | progression and reduce proteinuria.<br>3. Lifestyle: low-salt diet (< 5 g/day NaCl), weight control, smoking cessation.<br>4. Monitor proteinuria, eGFR, potassium every 1–3 months initially.<br><br>B. Immunomodulatory options (consider only if proteinuria persists > 0.75–1 g/day after ≥ 3 months of optimized supportive care, per KDIGO 2021)<br>• Oral corticosteroid course (e.g., prednisolone 0.6–0.8 mg/kg/day tapered over 6 months) or targeted-release budesonide (NEFIGAN regimen) if eGFR > 30 mL/min and infection risk acceptable; weigh against Crohn's remission status and long-term infliximab.<br>• Tonsillectomy with short-course steroids—option mainly practiced in Japan for persistent active IgAN.<br><br>C. Review concomitant medications<br>• Continue infliximab if Crohn's remains quiescent and no evidence it exacerbates IgAN; switch biologic only if future data suggest causality.<br>• Avoid NSAIDs and other nephrotoxins; ensure uric acid control (allopurinol/febuxostat) because hyperuricemia can accelerate CKD.<br><br>D. Management of CKD complications<br>• Treat anemia of CKD (iron studies, ESAs as needed).<br>• Educate on vaccination (pneumococcal, hepatitis B, influenza).<br>• Plan for renal replacement therapy if progression (timely referral for transplant evaluation, given Crohn's in remission).<br><br>Alternative therapies (lower evidence)<br>• Fish-oil (omega-3 FAs) 3–4 g/day for proteinuria reduction.<br>• Immunosuppressants beyond steroids (e.g., MMF, cyclophosphamide) are not recommended in typical IgAN without crescents.<br>Follow-up: reassess proteinuria and renal function at 3 months; escalate therapy only if targets unmet or renal decline accelerates. |
| Llama4-Marveric | Treatment Plan<br>The treatment plan for this patient should focus on managing IgA nephropathy, controlling hypertension, and monitoring kidney function.<br>**1. Renin-Angiotensin System (RAS) Inhibitors:** To control proteinuria and hypertension, given their protective effects on the kidneys.<br>**2. Immunosuppressive Therapy:** May be considered if there's significant kidney inflammation or rapid progression, though the evidence is more nuanced for IgA nephropathy.<br>**3. Monitoring:** Regular follow-up of kidney function, proteinuria, and hematuria to assess disease progression and response to treatment.<br>**4. Management of Crohn's Disease:** Continue monitoring and managing Crohn's disease, as its activity may influence IgA nephropathy.<br>**5. Lifestyle Modifications:** Including dietary adjustments, blood pressure control, and avoidance of nephrotoxic substances.<br><br>Rationale: The treatment is based on evidence-based guidelines for IgA nephropathy and considers the patient's specific condition, including the presence of Crohn's disease and hypertension.<br><br>Alternative options could include more aggressive immunosuppression or novel therapies under clinical investigation for IgA nephropathy. The choice of treatment should be tailored to the patient's specific clinical context and disease severity. |
| **Phase III – Q8: Evaluation of treatment** | |
| DeepSeek- | NA |

| | |
|---|---|
| R1 | |
| Gemini 2.5 Pro | NA |
| OpenAI GPT-o3 | NA |
| Llama4-Marveric | NA |
| **Phase III – Q9: Formulating plans for worsening conditions** | |
| DeepSeek-R1 | NA |
| Gemini 2.5 Pro | NA |
| OpenAI GPT-o3 | NA |
| Llama4-Marveric | NA |

| | |
|---|---|
| **Case 9: Delayed ureteric obstruction secondary to blood clot following renal biopsy: Case report and literature review** [10] | |
| **Phase I – Q1: Summarizing medical problems** | |
| DeepSeek-R1 | 56–60-year-old male with >10-year diabetes mellitus history, presenting with **chronic (2-month) bilateral symmetrical lower limb edema.** |
| Gemini 2.5 Pro | A 56–60-year-old male with a 10-year history of diabetes mellitus presents with a subacute (2-month) onset of progressive, bilateral lower limb edema, suggesting a systemic cause of fluid retention, likely related to a complication of his long-standing diabetes. |
| OpenAI GPT-o3 | 56–60-year-old man with >10-year history of diabetes mellitus presents with insidious, progressive bilateral lower-limb edema lasting 2 months. |
| Llama4-Marveric | The patient is a 56–60-year-old male with a 10-year history of diabetes mellitus, presenting with bilateral lower limb edema for 2 months. |
| **Phase I – Q2: Differential diagnoses and rationale** | |
| DeepSeek-R1 | Differential Diagnoses<br>**1. Diabetic nephropathy/nephrotic syndrome** (Most likely)<br>• *Rationale*: Long-standing diabetes is the top risk factor. Chronic edema suggests nephrotic-range proteinuria.<br>**2. Heart failure** (Must not be missed)<br>• *Rationale*: Diabetes predisposes to cardiovascular disease. Bilateral edema could indicate volume overload.<br>**3. Hepatic cirrhosis**<br>• *Rationale*: Chronic edema may reflect hypoalbuminemia or portal hypertension, though less directly linked to diabetes. |
| Gemini 2.5 Pro | Differential Diagnoses<br>I would consider the following three differential diagnoses:<br>**1. Diabetic Nephropathy causing Nephrotic Syndrome:** This is the most likely diagnosis. The long duration of diabetes mellitus (>10 years) is a classic risk factor for developing diabetic nephropathy, a leading cause of nephrotic syndrome. Nephrotic syndrome is characterized by heavy proteinuria, which leads to hypoalbuminemia and generalized edema.<br>**2. Congestive Heart Failure (CHF):** This is the must-not-miss diagnosis. Diabetes is a major independent risk factor for cardiovascular disease and heart failure. CHF leads to increased hydrostatic pressure and fluid retention, causing bilateral pitting edema. It is a common and potentially life-threatening condition that requires specific and urgent management.<br>**3. Chronic Liver Disease (Cirrhosis):** Non-alcoholic fatty liver disease (NAFLD) is strongly associated with diabetes and can progress to cirrhosis. Cirrhosis causes edema through hypoalbuminemia (due to decreased synthesis) and portal hypertension. |

<table>
<tr><td>OpenAI<br>GPT-o3</td><td>Differential Diagnosis<br>a. Diabetic nephropathy causing nephrotic-range proteinuria or CKD with salt/water retention – most likely given long-standing diabetes and typical bilateral edema.<br>b. Congestive heart failure (CHF) – common, potentially life-threatening; would also explain bilateral edema.<br>c. Hepatic cirrhosis / severe chronic liver disease – hypoalbuminemia leads to peripheral edema; must not be missed if present.<br>(The “must-not-miss” is CHF because of its acute morbidity and need for rapid treatment.)</td></tr>
<tr><td>Llama4-<br>Marveric</td><td>Differential Diagnoses<br>1. Diabetic Nephropathy: Given the patient's long-standing diabetes, diabetic nephropathy is a likely cause of his edema due to potential hypoalbuminemia from nephrotic syndrome.<br>2. Heart Failure: Fluid retention and edema can be symptoms of heart failure, which is more common in diabetic patients due to diabetic cardiomyopathy.<br>3. Venous Insufficiency or Deep Vein Thrombosis: Edema can also be caused by venous insufficiency or deep vein thrombosis, conditions that may be more prevalent in diabetic patients.<br><br>The most likely diagnosis is Diabetic Nephropathy, considering the patient's history of diabetes mellitus.<br><br>A diagnosis that must not be missed is Deep Vein Thrombosis, as it can lead to life-threatening pulmonary embolism if not treated promptly.</td></tr>
<tr><td colspan="2">Phase I – Q3: Necessary physical examinations and reasons</td></tr>
<tr><td>DeepSeek-<br>R1</td><td>Targeted History & Physical Exam
<table>
<tr><th>Action</th><th>Purpose</th></tr>
<tr><td>History: Ask about orthopnea, paroxysmal nocturnal dyspnea (PND)</td><td>Rule out cardiac failure (e.g., fluid redistribution at night).</td></tr>
<tr><td>History: Inquire about frothy urine or recent weight gain</td><td>Assess for nephrotic syndrome (proteinuria) vs. fluid retention.</td></tr>
<tr><td>Physical: Check for elevated JVP, lung crackles</td><td>Identify cardiac overload signs.</td></tr>
<tr><td>Physical: Examine for ascites/spider angiomata</td><td>Evaluate liver involvement.</td></tr>
<tr><td>Physical: Assess for periorbital edema and hypertension</td><td>Support nephrotic syndrome (e.g., diabetic nephropathy).</td></tr>
</table></td></tr>
<tr><td>Gemini<br>2.5 Pro</td><td>Additional History and Physical Examination<br>To differentiate these possibilities, I would gather more information:<br>• Additional History:<br>• Question: "Have you noticed any frothy or foamy appearance in your urine?"<br>• Purpose: To screen for heavy proteinuria, a hallmark of nephrotic syndrome.<br>• Question: "Do you experience shortness of breath when lying flat (orthopnea) or when exerting yourself? Have you ever woken up at night feeling like you can't breathe (paroxysmal nocturnal dyspnea)?"<br>• Purpose: To screen for symptoms of congestive heart failure.<br>• Question: "Have you noticed any abdominal swelling, yellowing of your skin or eyes (jaundice), or easy bruising?"<br>• Purpose: To identify signs of advanced liver disease (ascites, jaundice, coagulopathy).<br><br>• Physical Examination:<br>• Maneuver: Measure blood pressure and assess the Jugular Venous Pressure (JVP).<br>• Purpose: Hypertension is common in chronic kidney disease. An elevated JVP is a highly specific sign of volume overload, most commonly due to heart failure.<br>• Maneuver: Auscultate the heart and lungs.</td></tr>
</table>

<table>
<tr><td></td><td>• Purpose: To listen for an S3 gallop (a sign of heart failure) or bibasilar crackles (pulmonary edema).<br>• Maneuver: Palpate the abdomen for a fluid wave or shifting dullness.<br>• Purpose: To detect ascites, which points toward either severe fluid overload or primary liver disease.</td></tr>
<tr><td>OpenAI GPT-o3</td><td>Key History & Physical Examination<br>History<br>• Change in urine (foamy, dark, volume) → proteinuria / hematuria.<br>• Dyspnea, orthopnea, paroxysmal nocturnal dyspnea, weight gain → CHF.<br>• Abdominal distension, jaundice, alcohol use, viral hepatitis risk → liver disease.<br>• Medications (NSAIDs, TZDs) → drug-induced edema or renal injury.<br>• Glycemic control, retinopathy, neuropathy → severity/duration of diabetic complications.<br><br>Physical Examination<br>• Vital signs & blood pressure → hypertension of CKD vs low BP in liver disease.<br>• Jugular venous pressure, S3, lung crackles → CHF.<br>• Abdominal exam for hepatomegaly, splenomegaly, ascites, caput medusae → liver disease.<br>• Fundoscopy for diabetic retinopathy → supports diabetic nephropathy.<br>• Peripheral pulses, skin changes → vascular/venous causes.</td></tr>
<tr><td>Llama4-Marveric</td><td>Additional Questions and Physical Exams<br>To verify these hypotheses, additional questions and physical exams would include:<br>• Questions about the patient's urine output, proteinuria history, and any recent changes in blood sugar control.<br>• Assessment of cardiac function, such as history of heart failure, coronary artery disease, and symptoms like orthopnea or paroxysmal nocturnal dyspnea.<br>• Examination for signs of venous insufficiency or deep vein thrombosis, such as warmth, tenderness, or cord-like swelling in the legs.<br>• Physical examination for signs of fluid overload (e.g., jugular venous distension, pulmonary edema).<br><br>These would help clarify the etiology of the edema and guide further diagnostic testing.</td></tr>
<tr><td colspan="2">Phase I – Q4: Plan for investigations/tests</td></tr>
<tr><td>DeepSeek-R1</td><td>Diagnostic Testing Plan
<table>
<tr><th>Test</th><th>Purpose</th><th>Expected Result</th></tr>
<tr><td>Urinalysis + ACR</td><td>Detect proteinuria</td><td>Heavy proteinuria (nephrotic range) supports diabetic nephropathy.</td></tr>
<tr><td>Serum albumin, creatinine/eGFR</td><td>Evaluate nephrotic syndrome & renal function</td><td>Low albumin, elevated creatinine in nephropathy; normal in cardiac/liver causes.</td></tr>
<tr><td>BNP/NT-proBNP</td><td>Screen for heart failure</td><td>Elevated if cardiac etiology.</td></tr>
<tr><td>Liver function tests (ALT, AST, ALP)</td><td>Rule out hepatic cirrhosis</td><td>Normal in diabetic nephropathy; abnormal in liver disease.</td></tr>
<tr><td>Renal ultrasound</td><td>Assess kidney size/structure</td><td>Bilateral reduced size in chronic diabetic nephropathy.</td></tr>
</table>
<br>Tests prioritized for cost-effectiveness and non-invasiveness, beginning with urinalysis and basic blood work.</td></tr>
</table>

| | |
|---|---|
| Gemini 2.5 Pro | Diagnostic Testing Plan<br>My testing plan would be as follows:<br>**1. Urinalysis and Urine Protein-to-Creatinine Ratio (UPCR):**<br>• **Purpose:** To quantify proteinuria, which is essential for diagnosing nephrotic syndrome.<br>• **Expected Result:** In diabetic nephropathy, I expect to see significant proteinuria on urinalysis (3+ or 4+) and a UPCR >3.5 g/g, confirming nephrotic-range proteinuria.<br>**2. Comprehensive Metabolic Panel (CMP):**<br>• **Purpose:** To assess kidney function (serum creatinine, BUN), serum albumin levels, and liver enzymes.<br>• **Expected Result:** In nephrotic syndrome, I expect an elevated creatinine (indicating renal dysfunction) and a low serum albumin (<3.0 g/dL). Abnormal liver function tests would suggest liver disease.<br>**3. Brain Natriuretic Peptide (BNP or NT-proBNP):**<br>• **Purpose:** To screen for heart failure. This peptide is released in response to cardiac ventricular stretch.<br>• **Expected Result**: A normal or low BNP level makes symptomatic heart failure very unlikely. A high level would increase suspicion for CHF and prompt further cardiac evaluation.<br>**4. Echocardiogram:**<br>• **Purpose:** If heart failure is suspected (based on symptoms or an elevated BNP), an echocardiogram is necessary to assess cardiac structure, ejection fraction, and diastolic function.<br>• **Expected Result:** This would be normal in isolated nephrotic syndrome but would show systolic or diastolic dysfunction in a patient with CHF. |
| OpenAI GPT-o3 | Investigations<br>• Urinalysis + urine protein/creatinine ratio: confirm proteinuria; nephrotic-range supports diabetic nephropathy.<br>• Serum creatinine, eGFR, electrolytes: assess renal function.<br>• Serum albumin & lipid profile: hypoalbuminemia and hyperlipidemia typical of nephrotic syndrome.<br>• BNP and echocardiography: evaluate CHF; elevated BNP or reduced EF would confirm cardiac cause.<br>• Liver panel (AST, ALT, ALP, bilirubin), coagulation profile: screen for cirrhosis.<br>• Renal ultrasound: kidney size (small in chronic GN, enlarged in diabetic nephropathy), exclude obstruction.<br>Optional / next step<br>• Kidney biopsy if proteinuria is heavy but atypical for diabetic nephropathy (e.g., sudden onset, active urinary sediment). |
| Llama4-Marveric | Tests to Narrow Down the Diagnosis<br>**1. Urine Protein/Creatinine Ratio or 24-hour Urine Protein:** To assess for significant proteinuria indicative of diabetic nephropathy or other glomerulopathies.<br>**2. Serum Albumin and Lipid Profile:** To evaluate for hypoalbuminemia and hyperlipidemia, common in nephrotic syndrome.<br>**3. Renal Ultrasound:** To assess kidney size and echotexture, and rule out obstructive causes.<br>**4. Echocardiogram:** To evaluate cardiac function and structure, helping diagnose heart failure.<br>**5. Lower Limb Venous Doppler Ultrasound:** To diagnose deep vein thrombosis or venous insufficiency.<br><br>These tests are aimed at confirming the suspected diagnoses while considering the patient's underlying diabetes and the presenting symptom of bilateral lower limb edema. |

| **Phase II – Q5: Interpreting test results** | |
|---|---|
| DeepSeek-R1 | Interpretation of Test Results<br>• **Nephrotic Syndrome Confirmation:**<br>• **Proteinuria** (urine protein 3+, UPC ratio ≥300 mg/gCr, 24-hour protein 7.58 g) confirms nephrotic-range proteinuria (>3.5 g/day).<br>• **Hypoalbuminemia** (albumin 18 g/L): Drives edema via reduced plasma oncotic pressure.<br>• **Hyperlipidemia** (cholesterol 8.2 mmol/L): Compensatory hepatic synthesis due to urinary protein loss.<br>• **Atypical Hematuria** (urine RBC 96.9/μL): Uncommon in pure diabetic nephropathy, suggesting possible non-diabetic glomerular pathology (e.g., membranous nephropathy).<br>• **Preserved Renal Function** (Scr 87 μmol/L, eGFR ~70-80 mL/min): Inconsistent with advanced diabetic nephropathy, which typically shows elevated Scr.<br>• **Hypocalcemia** (1.86 mmol/L): Likely due to urinary loss of vitamin D-binding protein in nephrotic syndrome.<br>• **Normal Kidney Size on Ultrasound:** Rules out chronic atrophy but doesn't exclude early diabetic nephropathy or other glomerulopathies.<br><br>**Context:** In a diabetic patient, nephrotic syndrome is most commonly attributed to diabetic nephropathy. However, the preserved eGFR and hematuria are atypical, raising suspicion for a secondary glomerular disease. |
| Gemini 2.5 Pro | **Interpretation of Test Results**<br>The laboratory and physical findings collectively paint a clear picture of **Nephrotic Syndrome.** The key findings are:<br>• **Massive Proteinuria (7.58 g/day) and Severe Hypoalbuminemia (18 g/L):** These are the core features. The kidneys are leaking an exceptionally large amount of protein, primarily albumin, into the urine. This loss overwhelms the liver's ability to produce albumin, causing a sharp drop in its serum concentration.<br>• **Bilateral Pitting Edema and Hypertension (146/81 mmHg):** The severe hypoalbuminemia reduces plasma oncotic pressure, causing fluid to shift from the vasculature into the interstitial space, resulting in edema. The hypertension is a common consequence of glomerular disease and the associated fluid retention.<br>• **Hyperlipidemia (Total Cholesterol 8.2 mmol/L):** This is a reactive process where the liver, stimulated by low oncotic pressure, increases the synthesis of lipoproteins, leading to high cholesterol levels.<br>• **Significant Hematuria (Urine red cells 96.9/µL):** This finding is critically important. While classic diabetic nephropathy is a possible cause of nephrotic syndrome in this patient, it typically presents with a "bland" urinary sediment (minimal or no red blood cells). The presence of significant hematuria strongly suggests an active, inflammatory glomerular process and raises the possibility of a non-diabetic primary glomerulonephritis (e.g., Membranous Nephropathy, IgA Nephropathy, FSGS). |
| OpenAI GPT-o3 | Interpretation of test results<br>• Nephrotic-range proteinuria: urine protein/creatinine ≥300 mg/g and 24 h protein 7.58 g clearly satisfy the definition (>3.5 g/day).<br>• Marked hypoalbuminemia (Alb 18 g/L) and hypercholesterolaemia (TC 8.2 mmol/L) complete the biochemical triad of nephrotic syndrome and explain the bilateral pitting oedema.<br>• Serum creatinine 87 µmol/L indicates preserved glomerular filtration; therefore the process is at an early stage or predominantly proteinuric rather than nephritic.<br>• Microscopic haematuria (≈97 RBC/µL) is present; in long-standing diabetic nephropathy it is usually minimal, so this raises the possibility of a non-diabetic glomerulopathy superimposed on diabetes. |

| | |
|---|---|
| | • Normal kidney size on ultrasound is compatible with both diabetic nephropathy and most primary glomerular diseases.<br>• Mild anaemia (Hb 119 g/L) and hypocalcaemia (Ca 1.86 mmol/L) are common secondary findings in nephrotic syndrome due to chronic disease and urinary loss of vitamin-D–binding protein. |
| Llama4-Marveric | Interpretation of Test Results<br>The laboratory results reveal several key findings:<br>• **Severe Hypoalbuminemia (Albumin 18 g/L):** Indicates significant loss of albumin, likely through the urine given the high level of proteinuria.<br>• **Significant Proteinuria (24-hour protein excretion 7.58 g):** Confirms nephrotic syndrome, characterized by heavy proteinuria (>3.5 g/24 hours), hypoalbuminemia, and edema.<br>• **Hyperlipidemia (Total Cholesterol 8.2 mmol/L):** Commonly seen in nephrotic syndrome due to increased hepatic synthesis of lipids in response to hypoalbuminemia.<br>• **Mild Renal Impairment (Serum Creatinine 87 μmol/L):** Indicates some degree of kidney dysfunction but not severe.<br>• **Normal Coagulation Indicators:** Rules out significant coagulopathy that could complicate the clinical picture or treatment.<br>• **Urine Red Blood Cell Count (96.9/μL):** Suggests some degree of hematuria, which could be seen in various glomerulopathies.<br><br>These findings, in the context of the patient's long-standing diabetes, strongly support a diagnosis of diabetic nephropathy with nephrotic syndrome. |
| **Phase II – Q6: Reconsidering the differential diagnosis** | |
| DeepSeek-R1 | Updated Diagnostic Thinking<br>• **Revised Differentials:**<br>**i. Primary glomerulopathy** (e.g., membranous nephropathy): Elevated due to nephrotic syndrome with hematuria and preserved eGFR.<br>**ii. Diabetic nephropathy:** Still possible but less likely due to preserved eGFR and hematuria.<br>**iii. Amyloidosis/other secondary causes:** Lower probability (no systemic symptoms or organomegaly).<br><br>• **Working Diagnosis: Nephrotic syndrome likely due to non-diabetic glomerulopathy** (e.g., membranous nephropathy), with diabetes as a comorbidity.<br>*Rationale*: Hematuria and preserved eGFR are red flags against classic diabetic nephropathy, warranting biopsy to exclude alternative etiologies. |
| Gemini 2.5 Pro | **Revised Diagnostic Thinking and Working Diagnosis**<br>The new information confirms the syndrome but significantly alters the likely underlying cause. Initially, diabetic nephropathy was the most probable diagnosis due to the patient's history. However, the presence of significant hematuria alongside nephrotic-range proteinuria makes uncomplicated diabetic nephropathy less likely to be the sole diagnosis.<br><br>My diagnostic thinking has shifted to prioritize a primary glomerular disease.<br>• **Working Diagnosis: Nephrotic Syndrome, secondary to a primary glomerulonephritis requiring biopsy for definitive diagnosis.**<br>• **Rationale:** The combination of nephrotic syndrome with an "active" urinary sediment (significant hematuria) is a classic indication for a kidney biopsy in a patient with diabetes. This is to rule out a treatable non-diabetic kidney disease that may be co-existing with or mimicking diabetic nephropathy. The most likely possibilities now include **Membranous Nephropathy** or **Focal Segmental Glomerulosclerosis (FSGS)**, which require different management strategies than diabetic nephropathy. |
| OpenAI | Diagnostic reasoning and working diagnosis |

| | |
|---|---|
| GPT-o3 | The data establish nephrotic syndrome. Given the >10-year history of diabetes, diabetic nephropathy remains the leading cause, but the combination of:<br>− heavy, relatively sudden proteinuria,<br>− preserved eGFR, and<br>− significant microscopic haematuria<br>makes a superimposed primary glomerular disease (especially membranous nephropathy or focal-segmental glomerulosclerosis) plausible.<br>Working Diagnosis: Nephrotic syndrome, most likely diabetic nephropathy; non-diabetic kidney disease cannot be excluded and a renal biopsy is indicated for definitive classification. |
| Llama4-Marveric | Diagnostic Thinking Update<br>Given the significant proteinuria, hypoalbuminemia, hyperlipidemia, and edema, the most likely diagnosis is **Diabetic Nephropathy with Nephrotic Syndrome**. The presence of hematuria might also suggest a superimposed glomerulonephritis, but diabetic nephropathy remains the primary consideration given the patient's diabetic history. |
| **Phase II – Q7: Treatment planning** | |
| DeepSeek-R1 | Treatment Plan<br>**Immediate Management:**<br>**1. ACE Inhibitor (e.g., Ramipril):**<br>• *Rationale*: Reduces proteinuria and controls BP (target <130/80 mmHg) per KDIGO guidelines. EBM shows 30-40% proteinuria reduction in nephrotic syndrome.<br>**2. Diuretics (e.g., Furosemide):**<br>• *Rationale*: Addresses edema; monitor electrolytes to avoid AKI.<br>**3. Statin (e.g., Atorvastatin):**<br>• *Rationale*: Manages hyperlipidemia; reduces cardiovascular risk (EBM: ↓ LDL by 50-60%).<br>**4. Albumin Support + Salt Restriction:**<br>• *Rationale*: Mitigates edema (albumin <20 g/L warrants cautious infusion if diuretic-resistant).<br><br>**Definitive Diagnosis & Next Steps:**<br>• **Kidney Biopsy:** Critical to confirm pathology (e.g., membranous vs. diabetic nephropathy).<br>• **Diabetes Optimization:** Tight glycemic control (HbA1c <7%) to slow renal decline regardless of diagnosis.<br><br>**Alternative Options:**<br>• **Immunosuppression (if biopsy-confirmed MN):** Rituximab or cyclophosphamide (per KDIGO).<br>• **Avoid NSAIDs/Nephrotoxins:** Prevent AKI in nephrotic state.<br><br>**Rationale:** Biopsy-directed therapy is essential—empirical steroids are avoided due to diabetes and unclear diagnosis. Supportive care aligns with EBM for nephrotic syndrome complications. |
| Gemini 2.5 Pro | **Treatment Plan**<br>The treatment plan must be two-pronged: immediate management of the nephrotic syndrome complications and establishing a definitive diagnosis to guide specific therapy.<br><br>**A. Immediate Supportive and General Management:**<br>**i. Control Edema and Blood Pressure:**<br>• **Treatment:** Initiate a loop diuretic (e.g., Furosemide) to manage volume overload and a renin-angiotensin-aldosterone system (RAAS) inhibitor, such as an ACE inhibitor or an ARB. |

| | |
|---|---|
| | • **Rationale:** Diuretics will alleviate the symptomatic edema. RAAS blockade is crucial for its dual benefit of lowering systemic blood pressure and reducing intraglomerular pressure, which helps decrease proteinuria. This is a cornerstone of therapy for nearly all proteinuric kidney diseases.<br>**ii. Prevent Thromboembolism:**<br>• **Treatment:** Start prophylactic anticoagulation.<br>• **Rationale:** The patient's serum albumin is <20 g/L, placing him at high risk for venous and arterial thrombosis due to urinary loss of anticoagulant proteins. This is a life-threatening complication of severe nephrotic syndrome.<br>**iii. Manage Hyperlipidemia:**<br>• **Treatment:** Begin statin therapy.<br>• **Rationale:** To lower the high cholesterol and mitigate the significantly increased cardiovascular risk associated with nephrotic syndrome.<br>**iv. Dietary Modification:**<br>• **Treatment:** Institute a strict low-sodium diet (<2 g/day).<br>• **Rationale:** To work in concert with diuretics to control edema and hypertension.<br><br>**B. Definitive Diagnosis and Specific Therapy:**<br>**v. Most Critical Next Step: Kidney Biopsy.**<br>• **Rationale:** This is mandatory to establish the precise histological diagnosis. The treatment for primary Membranous Nephropathy (e.g., immunosuppression with rituximab or cyclophosphamide) is fundamentally different from the management of diabetic nephropathy (which focuses on glycemic control, RAAS blockade, and SGLT2 inhibitors). The biopsy result will be the sole determinant of long-term, disease-modifying therapy.<br>**vi. Serological Workup:**<br>• **Rationale:** While awaiting biopsy, blood tests for anti-PLA2R antibodies, ANA, and viral serologies (Hepatitis B/C, HIV) should be sent to investigate potential causes of a primary or secondary glomerulonephritis. |
| OpenAI GPT-o3 | Treatment plan and rationale<br>A. General nephrotic-syndrome management<br>• Salt (<2 g Na/day) and fluid restriction; monitor daily weight.<br>• Loop diuretic (e.g., furosemide) ± thiazide sequential blockade for oedema control.<br>• High-intensity statin (e.g., atorvastatin 40 mg) for secondary prevention and lipid control.<br>• Thrombosis prophylaxis: consider low-dose anticoagulation (e.g., rivaroxaban 10 mg/d) because Alb < 25 g/L and DM increase risk; weigh bleeding risk.<br>• Vaccinations (pneumococcal, influenza) to reduce infection risk.<br><br>B. Kidney- and cardio-protective therapy<br>• Start/uptitrate ACE inhibitor or ARB unless contraindicated; target BP <130/80 mm Hg and maximise antiproteinuric effect.<br>• Add SGLT2 inhibitor (e.g., dapagliflozin) if eGFR ≥30 mL/min to further reduce proteinuria and slow progression (strong RCT evidence in diabetic CKD).<br>• Optimise glycaemic control (HbA1c 6.5–7.5 %) with metformin (if eGFR adequate) and/or GLP-1 receptor agonist.<br><br>C. Diagnostic step<br>Renal biopsy after oedema is partially controlled, because the result will determine whether immunosuppression is needed.<br>− If biopsy confirms primary membranous nephropathy or FSGS: initiate risk-stratified immunosuppression (e.g., rituximab or cyclophosphamide-steroid regimen).<br>− If classic diabetic nephropathy: continue conservative therapy; no immunosuppression |

| | |
|---|---|
| | required.<br><br>D. Supportive measures<br>• Correct hypocalcaemia with cholecalciferol ± calcitriol after vitamin-D level.<br>• Erythropoiesis-stimulating agent only if anaemia becomes symptomatic or Hb <100 g/L.<br>• Diet: adequate protein (0.8 g/kg/d) to avoid further catabolism but not exacerbate proteinuria.<br><br>Alternative/adjunct options<br>• Endothelin receptor antagonist (e.g., atrasentan) in future if proteinuria remains refractory (currently investigational/selected use).<br>• Sodium restriction plus mineralocorticoid receptor antagonist (finerenone) for additional renoprotective benefit in diabetic CKD.<br>Frequent follow-up (weight, BP, Scr/eGFR, electrolytes, urine protein) is essential to titrate therapy and detect complications early. |
| Llama4-Marveric | Treatment Plan<br>**1. Manage Nephrotic Syndrome:**<br>• **Angiotensin-Converting Enzyme Inhibitors (ACEi) or Angiotensin Receptor Blockers (ARB):** To reduce proteinuria and slow the progression of kidney disease.<br>• **Diuretics:** To control edema, with careful monitoring of volume status and electrolytes.<br>• **Statins:** To manage hyperlipidemia.<br>**2. Optimize Glycemic Control:** Tight blood sugar control is crucial to slow the progression of diabetic nephropathy.<br>**3. Monitor and Manage Blood Pressure:** Targeting a blood pressure that balances renal protection with the risk of hypotension.<br>**4. Consider Renal Biopsy:** If there's a suspicion of a non-diabetic glomerulopathy or if the patient doesn't respond to initial therapy, a renal biopsy might be considered.<br>**5. Lifestyle Modifications:** Including dietary changes (e.g., low salt, controlled protein intake) and avoiding nephrotoxic substances.<br><br>The rationale is based on evidence that controlling proteinuria, blood pressure, and hyperlipidemia, along with tight glycemic control, can slow the progression of diabetic nephropathy and manage symptoms of nephrotic syndrome.<br><br>**Alternative Options:**<br>• Immunosuppressive therapy if a biopsy reveals a specific glomerulonephritis that could benefit from such treatment.<br>• SGLT2 inhibitors, which have been shown to have renoprotective effects in diabetic patients.<br><br>The treatment plan should be tailored to the patient's specific needs and monitored closely for efficacy and adverse effects. |
| **Phase III – Q8: Evaluation of treatment** | |
| DeepSeek-R1 | Monitoring Effectiveness, Safety, and Risk Management<br>**Effectiveness Monitoring:**<br>• **Glycemic control:** HbA1c every 3 months (target <7%).<br>• **Renal function:**<br>• Serum creatinine/eGFR weekly until stable, then every 3 months.<br>• Urine protein-to-creatinine ratio (UPCR) monthly (target reduction ≥50% from baseline).<br>• **Hematuria resolution:** Urinalysis + microscopy weekly for 4 weeks, then as needed.<br>• **Edema/volume status:** Daily weights and clinical assessment for fluid overload. |

**Safety Risks & Countermeasures:**

| Risk | Monitoring | Concrete Countermeasure |
|---|---|---|
| **Ureteral re-obstruction** | Flank pain, hematuria, rising Scr | Urgent renal ultrasound; if confirmed, repeat stent placement. |
| **Bleeding/hematoma** | Hb trend, hemodynamics, abdominal CT | Avoid anticoagulants; bed rest + transfusion if Hb drop >20 g/L. |
| **AKI progression** | Scr/eGFR 2–3×/week during instability | Discontinue nephrotoxins (e.g., NSAIDs); optimize hydration. |
| **Stent complications** | Fever, dysuria, persistent hematuria | Urine culture; stent exchange if infection/encrustation. |
| **Thromboembolism** | Oxygen saturation, limb swelling, D-dimer | Prophylactic LMWH if albumin <20 g/L; compress stockings. |

Gemini 2.5 Pro

Monitoring Plan for Treatment Effectiveness and Safety

**Monitoring Treatment Effectiveness:**

**1. Renal Function and Obstruction Resolution:**

• **Parameter:** Serum creatinine (Scr) and estimated GFR (eGFR).

• **Frequency:** Check at each follow-up visit (e.g., monthly for 3 months, then every 3 months).

• **Goal:** Maintain a stable Scr/eGFR at the patient's new baseline (approx. 88 μmol/L). A significant increase would trigger an investigation for recurrence.

**2. Underlying Diabetic Nephropathy:**

• **Parameter:** Urine protein-to-creatinine ratio (UPCR) or 24-hour urine protein excretion.

• **Frequency:** Monthly for 3 months, then every 3 months.

• **Goal:** A sustained reduction in proteinuria, which is the primary surrogate marker for slowing disease progression.

**3. Hematuria Resolution:**

• **Parameter:** Urinalysis with microscopy.

• **Frequency**: At each follow-up visit.

• **Goal:** Resolution of gross hematuria and significant reduction or normalization of microscopic hematuria.

**4. Overall Disease Control:**

• **Parameters:** Blood pressure and HbA1c.

• **Frequency:** At each follow-up visit.

• **Goals:** Blood pressure <130/80 mmHg (due to proteinuria) and an individualized HbA1c target (typically ~7.0%) to slow the progression of diabetic complications.

**Monitoring for Risks and Countermeasures:**

**1. Risk: Recurrent Bleeding and Ureteral Obstruction.**

• **Monitoring:** Vigilant monitoring for symptoms of flank pain or gross hematuria. Patient education on these warning signs is critical.

• **Countermeasures:** Avoid anticoagulants and antiplatelet agents unless there is an overriding and compelling indication. Any future invasive procedures would require extremely careful risk-benefit analysis and planning.

**2. Risk: Thromboembolism (Venous and Arterial).**

• **Monitoring:** Clinical surveillance for signs of DVT (unilateral leg swelling/pain) or PE (sudden dyspnea, pleuritic chest pain).

• **Countermeasures:** The patient's albumin level of 18 g/L puts him at very high risk. However, given the severe post-biopsy bleeding, initiating prophylactic anticoagulation is a complex decision. A careful risk-benefit discussion with the patient is essential. Anticoagulation might be cautiously considered only after a prolonged period (e.g., several weeks to months) of documented hematuria resolution.

| | |
|---|---|
| | **3. Risk: Progression to End-Stage Renal Disease (ESRD).**<br>• **Monitoring:** Track the slope of eGFR decline and the trend in proteinuria over time.<br>• **Countermeasures:** Ensure adherence to optimized medical therapy, including a RAAS inhibitor (ACEi or ARB), an SGLT2 inhibitor (now standard of care for proteinuric diabetic nephropathy), statin therapy, and strict glycemic and blood pressure control.<br>**4. Risk: Infection.**<br>• **Monitoring:** Watch for any signs of infection, particularly urinary tract infections, which can be a risk post-stenting.<br>• **Countermeasures:** Recommend annual influenza and pneumococcal vaccinations due to the immunocompromised state associated with nephrotic syndrome. |
| OpenAI GPT-o3 | Monitoring plan (effectiveness & safety)<br>Effectiveness<br>• Weight / lower-limb edema: daily at home, every visit in clinic—objective marker of salt-water control.<br>• Blood pressure: home BP diary; target <130/80 mm Hg with ACE-I/ARB ± SGLT2-i.<br>• Serum creatinine, eGFR, electrolytes: 1 week after any drug change, then every 1–3 months—tracks renal function and drug tolerance.<br>• Spot urine protein-to-creatinine ratio (or 24 h protein) and urinalysis with RBC count: monthly for 3 months, then quarterly—goal ≥50 % proteinuria reduction, absence of gross/microscopic hematuria.<br>• HbA1c and fasting glucose: every 3 months—assess glycaemic control.<br>• Lipid profile, albumin: every 6 months—response to statin and nutritional status.<br>• Imaging (renal-urinary ultrasound): immediately if flank pain, oliguria, or recurrent hematuria; routine at 3–6 months post-stent removal.<br><br>Safety / specific risks & countermeasures<br>1. Recurrent post-biopsy hemorrhage or ureteric obstruction<br>• Watch for new flank pain, gross hematuria, rising Scr.<br>• Countermeasure: urgent CT/ultrasound, rapid urology referral; prepare for repeat stenting or embolization.<br>2. Acute kidney injury (AKI) from obstruction, drugs, or hypovolemia<br>• Serial Scr/eGFR, urine output; avoid nephrotoxins; adjust ACE-I/ARB and diuretics promptly.<br>3. Infection (UTI, perinephric abscess, diabetic foot, pneumonia)<br>• Periodic urinalysis, fever surveillance; early cultures and antibiotics; vaccination (pneumococcal, influenza).<br>4. Venous thrombo-embolism (Alb < 25 g/L)<br>• Assess leg pain, dyspnea; D-dimer if suspected.<br>• Countermeasure: mobilization, compression stockings; consider prophylactic anticoagulation if albumin <20 g/L and no bleeding risk.<br>5. Drug adverse effects<br>• ACE-I/ARB/SGLT2-i: monitor $K^+$ and eGFR; treat hyperkalemia with diuretics/resin, hold drug if $K^+ > 6$ mmol/L.<br>• Statin: baseline and annual CK/LFT; stop if myopathy or severe transaminase rise.<br>• Diuretics: check $Na^+$, $K^+$, uric acid; adjust dose or add supplementation.<br>6. Hypoglycemia from intensified diabetes therapy<br>• Teach SMBG, recognize symptoms; adjust agents if eGFR changes.<br>Patient education<br>− Provide written warning signs (gross hematuria, anuria, severe flank pain, sudden weight gain, dyspnea).<br>− Direct phone line for rapid contact; instruct to seek ER care if any red-flag symptom occurs. |
| Llama4- | Monitoring Treatment Effectiveness and Safety |

| | |
|---|---|
| Marveric | To monitor the effectiveness and safety of the treatment, the following measures will be taken:<br>**1. Renal Function Monitoring:** Regularly check serum creatinine and urine output to assess kidney function.<br>**2. Hematuria Monitoring:** Monitor urine red blood cell count to assess resolution of hematuria.<br>**3. Imaging Studies:** Perform ultrasound or CT scans as needed to check for resolution of the hematoma and ureteral obstruction.<br>**4. Blood Pressure and Glycemic Control:** Regularly monitor blood pressure and blood glucose levels to ensure they are within target ranges.<br>**5. Electrolyte Monitoring:** Check electrolyte levels to prevent or manage any imbalances caused by diuretics or other treatments.<br><br>Specific risks to watch out for include:<br>• **Ureteral Stent Complications:** Such as stent migration, encrustation, or infection.<br>• **Bleeding Complications**: Continued or recurrent bleeding from the biopsy site or ureter.<br>• **Infection:** Urinary tract infections or sepsis related to the stent or catheter.<br><br>Countermeasures:<br>• **Regular Follow-Up:** Schedule regular follow-up appointments to monitor for complications and adjust treatment as necessary.<br>• **Antibiotic Prophylaxis:** Consider antibiotic prophylaxis for patients with ureteral stents to prevent urinary tract infections.<br>• **Stent Removal:** Plan for timely removal of the ureteral stent to minimize the risk of complications. |
| **Phase III – Q9: Formulating plans for worsening conditions** | |
| DeepSeek-R1 | Response to Unexpected Deterioration<br>**Thought Process:**<br>**1. Rapidly reassess causes:**<br>• If **worsening hematuria/flank pain**: Prioritize ureteral clot recurrence vs. new bleed (e.g., ruptured arteriovenous fistula).<br>• If **rising Scr:** Differentiate prerenal (dehydration), intrinsic (glomerular insult), or postrenal (obstruction).<br>• If **treatment failure:** Consider undiagnosed pathology (e.g., IgA nephropathy) or stent malfunction.<br>**2. Key triggers:** Hemoglobin drop >15 g/L, Scr increase >50%, or new systemic symptoms (e.g., fever).<br><br>**Action Plan:**<br>• **Immediate steps:**<br> • Labs: CBC, Scr, coagulation panel, urinalysis.<br> • Imaging: Renal ultrasound (bedside) → CT urogram if obstruction suspected.<br>• **Treatment revision:**<br> • **Obstruction recurrence:** Ureteral stent reinsertion ± clot evacuation.<br> • **Refractory bleeding:** Angioembolization for arterial bleed.<br> • **Infection:** Culture-directed antibiotics; stent removal if infected.<br>• **Diagnosis refinement:** Repeat biopsy if glomerular disease is suspected despite diabetic nephropathy.<br><br>**Rationale:** Early imaging and urology consultation prevent irreversible injury. Conservative management fails if obstruction persists >48 hours, necessitating intervention. |

| | |
|---|---|
| Gemini 2.5 Pro | Action Plan for Unexpected Clinical Changes<br>If the patient's condition fails to improve or suddenly deteriorates, I would adopt a systematic approach to reassess the situation rapidly.<br><br>**1. Immediate Stabilization and Focused Reassessment:**<br>• **Action:** First, assess vital signs and stabilize the patient as needed (the ABCs of resuscitation).<br>• **Thought Process:** I would conduct a focused history and physical examination to precisely characterize the change. Is the primary problem recurrent pain, worsening AKI, increased edema, or a new systemic symptom like chest pain? This initial categorization guides the entire diagnostic process.<br><br>**2. Differential Diagnosis and Targeted Investigation based on Clinical Scenario:**<br>• **Scenario A: Recurrence of Flank Pain, Gross Hematuria, and/or AKI.**<br>• **Thought Process:** My primary hypothesis would be a **recurrent post-renal obstruction** due to a new blood clot from the biopsy site. The differential would include a passed stone or, less likely, a complication from the prior stent removal.<br>• **Action Plan:**<br>a. **STAT Labs:** CBC (to check for acute blood loss), CMP (to quantify AKI), and urinalysis.<br>b. **Urgent Imaging:** A renal ultrasound is the first-line test to look for hydronephrosis. A non-contrast CT of the kidneys, ureters, and bladder (CT-KUB) would follow immediately if ultrasound is non-diagnostic, as it is excellent for identifying ureteral clots or stones.<br>c. **Consultation:** Immediate urology consultation is mandatory for potential intervention.<br>d. **Intervention:** If obstruction is confirmed, treatment would involve repeat ureteral stenting or percutaneous nephrostomy (PCN) to relieve the obstruction. If active, uncontrolled hemorrhage is suspected, an urgent consultation with interventional radiology for potential renal artery embolization would be necessary.<br><br>• **Scenario B: Worsening Proteinuria and Edema without a change in flank pain.**<br>• **Thought Process:** This points towards a progression of the underlying glomerular disease. The key questions are whether this is due to suboptimal treatment or an evolution of the disease itself. I would assess adherence to diet and medications, and the adequacy of the current treatment regimen.<br>• **Action Plan:**<br>a. **Review and Optimize:** Meticulously review medication adherence, blood pressure, and glycemic control. Intensify therapy by maximizing doses of RAAS inhibitors and diuretics, and ensuring the patient is on an SGLT2 inhibitor.<br>b. **Re-evaluate Diagnosis:** While the biopsy showed diabetic nephropathy, a refractory or rapidly progressive course is atypical. Although a repeat biopsy is extremely high-risk in this patient, I would have to consider the remote possibility of a sampling error or a superimposed, treatable glomerulonephritis if all other measures fail, after an extensive discussion of the life-threatening bleeding risk.<br><br>• **Scenario C: Sudden Onset of Dyspnea or Chest Pain.**<br>• **Thought Process:** My top two differential diagnoses, both life-threatening, would be pulmonary embolism (PE), given the severe nephrotic syndrome, and acute coronary syndrome (ACS), given the patient's diabetes.<br>• **Action Plan:**<br>a. **Immediate Workup:** Obtain an urgent ECG, cardiac enzymes, and D-dimer.<br>b. **Definitive Imaging:** Proceed to a CT pulmonary angiogram (CTPA) if PE is suspected. |

| | |
|---|---|
| | **c. Consult and Treat:** Involve cardiology immediately for suspected ACS. If PE is confirmed, it necessitates immediate therapeutic anticoagulation, a decision now forced by a life-threatening event despite the bleeding risk. |
| OpenAI GPT-o3 | Approach if clinical course deviates<br>Step-wise reassessment<br>1. Confirm the change: repeat vitals, weight, physical exam; stat labs (CBC, Scr/eGFR, electrolytes, albumin, coagulation, urinalysis).<br>2. Identify pattern:<br>• Hematuria/colic → think re-bleed or obstruction.<br>• Rising Scr without hematuria → prerenal/drug-induced vs intrinsic AKI.<br>• Worsening edema/proteinuria → progression or superimposed GN.<br>3. Focused investigations: urgent renal/urinary ultrasound ± contrast-enhanced CT; ECG/BNP if heart failure suspected; cultures if febrile; immunologic work-up if new active sediment.<br>4. Immediate actions while investigating:<br>• Hold nephrotoxic and RAAS-blocking drugs if AKI >30 % rise in Scr.<br>• Insert Foley and measure hourly urine; give IV fluids or diuretics as indicated.<br>• Consult urology for obstruction, interventional radiology for embolization, or nephrology for consideration of repeat biopsy if non-diabetic GN suspected.<br>5. Re-evaluate original diagnosis and treatment plan: be willing to escalate (e.g., immunosuppression) or de-escalate (stop anticoagulation) based on new data.<br>6. Communicate with patient/family: explain findings, expected course, and next steps; document changes.<br>This structured monitor-and-respond strategy maximizes therapeutic benefit while minimizing complications in the post-biopsy diabetic nephropathy patient. |
| Llama4-Marveric | Managing Unexpected Changes or Lack of Improvement<br>If the patient's condition does not improve as expected or suddenly changes, the following steps will be taken:<br>**1. Reassess the Diagnosis:** Review the initial diagnosis and consider alternative diagnoses or complicating factors.<br>**2. Imaging and Laboratory Tests:** Perform additional imaging studies or laboratory tests to identify any new or worsening conditions.<br>**3. Consult Other Specialists:** Consider consulting urologists, radiologists, or other specialists as needed to manage complications or provide additional insights.<br>**4. Adjust Treatment Plan:** Based on new findings, adjust the treatment plan to address any identified issues or complications.<br><br>The thought process involves:<br>• **Rapid Reassessment:** Quickly re-evaluating the patient's condition to identify the cause of the change.<br>• **Flexibility:** Being prepared to adjust the treatment plan in response to new information or changing clinical circumstances.<br><br>By closely monitoring the patient's condition and being prepared to adapt the treatment plan, the goal is to optimize outcomes and minimize complications. |

**Note:** Due to policy restrictions, specific age details are not provided. See the original publication for further information.

2. Tseng CW, Liao JH, Chao TK, Hsu SN. New Onset of Primary Membranous Nephropathy After

COVID-19 mRNA Vaccination in Affected Sjögren's Syndrome. *Nephrology (Carlton)*. 2025;30(5):e70048.
